## Supporting information for "Characterising COVID-19 empirical research production in Latin America and the Caribbean: a scoping review"

### S1 Supporting information

#### 1. Search strategies

Search strategy used to search articles in OVID (Medline and EMBASE)

| # | QUERY |
|---|---|
| 1 | ((latin or south or central or north or hispanic) adj america*) or carib* |
| 2 | exp Latin America/ or exp Caribbean Region/ |
| 3 | Belize* or "costa rica" or "costa rican" or Salvador* or Guatemala* or hondura* or mexic* or Nicaragua* or panama* or argentin* or Bolivia* or brazil* or Colombia* or chile* or Ecuador* or Guiana* or guyan* or Paraguay* or peru* or suriname* or Uruguay* or Venezuela* or cuba* or Haiti or "Puerto rico" or "Puerto rican" or Anguill* or Aruba* or Barbad* or Bermud* or "Cayman Islands" or Caymanian or Grenad* or Jamaica* or Antilles or "Saint Lucia" or "saint Lucian" or "Turks and Caicos" or Antigua* or Baham* or Dominica* or "virgin islands" or "virgin islanders" or Monstserrat* or "Saint Kitts and Nevis" or kittitian or Nevisian or "Saint Vincent and the Grenadines" or Trinidad* or Tobago* or trinis or trinbagonian or Curacao* or Guadeloupe* or Martiniqu* or "Sint Maarten" or "saint martin" or "saint martinois" |
| 4 | Coronavir* or 2019-nCoV or 2019nCoV or nCoV or SARS-CoV-2 or COVID19 or COVID-19 or “severe acute respiratory syndrome” or SARS |
| 5 | 1 or 2 or 3 |
| 6 | 4 and 5 |
| 7 | limit 6 to yr=”2019-Current” |

Search strategy used to search articles in LILACS (using VHL)

| # | QUERY |
|---|---|
| 1 | ((latin OR south OR central OR north OR hispanic) AND america*) OR carib* OR belize* OR "costa rica" OR "costa rican" OR salvador* OR guatemala* OR hondura* OR mexic* OR nicaragua* OR panama* OR argentin* OR bolivia* OR brazil* OR colombia* OR chile* OR ecuador* OR guiana* OR guyan* OR paraguay* OR peru* OR suriname* OR uruguay* OR venezuela* OR cuba* OR haiti OR "Puerto rico" OR "Puerto rican" OR anguill* OR aruba* OR barbad* OR bermud* OR "Cayman Islands" OR caymanian OR grenad* OR jamaica* OR antilles OR "Saint Lucia" OR "saint Lucian" OR "Turks and Caicos" OR antigua* OR baham* OR dominica* OR "virgin islands" OR "virgin islanders" OR monstserrat* OR "Saint Kitts and Nevis" OR kittitian OR nevisian OR "Saint Vincent and the Grenadines" OR trinidad* OR tobago* OR trinis OR trinbagonian OR curacao* OR guadeloupe* OR martiniqu* OR "Sint Maarten" OR "saint martin" OR "saint martinois") AND (coronavir* OR 2019-ncov OR 2019ncov OR ncov OR sars-cov-2 OR covid19 OR covid-19 OR “severe acute respiratory syndrome” OR sars) |

Search strategy used to search articles in CENTRAL (the Cochrane Library)

| # | QUERY |
|---|---|
| 1 | ((latin or south or central or north or hispanic) and america*) or carib* |
| 2 | MeSH descriptor: [Latin America] explode all trees |
| 3 | MeSH descriptor: [Caribbean Region] explode all trees |
| 4 | Belize* or "costa rica" or "costa rican" or Salvador* or Guatemala* or hondura* or mexic* or Nicaragua* or panama* or argentin* or Bolivia* or brazil* or Colombia* or chile* or Ecuador* or Guiana* or guyan* or Paraguay* or peru* or suriname* or Uruguay* or Venezuela* or cuba* or Haiti or "Puerto rico" or "Puerto rican" or Anguill* or Aruba* or |

| | |
|---|---|
| | Barbad* or Bermud* or "Cayman Islands" or Caymanian or Grenad* or Jamaica* or Antilles or "Saint Lucia" or "saint Lucian" or "Turks and Caicos" or Antigua* or Baham* or Dominica* or "virgin islands" or "virgin islanders" or Monstserrat* or "Saint Kitts and Nevis" or kittitian or Nevisian or "Saint Vincent and the Grenadines" or Trinidad* or Tobago* or trinis or trinbagonian or Curacao* or Guadeloupe* or Martiniqu* or "Sint Maarten" or "saint martin" or "saint martinois" |
| 5 | coronavir* or "2019-nCoV" or 2019nCoV or nCoV or "SARS-CoV-2" or COVID19 or "COVID-19" or “severe acute respiratory syndrome” or SARS |
| 6 | 1 or 2 or 3 or 4 |
| 7 | 5 and 6 |
| 8 | with Publication Year from 2019 to present , in Trials |

Search strategy used to search articles in Scielo

| # | **QUERY** |
|---|---|
| 1 | ((latin or south or central or north or hispanic) and america*) or carib* |
| 2 | Belize* or "costa rica" or "costa rican" or Salvador* or Guatemala* or hondura* or mexic* or Nicaragua* or panama* or argentin* or Bolivia* or brazil* or Colombia* or chile* or Ecuador* or Guiana* or guyan* or Paraguay* or peru* or suriname* or Uruguay* or Venezuela* or cuba* or Haiti or "Puerto rico" or "Puerto rican" or Anguill* or Aruba* or Barbad* or Bermud* or "Cayman Islands" or Caymanian or Grenad* or Jamaica* or Antilles or "Saint Lucia" or "saint Lucian" or "Turks and Caicos" or Antigua* or Baham* or Dominica* or "virgin islands" or "virgin islanders" or Monstserrat* or "Saint Kitts and Nevis" or kittitian or Nevisian or "Saint Vincent and the Grenadines" or Trinidad* or |

| | |
|---|---|
| | Tobago* or trinis or trinbagonian or Curacao* or Guadeloupe* or Martiniqu* or "Sint Maarten" or "saint martin" or "saint martinois" |
| 3 | coronavir* or "2019-nCoV" or 2019nCoV or nCoV or "SARS-CoV-2" or COVID19 or "COVID-19" or "severe acute respiratory syndrome" or SARS |
| 4 | (1 or 2) AND 3 |
| 5 | Published 2019 AND 2020 |

Search strategy used to search articles in Epistemonikos

| # | QUERY |
|---|---|
| 1 | (title:((((latin OR south OR central OR north OR hispanic) AND america*) OR carib*) OR (Belize* OR "costa rica" OR "costa rican" OR Salvador* OR Guatemala* OR hondura* OR mexic* OR Nicaragua* OR panama* OR argentin* OR Bolivia* OR brazil* OR Colombia* OR chile* OR Ecuador* OR Guiana* OR guyan* OR Paraguay* OR peru* OR suriname* OR Uruguay* OR Venezuela* OR cuba* OR Haiti OR "Puerto rico" OR "Puerto rican" OR Anguill* OR Aruba* OR Barbad* OR Bermud* OR "Cayman Islands" OR Caymanian OR Grenad* OR Jamaica* OR Antilles OR "Saint Lucia" OR "saint Lucian" OR "Turks AND Caicos" OR Antigua* OR Baham* OR Dominica* OR "virgin islands" OR "virgin islanders" OR Monstserrat* OR "Saint Kitts AND Nevis" OR kittitian OR Nevisian OR "Saint Vincent AND the Grenadines" OR Trinidad* OR Tobago* OR trinis OR trinbagonian OR Curacao* OR Guadeloupe* OR Martiniqu* OR "Sint Maarten" OR "saint martin" OR "saint martinois")) OR abstract:((((latin OR south OR central OR north OR hispanic) AND america*) OR carib*) OR (Belize* OR "costa rica" OR "costa rican" OR Salvador* OR Guatemala* OR hondura* OR mexic* OR Nicaragua* OR panama* OR argentin* OR Bolivia* OR brazil* OR Colombia* OR chile* OR Ecuador* OR Guiana* OR guyan* OR Paraguay* OR peru* OR suriname* OR |

| | |
|---|---|
| | Uruguay* OR Venezuela* OR cuba* OR Haiti OR "Puerto rico" OR "Puerto rican" OR Anguill* OR Aruba* OR Barbad* OR Bermud* OR "Cayman Islands" OR Caymanian OR Grenad* OR Jamaica* OR Antilles OR "Saint Lucia" OR "saint Lucian" OR "Turks AND Caicos" OR Antigua* OR Baham* OR Dominica* OR "virgin islands" OR "virgin islanders" OR Monstserrat* OR "Saint Kitts AND Nevis" OR kittitian OR Nevisian OR "Saint Vincent AND the Grenadines" OR Trinidad* OR Tobago* OR trinis OR trinbagonian OR Curacao* OR Guadeloupe* OR Martiniqu* OR "Sint Maarten" OR "saint martin" OR "saint martinois"))) AND (title:(coronavir* OR "2019-nCoV" OR 2019nCoV OR nCoV OR "SARS-CoV-2" OR COVID19 OR "COVID-19" OR "severe acute respiratory syndrome" OR SARS) OR abstract:(coronavir* OR "2019-nCoV" OR 2019nCoV OR nCoV OR "SARS-CoV-2" OR COVID19 OR "COVID-19" OR "severe acute respiratory syndrome" OR SARS)) |
| 2 | Primary studies From 2019 |

#### 2. Data extraction fields

| **DATA SOURCES** | |
|---|---|
| *Source where the data that is analyzed in the study comes from. This category is used to identify the country later* | |
| Inert sources | e.g., sewage, surfaces,air particles, etc. The paper needs to specificy that the country where the sources come from to be included |
| Animals | |
| Directly humans | Information is directly collected from humans |
| Databases | Human information that was not directly collected from humans (i.e., someone else collected the data before the research) |
| Documents | Documentary analysis (e.g., analyses of government documents, bulletins, reports, etc.) |
| Not described | If it is not described, the study should be excluded (e.g., the study analyze molecules coming from samples that the article does not specify where do they come from) |
| Other | |

| **BROAD METHODOLOGY PARADIGM** |
|---|
| *High-level methodology that the study uses* |
| Basic science |

| |
|---|
| Quantitative |
| Qualitative |
| Mixed-methods |

| **STUDY DESIGNS** | |
|---|---|
| *Depending on the methodology selected, we could use several options here* | |
| **Basic sciences** | We will not identify the study design if the study is about basic sciences for now |
| **Quantitative** | |
| Randomized controlled trial | |
| Non-randomized controlled trial | Patients allocated to intervention and control group without randomization |
| Controlled before and after (including diff in diff analysis) | At least two groups are compared before and after an intervention |
| Single before and after | Only one group is compared before and after an intervention/exposure |
| Interrupted time series | Time series are compared before and after the intervention is implemented. It is differently from a descriptive time series |
| Propensity score matching | Two groups are compared matching by several variables |
| Cohort study | |

| | |
|---|---|
| Case-control study | |
| Cross-sectional study | |
| Ecological study | Where the unit of analysis is not a patient but a group of patients (e.g., comparing the rates of two different countries, cities) |
| Descriptive time series | Time series that are used to only describe trends |
| Simulation model used to predict | |
| Validation of a simulation model | |
| Economic evaluation | Cost-utility, cost-effectiveness, cost-benefit analysis |
| Cost study | Budget analysis, just measuring how much does it cost an intervention |
| Other | |
| **Qualitative** | |
| Case-study | There are single or multiple case studies, where cases (institutions, organizations, etc) are compared in their processes |
| Narrative | Study to describe the main codes coming from speaches or information collected from people |
| Ethnography | |

| | |
|---|---|
| Phenomenological | |
| Grounded theory | |
| Other | |
| **Mixed methods** | |
| Exploratory | Quantitative is conducted after qualitative phase |
| Explanatory sequential | Qualitative is conducted after quantitative phase |
| Convergent | Qualitative and quantitative are conducted at the same time |
| Other | |

17

| **STATUS OF PUBLICATION** |
|---|
| *Whether or not the study has been finished or not* |
| Completed |
| Ongoing |
| Terminated |

18

| **WHO CLASSIFICATION** |
|---|
| *The main research subject is classified in one of these 10 areas.* |
| Virus natural history, transmission and diagnostics |

| |
|---|
| Animal and environmental research on the virus origin, and management measures at the human-animal interface |
| Epidemiological studies |
| Clinical management |
| Infection prevention and control, including health care workers' protection |
| Candidate therapeutics R&d |
| Candidate vaccines R&D |
| Ethics considerations for research |
| Social sciences in the outbreak response |
| Health systems arrangements |
| Other, including mental health |

| **RESEARCH QUESTION** | |
|---|---|
| *Type of research question. Knowledge that we are looking from the main intervention/exposure/phenomenon of interest* | |
| Benefits and harms of an intervention | Effectiveness or efficacy of an intervention |
| Costs and value for money | Budget impacts, economic evaluations, cost estimations,etc. |
| Magnitude of the problem | Measuring how big is a problem/issue (e.g., the impact of pandemic) |

| | |
|---|---|
| Views and preferences | Values and preferences of the population on a specific topic |
| How and why it works | Usually questions related to explore the mechanism of action of an intervention |
| Implementation barriers and facilitators | |
| Feasibility | How feasible is to implement an intervention |
| Diagnostic accuracy | |
| Prognosis | Study analyzing what factors determine the prospect of a health condition in the future (usually clinical) (e.g., does smoking increase the risk of becoming a critical COVID-19 patient?) |
| To describe a population | Prevalence, incidence on a specific population |
| Other | |

| **TYPES OF FUNDING** | |
|---|---|
| *Type of organization that is funding the research.* ***We might need to google the organization that is reported to be funding*** | |
| Government | or government agencies |
| International organizations | e.g., WHO, PAHO, etc. |
| Universities or research | |

| | |
|---|---|
| centres | |
| NGOs | |
| Private companies | e.g., pharmaceutical companies |
| Other | |

#### 3. Data on the number and rate of publications per country

| Country | Frequency | Population | Rate (publications per 1,000,000 population) | Rate excluding small countries with less than 1000000 population | Rate of research per FTE researchers*1000 |
|---|---|---|---|---|---|
| Antigua and Barbuda | 12 | 97,929 | 122.54 | | |
| Argentina | 168 | 45,195,774 | 3.72 | 3.72 | 3.26 |
| Bahamas | 12 | 393,244 | 30.52 | | |
| Barbados | 11 | 287,375 | 38.28 | | |
| Belize | 15 | 397,628 | 37.72 | | |
| Bolivia | 52 | 11,673,021 | 4.45 | 4.45 | 31.59 |
| Brazil | 885 | 212,559,417 | 4.16 | 4.16 | 6.38 |
| Chile | 162 | 19,116,201 | 8.47 | 8.47 | 23.83 |
| Colombia | 181 | 50,882,891 | 3.56 | 3.56 | 23.5 |
| Costa Rica | 41 | 5,094,118 | 8.05 | 8.05 | 6.71 |
| Cuba | 70 | 11,326,616 | 6.18 | 6.18 | |
| Dominica | 9 | 71,986 | 125.02 | | |
| Dominican | 41 | 10,847,910 | 3.78 | 3.78 | |

| | | | | | |
|---|---|---|---|---|---|
| Republic | | | | | |
| Ecuador | 106 | 17,643,054 | 6.01 | 6.01 | 38.74 |
| El Salvador | 35 | 6,486,205 | 5.40 | 5.4 | |
| Grenada | 9 | 112,523 | 79.98 | | |
| Guatemala | 39 | 17,915,568 | 2.18 | 2.18 | 94.89 |
| Guyana | 11 | 786,552 | 13.99 | | |
| Haiti | 16 | 11,402,528 | 1.40 | 1.4 | |
| Honduras | 35 | 9,904,607 | 3.53 | 3.53 | |
| Jamaica | 17 | 2,961,167 | 5.74 | 5.74 | |
| Mexico | 310 | 128,932,753 | 2.40 | 2.4 | 6.72 |
| Nicaragua | 26 | 6,624,554 | 3.92 | 3.92 | |
| Panama | 42 | 4,314,767 | 9.73 | 9.73 | 95.89 |
| Paraguay | 47 | 7,132,538 | 6.59 | 6.59 | 43.48 |
| Peru | 167 | 32,971,854 | 5.06 | 5.06 | |
| Saint Kitts and Nevis | 8 | 53,199 | 150.38 | | |
| Saint Lucia | 8 | 183,627 | 43.57 | | |
| Saint Vincent | 7 | 110,940 | 63.10 | | |

| | | | | | |
|---|---|---|---|---|---|
| and the Grenadines | | | | | |
| Suriname | 10 | 586,632 | 17.05 | | |
| Trinidad and Tobago | 13 | 1,399,488 | 9.29 | 9.29 | |
| Uruguay | 52 | 3,473,730 | 14.97 | 14.97 | 28.84 |
| Venezuela, Bolivarian Republic of | 50 | 28,435,940 | 1.76 | 1.76 | 5.76 |

#### 4. Included articles


1. No Authors. Willingness to pay for a COVID-19 vaccine high in Chile. PharmacoEconomics outcomes news. 2020;858: 35. doi:10.1007/s40274-020-7008-x
2. No Authors. What is the potential for a second peak in the evolution of SARS-CoV-2 in Brazil? Insights from a SIRASD model considering the informal economy. 2020.
3. No Authors. What is the people posting about symptoms related to Coronavirus in Bogota, Colombia? 2020.
4. No Authors. Update of the Evolution of SARS-CoV-2 Infection, COVID-19, and Mortality in Mexico Until May 15, 2020: An Ecological Study. Int J Trop Dis Heal. 2020. doi:10.9734/IJTDH/2020/V41I530277
5. No Authors. The First Few Hundred Cases for Coronavirus Disease 2019 (COVID-19) in Colombia. Iran J Public Health. 2020;49. doi:10.18502/IJPH.V49IS1.3669
6. No Authors. Predicting COVID-19 distribution in Mexico through a discrete and time-dependent Markov chain and an SIR-like model. 2020.
7. No Authors. Poverty levels, societal and individual heterogeneities explain the SARS-CoV-2 pandemic growth in Latin America. 2020.
8. No Authors. PADRÕES ESPAÇOS-TEMPORAIS DA ROTA DO COVID19 (CORONAVIRUS) NO ESTADO DE SANTA CATARINA, BRASIL. Hygeia Rev Bras Geogr Médica e da Saúde. 2020. doi:10.14393/HYGEIA0054251
9. No Authors. Modeling the transmission of new coronavirus in São Paulo State, Brazil -- Assessing epidemiological impacts of isolating young and elder persons. 2020.
10. No Authors. Modeling the early evolution of the COVID-19 in Brazil: results from a Susceptible-Infectious-Quarantined-Recovered (SIQR) model. 2020.
11. No Authors. How relevant is the decision of containment measures against COVID-19 applied ahead of time? 2020.
12. No Authors. Different scenarios in the Dynamics of SARS-Cov-2 Infection: an adapted ODE model. 2020.
13. No Authors. Data analysis and modeling of the evolution of COVID-19 in Brazil. 2020.
14. No Authors. COVID-19: estadística de morbi-mortalidad en Venezuela. 2020;5.
15. No Authors. COVID-19: A model for studying the evolution of contamination in Brazil. 2020.
16. No Authors. Clinical Management Strategies in Mild COVID-19 cases in Latin-America: A Decision Model. 2020.
17. No Authors. Cenário atual da covid-19 no estado do Ceará, Brasil. 2020;3. doi:10.31005/IAJMH.V3I0.92
18. No Authors. An Optimal Predictive Control Strategy for COVID-19 (SARS-CoV-2) Social Distancing Policies in Brazil. 2020.
19. No Authors. A utilização do planejamento territorial no combate da COVID-19: considerações sobre a situação dos leitos nos municípios de Pernambuco, Brasil. 2020;8. doi:10.22239/2317-269X.01546
20. Abrahao JS, Sacchetto L, Rezende IM, Rodrigues RAL, Crispim APC, Moura C, et al. Detection of SARS-CoV-2 RNA on public surfaces in a densely populated urban area of Brazil: A potential tool for monitoring the circulation of infected patients. Sci Total Environ. 2020; 142645. doi:http://dx.doi.org/10.1016/j.scitotenv.2020.142645

Jonatas Santos A, Livia Sacchetto P, Izabela Mauricio R, Rodrigo R, Ana Paula Correia C, Cesar M, et al. Detection of SARS-CoV-2 RNA on public surfaces in a densely populated urban area of Brazil. medRxiv. 2020. doi:10.1101/2020.05.07.20094631

21. Acosta G, Escobar G, Bernaola G, Alfaro J, Taype W, Marcos C, et al. Description of patients with severe COVID-19 treated in a national referral hospital in Peru. Rev peru med exp salud publica. 2020;37: 253–258. doi:10.17843/RPMESP.2020.372.5437
22. Acosta LD. [Response capacity to the COVID-19 pandemic in Latin America and the Caribbean]. Rev Panam Salud Publica. 2020;44: e109. doi:10.26633/RPSP.2020.109
23. Acosta LD. Capacidad de respuesta frente a la pandemia de COVID-19 en América Latina y el Caribe. 2020.
24. Acuna-Zegarra MA, Santana-Cibrian M, Velasco-Hernandez JX. Modeling behavioral change and COVID-19 containment in Mexico: A trade-off between lockdown and compliance. Math Biosci. 2020;325: 108370. doi:https://dx.doi.org/10.1016/j.mbs.2020.108370
25. Adan S-M, Jose Roberto Balmori de la M, Lauren H-V. Families under Confinement: COVID-19, Domestic Violence, and Alcohol Consumption. SSRN. 2020. doi:10.2139/ssrn.3688384
26. Adan S-M, Lauren H-V, Jose Roberto Balmori de la M, Abel R. COVID-19 and Women's Health: Examining Changes in Mental Health and Fertility. SSRN. 2020. doi:10.2139/ssrn.3656596
27. Adrian S-M, Braulio AMG, Erick Martinez R, Jose Omar Barreto R, Alicia Estela Lopez R, Paolo Alberti M, et al. THE LOW-HARM SCORE FOR PREDICTING MORTALITY IN PATIENTS DIAGNOSED WITH COVID-19: A MULTICENTRIC VALIDATION STUDY. medRxiv. 2020. doi:10.1101/2020.05.26.20111120
28. Agenor De Jr. N, Bernando Araldi da S, Felipe D-P, Luismar Marques P. A two-wave epidemiological model of COVID-19 outbreaks using MS-Excel(R). medRxiv. 2020. doi:10.1101/2020.05.08.20095133
29. Agostinho Antônio Cruz A, Jackeline Vieira A, Juliana do Nascimento S, Maria Clara Santos F, Camila de Meneses Caetano V, Pedro Henrique Moraes M, et al. COVID-19: ANALYSIS OF CONFIRMED CASES IN TERESINA, PIAUI, BRAZIL. 2020;6. doi:10.26694/REPIS.V6I0.10569
30. Aguiar S, Baiocchi G, Duprat JP, Coimbra FJF, Makdissi FB, Vartanian JG, et al. Value of preoperative testing for SARS-CoV-2 for elective surgeries in a cancer center during the peak of pandemic in Brazil. J Surg Oncol. 2020. doi:10.1002/jso.26146
31. Ahmed Sameer El K. Aspectos Psicocomportamentais durante a Pandemia da COVID-19: Uma análise dos efeitos provocados em moradores da região central de São Paulo. (Psychobehavioral Aspects during the COVID-19 Pandemic: An Analysis of the Effects Caused in Residents of the Cen. SSRN. 2020. doi:10.2139/ssrn.3612785
32. Ahumada PSM. Social isolation and quarantine effects over the subjective well-being of Chileans during the COVID-19 outbreak. 2020.
33. Akira O, Yusuke N, Tokuro F. Universality in COVID-19 spread in view of the Gompertz function. medRxiv. 2020. doi:10.1101/2020.06.18.20135210
34. Alberto F, Matther S-K. Factors linked to changes in mental health outcomes among Brazilians in quarantine due to COVID-19. medRxiv. 2020. doi:10.1101/2020.05.12.20099374
35. Alberto F, Matthew S-K. The Relationship Between Behavioural and Psychosocial Factors Among Brazilians in Quarantine Due to COVID-19. SSRN. 2020. doi:10.2139/ssrn.3566245
36. Alberto P-M, Marina M, Carolina F, Sergio G, Angelica R, Lisseth P, et al. SARS-CoV-2 spread across the Colombian-Venezuelan border. medRxiv. 2020. doi:10.1101/2020.07.09.20149856
37. Albitres-Flores L, Pisfil-Farroñay YA, Guillen-Macedo K, Niño-Garcia R, Alarcon-Ruiz CA. Perception of medical interns about the internship during the COVID-19 pandemic in Peru. 2020.
38. Alcibiades V, Giselle R, Xu Z, Digna W, Carolina De La G, Gabrielle B, et al. Performance of a point of care test for detecting IgM and IgG antibodies against SARS-CoV-2 and seroprevalence in blood donors and health care workers in Panama. medRxiv. 2020. doi:10.1101/2020.09.25.20201459
39. Aldo C, Marcel G, Eduardo L, Marcelo O, Gabriel YW, Julio C, et al. The Social Divide of Social Distancing: Lockdowns in Santiago during the COVID-19 Pandemic. SSRN. 2020. doi:10.2139/ssrn.3691373

40. Alejandra C-M, Carlos MG-L, Mercedes A, Ana Cristina S, Hector L-F. Municipality- level predictors of COVID-19 mortality in Mexico: a cautionary tale. medRxiv. 2020. doi:10.1101/2020.07.11.20151522
41. Alejandro C-M, Reinaldo Andres R-S. CCOFEE-GI Study: Colombian COVID19 First Experience in Gastroentrology. Characterization of digestive manifestations in patients diagnosed with COVID-19 at a highly complex institution in Bogota D.C., Colombia. medRxiv. 2020. doi:10.1101/2020.07.24.20161604
42. Alejandro L-F, David H, Fernanda M-P. Air Pollution Exposure and COVID-19: A Look at Mortality in Mexico City Using Individual-Level Data. SSRN. 2020. doi:10.2139/ssrn.3673616
43. Alejandro M-S, Carlos AF-M, Neftali Eduardo A-V, Arsenio V-V, Enrique CG, Alejandro C-M, et al. Adaptive responses to SARS-CoV-2 infection linked to accelerated aging measures predict adverse outcomes in patients with severe COVID-19. medRxiv. 2020. doi:10.1101/2020.11.03.20225375
44. Alejandro M, Alejandra S, Adriana A, Carolina G, Yazmin R, Javier G, et al. Socio-demographic and Clinical Characteristics of Adults with SARS-CoV-2 Infection in Two Hospitals in Bogota, Colombia. medRxiv. 2020. doi:10.1101/2020.08.13.20167445
45. Alejandro R, Alfredo M, Gabriela BS, Patricia MG, Juan ES, Jose MC. A numerical simulation of the COVID-19 epidemic in Argentina using the SEIR model. arXiv Popul Evol. 2020.
46. Alejandro T, María P, Felipe V, Luis Ruso M, Gustavo Rodríguez T, Mauro P, et al. COVID 19 en fase 2 sostenida. Experiencia quirúrgica inicial en el Hospital Maciel. 2020;4. doi:10.31837/CIR.URUG.4.2.6
47. Alessandro CP, Paula de Castro P, Daiane FDL, Alexandre VS, Marco R, Cezar VWR, et al. COVID-19 Seroprevalence in Military Police Forces, Southern Brazil. SSRN. 2020. doi:10.2139/ssrn.3697174
48. Alessi J, De Oliveira GB, Franco DW, Brino Do Amaral B, Becker AS, Knijnik CP, et al. Mental health in the era of COVID-19: Prevalence of psychiatric disorders in a cohort of patients with type 1 and type 2 diabetes during the social distancing. Diabetol Metab Syndr. 2020;12: 76. doi:http://dx.doi.org/10.1186/s13098-020-00584-6
49. Alger J, Cafferata ML, Alvarado T, Ciganda A, Corrales A, Desale H, et al. Using Prenatal Blood Samples to Evaluate COVID-19 Rapid Serologic Tests Specificity. Matern Child Health J. 2020;24: 1099–1103. doi:10.1007/s10995-020-02981-9
50. Aliaga-Castillo V, Fuentes Vejar B, Muñoz Campos J. Descriptive report of self-reported cases of physiotherapists infected by COVID-19 in Chile. Kinesiologia. 2020;39: 8–13.
51. Almeida ALC, Santo T, Mello MSS, Cedro A V, Lopes NL, Ribeiro A, et al. Repercussions of the COVID-19 Pandemic on the Care Practices of a Tertiary Hospital. Arq Bras Cardiol. 2020. doi:10.36660/abc.20200436
52. Almeida J dos S, Cardoso JA, Cordeiro EC, Lemos M, Araújo TME de, Sardinha AH de L. CARACTERIZAÇÃO EPIDEMIOLÓGICA DOS CASOS DE COVID-19 NO MARANHÃO: Uma breve análise. 2020.
53. Almeida S, Villibor C, Carstensen S, Petterle R. Proposal and psychometric validation of the Severe Acute Respiratory Syndrome - Coronavirus-2 Fear Scale (SCoV-2-FS). ResearchSquare. 2020. doi:10.21203/rs.3.rs-48227/v1
54. Almeida W da S de, Szwarcwald CL, Malta DC, Barros MB de A, Souza Júnior PRB de, Azevedo LO, et al. Changes of Brazilians' socioeconomic and health conditions during the COVID-19 pandemic. 2020.
55. Alomo M, Gagliardi G, Peloche S, Somers E, Alzina P, Prokopez CR. [Psychological effects during the COVID-19 outbreak in Argentina]. Efectos Psicol la pandemia COVID-19 en la Poblac Gen Argentina. 2020;77: 176–181. doi:https://dx.doi.org/10.31053/1853.0605.v77.n3.28561
56. Altair Souza De A, Vinicius JOW de C. Logistic Approach to COVID - 19 Epidemic Evolution in Brazil. medRxiv. 2020. doi:10.1101/2020.06.22.20135921
57. Alvarez-Diaz DA, Franco-Munoz C, Laiton-Donato K, Usme-Ciro JA, Franco-Sierra ND, Florez-Sanchez AC, et al. Molecular analysis of several in-house rRT-PCR protocols for SARS-CoV-2 detection in the context of genetic variability of the virus in Colombia. Infect Genet Evol. 2020;84: 104390. doi:https://dx.doi.org/10.1016/j.meegid.2020.104390

58. Alvarez-Lopez DI, Espinoza-Molina MP, Cruz-Loustaunau ID, Alvarez-Hernandez G. Diabetes and hypertension as factors associated with Covid-19 lethality in Sonora, Mexico, 2020. Salud Publica Mex. 2020;62: 456–457. doi:http://dx.doi.org/10.21149/11546
59. Alvaro Francisco Lopes de S, Layze Braz O, Guilherme S, Artur Acelino Francisco Luz Q, Herica Emilia Felix de C, Telma Maria Evangelista de A, et al. Casual sex among MSM during the period of social isolation in the COVID-19 pandemic: Nationwide study in Brazil and Portugal. medRxiv. 2020. doi:10.1101/2020.06.07.20113142
60. Alvaro Q-A, Oscar E, Marcela MM-R, Diana W, Diana Carolina M. Analysis of the interventions adopted due to the COVID-19 on ARI morbility for Colombia. medRxiv. 2020. doi:10.1101/2020.09.12.20193334
61. Alves JG, Ferreira Lima TP. International air traffic and COVID-19 geographical incidence in Brazil. J Microbiol Immunol Infect. 2020. doi:http://dx.doi.org/10.1016/j.jmii.2020.09.009
62. Alves THE, Souza TA de, Silva S de A, Ramos NA, Oliveira SV de. Analysis of home and hospital deaths from respiratory and cardiovascular causes during the COVID-19 pandemic in Minas Gerais. 2020.
63. Amador-Jiménez M, Millner N, Palmer C, Pennington RT, Sileci L. The Unintended Impact of Colombia's Covid-19 Lockdown on Forest Fires. Environ Resour Econ. 2020; 1–25. doi:10.1007/s10640-020-00501-5
64. Amaral-Prado HM, Borghi F, Mello T, Grassi-Kassisse DM. The impact of confinement in the psychosocial behaviour due COVID-19 among members of a Brazilian university. Int J Soc Psychiatry. 2020; 20764020971318. doi:10.1177/0020764020971318
65. Amna T, Eduardo AU, Carla Castillo L, Katia V-G, Ruiyan L, Richard R, et al. Early transmission dynamics of COVID-19 in Chile: From sub-exponential ascending growth dynamics to a stationary disease wave, March-April, 2020. medRxiv. 2020. doi:10.1101/2020.05.15.20103069
66. Amorim Filho L, Szwarcwald CL, Mateos SOG, Leon A, Medronho RA, Veloso VG, et al. Seroprevalence of anti-SARS-CoV-2 among blood donors in Rio de Janeiro, Brazil. Rev Saude Publica. 2020;54: 69. doi:10.11606/s1518-8787.2020054002643

Amorim Filho L, Szwarcwald CL, Mateos S de OG, Leon ACMP de, Medronho R de A, Veloso VG, et al. Seroprevalence of IgG and IgM anti-SARS-CoV-2 among voluntary blood donors in Rio de Janeiro, Brazil. 2020. p. 19.
67. Amrita R, Katherine R, Brooke J, Benjamin A, Sara W, Julia M, et al. Potential interruptions in HIV prevention and treatment services for gay, bisexual, and other men who have sex with men associated with COVID-19. medRxiv. 2020. doi:10.1101/2020.08.19.20178285
68. Ana Jessica P, Diego R, Sofia Mendes S, Kamila M, Karina B, Ana CMR, et al. Increased prolonged sitting in rheumatoid arthritis patients during the COVID-19 pandemic: a within-subjects, accelerometer-based study. medRxiv. 2020. doi:10.1101/2020.09.09.20191395
69. Ana Karolina Antunes E, Meriane D, Juliana Schons G, Alana Witt H, Karoline S, Larissa M, et al. Comparison Of Different Kits For SARS-CoV-2 RNA Extraction Marketed In Brazil. bioRxiv. 2020. doi:10.1101/2020.05.29.122358
70. Ana Maria Baptista M, Cesar GV, Fernando PH, Mariangela FS, Bernardo LH, Aluisio JDB, et al. High prevalence of symptoms among Brazilian subjects with antibodies against SARS-CoV-2: a nationwide household survey. medRxiv. 2020. doi:10.1101/2020.08.10.20171942
71. Ana O-G, Erwin C, Jesus A-D, Carlos G-C, Adrián G-Á, Raúl R-R, et al. Sex Differences in the Outcome of Patients with Confirmed SARS-CoV-2 Infection. SSRN. 2020. doi:10.2139/ssrn.3624222
72. Anacleto MA, Brito FA, de Queiroz AR, Passos E, Santos JRL. Diffusive process under Lifshitz scaling and pandemic scenarios. Physica A. 2020;559: 125092. doi:10.1016/j.physa.2020.125092
73. Analía R, Andrea Elvia María B, Pilar BB, Camila Soledad D, Melisa Adriana L, Martina P, et al. Epidemiological characteristics of the first 116 974 cases of COVID-19 in Argentina, 2020. Rev argent salud publica. 2020;12: 1–9.

74. Anderson Castro Soares de O, Lia Hanna Martins M, Eveliny Barroso da S, Daniele Cristina Tita G, Luiz Andre Ribeiro Z, Cor Jesus Fernandes F. Bayesian modeling of COVID-19 cases with a correction to account for under-reported cases. medRxiv. 2020. doi:10.1101/2020.05.24.20112029
75. Anderson Walter Costa S. Epidemiologic profile and social determinant of COVID-19 in Macapá, Amapá, Amazon, Brazil. 2020;04. doi:10.32749/NUCLEODOCONHECIMENTO.COM.BR/HEALTH/COVID-19-IN-MACAPA
76. Andrade EF, Pereira LJ, Oliveira APL, Orlando DR, Alves DAG, Guilarducci JS, et al. Perceived fear of COVID-19 infection according to sex, age and occupational risk using the Brazilian version of the Fear of COVID-19 Scale. Death Stud. 2020; 1–10. doi:http://dx.doi.org/10.1080/07481187.2020.1809786
77. Andrade JV, Moraes RCC. What has the Coronavirus take from us? Potential years of life lost in Minas Gerais. J nurs Heal. 2020;10: 20104014.
78. Andrade LA, Gomes DS, Goes MAO, Souza MSF, Teixeira DCP, Ribeiro CJN, et al. Surveillance of the first cases of COVID-19 in Sergipe using a prospective spatiotemporal analysis: the spatial dispersion and its public health implications. Rev Soc Bras Med Trop. 2020;53: e20200287. doi:http://dx.doi.org/10.1590/0037-8682-0287-2020
79. Andre Filipe de Moraes B, Joao Luiz M, Thiago Henrique Rizzi D, Alexandre Dias Porto Chiavegatto F. COVID-19 diagnosis prediction in emergency care patients: a machine learning approach. medRxiv. 2020. doi:10.1101/2020.04.04.20052092
80. Andre Z, Andréa Burmeister Morais H, Bernardo F, Claudionor Oliveira Gomes J, Cristian Rogério F, Frederico Golbspan S, et al. How Do Networking Capabilities Support SMEs in Surpassing the COVID-19 Crisis? Lessons Learned from Small Firms in a Developing Country. SSRN. 2020. doi:10.2139/ssrn.3669248
81. Andreia Faranha da C, Alan Patricio da S, Luana Dias de V, Marjorie Heloise M, Andreia Zarzour Abou Hala C. O acesso vertiginoso na aquisição de máscaras faciais e suas repercussões na saúde pública e de populações vulneráveis/The vertiginous access in the acquisition of face masks and their repercussions on pubic health and vulnerable populations. 2020;4.
82. Andreia C de M, Luiz Claudio ST, Jesse L da S, Lucas Z de A, Ana Carla P, Luciana ORR, et al. Cancer inpatient with COVID-19: a report from the Brazilian National Cancer Institute. medRxiv. 2020. doi:10.1101/2020.06.27.20141499

De Melo AC, Thuler LCS, Da Silva JL, De Albuquerque LZ, Pecego AC, Rodrigues LDOR, et al. Cancer inpatients with COVID-19: A report from the Brazilian National Cancer Institute. PLoS One. 2020;15: e0241261. doi:http://dx.doi.org/10.1371/journal.pone.0241261
83. Andres IV-O, Juliana Villanueva C, Silvana Zapata B, Zulma MC. Impact of contact tracing on COVID-19 mortality: An impact evaluation using surveillance data from Colombia. medRxiv. 2020. doi:10.1101/2020.08.14.20158535
84. Andres M, Pieter S, Alethea D, Vilma R, Maria Jose Torres H, Alicia L. Impact of COVID-19 on Maternal Mental Health in a Fragile and Conflict-Affected Setting: A Longitudinal Case-Control Study in Tumaco, Colombia. SSRN. 2020. doi:10.2139/ssrn.3706751
85. Andrés N, Gastón V-H. On the dynamics of the Coronavirus epidemic and the unreported cases: the Chilean case. arXiv Popul Evol. 2020.
86. Angel Jose P-C, Marc C, Christian G-C, Guido C-E, José R-S, Carmelo D, et al. Social interventions can lower COVID-19 deaths in middle-income countries. medRxiv. 2020. doi:10.1101/2020.04.16.20063727
87. Angel Santillan H. Caracterización epidemiológica de covid-19 en Ecuador. 2020;3. doi:10.31005/IAJMH.V3I0.99
88. Ángel GCP, Eric A-V, Ugo Avila-Ponce de L. A data driven analysis and forecast of an SEIARD epidemic model for COVID-19 in Mexico. arXiv Popul Evol. 2020.
89. Angela Araújo M, Amanda Stefani Torquato da S, Regiane Lima Gasques P. Implantação do protocolo de manejo de corpos pós-óbito no contexto do novo Coronavírus. 2020;10.

90. Anibal AT, Gabriel C, Ronald Prado de la G, Carolina H, Giovanny H, Luis MP, et al. Epidemiological characterization of asymptomatic carriers of COVID-19 in Colombia. medRxiv. 2020. doi:10.1101/2020.06.18.20134734
91. Annaelise Fritz M, Bruno Barbosa S, Teresa D, Miguel Barbosa R, Francisca Braga C. Empreendedorismo Social, Inovação e Benchmarking no Instagram para combater os efeitos negativos do COVID-19 numa visão luso-brasileira. 2020;6.
92. Antônio Augusto Moura da S, Lídio Gonçalves Lima N, Conceição de Maria Pedrozo e Silva de A, Léa Márcia Melo da C, Maylla Luana Barbosa Martins B, Allan Kardec Duailibe Barros F, et al. Population-based seroprevalence of SARS-CoV-2 is more than halfway through the herd immunity threshold in the State of Maranhao, Brazil. medRxiv. 2020. doi:10.1101/2020.08.28.20180463
93. Antonio da Silva S, Antonio Jr. P. Estimation and monitoring of COVID-19 transmissibility from publicly available data. medRxiv. 2020. doi:10.1101/2020.05.24.20112128
94. Antonio CCG, Karla Santa Cruz C, Kathleen Tereza da C, Bárbara Soares de Oliveira S, Janimayri Forastieri de A, Gustavo Fialho C, et al. COMPARATIVE ANALYSIS OF COVID-19 MORTALITY IN BRAZIL, RIO DE JANEIRO, CAMPOS DOS GOYTACAZES, MACAE, CABO FRIO AND RIO DAS OSTRAS. medRxiv. 2020. doi:10.1101/2020.09.17.20196444
95. Antunes BB de P, Peres IT, Baiao FA, Ranzani OT, Bastos LDSL, Silva A de AB da, et al. Progression of confirmed COVID-19 cases after the implementation of control measures. Rev Bras Ter intensiva. 2020. doi:https://dx.doi.org/10.5935/0103-507x.20200028

Igor Tona P, Fernanda Araújo B, Otavio Tavares R, Leonardo Dos Santos Lourenço B, Amanda de Araújo Batista da S, Guilherme Faveret Garcia de S, et al. Progressão dos casos confirmados de COVID-19 após implantação de medidas de controle./ Progressão dos casos confirmados de COVID-19 após implantação de medidas de controle./ Progression of confirmed COVID-19 cases after the implementation of control measu. Rev Bras Ter intensiva. 2020.
96. Antunez-Montes OY, Escamilla MI, Figueroa-Uribe AF, Arteaga-Menchaca E, Lavariega-Sarachaga M, Salcedo-Lozada P, et al. COVID-19 and Multisystem Inflammatory Syndrome in Latin American Children: A Multinational Study. Pediatr Infect Dis J. 2020. doi:http://dx.doi.org/10.1097/INF.0000000000002949
97. Aranda-Abreu GE, Aranda-Martinez JD, Araujo R, Hernandez-Aguilar ME, Herrera-Covarrubias D, Rojas-Duran F. Observational study of people infected with SARS-Cov-2, treated with amantadine. Pharmacol Reports. 2020. doi:http://dx.doi.org/10.1007/s43440-020-00168-1
98. Arantes VN, Martins BC, Seqatto R, Milhomen-Cardoso DM, Franzini TP, Zuccaro AM, et al. Impact of coronavirus pandemic crisis in endoscopic clinical practice: Results from a national survey in Brazil. Endosc Int open. 2020;8: E822–E829. doi:https://dx.doi.org/10.1055/a-1183-3324
99. Araújo Neto RA de, Melo GC de. Correlation between weather, population size and COVID-19 pandemic: a study of Brazilian capitals. J Heal Biol Sci. 2020;8: 1–5.
100. Araujo DB, Machado RRG, Amgarten DE, Malta FM, de Araujo GG, Monteiro CO, et al. SARS-CoV-2 isolation from the first reported patients in Brazil and establishment of a coordinated task network. Mem Inst Oswaldo Cruz. 2020;115: e200342. doi:http://dx.doi.org/10.1590/0074-02760200342
101. Araujo EM, Lilly Caldwell K, Pereira Alves dos Santos M, Magalhães de Souza I, Lima Ferreira Santa Rosa P, Beatriz Silva dos Santos A, et al. COVID-19 morbimortality by race/skin color/ethnicity: the experience of Brazil and the United States. 2020.
102. Araujo OR, Almeida CG, Lima-Setta F, Prata-Barbosa A, Colleti Junior J. The Impact of the Novel Coronavirus on Brazilian PICUs. Pediatr Crit Care Med. 2020. doi:http://dx.doi.org/10.1097/PCC.0000000000002583
103. Ardila-Sierra A, Niño-Leal L, Rivera-Triana D, Sarmiento-Medina MI, Alzate JP. Underlying conditions on the southern border between Colombia and Venezuela to face the COVID-19 pandemic. Rev salud pública. 2020;22: e486366–e486366.

104. Arellano-Llamas AA, HernÁNdez-Caballero Ál. COVID-19 en niños y adolescentes mexicanos hasta el 10 de mayo de 2020. Enfoque en pacientes con diabetes. Rev Mex Endocrinol Metab y Nutr. 2020;7: 80–86. doi:10.24875/RME.20000055
105. Arias Velásquez RM, Mejía Lara J V. Gaussian approach for probability and correlation between the number of COVID-19 cases and the air pollution in Lima. Urban Clim. 2020;33: 100664. doi:10.1016/j.uclim.2020.100664
106. Arias-Reyes C, Zubieta-DeUrioste N, Poma-Machicao L, Aliaga-Raudan F, Carvajal-Rodriguez F, Dutschmann M, et al. Does the pathogenesis of SAR-CoV-2 virus decrease at high-altitude? Respir Physiol Neurobiol. 2020;277: 103443. doi:10.1016/j.resp.2020.103443
107. Arizpe-Bravo D, Arizpe-Bravo A, Leon RB, Chacon-Cano R, Arizpe-Vilana D, Anthon F, et al. EXPERIENCE WITH COVID-19 CRITICALLY ILL PATIENTS IN A PRIVATE COMMUNITY HOSPITAL IN PUEBLA CITY, MEXICO. Chest. 2020;158: A2482–A2483. doi:http://dx.doi.org/10.1016/j.chest.2020.09.057
108. Armando da R. Regression Polynomial Analysis of the COVID-19 Epidemics: An Alternative Infection Modeling. SSRN. 2020. doi:10.2139/ssrn.3682535
109. Arregoces HA, Rojano R, Restrepo G. Impact of lockdown on particulate matter concentrations in Colombia during the COVID-19 pandemic. Sci Total Environ. 2020; 142874. doi:http://dx.doi.org/10.1016/j.scitotenv.2020.142874
110. Arturo Sr. C-V, Berenice Z-B, Jesus Carlos B-G, Guadalupe Mercedes Lucia G-A, Lourdes A-P, Eduardo F-O, et al. Serological Cytokine and chemokine profile in pregnant women with COVID19 in Mexico City. medRxiv. 2020. doi:10.1101/2020.07.14.20153585
111. Asanov I, Flores F, McKenzie D, Mensmann M, Schulte M. Remote-learning, time-use, and mental health of Ecuadorian high-school students during the COVID-19 quarantine. World Dev. 2021;138: 105225. doi:10.1016/j.worlddev.2020.105225
112. Astigueta-Perez J, Abad-Licham M, Chavez-Chirinos C, Beraun-Milla L, Lachos-Davila A, Diaz-Perez E, et al. Cancer disease progression and death during the COVID-19 pandemic: A multidisciplinary analysis for the Peruvian setting. Ecancermedicalscience. 2020;14: 1098. doi:http://dx.doi.org/10.3332/ECANCER.2020.1098
113. Atamari-Anahui N, Cruz-Nina ND, Condori-Huaraka M, Nunez-Paucar H, Rondon-Abuhadba EA, Ordonez-Linares ME, et al. [Characterization of coronavirus disease 2019 (COVID-19) in children and adolescents in Latin American and the Caribbean countries: A descriptive study]. Caracter la Enferm por coronavirus 2019 en ninos y Adolesc en paises Am Lat y El Caribe Estud Descr. 2020;20: e8025. doi:https://dx.doi.org/10.5867/medwave.2020.08.8025
114. Augusto Sr. SF, Leonardo Sousa Sr. S, Silvio Sr. SLS. New S.I.R. model used in the projection of COVID 19 cases in Brazil. medRxiv. 2020. doi:10.1101/2020.04.26.20080218
115. Auler AC, Cassaro FAM, da Silva VO, Pires LF. Evidence that high temperatures and intermediate relative humidity might favor the spread of COVID-19 in tropical climate: A case study for the most affected Brazilian cities. Sci Total Environ. 2020;729: 139090. doi:https://dx.doi.org/10.1016/j.scitotenv.2020.139090
116. Autrán-Gómez AM, Tobia I, Molina RC, Covarrubias FR, Benzing F, Serena Maruccia F, et al. Exploring Urological Experience in the COVID-19 Outbreak: American Confederation of Urology (CAU) Survey. Int Braz J Urol. 2020;46. doi:10.1590/S1677-5538.IBJU.2020.S119
117. Avid R-G, Natalia IV-C. Variation of Aerosol Pollution in Peru during the Quarantine Due to COVID-19. Int J Adv Comput Sci Appl. 2020;11. doi:10.14569/IJACSA.2020.0110407
118. Avila-Ponce de León U, Pérez Á GC, Avila-Vales E. An SEIARD epidemic model for COVID-19 in Mexico: Mathematical analysis and state-level forecast. Chaos Solitons Fractals. 2020;140: 110165. doi:10.1016/j.chaos.2020.110165
119. Badellino H, Gobbo ME, Torres E, Aschieri ME. Early indicators and risk factors associated with mental health problems during COVID-19 quarantine: Is there a relationship with the number of confirmed cases

and deaths? Int J Soc Psychiatry. 2020; 20764020966020. doi:http://dx.doi.org/10.1177/0020764020966020

120. Ballivian J, Alcaide ML, Cecchini D, Jones DL, Abbamonte JM, Cassetti I. Impact of COVID-19-Related Stress and Lockdown on Mental Health Among People Living With HIV in Argentina. J Acquir Immune Defic Syndr. 2020;85: 475–482. doi:http://dx.doi.org/10.1097/QAI.0000000000002493

121. Banerjee A, Ray S, Vorselaars B, Kitson J, Mamalakis M, Weeks S, et al. Use of Machine Learning and Artificial Intelligence to predict SARS-CoV-2 infection from Full Blood Counts in a population. Int Immunopharmacol. 2020;86: 106705. doi:10.1016/j.intimp.2020.106705

122. Baptista AS, Prado IM, Perazzo MF, Pinho TM, Paiva SM, Pordeus IA, et al. Can children's oral hygiene and sleep routines be compromised during the COVID-19 pandemic? Int J Paediatr Dent. 2020. doi:http://dx.doi.org/10.1111/ipd.12732

123. Baqui P, Bica I, Marra V, Ercole A, van der Schaar M. Ethnic and regional variations in hospital mortality from COVID-19 in Brazil: a cross-sectional observational study. Lancet Glob Heal. 2020;8: e1018–e1026. doi:10.1016/S2214-109X(20)30285-0

Pedro B, Ioana B, Valerio M, Ari E, Mihaela van der S. Ethnic and Regional Variation in Hospital Mortality from COVID-19 in Brazil. SSRN. 2020. doi:10.2139/ssrn.3606429

Gustavo Barcelos B, Ticiane Henriques Santa R, Pedro Goes M, Rafael Henriques J, Lidia Freire Abdalla N. Analytical sensibility and specificity of two RT-qPCR protocols for SARS-CoV-2 detection performed in an automated workflow. medRxiv. 2020. doi:10.1101/2020.03.07.20032326

129. Barraza NR, Pena G, Moreno V. A non-homogeneous Markov early epidemic growth dynamics model. Application to the SARS-CoV-2 pandemic. Chaos Solitons Fractals. 2020;139: 110297. doi:10.1016/j.chaos.2020.110297

130. Barros AJD, Victora CG, Menezes AMB, Horta BL, Hartwig F, Victora G, et al. Social distancing patterns in nine municipalities of Rio Grande do Sul, Brazil: the Epicovid19/RS study. Rev Saude Publica. 2020;54: 75. doi:10.11606/s1518-8787.2020054002810

131. Barros MB de A, Lima MG, Malta DC, Szwarcwald CL, Azevedo RCS de, Romero D, et al. Report on sadness/depression, nervousness/anxiety and sleep problems in the Brazilian adult population during the COVID-19 pandemic. Relato tristeza/depressao, Nerv e Probl sono na Popul adulta Bras durante a pandemia COVID-19. 2020;29: e2020427. doi:https://dx.doi.org/10.1590/s1679-49742020000400018

132. Bastidas JAO, Pita APP, Vargas LNO, Montenegro AF. Importance of nonlinear regression models in the interpretation of data from COVID-19 in Colombia. Rev Habanera Ciencias Medicas. 2020;19: e_3309.

Omar Yaxmehen B-C, Neftali Eduardo A-V, Arsenio V-V, Carlos AF-M, Alejandro M-S, Jessica Paola B-L. Profiling pre-symptomatic and asymptomatic cases with confirmed SARS-CoV-2 infection in Mexico City. medRxiv. 2020. doi:10.1101/2020.07.02.20145516

145. Bello-Chavolla OY, Bahena-Lopez JP, Antonio-Villa NE, Vargas-Vazquez A, Gonzalez-Diaz A, Marquez-Salinas A, et al. Predicting mortality due to SARS-CoV-2: A mechanistic score relating obesity and diabetes to COVID-19 outcomes in Mexico. J Clin Endocrinol Metab. 2020. doi:https://dx.doi.org/10.1210/clinem/dgaa346

Omar Yaxmehen B-C, Jessica Paola B-L, Neftali EA-V, Arsenio V-V, Armando G-D, Alejandro M-S, et al. Predicting mortality attributable to SARS-CoV-2: A mechanistic score relating obesity and diabetes to COVID-19 outcomes in Mexico. medRxiv. 2020. doi:10.1101/2020.04.20.20072223

146. Bello-Chavolla OY, Gonzalez-Diaz A, Antonio-Villa NE, Fermin-Martinez CA, Marquez-Salinas A, Vargas-Vazquez A, et al. Unequal impact of structural health determinants and comorbidity on COVID-19

severity and lethality in older Mexican adults: Considerations beyond chronological aging. J Gerontol A Biol Sci Med Sci. 2020. doi:https://dx.doi.org/10.1093/gerona/glaa163

Omar Yaxmehen B-C, Armando G-D, Neftali EA-V, Carlos AF-M, Alejandro M-S, Arsenio V-V, et al. Unequal impact of structural health determinants and comorbidity on COVID-19 severity and lethality in older Mexican adults: Looking beyond chronological aging. medRxiv. 2020. doi:10.1101/2020.05.12.20098699

Carlos Ernesto B-A, Oscar Mauricio E-S. Clinical and Epidemiological Analysis of COVID-19 Children Cases in Colombia PEDIACOVID. SSRN. 2020. doi:10.2139/ssrn.3678580

159. Borba MGS, Val FFA, Sampaio VS, Alexandre MAA, Melo GC, Brito M, et al. Effect of High vs Low Doses of Chloroquine Diphosphate as Adjunctive Therapy for Patients Hospitalized With Severe Acute Respiratory Syndrome Coronavirus 2 (SARS-CoV-2) Infection: A Randomized Clinical Trial. JAMA Netw open. 2020;3: e208857. doi:https://dx.doi.org/10.1001/jamanetworkopen.2020.8857

Mayla B, Fernando de Almeida V, Vanderson Sousa S, Marcia Araujo A, Gisely Cardoso M, Marcelo B, et al. Chloroquine diphosphate in two different dosages as adjunctive therapy of hospitalized patients with severe respiratory syndrome in the context of coronavirus (SARS-CoV-2) infection: Preliminary safety results of a randomized, double-blinded, phase IIb cl. medRxiv. 2020. doi:10.1101/2020.04.07.20056424

160. Borba PL de O, Bassi BG de C, Pereira BP, Vasters GP, Correia RL, Barreiro RG. "Practical and reflective" challenges for occupational therapy's undergraduation courses in pandemic times. 2020.
161. Borges GM, Crespo CD. Demographic and socioeconomic characteristics of Brazilian adults and COVID-19: a risk group analysis based on the Brazilian National Health Survey, 2013. Cad Saude Publica. 2020;36: e00141020. doi:http://dx.doi.org/10.1590/0102-311X00141020
162. Borges LL, Guimaraes CC V, Aguiar BGC, Felipe LAF. Military Nursing in "Operation Return to Brazil": aeromedical evacuation in the coronavirus pandemic. Rev Bras Enferm. 2020;73 2: e20200297. doi:http://dx.doi.org/10.1590/0034-7167-2020-0297
163. Borja-Villanueva CA, Gómez-Carrión CE, Alvarado-Muñoz ER, Bernuy-Torres LA. Knowledge about coronavirus disease (COvId-19) in dentists of Lima and Callao. Rev cient odontol. 2020;8: e019–e019.
164. Borracci RA, Giglio ND. Forecasting the effect of social distancing on COVID-19 autumn-winter outbreak in the metropolitan area of Buenos Aires. Estim del Ef del distanciamiento Soc sobre la epidemia COVID-19 otono-invierno en el area Metrop Buenos Aires. 2020;80 Suppl 3: 7–15.
165. Bottan N, Hoffmann B, Vera-Cossio D. The unequal impact of the coronavirus pandemic: Evidence from seventeen developing countries. PLoS One. 2020;15: e0239797. doi:http://dx.doi.org/10.1371/journal.pone.0239797
166. Bozovich GE, Alves De Lima A, Fosco M, Burgos LM, Martínez R, Dupuy De Lôme R, et al. [Collateral damage of COVID-19 pandemic in private healthcare centres of Argentina]. Medicina (B Aires). 2020;80 Suppl 3: 37–41.
167. Braga JU, Ramos Jr. AN, Ferreira AF, Lacerda VM, Freire RMC, Bertoncini BV. Propensity for COVID-19 severe epidemic among the populations of the neighborhoods of Fortaleza, Brazil, in 2020. BMC Public Health. 2020;20: 1486. doi:https://dx.doi.org/10.1186/s12889-020-09558-9
168. Brandao Neto D, Fornazieri MA, Dib C, Di Francesco RC, Doty RL, Voegels RL, et al. Chemosensory Dysfunction in COVID-19: Prevalences, Recovery Rates, and Clinical Associations on a Large Brazilian Sample. Otolaryngol - Head Neck Surg (United States). 2020. doi:http://dx.doi.org/10.1177/0194599820954825
169. Brinkley R, Elvis WD, Julianna S, Edith Z, Ynes M, Claudia M, et al. The impact of the COVID-19 pandemic on rabies reemergence in Latin America: the case of Arequipa, Peru. medRxiv. 2020. doi:10.1101/2020.08.06.20169581
170. Brito LGO, Romao GS, Fernandes CE, Silva-Filho AL. Impact of COVID-19 on Brazilian medical residencies in obstetrics and gynecology. Int J Gynaecol Obstet. 2020. doi:https://dx.doi.org/10.1002/ijgo.13283
171. Bruna Caruso M, Fabiana Infante S, Gabriel Perri E, Heloisa CSA, Marcia Thereza C, Milla Cordeiro A, et al. Influence of nutritional status on eating habits and food choice determinants among Brazilian women during the COVID-19 pandemic. medRxiv. 2020. doi:10.1101/2020.11.03.20225136
172. Bruno Campello de S, Fernando Menezes Campello de S. Does Social Isolation Really Curb COVID-19 Deaths? Direct Evidence from Brazil that it Might do the Exact Opposite. SSRN. 2020. doi:10.2139/ssrn.3706464
173. Bruno Campello de S, Fernando Menezes Campello de S. Physical Distancing and Future COVID-19 Deaths in Brazil: Evidence of a Paradoxical Effect. SSRN. 2020. doi:10.2139/ssrn.3711686
174. Bryan Adrian G-P, Arturo T-R, Samanta Mayanini P-G, Cristina D-R, Omar S-N, Marisol Manriquez R, et al. Anxiety, depression, attitudes, and internet addiction during the initial phase of the 2019 coronavirus disease (COVID-19) epidemic: A cross-sectional study in Mexico. medRxiv. 2020. doi:10.1101/2020.05.10.20095844

Askery C, Elayne T, Renato M, Lais M, Samurai B, Rafael C. Data-Driven Study of the the COVID-19 Pandemic via Age-Structured Modelling and Prediction of the Health System Failure in Brazil amid Diverse Intervention Strategies. medRxiv. 2020. doi:10.1101/2020.04.03.20052498

Routes for COVID-19 importation in Brazil ;Journal of Travel Medicine ;Oxford Academic. 2020.

206. Carlos Sr. AR, Airandes Sr. SP, Carlito Sr. LS, Edval Jr. GS, Livia Sr. AC, Paulo Sr. CN, et al. Covid-19 epidemic curve in Brazil: A sum of multiple epidemics, whose income inequality and population

density in the states are correlated with growth rate and daily acceleration. medRxiv. 2020. doi:10.1101/2020.09.09.20191353
207. Carlos Augusto Cardoso Sr. P, Estefano Aparecido Sr. V, Jose Andre Sr. L, Jefferson Oliveira do Sr. N. Characteristics and evolution of COVID-19 cases in Brazil: mathematical modeling and simulation. medRxiv. 2020. doi:10.1101/2020.10.14.20212829
208. Carlos Eduardo R, Marcella Cini O, Tatiana de Araujo E, Suzana Rosa A, Marcele Gonçalves da S, Eny Regina da Silva Q, et al. Spatial Analysis of COVID-19 Incidence and the Sociodemographic Context in Brazil. SSRN. 2020. doi:10.2139/ssrn.3709861
209. Carlos Enrique Bustamante O, Jordy Jose Cevallos C, Cesar M, Jeff S, Edwin M, Anuj M. Modeling and Preparedness: The Transmission Dynamics of COVID-19 Outbreak in Provinces of Ecuador. medRxiv. 2020. doi:10.1101/2020.07.09.20150078
210. Carlos Jesús A-A, Julissa C-U, Luis H, Frank Z-C, Cender UQ-J. Infodemiological study of COVID-19 in Latin America and The Caribbean. medRxiv. 2020. doi:10.1101/2020.08.11.20173054
211. Carlos Magno Castelo Branco F, Raul Borges G, Rafael de Castro C, Claudia Pio F, Gabriel Berg de A, Edmur P. Elementary spatial structures and dispersion of COVID-19: health geography directing responses to public health emergency in Sao Paulo State, Brazil. medRxiv. 2020. doi:10.1101/2020.04.26.20080895
212. Carlos von Krakauer Sr. H, Marcella Lima B, Rafaella Dourado L. Distress among Brazilian university students due to the Covid-19 pandemic: survey results and reflections. medRxiv. 2020. doi:10.1101/2020.06.19.20135251
213. Carlos F-M, Diego Alejandro A-D, Katherine L-D, Magdalena W, Patricia E, Jose AU-C, et al. Substitutions in Spike and Nucleocapsid proteins of SARS-CoV-2 circulating in Colombia. medRxiv. 2020. doi:10.1101/2020.06.02.20120782
214. Carlos IM. Inhomogeneous mixing and asynchronic transmission between local outbreaks account for the spread of COVID-19 epidemics. medRxiv. 2020. doi:10.1101/2020.08.04.20168443
215. Carlos MH-S. Statistics associated with the lethality of COVID-19 by age group and gender in Mexico. medRxiv. 2020. doi:10.1101/2020.06.28.20142117
216. Carlos MH-S, Efren M-Z. Using COVID-19 deaths as a surrogate to measure the progression of the pandemics. medRxiv. 2020. doi:10.1101/2020.09.27.20202564
217. Carlos MH-S, Paolo V, Efren M-Z. On the estimation of the total number of SARS-CoV-2 infections. medRxiv. 2020. doi:10.1101/2020.04.23.20077446
218. Carlos M-R, Hilario M-L. Data Mining for the Study of the Epidemic (SARS- CoV-2) COVID-19: Algorithm for the Identification of Patients (SARS-CoV-2) COVID 19 in Mexico. SSRN. 2020. doi:10.2139/ssrn.3619549
219. Carlos M-R, Hilario M-L. Impact of (SARS-CoV-2) COVID-19 on the Five Main Indigenous Language-Speaking Areas in Veracruz Mexico: The Case of the Totonacapan Area. SSRN. 2020. doi:10.2139/ssrn.3672636
220. Carlos-Maria A, Sergio S, Gabriela N, Raul R-V, Carolina G-S. Job insecurity, financial threat and mental health in the COVID-19 context: The buffer role of perceived social support. medRxiv. 2020. doi:10.1101/2020.07.31.20165910
221. Carmen Cecilia C-D, Jeimmy De L-S, Adalberto C-A. Social capital and psychological distress during Colombian coronavirus disease lockdown. medRxiv. 2020. doi:10.1101/2020.09.04.20187914
222. Carmo RF, Nunes BEBR, Machado MF, Armstrong AC, Souza CDF. Expansion of COVID-19 within Brazil: the importance of highways. J Travel Med. 2020. doi:https://dx.doi.org/10.1093/jtm/taaa106
223. Carolina P, Marcos Antonio Ferreira J, Andreia Insabralde de Queiroz C, Luciana Scarlazzari C, Mercy da Costa S, Felipe Machado M, et al. Temporal analysis of the clinical evolution of confirmed cases of COVID-19 in the state of Mato Grosso do Sul - Brazil. medRxiv. 2020. doi:10.1101/2020.09.21.20198812

224. Carrascosa MMC, Campos T de, Sampaio JE, Souza RRF, Ribeiro VL, Maia MLN, et al. Medical Interns and COVID-19: results of national research. Rev Assoc Med Bras. 2020;66: 812–817. doi:https://dx.doi.org/10.1590/1806-9282.66.6.812
225. Carriel J, Muñoz-Jaramillo R, Bolaños-Ladinez O, Heredia-Villacreses F, Menéndez-Sanchón J, Martin-Delgado J, et al. [CURB-65 as a predictor of 30-day mortality in patients hospitalized with COVID-19 in Ecuador: COVID-EC StudyAbstract]. Rev Clin Esp. 2020. doi:10.1016/j.rce.2020.10.001
226. Carrillo-Vega MF, Salinas-Escudero G, Garcia-Pena C, Gutierrez-Robledo LM, Parra-Rodriguez L. Early estimation of the risk factors for hospitalization and mortality by COVID-19 in Mexico. PLoS One. 2020;15: e0238905. doi:https://dx.doi.org/10.1371/journal.pone.0238905

Maria Fernanda C-V, Guillermo S-E, Carmen G-P, Luis Miguel G-R, Lorena P-R. Early estimation of the risk factors for hospitalisation and mortality by COVID-19 in Mexico. medRxiv. 2020. doi:10.1101/2020.05.11.20098145
227. Carvalho Malta D, Saar Gomes C, Landmann Szwarcwald C, Berti de Azevedo Barros M, Gomes da Silva A, Junio Sady Prates E, et al. Social distancing, feeling of sadness and lifestyles of the Brazilian population during the COVID-19 pandemic. 2020.
228. Carvalho EC, Souza PHD de O, Varella TCMYML, Souza NVD de O, Farias SNP de, Soares SSS. COVID-19 pandemic and the judicialization of health care: an explanatory case study. Rev Lat Am Enfermagem. 2020;28: e3354. doi:https://dx.doi.org/10.1590/1518-8345.4584.3354
229. Carvalho HEF, Schneider G, Sousa AR, Camargo ELS, Nunes R V, Possani MA, et al. Suspected COVID-19 flu-like syndrome in men who have sex with men and have been involved in casual sex. Rev Bras Enferm. 2020;73: e20200913. doi:http://dx.doi.org/10.1590/0034-7167-2020-0913
230. Carvalho L de F, Pianowski G, Goncalves AP. Personality differences and COVID-19: are extroversion and conscientiousness personality traits associated with engagement with containment measures? Trends psychiatry Psychother. 2020. doi:https://dx.doi.org/10.1590/2237-6089-2020-0029
231. Cassiani-Miranda CA, Campo-Arias A, Tirado-Otálvaro AF, Botero-Tobón LA, Upegui-Arango LD, Rodríguez-Verdugo MS, et al. Stigmatisation associated with COVID-19 in the general Colombian population. Int J Soc Psychiatry. 2020; 20764020972445. doi:10.1177/0020764020972445
232. Castillo AE, Parra B, Tapia P, Acevedo A, Lagos J, Andrade W, et al. Phylogenetic analysis of the first four SARS-CoV-2 cases in Chile. J Med Virol. 2020. doi:https://dx.doi.org/10.1002/jmv.25797
233. Castro R, Luz PM, Wakimoto MD, Veloso VG, Grinsztejn B, Perazzo H. COVID-19: a meta-analysis of diagnostic test accuracy of commercial assays registered in Brazil. Braz J Infect Dis. 2020;24: 180–187. doi:https://dx.doi.org/10.1016/j.bjid.2020.04.003
234. Castro-Paris R, Munoz-Velandia O, Garcia-Pena A, Fernández-Ávila D. Assessment of the Academic Satisfaction of Graduate Students During the SARS-CoV-2 Pandemic, In An Internal Medicine Department. ResearchSquare. 2020. doi:10.21203/rs.3.rs-69949/v1
235. Castro-Rodriguez Y, Valenzuela-Torres O. Implications of COVID-19 pandemic for dental care: A perspective for clinical dentists. Rev Habanera Ciencias Medicas. 2020;19: A58.
236. Catarina Vezetiv Jr. M, Aluizio M, Vitoria Ferreira C, Maria do Carmo Pinho F, Elke S, Luciana Yuki T. High prevalence of food insecurity, the adverse impact of COVID-19 in Brazilian favela. medRxiv. 2020. doi:10.1101/2020.07.31.20166157
237. Cavalcante FP, Novita GG, Millen EC, Zerwes FP, de Oliveira VM, Sousa ALL, et al. Management of early breast cancer during the COVID-19 pandemic in Brazil. Breast Cancer Res Treat. 2020. doi:10.1007/s10549-020-05877-y
238. Cavalcante JR, Cardoso-Dos-Santos AC, Bremm JM, Lobo AP, Macário EM, Oliveira WK, et al. COVID-19 in Brazil: evolution of the epidemic up until epidemiological week 20 of 2020. Epidemiol e Serv saude Rev do Sist Unico Saude do Bras. 2020;29: e2020376. doi:10.5123/s1679-49742020000400010

239. Cavalcante JR, Abreu A de JL de. COVID-19 in the city of Rio de Janeiro: spatial analysis of first confirmed cases and deaths. Epidemiol e Serv saude Rev do Sist Unico Saude do Bras. 2020;29: e2020204. doi:https://dx.doi.org/10.5123/S1679-49742020000300007

Cavalcante JR, Abreu A de JL de. COVID-19 in Rio de Janeiro municipality: spatial distribution of the first deaths and cases confirmed. 2020.

240. Cavalcanti AB, Zampieri FG, Rosa RG, Azevedo LCP, Veiga VC, Avezum A, et al. Hydroxychloroquine with or without Azithromycin in Mild-to-Moderate Covid-19. N Engl J Med. 2020. doi:10.1056/NEJMoa2019014

Alexandre BC, Fernando GZ, Luciani CPA, Regis GR, Alvaro A, Viviane CV, et al. Hydroxychloroquine alone or in combination with azithromycin to prevent major clinical events in hospitalised patients with coronavirus infection (COVID-19): rationale and design of a randomised, controlled clinical trial. medRxiv. 2020. doi:10.1101/2020.05.19.20106997

253. Chate RC, Fonseca EKUN, Passos RBD, Teles GB da S, Shoji H, Szarf G. Presentation of pulmonary infection on CT in COVID-19: initial experience in Brazil. J Bras Pneumol. 2020;46: e20200121. doi:https://dx.doi.org/10.36416/1806-3756/e20200121
254. Chavarri-Guerra Y, Ramos-Lopez WA, Covarrubias-Gomez A, Sanchez-Roman S, Quiroz-Friedman P, Alcocer-Castillejos N, et al. Providing Supportive and Palliative Care Using Telemedicine for Patients with Advanced Cancer During the COVID-19 Pandemic in Mexico. Oncologist. 2020. doi:http://dx.doi.org/10.1002/onco.13568
255. Chavin DG, Dale V, Michelle T, Davlin T. The Epidemiology Characteristics of Positive COVID-19 patients in a Caribbean Territory. medRxiv. 2020. doi:10.1101/2020.08.06.20148288
256. Chen X, Zhang SX, Jahanshahi AA, Alvarez-Risco A, Dai H, Li J, et al. Belief in a COVID-19 Conspiracy Theory as a Predictor of Mental Health and Well-Being of Health Care Workers in Ecuador: Cross-Sectional Survey Study. JMIR public Heal Surveill. 2020;6: e20737. doi:https://dx.doi.org/10.2196/20737
257. Chen YT, Yen YF, Yu SH, Su EC. An Examination on the Transmission of COVID-19 and the Effect of Response Strategies: A Comparative Analysis. Int J Environ Res Public Health. 2020;17: 1–14. doi:10.3390/ijerph17165687
258. Chiesa-Estomba CM, Lechien JR, Portillo-Mazal P, Martinez F, Cuauro-Sanchez J, Calvo-Henriquez C, et al. Olfactory and gustatory dysfunctions in COVID-19. First reports of Latin-American ethnic patients. Am J Otolaryngol. 2020;41: 102605. doi:https://dx.doi.org/10.1016/j.amjoto.2020.102605
259. Chioro A, Calife K, Santos Barros CR dos, Martins LC, Calvo M, Estanislau E, et al. COVID-19 at a metropolitan region: public polices and social vulnerability within an iniquity context. 2020.
260. Chisini LA, Sartori LRM, Costa FDS, Salvi LC, Demarco FF. COVID-19 pandemic impact on prosthetic treatments in the Brazilian Public Health System. Oral Dis. 2020. doi:http://dx.doi.org/10.1111/odi.13668
261. Christian Omar R-P, Erika Areli R-G, Cristina Elizabeth M-M, Monica Patricia B-R, Rafael García R, Carolina B-D, et al. Laboratory Parameters and Complication Risk Factors in Mexican COVID-19 Patients: A Cross-Sectional Study. SSRN. 2020. doi:10.2139/ssrn.3634850
262. Christian A-R, Favio C-R, Liliana P-M, Fernanda A-R, Danuzia AM, Natalia Zubieta D, et al. Decreased incidence, virus transmission capacity, and severity of COVID-19 at altitude on the American continent. medRxiv. 2020. doi:10.1101/2020.07.22.20160168
263. Christian C-C, Medalit L-V, Julio M-A, Blanca S-M, Diana P-A, Ramiro P-M, et al. Clinical-epidemiological and treatment characteristics of children with COVID-19 in a tertiary referral center in Peru. medRxiv. 2020. doi:10.1101/2020.09.18.20186866
264. Christian SP. Analysis of the SARS-CoV-2 outbreak in Rio Grande do Sul / Brazil. arXiv Popul Evol. 2020.
265. Christielly Mendonca B, Marco Tulio Pacheco C, Jose Alexandre Felizola D-F, Thiago Fernando R. The necessary cooperation between governments and public in the fight against COVID-19: why non-pharmaceutical interventions may be ineffective. medRxiv. 2020. doi:10.1101/2020.08.17.20176347
266. Chu AMY, Tsang JTY, Chan JNL, Tiwari A, So MKP. Analysis of travel restrictions for COVID-19 control in Latin America through network connectedness. J Travel Med. 2020. doi:http://dx.doi.org/10.1093/jtm/taaa176
267. Ciampi E, Uribe-San-Martín R, Soler B, Fernández R, García P, Navarrete-Asenjo C, et al. COVID-19 in MS and NMOSD: A multicentric online national survey in Chile. Mult Scler Relat Disord. 2020;45: 102392. doi:10.1016/j.msard.2020.102392
268. Ciampi E, Uribe-San-Martin R, Carcamo C. COVID-19 pandemic: The experience of a multiple sclerosis centre in Chile. Mult Scler Relat Disord. 2020;42: 102204. doi:https://dx.doi.org/10.1016/j.msard.2020.102204
269. Cintra HPC, Fontinele FN. Estimative of real number of infections by COVID-19 in Brazil and possible scenarios. Infect Dis Model. 2020;5: 720–736. doi:http://dx.doi.org/10.1016/j.idm.2020.09.004

Pedro Henrique Pinheiro C, Felipe Fontinele N. Estimative of real number of infections by COVID-19 on Brazil and possible scenarios. medRxiv. 2020. doi:10.1101/2020.05.03.20052779
270. Civantos AM, Bertelli A, Goncalves A, Getzen E, Chang C, Long Q, et al. Mental health among head and neck surgeons in Brazil during the COVID-19 pandemic: A national study. Am J Otolaryngol - Head Neck Med Surg. 2020;41: 102694. doi:http://dx.doi.org/10.1016/j.amjoto.2020.102694
271. Clarice Ribeiro Alves C, Francisco das Chagas Bezerra N, Eliezio Nascimento B, Adryele Gomes M. Impactos da pandemia da covid-19 sob a égide do código de defesa do consumidor. Res Soc Dev. 2020;9. doi:10.33448/RSD-V9I6.3578
272. Clarisse Lins de L, Cecilia Cordeiro da S, Ana Clara Gomes da S, Eduardo Luiz S, Gabriel Souza M, Lucas Job Brito de A, et al. COVID-SGIS: A smart tool for dynamic monitoring and temporal forecasting of Covid-19. medRxiv. 2020. doi:10.1101/2020.05.30.20117945
273. Claudia R-R, Beatriz Piedad U. Modelling strategies to predict hospital demand during the COVID-19 outbreak in Bogota, Colombia. medRxiv. 2020. doi:10.1101/2020.04.14.20065466
274. Cobre AF, Boger B, Fachi MM, Vilhena RO, Domingos EL, Tonin FS, et al. Risk factors associated with delay in diagnosis and mortality in patients with COVID-19 in the city of Rio de Janeiro, Brazil. Cien Saude Colet. 2020;25: 4131–4140. doi:http://dx.doi.org/10.1590/1413-812320202510.2.26882020
275. Coelho FC, Lana RM, Cruz OG, Villela DAM, Bastos LS, Pastore Y Piontti A, et al. Assessing the spread of COVID-19 in Brazil: Mobility, morbidity and social vulnerability. PLoS One. 2020;15: e0238214. doi:https://dx.doi.org/10.1371/journal.pone.0238214
276. Cohen G, Russo MJ, Campos JA, Allegri RF. COVID-19 Epidemic in Argentina: Worsening of Behavioral Symptoms in Elderly Subjects With Dementia Living in the Community. Front Psychiatry. 2020;11: 866. doi:http://dx.doi.org/10.3389/fpsyt.2020.00866
277. Cohen G, Russo MJ, Campos JA, Allegri RF. Living with dementia: increased level of caregiver stress in times of COVID-19. Int psychogeriatrics. 2020; 1–11. doi:10.1017/S1041610220001593
278. Colunga-Salas P, Hernandez-Canchola G. Bats and humans during the SARS-CoV-2 outbreak: The case of bat-coronaviruses from Mexico. Transbound Emerg Dis. 2020. doi:http://dx.doi.org/10.1111/tbed.13751
279. Connerton P, Vicente de Assunção J, Maura de Miranda R, Dorothée Slovic A, José Pérez-Martínez P, Ribeiro H. Air Quality during COVID-19 in Four Megacities: Lessons and Challenges for Public Health. Int J Environ Res Public Health. 2020;17: 1–24. doi:10.3390/ijerph17145067
280. Conselho Nacional de Secretários de S. Segunda fase da pesquisa "Evolução da Prevalência de Infecção por Covid-19 no Brasil: Estudo de Base Populacional&quot. 2020.
281. Conte D, Bahia L, Laurentino de Carvalho E, Monte Cardoso A, Souza PM. Public and private supply of beds and access to health care in the COVID-19 pandemic in Brazil. 2020.
282. Contreras S, Biron-Lattes JP, Villavicencio HA, Medina-Ortiz D, Llanovarced-Kawles N, Olivera-Nappa Á. Statistically-based methodology for revealing real contagion trends and correcting delay-induced errors in the assessment of COVID-19 pandemic. Chaos Solitons Fractals. 2020;139: 110087. doi:10.1016/j.chaos.2020.110087
283. Contreras-Rodríguez L, Sandoval-Hernández A, Maya-Hoyos M, Soto C. Remote learning of biochemistry during the COVID-19 pandemic: case of undergraduate students in Bogota, Colombia. ResearchSquare. 2020. doi:10.21203/rs.3.rs-72902/v1
284. Cordova-Lepe F, Gutierrez-Aguilar R, Gutierrez-Jara JP. Number of COVID-19 cases in Chile at 120 days with data at 21/03/2020 and threshold of daily effort to flatten the epi-curve. Medwave. 2020;20: e7861. doi:https://dx.doi.org/10.5867/medwave.2020.02.7861
285. Cornejo N, Candia P. Descripción de la aplicación de screening universal para SARS-CoV-2 en mujeres embarazadas que ingresan para interrupción en Hospital de Carabineros – Santiago de Chile. Rev Chil Obstet Ginecol. 2020;85: S2–S8.
286. Corpus-Mendoza AN, Ruiz-Segoviano HS, Rodriguez-Contreras SF, Yanez-Davila D, Hernandez-Granados A. Decrease of mobility, electricity demand, and NO2 emissions on COVID-19 times and their

feedback on prevention measures. Sci Total Environ. 2020; 143382. doi:http://dx.doi.org/10.1016/j.scitotenv.2020.143382
287. Corrêa PRL, Ishitani LH, Abreu DMX, Teixeira RA, Marinho F, França EB. The importance of surveillance in cases of and mortality from the COVID-19 epidemic in Belo Horizonte, Brazil, 2020. Rev Bras Epidemiol. 2020;23: e200061. doi:10.1590/1980-549720200061
288. Corrêa PRL, Ishitani LH, Abreu DMX de, Teixeira RA, Marinho F, França EB. The importance of surveillance in cases and mortality from COVID-19 epidemic in Belo Horizonte, 2020. 2020.
289. Correa-Araneda F, Ulloa-Yañez A, Núñez D, Boyero L, Tonin A, Cornejo A, et al. Environmental determinants of COVID-19 transmission across a wide climatic gradient in Chile. ResearchSquare. 2020. doi:10.21203/rs.3.rs-30393/v1
290. Correa-Cuadros JP, Muñoz-Rodríguez MA. SARS-C0V.2/COVID-I9 in Colombia: tendencies, predictions, and tensions about health system. Rev salud pública. 2020;22: e386614–e386614.
291. Correa-Cuadros JP, Muñoz-Rodríguez MA. SARS-COV2/COVID-19 en Colombia: tendencias, predicciones y tensiones sobre el sistema sanitario. Rev salud pública. 2020;22: e386614–e386614.
292. Cortés JA, Espitia P, Rosero-Lasso YL. Citywide preparedness for a pandemic: A crosssectional survey of knowledge, attitudes, and practices about respiratory infection prevention in Bogotá, Colombia. Biomedica. 2020;40: 159–165. doi:10.7705/biomedica.5526
293. Cortes-Alvarez NY, Pineiro-Lamas R, Vuelvas-Olmos CR. Psychological effects and associated factors of COVID-19 in a Mexican sample. Disaster Med Public Health Prep. 2020; 1–27. doi:https://dx.doi.org/10.1017/dmp.2020.215
294. Cortés-Tellés A, López-Romero S, Mancilla-Ceballos R, Ortíz-Farías DL, Núñez-Caamal N, Figueroa-Hurtado E. Risk factors for mortality among hospitalized patients with COVID-19. An overview in Mexican population. Tuberc Respir Dis (Seoul). 2020. doi:10.4046/trd.2020.0095
295. Costa AN, de Sa ERA, Bezerra RDS, Souza JL, Lima F das CA. Constituents of buriti oil (Mauritia flexuosa L.) like inhibitors of the SARS-Coronavirus main peptidase: an investigation by docking and molecular dynamics. J Biomol Struct Dyn. 2020; 1–8. doi:https://dx.doi.org/10.1080/07391102.2020.1778538
296. Costa CLA, Costa TM, Barbosa Filho VC, Bandeira PFR, Siqueira RCL. Influence of social distancing on the physical activity level during the COVID-19 pandemic. Rev bras ativ fís saúde. 2020;25: 1–6.
297. Costa IPA, Maêda S, Teixeira L, Gomes CFS, Santos MD. Choosing a hospital assistance ship to fight the covid-19 pandemic. Rev Saude Publica. 2020;54: 79. doi:10.11606/s1518-8787.2020054002792
298. Costa MF. Health belief model for coronavirus infection risk determinants. Rev Saude Publica. 2020;54: 47. doi:https://dx.doi.org/10.11606/s1518-8787.2020054002494
299. Costa MA, Lui L, Santos RM dos, Curi RLC, Albuquerque CG de, Tavares SR, et al. Apontamentos sobre a dimensão territorial da pandemia da Covid-19 e os fatores que contribuem para aumentar a vulnerabilidade socioespacial nas unidades de desenvolvimento humano de áreas metropolitanas brasileiras. 2020. p. 59.
300. Costa SM, Lacerda GT de, Villafort RN, Silveira RL, Amaral MBF. What Do We Know About COVID-19?: Maxillofacial Surgeons Survey. J Craniofac Surg. 2020;31: e661–e663. doi:https://dx.doi.org/10.1097/SCS.0000000000006658
301. Costa SF, Buss L, Espinoza EPS, Vieira Jr. JM, de Oliveira da Silva LC, de Souza RM, et al. Performance of a qualitative rapid chromatographic immunoassay to diagnose COVID-19 in patients in a middle-income country. J Clin Virol. 2020;131: 104592. doi:https://dx.doi.org/10.1016/j.jcv.2020.104592
302. Cota G, Freire ML, de Souza CS, Pedras MJ, Saliba JW, Faria V, et al. Diagnostic performance of commercially available COVID-19 serology tests in Brazil. Int J Infect Dis. 2020. doi:http://dx.doi.org/10.1016/j.ijid.2020.10.008
303. Cota W. Monitoring the number of COVID-19 cases and deaths in Brazil at municipal and federative units level. 2020.

304. Cotrin P, Bahls AC, da Silva DO, Pereira Girao VM, Maio Pinzan-Vercelino CR, de Oliveira RCG, et al. The use of facemasks during the COVID-19 pandemic by the Brazilian population. J Multidiscip Healthc. 2020;13: 1169–1178. doi:http://dx.doi.org/10.2147/JMDH.S281524

305. Cotrin P, Moura W, Gambardela-Tkacz CM, Pelloso FC, Santos LD, Carvalho MDB, et al. Healthcare Workers in Brazil during the COVID-19 Pandemic: A Cross-Sectional Online Survey. Inquiry. 2020;57: 46958020963711. doi:http://dx.doi.org/10.1177/0046958020963711

306. Cotrin P, Peloso RM, Pini NIP, Oliveira RC, de Oliveira RCG, Valarelli FP, et al. Urgencies and emergencies in orthodontics during the coronavirus disease 2019 pandemic: Brazilian orthodontists' experience. Am J Orthod Dentofacial Orthop. 2020. doi:10.1016/j.ajodo.2020.06.028

307. Cotta RM, Naveira-Cotta CP, Magal P. Mathematical Parameters of the COVID-19 Epidemic in Brazil and Evaluation of the Impact of Different Public Health Measures. Biology (Basel). 2020;9. doi:10.3390/biology9080220

Renato Machado C, Carolina Palma N-C, pierre magal. Modelling the COVID-19 epidemics in Brasil: Parametric identification and public health measures influence. medRxiv. 2020. doi:10.1101/2020.03.31.20049130

Carlos Henrique de Brito C. Social distancing in são paulo state: Demonstrating the reduction in cases using time series analysis of deaths due to covid-19/ Distanciamento social no estado de são paulo: Uso de série temporal dos óbitos por causa da covid-19 para demonstrar a redução. Rev Bras Epidemiol. 2020.

318. Cruz-Pacheco G, Bustamante-Castaneda JF, Caputo JG, Jimenez-Corona ME, Ponce-de-Leon-Rosales S. DISPERSION OF A NEW CORONAVIRUS SARS-COV-2 BY AIRLINES IN 2020: TEMPORAL

ESTIMATES OF THE OUTBREAK IN MEXICO. Rev Invest Clin. 2020;72: 138–143. doi:https://dx.doi.org/10.24875/RIC.20000113
319. Cuartas DE, Arango-Londoño D, Guzmán-Escarria G, Muñoz E, Caicedo D, Ortega D, et al. SARS-coV-2 spatio - temporal analysis in Cali, Colombia. Rev salud pública. 2020;22: e286431–e286431.

Gissel G-E, Edgar M, Diana C, Delia O, Andrés F-L, Jorge M, et al. Análisis espacio-temporal del SARS-COV-2 en Cali, Colombia/ SARS-coV-2 spatio - temporal analysis in Cali, Colombia. 2020;22.
320. Cuenca-Jaque CR, Osorio-Tarrillo ML, Pastor-Ramos JL, G DELPP-P, Torres-Vasquez LE. Economic and health aspects in quarantine times per covid 19 in the peruvian population, year 2020. Rev la Fac Med Humana. 2020;20: 630–639. doi:http://dx.doi.org/10.25176/RFMH.V20I4.3067
321. Cuesta J, Pico J. The Gendered Poverty Effects of the COVID-19 Pandemic in Colombia. Eur J Dev Res. 2020; 1–34. doi:10.1057/s41287-020-00328-2
322. Cuevas-Barragan CE, Buenrostro-Nava MT, Palos-Gomez GM, Ramirez-Padilla EA, Mendoza-Macias BI, Rivas-Caceres RR. Use of Nasoil via intranasal to control the harmful effects of Covid-19. Microb Pathog. 2020;149: 104504. doi:http://dx.doi.org/10.1016/j.micpath.2020.104504
323. Da Cunha de Sa-Caputo D, Sonza A, Bachur JA, Bernardo-Filho M. Development, validation and reliabilty of a questionnaire to evaluate the changes on the level of physical exercises and in daily life habits due to COVID-19 pandemic social distancing. Acta Biomed. 2020;91: e2020004. doi:https://dx.doi.org/10.23750/abm.v91i3.9888
324. da Cunha AR, Antunes JLF, Martins MD, Petti S, Hugo FN. The impact of the COVID-19 pandemic on oral biopsies in the Brazilian National Health System. Oral Dis. 2020. doi:http://dx.doi.org/10.1111/odi.13620
325. da Silva Dittz E, Leão da Silveira Rocha A. The repercussions in daily routine of mothers of babies admitted in Neonatal Intensive Care Unit during the social isolation to avoid COVID-19. 2020.
326. da Silva RG, Ribeiro M, Mariani VC, Coelho LDS. Forecasting Brazilian and American COVID-19 cases based on artificial intelligence coupled with climatic exogenous variables. Chaos Solitons Fractals. 2020;139: 110027. doi:10.1016/j.chaos.2020.110027
327. Dal'Bosco EB, Floriano LSM, Skupien S V, Arcaro G, Martins AR, Anselmo ACC. Mental health of nursing in coping with COVID-19 at a regional university hospital. Rev Bras Enferm. 2020;73 2: e20200434. doi:http://dx.doi.org/10.1590/0034-7167-2020-0434
328. Dalton Garcia Borges de S, Francisco Tarcísio Alves J, Nei Yoshihiro S. Forecasting COVID-19 cases at the Amazon region: a comparison of classical and machine learning models. bioRxiv. 2020. doi:10.1101/2020.10.09.332908
329. Dan M-K, Bruno B-T, Mercedes M-G, Andrea Zaldivar-Perez P, Juan OT. U-shaped-aggressiveness of SARS-CoV-2: Period between onset of nonspecific-specific symptoms for COVID-19. A population-based cohort study. medRxiv. 2020. doi:10.1101/2020.10.28.20221697
330. Daniel AA, Andrea B. Emerging Mental Health Challenges, Strategies and Opportunities in the context of the COVID-19 Pandemic: Perspectives from South American Decision-makers. medRxiv. 2020. doi:10.1101/2020.07.16.20155630
331. Daniel CPJ, Moreno SR, Mateus SS, Luciana LC, Nivea B da S, Ismael HS, et al. Assessing the nationwide impact of COVID-19 mitigation policies on the transmission rate of SARS-CoV-2 in Brazil. medRxiv. 2020. doi:10.1101/2020.06.26.20140780
332. Daniel G-C, Daniel R-A, Marco B, Juan G, Daniel M, Narcisa G, et al. Adapting for the COVID-19 pandemic in Ecuador, a characterization of hospital strategies and patients. medRxiv. 2020. doi:10.1101/2020.07.25.20161661
333. Daniele Melo S, Karla VBL, Ana LSF, Juliana CDG, Thalyta MRLU, Yan Correa R, et al. Clinical characteristics of Severe Acute Respiratory Syndrome by COVID-19 in Indigenous of Brazil. medRxiv. 2020. doi:10.1101/2020.10.24.20218701

334. Danilo F, Claudia G, Leyda EA, Jean PC, Yamilka D, Yaset C, et al. Early transmission dynamics, spread, and genomic characterization of SARS-CoV-2 in Panama. medRxiv. 2020. doi:10.1101/2020.07.31.20160929
335. Dantas F. Outcomes research of homeopathic treatment in suspected or confirmed patients with COVID-19 IN Brazil: Clinical research protocol. 2020.

Dantas F. Resultados terapêuticos da homeopatia em pacientes suspeitos ou confirmados de COVID-19 no Brasil: Protocolo para estudo observacional prospectivo. 2020. p. 46.
336. Dantas G, Siciliano B, Franca BB, da Silva CM, Arbilla G. The impact of COVID-19 partial lockdown on the air quality of the city of Rio de Janeiro, Brazil. Sci Total Environ. 2020;729: 139085. doi:https://dx.doi.org/10.1016/j.scitotenv.2020.139085
337. Daon Y, Thompson RN, Obolski U. Estimating COVID-19 outbreak risk through air travel. J Travel Med. 2020. doi:https://dx.doi.org/10.1093/jtm/taaa093
338. David de Paulo F, Fábio Fernandes de A. Will COVID-19 affect food supply in distribution centers of Brazilian regions affected by the pandemic? Trends Food Sci Technol. 2020;103: 361–366. doi:10.1016/J.TIFS.2020.05.023
339. Dayrell A, Cristina de Souza Andrade A, Leandro Machado E, Lúcia Meireles A, Silva Magalhães A, do Prado Ribeiro E, et al. Through the COVID-19 magnifying glass: an examination of disparities in the Brazilian capitals. 2020.
340. de Alencar JCG, Moreira CL, Muller AD, Chaves CE, Fukuhara MA, da Silva EA, et al. Double-blind, randomized, placebo-controlled trial with N-acetylcysteine for treatment of severe acute respiratory syndrome caused by COVID-19. Clin Infect Dis. 2020. doi:http://dx.doi.org/10.1093/cid/ciaa1443
341. De Boni RB, Balanza-Martinez V, Mota JC, Cardoso TA, Ballester P, Carbonell B, et al. Depression, Anxiety and Lifestyle Among Essential Workers: A Websurvey From Brazil And Spain During The Covid-19 Pandemic. J Med Internet Res. 2020. doi:http://dx.doi.org/10.2196/22835
342. de Brito CAA, de Brito MCM, Fernandes Martins TH, de Brito CCM, Militao Albuquerque MF, de Cassia Coelho Moraes de Brito R. Clinical laboratory and dispersion pattern of COVID-19 in a family cluster in the social-distancing period. J Infect Dev Ctries. 2020;14: 987–993. doi:http://dx.doi.org/10.3855/JIDC.13580
343. de Farias ECF, Pedro Piva J, de Mello M, do Nascimento L, Costa CC, Machado MMM, et al. Multisystem Inflammatory Syndrome Associated With Coronavirus Disease in Children: A Multi-centered Study in Belém, Pará, Brazil. Pediatr Infect Dis J. 2020. doi:10.1097/INF.0000000000002865
344. De la Hoz-Restrepo F, Alvis-Zakzuk NJ, De la Hoz-Gomez JF, De la Hoz A, Gomez Del Corral L, Alvis-Guzman N. Is Colombia an example of successful containment of the 2020 COVID-19 pandemic? A critical analysis of the epidemiological data, March to July 2020. Int J Infect Dis. 2020;99: 522–529. doi:http://dx.doi.org/10.1016/j.ijid.2020.08.017
345. de la Miyar JRB, Hoehn-Velasco L, Silverio-Murillo A. Druglords Don’t stay at home: COVID-19 pandemic and crime patterns in Mexico City. J Crim Justice. 2020; 101745. doi:10.1016/j.jcrimjus.2020.101745
346. de Lima Filho BF, Bessa NPOS, Fernandes ACT, da Silva Patricio IF, de Oliveira Alves N, da Costa Cavalcanti FA. Knowledge levels among elderly people with Diabetes Mellitus concerning COVID-19: an educational intervention via a teleservice. Acta Diabetol. 2020. doi:http://dx.doi.org/10.1007/s00592-020-01580-y
347. de Lucena EHG, Freire AR, Freire DEWG, de Araujo ECF, Lira GNW, Brito ACM, et al. Offer and use of oral health in primary care before and after the beginning of the COVID-19 pandemic in Brazil. Pesqui Bras Odontopediatria Clin Integr. 2020;20: 1–6. doi:http://dx.doi.org/10.1590/pboci.2020.163
348. de Maio Nascimento M. Covid-19: U3A students’ report on the impacts of social isolation on physical and mental health and access to information about the virus during the pandemic. Educ Gerontol. 2020;46: 499–511. doi:10.1080/03601277.2020.1795371

William Marciel de S, Lewis Fletcher B, Darlan da Silva C, Jean Paul C, Sabrina L, Alexander Z, et al. Epidemiological and clinical characteristics of the early phase of the COVID-19 epidemic in Brazil. medRxiv. 2020. doi:10.1101/2020.04.25.20077396
358. Del Brutto OH, Costa AF, Mera RM, Recalde BY, Bustos JA, Garcia HH. Late incidence of SARS-CoV-2 infection in a highly-endemic remote rural village. A prospective population-based cohort study. Pathog Glob Health. 2020. doi:http://dx.doi.org/10.1080/20477724.2020.1826152
359. Del Brutto OH, Costa AF, Mera RM, Recalde BY, Bustos JA, Garcia HH. SARS-CoV-2-related mortality in a rural Latin American population. Int J Infect Dis. 2020;99: 226–228. doi:http://dx.doi.org/10.1016/j.ijid.2020.08.003
360. Del Brutto OH, Costa AF, Mera RM, Recalde BY, Bustos JA, García HH. Household Clustering of SARS-CoV-2 in Community Settings: A Study from Rural Ecuador. Am J Trop Med Hyg. 2020. doi:10.4269/ajtmh.20-0688
361. Del Brutto OH, Costa AF, Mera RM, Recalde BY, Bustos JA, García HH. SARS-CoV-2 in rural Latin America. A population-based study in coastal Ecuador. Clin Infect Dis. 2020. doi:10.1093/cid/ciaa1055
362. Del Brutto OH, Costa AF, Recalde BY, Mera RM. Frailty and SARS-CoV-2 infection. A population-based study in a highly endemic village. J Neurol Sci. 2020;418: 117136. doi:http://dx.doi.org/10.1016/j.jns.2020.117136
363. Del Brutto OH, Mera RM, Recalde BY, Costa AF. Social Determinants of Health and Risk of SARS-CoV-2 Infection in Community-Dwelling Older Adults Living in a Rural Latin American Setting. J Community Health. 2020. doi:10.1007/s10900-020-00887-9
364. Delgado D, Wyss Quintana F, Perez G, Sosa Liprandi A, Ponte-Negretti C, Mendoza I, et al. Personal Safety during the COVID-19 Pandemic: Realities and Perspectives of Healthcare Workers in Latin America. Int J Environ Res Public Health. 2020;17. doi:https://dx.doi.org/10.3390/ijerph17082798

365. Delgado-Enciso I, Paz-Garcia J, Barajas-Saucedo C, Mokay-Ramírez K, Meza-Robles C, Lopez-Flores R, et al. Patient-Reported Health Outcomes After Treatment of COVID-19 with Nebulized and/or Intravenous Neutral Electrolyzed Saline Combined with Usual Medical Care Versus Usual Medical care alone: A Randomized, Open-Label, Controlled Trial. ResearchSquare. 2020. doi:10.21203/rs.3.rs-68403/v1
366. Delgado-Gallegos JL, Montemayor-Garza RJ, Padilla-Rivas GR, Franco-Villareal H, Islas JF. Prevalence of stress in healthcare professionals during the covid-19 pandemic in Northeast Mexico: A remote, fast survey evaluation, using an adapted covid-19 stress scales. Int J Environ Res Public Health. 2020;17: 1–12. doi:http://dx.doi.org/10.3390/ijerph17207624
367. Demenech LM, Dumith S de C, Vieira MECD, Neiva-Silva L. Income inequality and risk of infection and death by COVID-19 in Brazil. Desigual Econ e risco Infecc e morte por COVID-19 no Bras. 2020;23: e200095. doi:https://dx.doi.org/10.1590/1980-549720200095
368. Denova-Gutiérrez E, Lopez-Gatell H, Alomia-Zegarra JL, López-Ridaura R, Zaragoza-Jimenez CA, Dyer-Leal DD, et al. The association between obesity, type 2 diabetes, and hypertension with severe COVID-19 on admission among Mexicans. Obesity (Silver Spring). 2020;28: 1826–1832. doi:10.1002/oby.22946
369. Di Mascio D. Maternal and Perinatal Outcomes of Pregnant Women with SARS-COV-2 infection. Ultrasound Obstet Gynecol. 2020. doi:http://dx.doi.org/10.1002/uog.23107
370. Di Tommaso F, Maurice MF, Sastre P, Prado AH, Domine E, Aguero P, et al. Whatsapp consultations in the department of electrophysiology of a public hospital of the City of Buenos Aires in times of COVID-19. Rev Argent Cardiol. 2020;88: 225–228. doi:http://dx.doi.org/10.7775/rac.v88.i3.17976
371. Di M, Gonzalo y otros. PROTECCIÓN Y SITUACIÓN LABORAL DE LOS MÉDICOS ONCÓLOGOS DURANTE LA PANDEMIA DE COVID-19 EN LA ARGENTINA. 2020;8.
372. Diana C, Lenice de Castro Muniz de Q, Lílian Moura de Lima S, Tuany Nunes C, Deisi Cardoso S. Educational actions to combat COVID-19: experience report. Res Soc Dev. 2020;9. doi:10.33448/RSD-V9I8.5207
373. Diana MR-G, Sandra CG-C, Natalia M, Erika VJ-P, Jaime AC-O. COVID-19 in Latin America: Contrasting phylodynamic inference with epidemiological surveillance. medRxiv. 2020. doi:10.1101/2020.05.23.20111443
374. Díaz Castrillón C, Cortés N, Rey S, Pineda M, Díaz Castrillón J, Sierra S. Percepción de la pandemia COVID-19 en los servicios de cirugía en Colombia. Rev colomb cir. 2020;35: 290–301.
375. Díaz Castrillón C, Cortés N, Rey S, Pineda M, Díaz Castrillón J, Sierra S. Perception of the COVID-19 pandemic in the departments of surgery in Colombia. Rev colomb cir. 2020;35: 290–301.
376. Díaz Morales Y, Hidalgo Mesa CJ, Reyes Roque AC, Filgueira Morilla LM, Tamayo Díaz YB, Aguilera Calzadilla Y. Training for professionals and other health workers as regard as hospital management of COVID-19. Edumecentro. 2020;12: 8–24.
377. Díaz FJ, Aguilar-Jiménez W, Flórez-Álvarez L, Valencia G, Laiton-Donato K, Franco-Muñoz C, et al. Isolation and characterization of an early SARS-CoV-2 isolate from the 2020 epidemic in Medellín, Colombia. Biomedica. 2020;40: 148–158. doi:10.7705/biomedica.5834
378. Diaz LA, Garcia-Salum T, Fuentes-Lopez E, Ferres M, Medina RA, Riquelme A, et al. Symptom profiles and risk factors for hospitalization in patients with SARS-CoV-2 and COVID-19: A Large Cohort from South America. Gastroenterology. 2020. doi:https://dx.doi.org/10.1053/j.gastro.2020.05.014
379. Diaz-Corvillon P, Monckeberg M, Barros A, Illanes SE, Soldati A, Nien J-K, et al. Routine screening for SARS CoV-2 in unselected pregnant women at delivery. PLoS One. 2020;15: e0239887. doi:https://dx.doi.org/10.1371/journal.pone.0239887
380. Diaz-Guio DA, Ricardo-Zapata A, Ospina-Velez J, Gomez-Candamil G, Mora-Martinez S, Rodriguez-Morales AJ. Cognitive load and performance of health care professionals in donning and doffing PPE before and after a simulation-based educational intervention and its implications during the COVID-19 pandemic for biosafety. Le Infez Med. 2020;28: 111–117.

D'Imperio H, Gagliardi J, Zoni R, Charask A, Costa YC, Marturano MP, et al. Results of the COVID-19 survey. Impact on cardiovascular care in the argen-iam-st national infarction registry. Rev Argent Cardiol. 2020;88: 217–224. doi:http://dx.doi.org/10.7775/rac.v88.i3.18150
395. Djilali S, Ghanbari B. Coronavirus pandemic: A predictive analysis of the peak outbreak epidemic in South Africa, Turkey, and Brazil. Chaos Solitons Fractals. 2020;138: 109971. doi:https://dx.doi.org/10.1016/j.chaos.2020.109971
396. Dolce Filho R, Nechar RC, Ribeiro Filho A. Preliminary study to assess symptoms and prevalent drugs of the "epidemic genius" of the COVID-19 pandemic in Brazil. 2020; 21.
397. Dolhnikoff M, Duarte-Neto AN, de Almeida Monteiro RA, da Silva LFF, de Oliveira EP, Saldiva PHN, et al. Pathological evidence of pulmonary thrombotic phenomena in severe COVID-19. J Thromb Haemost. 2020;18: 1517–1519. doi:http://dx.doi.org/10.1111/jth.14844

398. Douglas F, Paula F, Paulo Jr. O, Jennifer R, Elicardo G, Andres P. Temporal and spatial characteristics of the spread of COVID-19 in Rio de Janeiro state and city. medRxiv. 2020. doi:10.1101/2020.05.13.20101113
399. Dowall MPM, Barrionuevo-Poquet A, Carnero-Fuentes O, Pareja-Begazo G, Coayla-Cano C, Gallo-Lopez A, et al. Clinical-pathological characterization, viral genotipification and genetic heterogeneity as risk determinants in covid-19: Study design and initial findings. Rev la Fac Med Humana. 2020;20: 433–443. doi:http://dx.doi.org/10.25176/RFMH.v20i3.3040
400. Duarte MMS, Haslett MIC, Freitas LJA de, Gomes NTN, Silva DCC da, Percio J, et al. Description of COVID-19 hospitalized health worker cases in the first nine weeks of the pandemic, Brazil, 2020. Descricao dos casos Hosp pela COVID-19 em profissionais saude nas Prim nove Sem da pandemia, Bras 2020. 2020;29: e2020277. doi:https://dx.doi.org/10.1590/S1679-49742020000500011
401. Duarte M de Q, Santo MA da S, Lima CP, Giordani JP, Trentini CM. Covid-19 and the impacts on mental health: a sample from Rio Grande do Sul, Brazil. COVID-19 e os impactos na saude Ment uma amostra do Rio Gd do Sul, Bras. 2020;25: 3401–3411. doi:https://dx.doi.org/10.1590/1413-81232020259.16472020
402. Duarte RB, Medeiros LMF, Araújo MJA de M, Cavalcante ASP, Souza EC de, Alencar OM de, et al. Community health agents in front of COVID-19: experiences with nursing professionals. Enferm foco. 2020;11: 252–256.
403. Duarte-Filho GC, Brum AA, Ospina R, Almeida FAG, Macêdo AMS, Vasconcelos GL. Recife and Belém are currently the only capitals that are already in the saturation phase of Covid-19 in Brazil. 2020.
404. Duarte-Filho GC, Brum AA, Ospina R, Almeida FAG, Macêdo AMS, Vasconcelos GL. Situation of the Covid-19 Epidemic in Brazil in August 2020: Most Northern and Northeastern States in a Saturation Phase, while Southern States in Acceleration or Slight Deceleration. 2020.
405. Duarte-Neto AN, Monteiro RAA, da Silva LFF, Malheiros DMAC, de Oliveira EP, Theodoro-Filho J, et al. Pulmonary and systemic involvement in COVID-19 patients assessed with ultrasound-guided minimally invasive autopsy. Histopathology. 2020;77: 186–197. doi:http://dx.doi.org/10.1111/his.14160
406. Duczmal LH, Almeida ACL, Duczmal DB, Alves CRL, Magalhaes FCO, Lima MS de, et al. Vertical social distancing policy is ineffective to contain the COVID-19 pandemic. Cad Saude Publica. 2020;36: e00084420. doi:https://dx.doi.org/10.1590/0102-311x00084420
407. Durán Morera N, Botello Ramírez E. Detection of emerging “active” clusters with high incidence rates, for rapid surveillance of COVID-19. Medicentro (Villa Clara). 2020;24: 642–655.
408. Eddy L-N, Paola S-P, Santiago R-V, Jose L-N, Paul V-M, Giuseppe B-B, et al. Paradigms About the COVID-19 Pandemic: What Medical Students Know and Feel. SSRN. 2020. doi:10.2139/ssrn.3594642

Eddy L-N, Paola S-P, Santiago R-V, Jose L-N, Paul V-M, Giuseppe B-B, et al. Paradigms about the COVID-2 pandemic: knowledge, attitudes and practices from medical students. medRxiv. 2020. doi:10.1101/2020.05.21.20105858
409. Edilson C. The SARS-COV-2 outbreak around the Amazon rainforest: the relevance of the airborne transmission. medRxiv. 2020. doi:10.1101/2020.08.06.20169433
410. Eduardo Atem De C, Rogerio Atem De C. Identification of Patterns in Epidemic Cycles and Methods for Estimating Their Duration: COVID-19 Case Study. medRxiv. 2020. doi:10.1101/2020.07.13.20153080
411. Eduardo Juscamayta L, David T, Faviola Valdivia G, Nancy Rojas S, Dennis C, Lenin Maturrano H, et al. Phylogenomic reveals multiple introductions and early spread of SARS-CoV-2 into Peru. bioRxiv. 2020. doi:10.1101/2020.09.14.296814
412. Eduardo AU, Gerardo C, Kenji M. Case fatality risk by age from COVID-19 in a high testing setting in Latin America: Chile, March-May, 2020. medRxiv. 2020. doi:10.1101/2020.05.25.20112904
413. Edwin Sr. NBR, Anthony Sr. JFE, Fernando EAL, oacutepez Sr., Pa, uacutel HV, et al. Environmental and social analysis as risk factors for the spread of the novel coronavirus (SARS-CoV-2) using remote

sensing, GIS and analytical hierarchy process (AHP): Case of Peru. medRxiv. 2020. doi:10.1101/2020.05.31.20118653
414. Efren M-Z, Oliver M-C, Ivan D-E, Carlos MH-S. Predictors of severe symptomatic laboratory-confirmed SARS-COV-2 reinfection. medRxiv. 2020. doi:10.1101/2020.10.14.20212720
415. Elachola H, Gozzer E, Rahman NMM, Ditekemena J, Pando-Robles V, Pa K, et al. Photo-epidemiology to estimate face covering use in select areas in Asia versus the Americas and Africa during the COVID-19 pandemic. J Travel Med. 2020. doi:10.1093/jtm/taaa121
416. Eliandro Rodrigues C, Miguel C, Paulo N, Neyva R, Jeinny P, Austin W, et al. One Study of COVID-19 Spreading at The United States - Brazil - Colombia. medRxiv. 2020. doi:10.1101/2020.08.29.20184465
417. Elías-Gutiérrez M, Mendoza-Carranza M, Takahashi-Aguilar U, Valdez-Moreno M, Prado-Bernal J, Mora-Loya C. Why does the COVID-19 outbreak in Mexico seem to be slower? Some hypotheses for testing. ResearchSquare. 2020. doi:10.21203/rs.3.rs-51450/v1
418. Elsa Berenice G-P, Verónica M-R, Ignacio M-G-H, Luz María G-A, Marco Antonio Á-A, Lucía C-N, et al. Vulnerabilidad, conocimiento sobre medidas de mitigación y exposición ante COVID-19 en adultos de México: Resultados de la ENSARS-CoV-2. Salud Publica Mex. 2020. doi:10.21149/11875
419. Elvira Maria G-S, Simone Schneider W, Clovis P, Guilherme Wataru G, Eduardo Jun S, Tiago Borges Ribeiro G, et al. Overview on COVID-19 outbreak indicators across Brazilian federative units. medRxiv. 2020. doi:10.1101/2020.06.02.20120220
420. Emer C, Maia KP, Santana PC, Santana EM, Silva DGM da, Cosmo LG, et al. Vulnerability of the micro-regions of the South of Brazil to the new coronavirus (SARS-CoV-2) pandemic. 2020.
421. Emerson Lucas Silva C, Bruna Isabela Adolpho de O, Igor Fessina S, Anderson Reis de S, Jules Ramon Brito T, Isabel Amelia Costa M, et al. LOW PSYCHOLOGICAL WELL-BEING IN MEN WHO HAVE SEX WITH MEN (MSM) DURING THE SHELTER-IN-PLACE ORDERS TO PREVENT THE COVID-19 SPREAD: RESULTS FROM A NATIONWIDE STUDY. medRxiv. 2020. doi:10.1101/2020.09.21.20198929
422. Emilio G, Adrian R, Tiago T. Information and Behavioral Responses during a Pandemic: Evidence from Delays in COVID-19 Death Reports. SSRN. 2020. doi:10.2139/ssrn.3645317
423. Endo PT, Silva I, Lima L, Bezerra L, Gomes R, Ribeiro-Dantas M, et al. #StayHome: Monitoring and benchmarking social isolation trends in Caruaru and the Região Metropolitana do Recife during the COVID-19 pandemic. Rev Soc Bras Med Trop. 2020;53: e20200271. doi:10.1590/0037-8682-0271-2020
424. Enner A, Jose M, Luiz R, Edward P, Thanan R, Fernando C, et al. Investigating spatiotemporal patterns of the COVID-19 in Sao Paulo State, Brazil. medRxiv. 2020. doi:10.1101/2020.05.28.20115626
425. Eric Luis Barroso C, Juliana Cristina Cardoso F. Surge capacities and predicted demands of Brazil's health system associated with severe COVID-19 cases. medRxiv. 2020. doi:10.1101/2020.04.02.20050351
426. Ernandes Gonçalves D. Casos confirmados do novo Coronavírus: interiorização da doença em um município de Minas Gerais. J Nurs Heal. 2020;10: 1–12.
427. Escalera-Antezana JP, Lizon-Ferrufino NF, Maldonado-Alanoca A, Alarcón-De-la-Vega G, Alvarado-Arnez LE, Balderrama-Saavedra MA, et al. Clinical features of cases and a cluster of Coronavirus Disease 2019 (COVID-19) in Bolivia imported from Italy and Spain. Travel Med Infect Dis. 2020;35: 101653. doi:10.1016/j.tmaid.2020.101653

Escalera-Antezana JP, Lizon-Ferrufino NF, Maldonado-Alanoca A, Alarcon-De-la-Vega G, Alvarado-Arnez LE, Balderrama-Saavedra MA, et al. Clinical features of the first cases and a cluster of Coronavirus Disease 2019 (COVID-19) in Bolivia imported from Italy and Spain. Travel Med Infect Dis. 2020;35: 101653. doi:https://dx.doi.org/10.1016/j.tmaid.2020.101653
428. Escalera-Antezana JP, Cerruto-Zelaya PE, Apaza-Huasco M, Miranda-Rojas SH, Flores-Cardenas CA, Rivera-Zabala L, et al. Healthcare workers' and students' knowledge regarding the transmission,

epidemiology and symptoms of COVID-19 in 41 cities of Bolivia and Colombia. Travel Med Infect Dis. 2020; 101702. doi:https://dx.doi.org/10.1016/j.tmaid.2020.101702

429. Escalera-Antezana JP, Lizon-Ferrufino NF, Maldonado-Alanoca A, Alarcon-De-la-Vega G, Alvarado-Arnez LE, Balderrama-Saavedra MA, et al. Risk factors for mortality in patients with Coronavirus Disease 2019 (COVID-19) in Bolivia: An analysis of the first 107 confirmed cases. Le Infez Med. 2020;28: 238–242.

430. Escobar G, Matta J, Taype Huamaní W, Ayala R, Amado J. Clinical - Epidemiological Characteristics Of Patients Deceased By COVID-19 In A National Hospital In Lima, Peru. Rev Fac Med Hum. 2020;20: 180–185.

431. Escosteguy CC, Eleuterio T de A, Pereira AGL, Marques MRVE, Brandão AD, Batista JPM. COVID-19: sectional study of suspected cases admitted to a federal hospital in Rio de Janeiro, Brazil, and factors associated with hospital death. 2020. doi:10.1590/s1679-49742021000100022

432. Espinosa MM, Oliveira EC de, Melo JS, Damaceno RD, Terças-Trette ACP. Prediction of COVID -19 cases and deaths in Mato Grosso state and Brazil. J Heal Biol Sci. 2020;8.

439. Evangelina Cervantes H, Pavel Roel Gutiérrez S. Resistir la Covid-19: intersecciones en la educación de Ciudad Juárez, México. Rev Int Educ para la Justicia Soc. 2020;9.

440. Ezequiel A, Franco M. COVID-19 mild cases determination from correlating COVID-line calls to reported cases. medRxiv. 2020. doi:10.1101/2020.07.26.20162008

441. Ezequiel A, Franco M, Nicolas K, Senastian C, Daniela O, Enio G. Estimating COVID-19 cases and outbreaks on-stream through phone-calls. medRxiv. 2020. doi:10.1101/2020.10.09.20210351

442. Fabian D, Alvaro J-M. Psychological distress during the COVID-19 epidemic in Chile: the role of economic uncertainty. medRxiv. 2020. doi:10.1101/2020.09.27.20202648

443. Fabiana G, Fabio Macedo M, Silvano Barbosa O, Victor Bertolo Gomes P, Wildo A, Helder N, et al. The impact of early social distancing at COVID-19 Outbreak in the largest Metropolitan Area of Brazil. medRxiv. 2020. doi:10.1101/2020.04.06.20055103

444. Fabio Rocha de P, Marcia Gomide da Silva M. Análise de Redes Sociais: a formação de grupos do Facebook frente à epidemia da COVID-19 no Brasil. 2020;32.

Fernandes D, Ferreira P, Kirsztajn G. Pre-Exposure Prophylaxis During COVID-19 Pandemic: Can PrEP Prevent Flu-Like Symptoms? ResearchSquare. 2020. doi:10.21203/rs.3.rs-27798/v1
465. Fernandes EG, Santos J da S, Sato HK. Outbreak investigation in cargo ship in times of COVID-19 crisis, Port of Santos, Brazil. Rev Saude Publica. 2020;54: 34. doi:https://dx.doi.org/10.11606/s1518-8787.2020054002461
466. Fernandes J, Silva R, Silva A, Villela D, Mendonça V, Lacerda A. Altitude conditions seem to determine the evolution of COVID-19 in Brazil: a significance relationship with cases and deaths. ResearchSquare. 2020. doi:10.21203/rs.3.rs-76633/v1
467. Fernandes L, Araújo F, Silva M. Insights into the dynamics of SARS-CoV-2 pandemicvia Shannon-Fisher causality plane. ResearchSquare. 2020. doi:10.21203/rs.3.rs-36581/v1
468. Fernandez RS, Crivelli L, Guimet NM, Allegri RF, Pedreira ME. Psychological distress associated with COVID-19 quarantine: Latent profile analysis, outcome prediction and mediation analysis. J Affect Disord. 2020;277: 75–84. doi:http://dx.doi.org/10.1016/j.jad.2020.07.133
469. Fernando Pérez F, Adrial Despaigne B, Ernesto Sánchez H, Odalis Cardona G, Jacno Erik Ferrer C. Variabilidad de los resultados de la PCR evolutiva en pacientes con la COVID-19. 2020;24.
470. Fernando Timoteo F, Tiago Almeida de O, Cristiane Esteves T, Andre Filipe de Moraes B, Gabriel Dalla C, Alexandre Chiavegatto F. A multipurpose machine learning approach to predict COVID-19 negative prognosis in Sao Paulo, Brazil. medRxiv. 2020. doi:10.1101/2020.08.26.20182584
471. Fernando AB, Leonardo SLB, Pedro K, Leila FD, Fernando GZ, Marcio S, et al. Characteristics and Outcomes of 4,942 Critically Ill Adult Patients with COVID-19 in Brazil. SSRN. 2020. doi:10.2139/ssrn.3670662
472. Fernando R, Luis Gabriel P-L, Olga Lucía A-R, Diana Marcela M-R, Virginia Z. Differential Diagnosis between Dengue and COVID-19 Infection: Diagnosis Challenge in Tropical Regions. SSRN. 2020. doi:10.2139/ssrn.3590469
473. Fernando T, Adrian Ezequiel Y, Daniel L, Pablo L, Pedro B, Marcelo C, et al. Sooner than you think: a very early affective reaction to the COVID-19 pandemic and quarantine in Argentina. medRxiv. 2020. doi:10.1101/2020.07.31.20166272
474. Feroze N. Forecasting the patterns of COVID-19 and causal impacts of lockdown in top five affected countries using Bayesian Structural Time Series Models. Chaos Solitons Fractals. 2020;140: 110196. doi:10.1016/j.chaos.2020.110196
475. Ferraz LMR. Saúde e política na crise da Covid-19: apontamentos sobre a pandemia na imprensa brasileira. RECIIS (Online). 2020;14: 273–278.
476. Ferreira JC, Ho Y-L, Besen BAMP, Malbuisson LMS, Taniguchi LU, Mendes P V, et al. Characteristics and outcomes of patients with COVID-19 admitted to the ICU in a university hospital in Sao Paulo, Brazil - study protocol. Clinics (Sao Paulo). 2020;75: e2294. doi:https://dx.doi.org/10.6061/clinics/2020/e2294
477. Ferreira S. On the efficiency of restrictive sanitary barriers to contain the progress of COVID-19: A simple mathematical modeling. 2020.
478. Ferrer Castro JE, Sánchez Hernández E, Poulout Mendoza A, del Río Caballero G, Figueredo Sánchez D. Clinical and epidemiological characterization of patients confirmed with COVID-19 in Santiago de Cuba. Medisan. 2020;24: 473–485.

Flávio CC, Raquel ML, Oswaldo GC, Daniel V, Leonardo SB, Ana Pastore y P, et al. Assessing the Potential Impact of COVID-19 in Brazil: Mobility, Morbidity and the Burden on the Health Care System. SSRN. 2020. doi:10.2139/ssrn.3559609
484. Fonseca M, Beatriz E. The cuban radio against the COVID-19. A case study. 2020.
485. Forlenza O V, Stella F, HCFMUSP LIMPC. Impact of SARS-CoV-2 pandemic on mental health in the elderly: perspective from a psychogeriatric clinic at a tertiary hospital in Sao Paulo, Brazil. Int psychogeriatrics. 2020; 1–5. doi:https://dx.doi.org/10.1017/S1041610220001180
486. Fortaleza CMCB, Guimaraes RB, de Almeida GB, Pronunciate M, Ferreira CP. Taking the inner route: spatial and demographic factors affecting vulnerability to COVID-19 among 604 cities from inner Sao Paulo State, Brazil. Epidemiol Infect. 2020;148: e118. doi:https://dx.doi.org/10.1017/S095026882000134X
487. Fowler Z, Moeller E, Roa L, Castañeda-Alcántara ID, Uribe-Leitz T, Meara JG, et al. Projected impact of COVID-19 mitigation strategies on hospital services in the Mexico City Metropolitan Area. PLoS One. 2020;15: e0241954. doi:10.1371/journal.pone.0241954
488. Francis Ribeiro S, Daisy M-S, Douglas dos Santos S, Juliana Beust L, Gustavo Goncalves C, Luciano Santos Pinto G, et al. Physical Activity Decreases the Prevalence of COVID-19-associated Hospitalization: Brazil EXTRA Study. medRxiv. 2020. doi:10.1101/2020.10.14.20212704
489. Francisca Marli Rodrigues de A, Tarssio Brito B, Andrés H-F, Andrea U, Yu Ting L. Twitter in Brazil: Discourses on China in Times of Coronavirus. SSRN. 2020. doi:10.2139/ssrn.3608566
490. Francisco HCF, Juvenia BF. Instantaneous R calculation for COVID-19 epidemic in Brazil. medRxiv. 2020. doi:10.1101/2020.04.23.20077172
491. Franco AS, Henrique DM, Almeida LF de, Prado LM do, Gomes RVS, Silva RCL. Safety in the Rapid sequence intubation recommended in COVID-19: Experience report. rev Cuid (Bucaramanga 2010). 2020;11: e1260–e1260.
492. Franco-Munoz C, Alvarez-Diaz DA, Laiton-Donato K, Wiesner M, Escandon P, Usme-Ciro JA, et al. Substitutions in Spike and Nucleocapsid proteins of SARS-CoV-2 circulating in South America. Infect Genet Evol. 2020;85: 104557. doi:http://dx.doi.org/10.1016/j.meegid.2020.104557
493. Freire-Paspuel B, Vega-Mariño P, Velez A, Castillo P, Masaquiza C, Cedeño-Vega R, et al. "One health" inspired SARS-CoV-2 surveillance: The Galapagos Islands experience. One Heal (Amsterdam, Netherlands). 2020; 100185. doi:10.1016/j.onehlt.2020.100185

494. Freitas ARR, Medeiros NM, Frutuoso LCV, Beckedorff OA, Martin LMA de, Medeiros MM de, et al. Use of excess mortality associated with the COVID-19 epidemic as an epidemiological surveillance strategy - preliminary results of the evaluation of six Brazilian capitals. 2020. doi:10.1590/SciELOPreprints.442
495. Freitas VLS, Konstantyner TCRO, Mendes JF, Sepetauskas CSDN, Santos LBL. The correspondence between the structure of the terrestrial mobility network and the spreading of COVID-19 in Brazil. Cad Saude Publica. 2020;36: e00184820. doi:http://dx.doi.org/10.1590/0102-311X00184820

Vander LSF, Jeferson F, Catia SNS, Leonardo BLS. The correspondence between the structure of the terrestrial mobility network and the emergence of COVID-19 in Brazil. medRxiv. 2020. doi:10.1101/2020.05.17.20104612
496. Freitas-Jesus J V, Rodrigues L, Surita FG. The experience of women infected by the COVID-19 during pregnancy in Brazil: a qualitative study protocol. Reprod Health. 2020;17: 108. doi:10.1186/s12978-020-00958-z
497. Friedman J, Calderon-Villarreal A, Bojorquez I, Vera Hernandez C, Schriger DL, Tovar Hirashima E. Excess Out-of-Hospital Mortality and Declining Oxygen Saturation: The Sentinel Role of Emergency Medical Services Data in the COVID-19 Crisis in Tijuana, Mexico. Ann Emerg Med. 2020;76: 413–426. doi:http://dx.doi.org/10.1016/j.annemergmed.2020.07.035

Joseph F, Alheli C-V, Ietza B, Carlos Vera H, David S, Eva Tovar H. Excess Out-Of-Hospital Mortality and Declining Oxygen Saturation Documented by EMS During the COVID-19 Crisis in Tijuana, Mexico. medRxiv. 2020. doi:10.1101/2020.05.13.20098186
498. Friedman J, Calderon-Villarreal A, Bojorquez I, Vera Hernandez C, Schriger D, Tovar Hirashima E. Excess Out-Of-Hospital Mortality and Declining Oxygen Saturation: The Sentinel Role of EMS Data in the COVID-19 Crisis in Tijuana, Mexico. medRxiv  Prepr Serv Heal Sci. 2020. doi:https://dx.doi.org/10.1101/2020.05.13.20098186
499. Friedrich F, Ongaratto R, Scotta MC, Veras TN, Stein R, Lumertz MS, et al. Early Impact of social distancing in response to COVID-19 on hospitalizations for acute bronchiolitis in infants in Brazil. Clin Infect Dis. 2020. doi:http://dx.doi.org/10.1093/cid/ciaa1458
500. Furtado RHM, Berwanger O, Fonseca HA, Correa TD, Ferraz LR, Lapa MG, et al. Azithromycin in addition to standard of care versus standard of care alone in the treatment of patients admitted to the hospital with severe COVID-19 in Brazil (COALITION II): a randomised clinical trial. Lancet. 2020;396: 959–967. doi:http://dx.doi.org/10.1016/S0140-6736%2820%2931862-6
501. Gabriel Berg de A, Thomas V, Claudia F, Carlos Magno Castelo Branco F. Several countries in one: a mathematical modeling analysis for COVID-19 in inner Brazil. medRxiv. 2020. doi:10.1101/2020.04.23.20077438
502. Gaitan-Rossi P, Vilar-Compte M, Teruel G, Perez-Escamilla R. Food insecurity measurement and prevalence estimates during the COVID-19 pandemic in a repeated cross-sectional survey in Mexico. Public Health Nutr. 2020; 1–25. doi:http://dx.doi.org/10.1017/S1368980020004000
503. Galan-Rodas E, Tarazona-Fernandez A, Palacios-Celi M. Risk and death of doctors 100 days after the state of emergency by COVID-19 in Peru. Acta Medica Peru. 2020;37: 119–121. doi:http://dx.doi.org/10.35663/amp.2020.372.1033
504. Galhardi CP, Freire NP, Minayo MCS, Fagundes MCM. Fact or Fake? An analysis of disinformation regarding the Covid-19 pandemic in Brazil. Cien Saude Colet. 2020;25: 4201–4210. doi:http://dx.doi.org/10.1590/1413-812320202510.2.28922020
505. Galván-Tejada CE, Zanella-Calzada LA, Villagrana-Bañuelos KE, Moreno-Báez A, Luna-García H, Celaya-Padilla JM, et al. Demographic and Comorbidities Data Description of Population in Mexico with SARS-CoV-2 Infected Patients(COVID19): An Online Tool Analysis. Int J Environ Res Public Health. 2020;17. doi:10.3390/ijerph17145173

García-Reyna B, Castillo-García G, Barbosa-Camacho F, Cervantes-Cardona G, Cervantes-Pérez E, Torres-Mendoza B, et al. Fear of COVID-19 Scale for Hospital Staff in Regional Hospitals in Mexico: A Survey Study. ResearchSquare. 2020. doi:10.21203/rs.3.rs-34065/v1
517. García-Salirrosas EE, Sánchez-Poma RA. PREVALENCIA DE LOS TRASTORNOS MUSCULOESQUELÉTICO EN DOCENTES UNIVERSITARIOS QUE REALIZAN TELETRABAJO EN TIEMPOS DE COVID-19. 2020.
518. Gaspar GDS, Figueiredo N, de Lucena EHG, Ceissler CAS, Cavalcanti RP, de Goes PSA. Characterization of dental surgeons of pernambuco state in the covid-19 pandemic context: Preliminary data. Pesqui Bras Odontopediatria Clin Integr. 2020;20: 1–7. doi:http://dx.doi.org/10.1590/pboci.2020.152

Gaspar G da S, Figueiredo N, Lucena EHG de, Ceissler CAS, Cavalcanti RP, Goes PSA de. Characterization of dental surgeons of Pernambuco in the COVID-19 pandemic context: preliminary data. 2020.
519. Gaston V-H, Andres N. Extension and implementation of a system modelling the COVID-19 pandemic in Chile. medRxiv. 2020. doi:10.1101/2020.06.21.20136606

Karla Fabiana G, Carolina Ferreira N, Diego Augusto Nunes R, Sofia Mendes S, Gabriel Perri E, Rafael G, et al. Outpatient screening of health status and lifestyle among post-bariatric patients during the Covid-19 pandemic in Sao Paulo, Brazil. medRxiv. 2020. doi:10.1101/2020.07.30.20165068
532. Goiás . Secretaria da S. Comorbidade, idade e letalidade no COVID-19 no estado de Goiás. 2020.
533. Gois AN, Laureano EE, Santos DDS, Sanchez DE, Souza LF, Vieira RCA, et al. Lockdown as an Intervention Measure to Mitigate the Spread of COVID-19: a modeling study. Rev Soc Bras Med Trop. 2020;53: e20200417. doi:http://dx.doi.org/10.1590/0037-8682-0417-2020
534. Gomes CM, Favorito LA, Henriques JVT, Canalini AF, Anzolch KMJ, de Carvalho Fernandes R, et al. Impact of COVID-19 on clinical practice, income, health and lifestyle behavior of Brazilian urologists. Int Braz J Urol. 2020;46. doi:https://dx.doi.org/10.1590/S1677-5538.IBJU.2020.99.15
535. Gomes DS, Andrade LA, Ribeiro CJN, Peixoto MVS, Lima SVMA, Duque AM, et al. Risk clusters of COVID-19 transmission in Northeastern Brazil: Prospective space-time modeling. Epidemiol Infect. 2020. doi:http://dx.doi.org/10.1017/S0950268820001843

Laura G-L, Isabel B, Irina V, Maria Genoveva G-A, Byron F, Blanca R-T. First SARS-CoV-2 detection in river water: implications in low sanitation countries. medRxiv. 2020. doi:10.1101/2020.06.14.20131201

568. Guerrero-Nancuante C, Manriquez P R. [An epidemiological forecast of COVID-19 in Chile based on the generalized SEIR model and the concept of recovered]. Medwave. 2020;20: e7898. doi:https://dx.doi.org/10.5867/medwave.2020.04.7898
569. Guerrero-Torres L, Caro-Vega Y, Crabtree-Ramirez B, Sierra-Madero JG. Clinical Characteristics and Mortality of Healthcare Workers with SARS-CoV-2 infection in Mexico City. Clin Infect Dis. 2020. doi:http://dx.doi.org/10.1093/cid/ciaa1465
570. Guilherme Pertinni de Morais G, Samara Sousa Vasconcelos G, Samila Sousa V, Elisson de Sousa Mesquita S, Rebeca Galdino M, Rosangela Lago da S. INCIDÊNCIA E LETALIDADE DA COVID-19 NO CEARÁ, 2020. 2020;14.
571. Guilherme SC, Wesley Sr. C, Silvio CF. Metapopulation modeling of COVID-19 advancing into the countryside: an analysis of mitigation strategies for Brazil. medRxiv. 2020. doi:10.1101/2020.05.06.20093492
572. Guillermo Alberto Pérez F. ¿Es la comorbilidad cardiovascular la causante de la elevación de la proteína C reactiva en pacientes positivos a la COVID-19? 2020;14.
573. Guimaraes RM, Magalhaes M de AFM, Xavier DR, Saldanha R de F, Catao R de C. Is it time to talk about the end of social distancing? A joinpoint analysis of COVID-19 time series in Brazilian capitals. Rev Soc Bras Med Trop. 2020;53: e20200469. doi:https://dx.doi.org/10.1590/0037-8682-0469-2020
574. Guiroy A, Gagliardi M, Coombes N, Landriel F, Zanardi C, Camino Willhuber G, et al. COVID-19 Impact Among Spine Surgeons in Latin America. Glob Spine J. 2020. doi:http://dx.doi.org/10.1177/2192568220928032
575. Guiroy A, Gagliardi M, Coombes N, Landriel F, Zanardi C, Camino Willhuber G, et al. Psychosocial impact of the COVID-19 pandemic on Argentine spinal surgeons. Rev argent neurocir. 2020;34: 187–193.
576. Gustavo Lugo G, Carmen H-C, Carlos T-S, Felipe J, Hector S, Cristina A, et al. Clinical characteristics and outcomes of patients with COVID-19 and ARDS admitted to a third level health institution in Mexico City. medRxiv. 2020. doi:10.1101/2020.09.12.20193409
577. Gustavo C, Maria-Isabel L, Mauricio G-N, Claudia Alvarado De La B, Santiago A-R, amy peralta-prado, et al. Acute kidney injury in patients with severe COVID-19 in Mexico. medRxiv. 2020. doi:10.1101/2020.08.28.20167379
578. Gutiérrez ED, Puche R, Hernández F. Estimation of COVID-19 cases in South American countries using ARIMA models. Obs del Conoc Rev Espec en Gestión Soc del Conoc. 2020;5: 11–25.
579. Haider N, Yavlinsky A, Simons D, Osman AY, Ntoumi F, Zumla A, et al. Passengers' destinations from China: low risk of Novel Coronavirus (2019-nCoV) transmission into Africa and South America. Epidemiol Infect. 2020;148: e41. doi:https://dx.doi.org/10.1017/S0950268820000424
580. Hallal PC, Hartwig FP, Horta BL, Silveira MF, Struchiner CJ, Vidaletti LP, et al. SARS-CoV-2 antibody prevalence in Brazil: results from two successive nationwide serological household surveys. Lancet Glob Heal. 2020;8: e1390–e1398. doi:http://dx.doi.org/10.1016/S2214-109X%2820%2930387-9
581. Hallal PC, Barros FC, Silveira MF, Barros AJD de, Dellagostin OA, Pellanda LC, et al. EPICOVID19 protocol: repeated serological surveys on SARS-CoV-2 antibodies in Brazil. Cien Saude Colet. 2020;25: 3573–3578. doi:https://dx.doi.org/10.1590/1413-81232020259.25532020
582. Hallal PC, Horta BL, Barros AJD, Dellagostin OA, Hartwig FP, Pellanda LC, et al. Trends in the prevalence of COVID-19 infection in Rio Grande do Sul, Brazil: repeated serological surveys. Cien Saude Colet. 2020;25: 2395–2401. doi:https://dx.doi.org/10.1590/1413-81232020256.1.09632020
583. Heberto AB, Carlos PCJ, Antonio CRJ, Patricia PP, Enrique TR, Danira MPJ, et al. Implications of myocardial injury in Mexican hospitalized patients with coronavirus disease 2019 (COVID-19). IJC Hear Vasc. 2020;30: 100638. doi:http://dx.doi.org/10.1016/j.ijcha.2020.100638
584. Hector Eduardo C, Roberto Raul H, Maria Eugenia F. Safety and Efficacy of the combined use of ivermectin, dexamethasone, enoxaparin and aspirin against COVID-19. medRxiv. 2020. doi:10.1101/2020.09.10.20191619

585. Helcio RBO, Marcelo C, George SD, Luiz FSF. Estimation Of State Variables And Model Parameters For The Evolution Of COVID-19 In The City Of Rio de Janeiro. medRxiv. 2020. doi:10.1101/2020.05.21.20108407
586. Henrique Matheus S, Rafael Silva P, Raquel de Abreu G, Adolfo S, Fabio Andre P. Projection of hospitalization by COVID-19 in Brazilfollowing different social distances policies. medRxiv. 2020. doi:10.1101/2020.04.26.20080143
587. Hernández-Flores ML, Escobar-Sánchez J, Paredes-Zarco JE, Franyuti Kelly GA, Carranza-Ramírez L. Prediction and Potential Spatially Explicit Spread of COVID-19 in Mexico's Megacity North Periphery. Healthc (Basel, Switzerland). 2020;8. doi:10.3390/healthcare8040453
588. Hernández-Galdamez DR, González-Block MÁ, Romo-Dueñas DK, Lima-Morales R, Hernández-Vicente IA, Lumbreras-Guzmán M, et al. Increased Risk of Hospitalization and Death in Patients with COVID-19 and Pre-existing Noncommunicable Diseases and Modifiable Risk Factors in Mexico. Arch Med Res. 2020. doi:10.1016/j.arcmed.2020.07.003
589. Hernández-García I, Giménez-Júlvez T. Characteristics of YouTube Videos in Spanish on How to Prevent COVID-19. Int J Environ Res Public Health. 2020;17: 1–10. doi:10.3390/ijerph17134671
590. Hernandez-Garduno E. Comorbidities that predict acute respiratory syndrome coronavirus 2 test positivity in Mexican Children: A case-control study. Pediatr Obes. 2020. doi:http://dx.doi.org/10.1111/ijpo.12740
591. Hernandez-Garduno E. Obesity is the comorbidity more strongly associated for Covid-19 in Mexico. A case-control study. Obes Res Clin Pract. 2020. doi:https://dx.doi.org/10.1016/j.orcp.2020.06.001
592. Hernandez-Huerta MT, Perez-Campos Mayoral L, Romero Diaz C, Martinez Cruz M, Mayoral-Andrade G, Sanchez Navarro LM, et al. Analysis of SARS-CoV-2 mutations in Mexico, Belize, and isolated regions of Guatemala and its implication in the diagnosis. J Med Virol. 2020. doi:http://dx.doi.org/10.1002/jmv.26591
593. Hernández-Sánchez BR, Cardella GM, Sánchez-García JC. Psychological Factors that Lessen the Impact of COVID-19 on the Self-Employment Intention of Business Administration and Economics' Students from Latin America. Int J Environ Res Public Health. 2020;17: 1–22. doi:10.3390/ijerph17155293
594. Hernández-Vásquez A, Azañedo D, Vargas-Fernández R, Bendezu-Quispe G. Association of Comorbidities With Pneumonia and Death Among COVID-19 Patients in Mexico: A Nationwide Cross-sectional Study. J Prev Med Public Health. 2020;53: 211–219. doi:10.3961/jpmph.20.186
595. Hernandez-Vasquez A, Gamboa-Unsihuay JE, Vargas-Fernandez R, Azanedo D. Excess mortality in Metropolitan Lima during the COVID-19 pandemic: A district level comparison. Medwave. 2020;20: e8031. doi:http://dx.doi.org/10.5867/medwave.2020.08.8032
596. Hernandez-Vasquez A, Vargas-Fernandez R, Bendezu-Quispe G. Association between hypertension and pneumonia caused by SARS-CoV-2 in Mexican population. J Hypertens. 2020;38: 1857–1858. doi:http://dx.doi.org/10.1097/HJH.0000000000002544
597. Herrera-Valenzuela T, Narrea Vargas JJ, Merlo R, Valdés-Badilla PA, Pardo-Tamayo C, Franchini E. Effect of the COVID-19 quarantine on body weight among combat sports athletes. Nutr Hosp. 2020. doi:10.20960/nh.03207
598. Hildegart A, Santos E, Fernando HN. COVID-19 with Uncertain Phases: Estimation Issues with An Illustration for Argentina. SSRN. 2020. doi:10.2139/ssrn.3633500
599. Hillesheim D, Tomasi YT, Figueiró TH, Paiva KM de. Síndrome respiratória aguda grave por COVID-19 em crianças e adolescentes no Brasil: perfil dos óbitos e letalidade hospitalar até a 38a Semana Epidemiológica de 2020. Epidemiol e Serviços Saúde. 2020;29. doi:10.1590/s1679-49742020000500021
600. Horta BL, Silveira MF, Barros AJD, Barros FC, Hartwig FP, Dias MS, et al. Prevalence of antibodies against SARS-CoV-2 according to socioeconomic and ethnic status in a nationwide Brazilian survey. Rev Panam Salud Publica. 2020;44: e135. doi:10.26633/RPSP.2020.135
601. Huamanchumo-Suyon ME, Urrunaga-Pastor D, Ruiz-Perez PJ, Rodrigo-Gallardo PK, Toro-Huamanchumo CJ. Impact of the COVID-19 pandemic on general surgery residency program in Peru: A

cross-sectional study. Ann Med Surg. 2020;60: 130–134. doi:http://dx.doi.org/10.1016/j.amsu.2020.10.031
602. Huamani C, Huamani C, Timana-Ruiz R, Pinedo J, Pinedo J, Perez J, et al. Estimated conditions to control the COVID-19 pandemic in pre- and postquarantine scenarios in Peru. Rev peru med exp salud publica. 2020;37: 195–202. doi:10.17843/RPMESP.2020.372.5405
603. Huerta Saenz IH, Elias Estrada JC, Del Castillo KC, Taya RM, Coronado JC. Maternal and perinatal characteristics of pregnant women with COVID-19 in a national hospital in Lima, Peru. Rev Peru Ginecol y Obstet. 2020;66: 19–24. doi:http://dx.doi.org/10.31403/rpgo.v66i2245
604. Hurtado-Ortiz A, Moreno-Montoya J, Prieto-Alvarado FE, Idrovo Á J. Benchmarking of public health surveillance of COVID-19 in Colombia: First semester. Biomedica. 2020;40: 131–138. doi:10.7705/biomedica.5812
605. Hyun Mo Y, Luis Pedro Lombardi J, Ariana Campos Y. The SIR model estimates incorrectly the basic reproduction number for the covid-19 epidemic. medRxiv. 2020. doi:10.1101/2020.10.11.20210831
606. Hyun Mo Y, Luis Pedro Pedro Lombardi J, Ariana Campos Y, Fabio Fernandes Morato C. Evaluating the impacts of release in Sao Paulo State (Brazil) on the epidemic of covid-19 based on mathematical model. medRxiv. 2020. doi:10.1101/2020.08.03.20167221
607. Hyun Mo Y, Luis Pedro Pedro Lombardi J, Fábio Fernandes Morato C, Ariana Campos Y. Evaluating epidemiological scenarios of isolation and further releases considering protection actions to control transmission of CoViD-19 in Sao Paulo State, Brazil. medRxiv. 2020. doi:10.1101/2020.05.19.20099309
608. Idrovo Á J, Moreno-Montoya J, Pinzón-Flórez CE. Performance of rapid IgM-IgG combined antibody tests in the occupational surveillance of COVID-19 in Colombian enterprises. Biomedica. 2020;40: 139–147. doi:10.7705/biomedica.5829
609. Idrovo AJ, Manrique-Hernandez EF, Fernandez Nino JA. Report From Bolsonaro’s Brazil: The Consequences of Ignoring Science. Int J Health Serv. 2020; 20731420968446. doi:http://dx.doi.org/10.1177/0020731420968446
610. Ietza B, Cesar I, Isabel V, Silvana L, Chiara S. MIGRANTS IN TRANSIT AND ASYLUM SEEKERS IN MEXICO: AN EPIDEMIOLOGICAL ANALYSIS OF THE COVID-19 PANDEMIC. medRxiv. 2020. doi:10.1101/2020.05.08.20095604
611. Igor Gadelha P, Joris MG, Andouglas Goncalves Jr. S, Cosimo D, Gabriel Santos G, Luiz MGG. Forecasting Covid-19 dynamics in Brazil: a data driven approach. medRxiv. 2020. doi:10.1101/2020.05.11.20098392
612. Igor Hermann Huerta S, Jose Carlos Elias E, Katherine Campos Del C, Rossana Muñoz T, Julia Cristina C. Características materno perinatales de gestantes COVID-19 en un hospital nacional de Lima, Perú. Rev Peru Ginecol y Obstet. 2020;66. doi:10.31403/RPGO.V66I2245
613. Igor Silva C, Vinicius Ferreira A, Karina Baltor C, Jean Ezequiel L, Stefan Vilges de O. A vulnerability analysis for the management of and response to the COVID-19 epidemic in the second most populous state in Brazil. medRxiv. 2020. doi:10.1101/2020.07.20.20158345
614. Iragorri N, Gomez-Restrepo C, Barrett K, Herrera S, Hurtado I, Khan Y, et al. Covid-19: Adaptation of a model to predict healthcare resource needs in valle del Cauca, Colombia. Colomb Med. 2020;51: 1–12. doi:http://dx.doi.org/10.25100/cm.v51i3.e4534
615. Iraldo B, Hernandez-Bernal F, Nodarse-Cuni H, Duncan-Roberts Y, Suarez C, Campa-Legrá I, et al. Evaluation of the Effect and Safety of HeberFERON vs Heberon Alpha in Patients Infected with Corona Virus SARS-CoV-2 (Study ESPERANZA/HOPE): Study Protocol for a Randomized Controlled Trial. ResearchSquare. 2020. doi:10.21203/rs.3.rs-28958/v1
616. Irigoyen-Camacho ME, Velazquez-Alva MC, Zepeda-Zepeda MA, Cabrer-Rosales MF, Lazarevich I, Castano-Seiquer A. Effect of income level and perception of susceptibility and severity of covid-19 on stay-at-home preventive behavior in a group of older adults in Mexico City. Int J Environ Res Public Health. 2020;17: 1–16. doi:http://dx.doi.org/10.3390/ijerph17207418

617. Irvin R. On the Economic Benefits and Costs of COVID-19 Mitigation Measures in Mexico. SSRN. 2020. doi:10.2139/ssrn.3592209
618. Isaac JN, Pablo FB-Z, Yanink C-V. Impact of false negative results of RT-PCR test for SARS-CoV-2 in COVID-19 case count as applied to Mexico. medRxiv. 2020. doi:10.1101/2020.09.17.20197038
619. Isaac N, Pablo FB-Z, Yanink C-V. Corrected Estimation of COVID-19 Cases in Mexico: A Nation-Wide Open Data Modelling Study. SSRN. 2020. doi:10.2139/ssrn.3691134
620. Isabel RF, Mary C, Wesler L, Fernet Renand L, Bethany H-G, Wanying L, et al. Adapting Lot Quality Assurance Sampling to accommodate imperfect tests: application to COVID-19 serosurveillance in Haiti. medRxiv. 2020. doi:10.1101/2020.09.11.20193052
621. Isadora CC, Eloiza DF, Janaína C da S, Guilherme S, Daisy MS, Guilherme FS. Predictive model of COVID-19 incidence and socioeconomic description of municipalities in Brazil. medRxiv. 2020. doi:10.1101/2020.06.28.20141952
622. Isaure D, Julia E, Werner P. The Distributional Consequences of Social Distancing on Poverty and Labour Income Inequality in Latin America and the Caribbean. SSRN. 2020. doi:10.2139/ssrn.3710062
623. Isaure D, Werner P. Working From Home Under COVID-19: Who Is Affected? Evidence From Latin American and Caribbean Countries. SSRN. 2020. doi:10.2139/ssrn.3610885
624. Isea R. The Transmission Dynamics of Covid-19 from a MathematicalPerspective. Obs del Conoc Rev Espec en Gestión Soc del Conoc. 2020;5: 15–23.
625. Ismael C, Silva PAIA, Silva CM da, Melo MSV de, Neto BAF, Melo JV de, et al. Universal Screening of SARS-CoV-2 of Oncology Healthcare Workers — a Brazilian experience. 2020. doi:10.1590/SciELOPreprints.293
626. Ismael I-N, Kathia GF-R, Violeta R-H, Hilda CO-B, Alfonso S-Z, Magaly P-O, et al. Ethnic disparities in COVID-19 mortality in Mexico: a cross-sectional study based on national data. medRxiv. 2020. doi:10.1101/2020.08.26.20182543
627. Itiki R, Roy Chowdhury P. Fast deployment of COVID-19 disinfectant from common ethanol of gas stations in Brazil: COVID-19 disinfectant from common ethanol. Heal Policy Technol. 2020. doi:10.1016/j.hlpt.2020.07.002
628. Ivan F, Gustavo Almeida Lopes F, Guilherme F, Pedro Ivo S. Ideology, Isolation, and Death. An Analysis of the Effects of Bolsonarism in the COVID-19 Pandemic. SSRN. 2020. doi:10.2139/ssrn.3654538
629. Iwona H, Thomas AM, Henrique HH, Ricardo S, Swapnil M, Charles W, et al. Inference of COVID-19 epidemiological distributions from Brazilian hospital data. medRxiv. 2020. doi:10.1101/2020.07.15.20154617
630. Izabela Sobiech P. Untimely Reopening? Change in the Number of New COVID-19 Cases after Reopening in One Brazilian State. SSRN. 2020. doi:10.2139/ssrn.3623930
631. Jacno Erik Ferrer C, Ernesto Sánchez H, Abel Poulout M, Germán del Río C, Daicy Figueredo S. Caracterización clínica y epidemiológica de pacientes confirmados con la COVID-19 en la provincia de Santiago de Cuba. 2020;24.
632. Jacobo G-C, Ricardo M, Ernesto L-V, Fabricia R, Rogerio L. The Effect of Management Control Systems during the COVID-19 Pandemic: Does the Market Appreciate the Breadth of Vision? SSRN. 2020. doi:10.2139/ssrn.3675688
633. Jacques N, Bruce M, Joshua C, Fabio C, Salah D, Barry D, et al. Worldwide Effectiveness of Various Non-Pharmaceutical Intervention Control Strategies on the Global COVID-19 Pandemic: A Linearised Control Model. medRxiv. 2020. doi:10.1101/2020.04.30.20085316
634. Jaime AY, Sun Ah C, Fiorella I-B, Christian M. Demographic and Geographic COVID-19 Death Risk Factors in Peru. A Nationwide Analysis. SSRN. 2020. doi:10.2139/ssrn.3648543
635. Jaime B, Max S, Jesus A, Guadalupe G, Gustavo O, Rosa MW-C, et al. Risk of infection and hospitalization by Covid-19 in Mexico: a case-control study. medRxiv. 2020. doi:10.1101/2020.05.24.20104414

636. Jairo Jesus Gomez T, Ronny Alejandro Dieguez G, Manuel Ramon Perez A, Odalis Tamayo V, Aida Elizabeth Iparraguirre T. Evaluación del nivel de conocimiento sobre COVID-19 durante la pesquisa en la población de un consultorio. 16 Abril. 2020;59.
637. Javier M-G, Jorge Oscar G-M, Jessica M-S, Rodrigo Estefano R-C, Eduardo A-A, Hector Isaac R-G, et al. Use of antivirals and antibiotics for COVID-19 in Mexico City: A Real-World Multicenter Cohort Study. medRxiv. 2020. doi:10.1101/2020.10.13.20211797
638. Javier M-G, Juan Mauricio V-Z, Addi Rhode N-C, Orietta S-B, Gerardo R-V, Francisco Javier T-L, et al. Development and Validation of the Patient History COVID-19 (PH-Covid19) Scoring System: A Multivariable Prediction Model of Death in Mexican Patients with COVID-19. medRxiv. 2020. doi:10.1101/2020.09.05.20189142
639. Javier O-C, Javier D-N, Ana S-T, Rosa P-A, Laura P-A, Juan L-L, et al. Inequality in times of pandemics: How online media are starting to treat the economic consequences of the coronavirus crisis. Prof La Inf. 2020;29. doi:10.3145/EPI.2020.JUL.03
640. Jean Franco Q-C, Evelyn F-R, Diego M, Jesús M-Z, Katia Medalith H-C, Scherlli EC, et al. Prevalence of Self-Medication During the COVID-19 Pandemic in Peru. SSRN. 2020. doi:10.2139/ssrn.3688880
641. Jeany D, Luiz Claudio N, Rinaldo Focaccia S, Gisely Cardoso de M, Estela Natacha Brandt B, José Carlos N, et al. Covid-19 automated diagnosis and risk assessment through Metabolomics and Machine-Learning. medRxiv. 2020. doi:10.1101/2020.07.24.20161828
642. Jeff H-V, David V-Z, Angela P, María Alejandra L-C. Psychometric Properties of a Spanish Version of the Fear of COVID-19 Scale in General Population of Lima, Peru. Int J Ment Health Addict. 2020. doi:10.1007/S11469-020-00354-5
643. Jeronimo CMP, Farias MEL, Val FFA, Sampaio VS, Alexandre MAA, Melo GC, et al. Methylprednisolone as Adjunctive Therapy for Patients Hospitalized With COVID-19 (Metcovid): A Randomised, Double-Blind, Phase IIb, Placebo-Controlled Trial. Clin Infect Dis. 2020. doi:10.1093/cid/ciaa1177
644. Jessica Sr. R-S, Valery Sr. D-O, Diayan C-G, Jennifer Sr. R-V, John B-O, Randol Sr. V-A, et al. Hospital practice in COVID-19 times: Perceptions of the midwifery interns in Peru. medRxiv. 2020. doi:10.1101/2020.06.05.20094482
645. Jessyka Viana Valadares F, Thiago Franco O. O avanço do COVID-19 na Amazônia Legal : Uma análise do crescimento de casos na cidade de Gurupi- Tocantins. 2020;8. doi:10.18606/2318-1419/AMAZONIA.SCI.HEALTH.V8N2P64-76
646. Jesus Arturo Sr. R-Q, Crystell Sr. G-P, German Alberto Sr. N-R, Carlos Alfonso T-Z, Oscar Israel F-B, Victor N-O, et al. Features of patients that died for COVID-19 in a Hospital in the south of Mexico: A observational cohort study. medRxiv. 2020. doi:10.1101/2020.09.21.20199117
647. Jesús Ojino S-G, Alan Omar G-V, Alondra G-B, Juan Pablo R-G, Eva J-H, Octavio G-C. Clinical Characteristics of Severe COVID-19 Patients in an ICU in Mexico. SSRN. 2020. doi:10.2139/ssrn.3605185
648. Jesús C-M, Panayotis K, Qian-Yong C, George K, Víctor V-D, Zoi R, et al. Lockdown Measures and their Impact on Single- and Two-age-structured Epidemic Model for the COVID-19 Outbreak in Mexico. medRxiv. 2020. doi:10.1101/2020.08.11.20172833
649. Jesus JG de, Sacchi C, Candido D da S, Claro IM, Sales FCS, Manuli ER, et al. Importation and early local transmission of COVID-19 in Brazil, 2020. Rev Inst Med Trop Sao Paulo. 2020;62: e30. doi:https://dx.doi.org/10.1590/s1678-9946202062030
650. Jimmy M-D, Eduardo V, Aurora M, Piedad S, Stefany P, Diana P, et al. Personal Protective Equipment, Diagnostic and Treatments Facilities for COVID-19 Patients. A Cross-Sectional Study in Brazil, Colombia and Ecuador. SSRN. 2020. doi:10.2139/ssrn.3667626
651. João Vitor A, Clarah Chaveiro MR. O que o Coronavírus tem nos tirado? Anos potenciais de vida perdidos em Minas Gerais. J Nurs Heal. 2020;10: 1–12. doi:10.15210/jonah.v10i4.19043

669. Josimar ECS. Infoveillance based on Social Sensors to Analyze the impact of Covid19 in South American Population. medRxiv. 2020. doi:10.1101/2020.04.06.20055749
670. Josimar ECS, Anabel P-B. Text Mining Approach to Analyze Coronavirus Impact: Mexico City as Case of Study. medRxiv. 2020. doi:10.1101/2020.05.07.20094466
671. Josimar ECS, Jimmy O. Covid19 Surveillance in Peru on April using Text Mining. medRxiv. 2020. doi:10.1101/2020.05.24.20112193
672. Josimar ECS, Roselyn L-M. Infoveillance to Analyze Covid19 Impact on Central America Population. medRxiv. 2020. doi:10.1101/2020.05.26.20113514
673. Juan Alonso L-A. Modeling the progression of SARS-CoV-2 infection in patients with COVID-19 risk factors through predictive analysis. medRxiv. 2020. doi:10.1101/2020.07.14.20154021
674. Juan Alonso L-A, Roha Saeed M, Bahar R, Maimoona I, Antara C. The impact of COVID-19 in diabetic kidney disease and chronic kidney disease: A population-based study. medRxiv. 2020. doi:10.1101/2020.09.12.20193235
675. Juan Carlos Sr. R, Rocio M, Danny R, Mariana C, Marcela S, Lina C, et al. Social response to early-stage government control measures of COVID-19 in Colombia: population survey, April 8-20 2020. medRxiv. 2020. doi:10.1101/2020.06.18.20135145
676. Juan David R, Carolina F, Marina M, Carolina H, Adriana C, Sergio G, et al. The arrival and spread of SARS-CoV2 in Colombia. medRxiv. 2020. doi:10.1101/2020.06.11.20125799
677. Juan David R, Marina M, Carolina H, Carolina F, Sergio G, Angelica R, et al. Genetic diversity among SARS-CoV2 strains in South America may impact performance of Molecular detection. medRxiv. 2020. doi:10.1101/2020.06.18.20134759
678. Juan Felipe Medina M, Manuel Eduardo Cortes C, Manuel Cortes I, Annia del Carmen Perez F, Marianelis Manzano C. Study on predictive models for COVID-19 in Cuba. 2020;18.
679. Juan Felipe Medina M, Manuel Eduardo Cortés C, Manuel Cortés I, Annia del Carmen Pérez F, Marianelis Manzano C. Estudio sobre modelos predictivos para la COVID-19 en Cuba. 2020;18.
680. Juan Fidel O-R, Horacio Rendon A, Luis Adrian De J-G, Jose Manuel Reyes R, Arely Montserrat Espinoza O, Luis Antonio Ochoa R, et al. Serum lipid profile changes and their clinical diagnostic significance in COVID-19 Mexican Patients. medRxiv. 2020. doi:10.1101/2020.08.24.20169789
681. Juan Pablo G, Stefano MB. Non-communicable diseases and inequalities increase risk of death among COVID-19 patients in Mexico. medRxiv. 2020. doi:10.1101/2020.05.27.20115204
682. Juan AGC. SEI1I2HRSVM model applied to the coronavirus pandemic (COVID-19) in Paraguay. arXiv Popul Evol. 2020.
683. Juan F-S, Jose Enrique Olvera V, Juan Javier G-B, Guadalupe C-V, Juan Paulo S-H, Joaquin P-O. The Hybrid Forecasting Method SVR-ESAR forCovid-19. medRxiv. 2020. doi:10.1101/2020.05.20.20103200
684. Juan MCL. Sars-Cov-2 in Argentina: Following Virus Spreading using Granger Causality. medRxiv. 2020. doi:10.1101/2020.10.06.20207993
685. Juan MCL. Sars-Cov-2 in Argentina: Lockdown, Mobility, and Contagion. medRxiv. 2020. doi:10.1101/2020.10.03.20203232
686. Julian Alfredo F-N, Jhon AG-G, Alvaro Javier I-V. Multimorbidity patterns among COVID-19 deaths: considerations for a better medical practice. medRxiv. 2020. doi:10.1101/2020.07.28.20163816
687. Juliana Marques de A, Roberta Andrade de S, Livia Gomes V-M, Landeira-Fernandez J, Alberto F. Effects of physical activity and exercise on well-being in the context of the Covid-19 pandemic. medRxiv. 2020. doi:10.1101/2020.06.08.20125575
688. Juliane Fonseca O, Daniel CPJ, Rafael VV, Moreno SR, Matheus FT, Nivea B da S, et al. Evaluating the burden of COVID-19 on hospital resources in Bahia, Brazil: A modelling-based analysis of 14.8 million individuals. medRxiv. 2020. doi:10.1101/2020.05.25.20105213
689. Julio Cesar Sampaio D, Wellington Betencurte da S, Jose Mir Justino da C. Monitoring and forecasting the number of reported and unreported cases of the COVID-19 epidemic in Brazil using Particle Filter. medRxiv. 2020. doi:10.1101/2020.05.27.20115212

690. Junior J, Freire G, Gonçalves M, Oliveira D, Moraes J, Fidelix Y. Do Age and Symptoms of Anxiety, Stress and Depression Predict Risk Behaviors for Eating Disorders in Brazilian Men and Women During the Covid-19 Pandemic? ResearchSquare. 2020. doi:10.21203/rs.3.rs-100005/v1
691. Junqueira-Kipnis AP, dos Anjos LRB, Barbosa LCS, da Costa AC, Borges KCM, Cardoso ARO, et al. BCG revaccination of health workers in Brazil to improve innate immune responses against COVID-19: A structured summary of a study protocol for a randomised controlled trial. Trials. 2020;21: 881. doi:http://dx.doi.org/10.1186/s13063-020-04822-0
692. Kalbusch A, Henning E, Brikalski MP, Luca FVD, Konrath AC. Impact of coronavirus (COVID-19) spread-prevention actions on urban water consumption. Resour Conserv Recycl. 2020;163: 105098. doi:http://dx.doi.org/10.1016/j.resconrec.2020.105098
693. Kammar-Garcia A, Vidal-Mayo J de J, Vera-Zertuche JM, Lazcano-Hernandez M, Vera-Lopez O, Segura-Badilla O, et al. IMPACT OF COMORBIDITIES IN MEXICAN SARS-COV-2-POSITIVE PATIENTS: A RETROSPECTIVE ANALYSIS IN A NATIONAL COHORT. Rev Invest Clin. 2020;72: 151–158. doi:https://dx.doi.org/10.24875/RIC.20000207
694. Karina C, Marcos Montani C, Claudia Renata dos Santos B, Lourdes Conceicao M, Arthur C, Evaldo E, et al. COVID-19 Seroprevalence in Baixada Santista Metropolitan Area, Sao Paulo, Brazil. medRxiv. 2020. doi:10.1101/2020.08.28.20184010
695. katherine L-D, Christian Julian Villabona A, Jose AUC, Carlos Franco M, Diego Alejandro A-D, Liz SV-A, et al. Genomic epidemiology of SARS-CoV-2 in Colombia. medRxiv. 2020. doi:10.1101/2020.06.26.20135715
696. Katherine S-R, Lenin G-B, Jhon G, Fernanda S-G, Raul F, Andres Sr. L-C, et al. Interim Analysis of Pandemic Coronavirus Disease 2019 (COVID-19) and the SARS-CoV-2 virus in Latin America and the Caribbean: Morbidity, Mortality and Molecular Testing Trends in the Region. medRxiv. 2020. doi:10.1101/2020.04.25.20079863
697. Kelly Cristine Lemos S, Maria Eliara Gomes L. A INSERÇÃO DE DUAS PSICÓLOGAS RESIDENTES EM TEMPOS DE COVID-19. 2020;14.
698. Kenzo A, Eduardo AU, Rodrigo V, Rodrigo W. The effect of COVID-19 on the economy: evidence from an early adopter of localized lockdowns. medRxiv. 2020. doi:10.1101/2020.09.21.20198887
699. Kerr L, Kendall C, Silva AAMD, Aquino EML, Pescarini JM, Almeida RLF, et al. COVID-19 in Northeast Brazil: achievements and limitations in the responses of the state governments. Cien Saude Colet. 2020;25: 4099–4120. doi:http://dx.doi.org/10.1590/1413-812320202510.2.28642020
700. Kevin SR, Sebastian D, Gyan B. The transmission of SARS-CoV-2 is likely comodulated by temperature and by relative humidity. medRxiv. 2020. doi:10.1101/2020.05.23.20111278
701. Khondoker Nazmoon N, Islam Md R. HAS COUNTRYWIDE LOCKDOWN WORKED AS A FEASIBLE MEASURE IN BENDING THE COVID-19 CURVE IN DEVELOPING COUNTRIES? medRxiv. 2020. doi:10.1101/2020.06.23.20138685
702. Klôh VP, Silva GD, Ferro M, Araújo E, de Melo CB, de Andrade Lima JRP, et al. The virus and socioeconomic inequality: An agent-based model to simulate and assess the impact of interventions to reduce the spread of COVID-19 in Rio de Janeiro, Brazil. Brazilian J Heal Rev. 2020;3: 3647–3673.
703. Kosugi EM, Lavinsky J, Romano FR, Fornazieri MA, Luz-Matsumoto GR, Lessa MM, et al. Incomplete and late recovery of sudden olfactory dysfunction in COVID-19. Braz J Otorhinolaryngol. 2020. doi:https://dx.doi.org/10.1016/j.bjorl.2020.05.001
704. Kowalski LP, Imamura R, Castro Junior G, Marta GN, Chaves ALF, Matos LL, et al. Effect of the COVID-19 Pandemic on the Activity of Physicians Working in the Areas of Head and Neck Surgery and Otorhinolaryngology. Int Arch Otorhinolaryngol. 2020;24: e258–e266. doi:10.1055/s-0040-1712169
705. Krüger-Malpartida H, Pedraz-Petrozzi B, Arevalo-Flores M, Salmavides-Cuba F, Anculle-Arauco V, Dancuart-Mendoza M. Effects on the mental health after COVID-19 lockdown period: results from a population survey study in Lima, Peru. ResearchSquare. 2020. doi:10.21203/rs.3.rs-81117/v1

Onicio Batista Leal N, Francisco Assis S, Julia Yeri L, Jones A, Wayner Vieira S. Prioritizing COVID-19 tests based in Participatory Surveillance and Spatial Scanning. medRxiv. 2020. doi:10.1101/2020.05.25.20109058
715. Leandro Pereira G, Andre Vinicius G, Matheus Pacheco de A, Lucas Alexandre P, Ana Cristina V, Roberto Z, et al. ESTIMATING UNDERDIAGNOSIS OF COVID-19 WITH NOWCASTING AND MACHINE LEARNING: EXPERIENCE FROM BRAZIL. medRxiv. 2020. doi:10.1101/2020.07.01.20144402
716. Leandro Simões P, Nobuiuki Costa I. O surto da COVID-19 e as respostas da administração municipal: munificência de recursos, vulnerabilidade social e eficácia de ações públicas. 2020.
717. Leila F D, Igor T P, Leonardo S.L B, Janaina F M, Guilherme F.G. De S, Joao Gabriel M G, et al. App-based symptom tracking to optimize SARS-CoV-2 testing strategy using machine learning. medRxiv. 2020. doi:10.1101/2020.09.01.20186049
718. Lemos ACB, do Espirito Santo DA, Salvetti MC, Gilio RN, Agra LB, Pazin-Filho A, et al. Therapeutic versus prophylactic anticoagulation for severe COVID-19: a randomized phase II clinical trial (HESACOVID). Thromb Res. 2020;196: 359-366. doi:10.1016/j.thromres.2020.09.026
719. Lemos DRQ, D'Angelo SM, Farias LABG, Almeida MM, Gomes RG, Pinto GP, et al. Health system collapse 45 days after the detection of COVID-19 in Ceara, Northeast Brazil: a preliminary analysis. Rev Soc Bras Med Trop. 2020;53: e20200354. doi:https://dx.doi.org/10.1590/0037-8682-0354-2020
720. Lenisse MR, Lilibeth O, Maxwell A, Yenifel L, Wilma R, Pablo J de JR. Misinformation on COVID-19 origin and social distancing: A cross-sectional study. medRxiv. 2020. doi:10.1101/2020.10.06.20207894
721. Leonardo JB, Felipe S, Hugo ES. Public Transport Policies after COVID-19 Confinement. SSRN. 2020. doi:10.2139/ssrn.3693098

722. Leung C. The younger the milder clinical course of COVID-19: Even in newborns? Pediatr Allergy Immunol. 2020. doi:http://dx.doi.org/10.1111/pai.13371
723. Lewis FB, Carlos Augusto Jr. P, Claudia MMA, Alfredo Jr. M, Tassila S, Cesar de A-N, et al. COVID-19 herd immunity in the Brazilian Amazon. medRxiv. 2020. doi:10.1101/2020.09.16.20194787
724. Liana RW, Clare W. Do Men and Women Lockdown Differently? An Examination of Panamas COVID-19 Sex-Segregated Social Distancing Policy. medRxiv. 2020. doi:10.1101/2020.06.30.20143388
725. Lima DLF, Dias AA, Rabelo RS, Cruz ID da, Costa SC, Nigri FMN, et al. Covid-19 in the State of Ceara: behaviors and beliefs in the arrival of the pandemic. Cien Saude Colet. 2020;25: 1575–1586. doi:https://dx.doi.org/10.1590/1413-81232020255.07192020
726. Lima DLF, Veras PJL, Marques TM, Costa SC, Santos HPG dos, Neri JR. Transmission care: what led Ceará to be the epicenter of COVID-19? Rev bras promoç saúde. 2020;33: 1–14.
727. Lima E de P, Vasconcelos AG, Corrêa LRT, Batista AG. Baixas na linha de frente: absenteísmo entre bombeiros durante o combate à pandemia da COVID-19. Rev Bras Saúde Ocup. 2020;45. doi:10.1590/2317-6369000016420
728. Lima FET, Albuquerque NLS de, Florencio S de SG, Fontenele MGM, Queiroz APO, Lima GA, et al. Time interval between onset of symptoms and COVID-19 testing in Brazilian state capitals, August 2020. 2020.
729. Lima KC de, Medeiros A de A, Barbosa IR. Trend analysis of cases and deaths by Coronavirus Disease-2019 (COVID-19) in the main affected countries and in Brazil: an analysis of the first 50 days of the pandemic. J Heal Biol Sci. 2020;8.
730. Lima L. Modeling and forecasting of epidemic spreading of the SARS-CoV-2 based in the nonlinear stochastic dynamics. ResearchSquare. 2020. doi:10.21203/rs.3.rs-29533/v1
731. Lima L. Modelling based in the stochastic dynamics for the time evolution of the COVID-19. ResearchSquare. 2020. doi:10.21203/rs.3.rs-23014/v1
732. Lima L. Stochastic analysis using public data for forecasting of epidemic spreading of the novel coronavirus disease. ResearchSquare. 2020. doi:10.21203/rs.3.rs-78321/v1
733. Lima MA, Silva MTT, Oliveira R V, Soares CN, Takano CL, Azevedo AE, et al. Smell dysfunction in COVID-19 patients: More than a yes-no question. J Neurol Sci. 2020;418: 117107. doi:http://dx.doi.org/10.1016/j.jns.2020.117107
734. Lima-Costa MF, Macinko J, Andrade FB, Souza Junior PRB, Vasconcellos MTL, Oliveira CM. ELSI-COVID-19 initiative: methodology of the telephone survey on coronavirus in the Brazilian Longitudinal Study of Aging. Cad Saude Publica. 2020;36Suppl 3: e00183120. doi:http://dx.doi.org/10.1590/0102-311X00183120

Lima-Costa MF, Mambrini JVM, Andrade FB, Peixoto SW V, Macinko J. Social distancing, use of face masks and hand washing among participants in the Brazilian Longitudinal Study of Aging: the ELSI-COVID-19 initiative. Cad Saude Publica. 2020;36Suppl 3: e00193920. doi:http://dx.doi.org/10.1590/0102-311X00193920
735. Lincoln Luis S, Amanda Carvalho D, Pedro Henrique I, Guilherme Luiz Rodrigues R, Gabriel Antonio Fernandes M, Iago Amado Peres G, et al. Brazil Health Care System preparation against COVID-19. medRxiv. 2020. doi:10.1101/2020.05.09.20096719
736. Lino DODC, Barreto R, Souza FDD, Lima CJMD, Silva Junior GBD. Impact of lockdown on bed occupancy rate in a referral hospital during the COVID-19 pandemic in northeast Brazil. Brazilian J Infect Dis. 2020;24: 466–469. doi:http://dx.doi.org/10.1016/j.bjid.2020.08.002
737. Lins S, Aquino S. Development and initial psychometric properties of a panic buying scale during COVID-19 pandemic. Heliyon. 2020;6: e04746. doi:10.1016/j.heliyon.2020.e04746
738. Lins-Filho PC, Araújo MMS de, Macêdo TS de, Melo MCF de, Ferreira AKA, Silva ELMS da, et al. The impact of socioeconomic vulnerability on COVID-19 outcomes and social distancing in Brazil. 2020.

Wladimir L, Jose Dias do N, Jaber B, Leandro de A, Pedro Paulo C, Ion de A. COVID-19 pandemics modeling with SEIR(+CAQH), social distancing, and age stratification. The effect of vertical confinement and release in Brazil. medRxiv. 2020. doi:10.1101/2020.04.09.20060053
768. Macarena P-L. Tweets de la autoridad sanitaria en Chile en los albores de la crisis del coronavirus. Rev Esp Comun en Salud. 2020;11: 117–127. doi:10.20318/RECS.2020.5447
769. Macarena RV, Anne P, Inia P, Maria S, Mario B, Mabel A, et al. Clinical and Epidemiological Characteristics of the First Month of the Covid-19 Pandemic in Chile. medRxiv. 2020. doi:10.1101/2020.08.11.20171975
770. Machado CJ, Pereira CC de A, Viana B de M, Oliveira GL, Melo DC, Carvalho JFMG de, et al. Estimates of the impact of COVID-19 on mortality of institutionalized elderly in Brazil. Estim impacto da COVID-

19 na mortalidade idosos Inst no Bras. 2020;25: 3437–3444. doi:https://dx.doi.org/10.1590/1413-81232020259.14552020
771. Machado MH, Pereira EJ, Ximenes Neto FRG, Wermelinger MC de MW. Enfermagem em tempos de COVID-19 no Brasil: um olhar da gestão do trabalho. Enferm foco. 2020;11: 32–39.
772. Maciel EL, Jabor P, Goncalves Junior E, Tristao-Sa R, Lima R de CD, Reis-Santos B, et al. Factors associated with COVID-19 hospital deaths in Espirito Santo, Brazil, 2020. Fatores Assoc ao obito Hosp por COVID-19 no Espirito Santo, 2020. 2020;29: e2020413. doi:https://dx.doi.org/10.1590/S1679-49742020000400022
773. Maciel EL, Jabor P, Júnior EG, Tristão-Sá R, Lima R de CD, Reis-Santos B, et al. Mortality in patients admitted to hospital by COVID-19 in Espírito Santo, Brazil, 2020. 2020.
774. Maciel FBM, Santos HLPCD, Carneiro RADS, Souza EA, Prado NMBL, Teixeira CFS. Community health workers: reflections on the health work process in Covid-19 pandemic times. Cien Saude Colet. 2020;25: 4185–4195. doi:http://dx.doi.org/10.1590/1413-812320202510.2.28102020
775. Maciel JAC, Castro-Silva II, Farias MR de. Initial analysis of the spatial correlation between the incidence of COVID-19 and human development in the municipalities of the state of Ceara in Brazil. Rev Bras Epidemiol. 2020;23: e200057. doi:https://dx.doi.org/10.1590/1980-549720200057
776. Macinko J, Woolley NO, Seixas B V, Andrade FB, Lima-Costa MF. Health care seeking due to COVID-19 related symptoms and health care cancellations among older Brazilian adults: the ELSI-COVID-19 initiative. Cad Saude Publica. 2020;36Suppl 3: e00181920. doi:http://dx.doi.org/10.1590/0102-311X00181920
777. Madhuvanti MM, Selvi MJ, Christina H, Natalie G, Heather H, Kim RQ, et al. COVID-19 containment in the Caribbean: the experience of Small Island Developing States. medRxiv. 2020. doi:10.1101/2020.05.27.20114538
778. Mækelæ MJ, Reggev N, Dutra N, Tamayo RM, Silva-Sobrinho RA, Klevjer K, et al. Perceived efficacy of COVID-19 restrictions, reactions and their impact on mental health during the early phase of the outbreak in six countries. R Soc open Sci. 2020;7: 200644. doi:10.1098/rsos.200644
779. Magalhaes JJFD, Mendes RPG, Silva CTAD, Silva SJRD, Guarines KM, Pena L. Epidemiological and clinical characteristics of the first 557 successive patients with COVID-19 in Pernambuco state, Northeast Brazil. Travel Med Infect Dis. 2020;38: 101884. doi:http://dx.doi.org/10.1016/j.tmaid.2020.101884
780. Magalhaes M, Arruda do Rego L, Vieira de Reboucas C, de Brito Alves R, Adami F, Cruz FJSM. Clinical and epidemiologic aspects of patients with cancer and COVID-19 in a Brazilian cancer center. Ann Oncol. 2020;31: S1027. doi:http://dx.doi.org/10.1016/j.annonc.2020.08.1834
781. Malagón-Rojas J, Gómez-Rendón C, Parra EL, Almentero J, Palma R, López R, et al. SARS-CoV-2 and RT-PCR in asymptomatic patients: Results of a cohort of workers at El Dorado International Airport in Bogotá, 2020. Biomedica. 2020;40: 166–172. doi:10.7705/biomedica.5802
782. Malgor RD, Sobreira ML, Mouawad NJ, Johnson AP, Wohlauer M V, Coogan SM, et al. Brazilian vascular surgeons experience during the coronavirus (COVID-19) pandemic. Vascular. 2020. doi:http://dx.doi.org/10.1177/1708538120954961
783. Malta DC, Szwarcwald CL, Barros MB de A, Gomes CS, Machado IE, Souza Junior PRB de, et al. The COVID-19 Pandemic and changes in adult Brazilian lifestyles: a cross-sectional study, 2020. A pandemia da COVID-19 e as mudancas no estilo vida dos Bras adultos um Estud transversal, 2020. 2020;29: e2020407. doi:https://dx.doi.org/10.1590/S1679-49742020000400026
784. Mamani-Benito O, Apaza Tarqui EE, Carranza Esteban RF, Rodriguez-Alarcon JF, Mejía CR. Perceived job insecurity in employment due to the impact of COVID-19: validation of an instrument on Peruvian workers (LABOR-PE-COVID-19). Rev Asoc Esp Espec Med Trab. 2020;29: 184–193.
785. Manchein C, Brugnago EL, da Silva RM, Mendes CFO, Beims MW. Strong correlations between power-law growth of COVID-19 in four continents and the inefficiency of soft quarantine strategies. Chaos. 2020;30: 41102. doi:https://dx.doi.org/10.1063/5.0009454

786. Manrique-Abril FG, Agudelo-Calderon CA, González-Chordá VM, Gutiérrez-Lesmes O, Téllez-Piñerez CF, Herrera-Amaya G. SIR model of the COVID-19 pandemic in Colombia. Rev salud pública. 2020;22: e185977–e185977.
787. Manrique-Hernández EF, Moreno-Montoya J, Hurtado-Ortiz A, Prieto-Alvarado FE, Idrovo Á J. Performance of the Colombian surveillance system during the COVID-19 pandemic: A rapid evaluation of the first 50 days. Biomedica. 2020;40: 96–103. doi:10.7705/biomedica.5582
788. Manuel Adrian A-Z, Andreu C-G, Esteban H-V, Mario S-C, Jorge XV-H. The SARS-CoV-2 epidemic outbreak: a review of plausible scenarios of containment and mitigation for Mexico. medRxiv. 2020. doi:10.1101/2020.03.28.20046276
789. Manuel A, Santiago V, Fernando V-E, Ricardo S-R, Gonzalo PB, Jonas C, et al. SARS-CoV-2 Detection in Sewage in Santiago, Chile - Preliminary results. medRxiv. 2020. doi:10.1101/2020.07.02.20145177
790. Maquilon C, Gongora J, Antolini M, Alvarado B, Valdes N, Benavente A, et al. Risk factors on admission and condition at discharge of 529 consecutive COVID-19 patients at a tertiary care center in Santiago, Chile. ResearchSquare. 2020. doi:10.21203/rs.3.rs-71187/v1
791. Marcel G, Mirko B-L, Magdalena B, Leonardo JB. COVID-19: Short-Term Forecast of ICU Beds in Times of Crisis. SSRN. 2020. doi:10.2139/ssrn.3693447
792. Marcelo Domingos M. TAMING COVID-19 EPIDEMIC IN SAO PAULO WITH ALOGISTIC MODEL AND NON-PHARMACEUTICAL MEASURES. medRxiv. 2020. doi:10.1101/2020.05.06.20093666
793. Marcelo Henrique Santos P, Duschinka Ribeiro Duarte G, Cassia D, Matheus Filgueira B, Filipe Zimmer D, Lais Ceschini M, et al. Multiple introductions followed by ongoing community spread of SARS-CoV-2 at one of the largest metropolitan areas in the Northeast of Brazil. medRxiv. 2020. doi:10.1101/2020.08.25.20171595
794. Marcelo M, Mehran S, Cesar C, Joao AMG. ASSESSING THE EFFICIENCY OF DIFFERENT CONTROL STRATEGIES FOR THE COVID-19 EPIDEMIC. Electron J Differ Equations. 2020.

César C, João AMG, Marcelo M, Mehran S. Assessing the Efficiency of Different Control Strategies for the Coronavirus (COVID-19) Epidemic. arXiv Popul Evol. 2020.
795. Marcia CC, Lucas Resende de C, Taylor C, Rebecca K, Giovanny VAF, Eduardo Marques M, et al. Demand for hospitalization services for COVID-19 patients in Brazil. medRxiv. 2020. doi:10.1101/2020.03.30.20047662
796. Marcos Montani C, Monica M, Bruno B, Estela Capelas B, Antonio Pires B. Positivity of SARS-CoV-2, by RT-PCR among workers of a Public Hospital in the city of Santos, SP, Brazil 2020. medRxiv. 2020. doi:10.1101/2020.06.30.20143529
797. Marcos Roberto M, Ricardo Vicente F, Rogerio HT, Luiza A, Michael Richard D, Eric MD. Detecting space-time clusters of COVID-19 in Brazil: mortality, inequality, socioeconomic vulnerability, and the relative risk of the disease in Brazilian municipalities. medRxiv. 2020. doi:10.1101/2020.06.14.20131102
798. Marcos AC, Antonio C, Christen JA. Forecasting hospital demand in metropolitan areas during the current COVID-19 pandemic and estimates of lockdown-induced 2nd waves. medRxiv. 2020. doi:10.1101/2020.07.16.20155721
799. Marcos M, Pablo Rodriguez M, Carlos M-G, Victor L, Francisco G. Mathematical estimation of COVID-19 prevalence in Latin America. medRxiv. 2020. doi:10.1101/2020.06.09.20126326
800. Margaret S, Carlos D, Matthew BBM, Adrian G, Miguel AS-M, Michael KM, et al. The association of UV with rates of COVID-19 transmission and deaths in Mexico: the possible mediating role of vitamin D. medRxiv. 2020. doi:10.1101/2020.05.25.20112805
801. María Alejandra B, Carolina V, Ana Rita S, Josefa H, Flavio MM, Francesco P. COVID-19 Pandemic in Five Countries of Latin America. SSRN. 2020. doi:10.2139/ssrn.3663292
802. Maria de Fátima Rodrigues D, Maira Freire C, Karen Saori Shiraishi S, Carolina Rocha Brito M, Alessandro Cavalcanti L, Maria Fernanda Bádue P, et al. Pediatric Heart in COVID-19: New Insights

from a Systematic Echocardiographic Study in a Tertiary Brazilian Hospital. SSRN. 2020. doi:10.2139/ssrn.3658311
803. María Elvira B, Luis R, Nicole Le C, Constanza M-V, María Elena C, Marcela F, et al. Early Anti-SARS-CoV-2 Convalescent Plasma in Patients Admitted for COVID-19: A Randomized Phase II Clinical Trial. medRxiv. 2020. doi:10.1101/2020.09.17.20196212
804. Maria Gerusa Brito A, Francisco Isaac Fernandes G, Camila Siqueira Silva C, Leticia Pinho Maia Paixao de M, Silmara Aparecida Milori C. Where do Brazilian dental students seek information about COVID-19? medRxiv. 2020. doi:10.1101/2020.08.24.20179614
805. Maria Gerusa Brito A, Francisco Isaac Fernandes G, Leticia Pinho Maia Paixao de M, Silmara Aparecida Milori C. Brazilian dental students and COVID-19: a survey on knowledge and perceptions. medRxiv. 2020. doi:10.1101/2020.07.28.20163964
806. Mariangela S, Aluisio B, Bernardo H, Lucia P, Gabriel V, Odir D, et al. Repeated population-based surveys of antibodies against SARS-CoV-2 in Southern Brazil. medRxiv. 2020. doi:10.1101/2020.05.01.20087205
807. Marinelli NP, Albuquerque LP de A, Sousa IDB de, Batista FM de A, Mascarenhas MDM, Rodrigues MTP. Evolution of indicators and service capacity at the beginning of the COVID-19 epidemic in Northeast Brazil, 2020. Epidemiol e Serv saude Rev do Sist Unico Saude do Bras. 2020;29: e2020226. doi:https://dx.doi.org/10.5123/S1679-49742020000300008
808. Marín-Sánchez A. [Basic clinical characteristics in the first 100 fatal cases of COVID-19 in Colombia]. Rev Panam Salud Publica. 2020;44: e87. doi:10.26633/RPSP.2020.87
809. Mario Arturo Ruiz E. Can COVID-19 Shows Income Inequality? SSRN. 2020. doi:10.2139/ssrn.3638160
810. Mario Arturo Ruiz E. How COVID-19 Quarantine(s) Can Generate Poverty? SSRN. 2020. doi:10.2139/ssrn.3580703
811. Mario Arturo Ruiz E. Is Poverty the Best Allied of COVID-19 in Developing Countries? The Case of Guatemala. SSRN. 2020. doi:10.2139/ssrn.3645045
812. Mario S-C, Manuel Adrian A-Z, Jorge XV-H. Flattening the curve and the effect of atypical events on mitigation measures in Mexico: a modeling perspective. medRxiv. 2020. doi:10.1101/2020.05.21.20109678
813. Marisa S, Tereza Fellipe G, Helena Cramer R, Fabiana M, Adriana C, Izabella B, et al. Epidemic Curve of Contamination in a Hospital That Served as Sentinel of the Spread of the SARS-Cov-2 Epidemic in the City of Rio de Janeiro. medRxiv. 2020. doi:10.1101/2020.10.19.20215079
814. Marques C, Kakehasi AM, Gomides APM, Paiva EDS, Dos Reis Neto ET, Pileggi GCS, et al. ReumaCoV Brasil Registry: a Brazilian cohort of Patients with Immuno-mediated Chronic Inflammatory Diseases Infected by SARS-CoV-2. JMIR Res Protoc. 2020. doi:10.2196/24357

Marques C, Kakehasi A, Gomides A, Martins D, Paiva E, Neto E, et al. ReumaCoV Brasil Registry: Brazilian Study of Patients with Immuno-mediated Chronic Inflammatory Diseases Infected by SARS-CoV-2 . ResearchSquare. 2020. doi:10.21203/rs.3.pex-1104/v1
815. Marques NP, Silveira DMM, de Lima Martelli PJ, Martelli DRB, Hilan de Lucena E, Martelli-Junior H. Brazilian Oral Medicine and public health system: the enormous impact of the COVID-19 Era. Oral Dis. 2020. doi:http://dx.doi.org/10.1111/odi.13677
816. Marquez S, Prado-Vivar B, Guadalupe JJ, Gutierrez B, Becerra-Wong M, Jibaja M, et al. Metagenome of a bronchoalveolar lavage fluid sample from a confirmed COVID-19 case in quito, ecuador, obtained using oxford nanopore MinION technology. Microbiol Resour Announc. 2020;9: e00996-20. doi:http://dx.doi.org/10.1128/MRA.00996-20
817. Marquez S, Prado-Vivar B, Guadalupe JJ, Gutierrez Granja B, Jibaja M, Tobar M, et al. Genome sequencing of the first SARS-CoV-2 reported from patients with COVID-19 in Ecuador. medRxiv Prepr Serv Heal Sci. 2020. doi:https://dx.doi.org/10.1101/2020.06.11.20128330

818. Marquiony S, Eudes ESL, Kenio CL, Andiara ACB, Monica BB, Diego B. Survival and predictors of deaths of patients hospitalized due to COVID-19 from a retrospective and multicenter cohort study in Brazil. medRxiv. 2020. doi:10.1101/2020.06.07.20125047
819. Martin RS, Soledad EG, Lorena R, Noelia SF, Veronica G, Patricia MC, et al. EFFECT OF CONVALESCENT PLASMA ON MORTALITY IN PATIENTS WITH COVID-19 PNEUMONIA. medRxiv. 2020. doi:10.1101/2020.10.08.20202606
820. Martinez D, Sarria GJ, Wakefield D, Flores C, Malhotra S, Li B, et al. COVID's Impact on Radiation Oncology: A Latin American Survey Study. Int J Radiat Oncol Biol Phys. 2020;108: 374–378. doi:https://dx.doi.org/10.1016/j.ijrobp.2020.06.058
821. Martinez EZ, Silva FM, Morigi TZ, Zucoloto ML, Silva TL, Joaquim AG, et al. Physical activity in periods of social distancing due to COVID-19: a cross-sectional survey. Cien Saude Colet. 2020;25: 4157–4168. doi:http://dx.doi.org/10.1590/1413-812320202510.2.27242020
822. Martinez EZ, Aragon DC, Nunes AA. Long-term forecasts of the COVID-19 epidemic: a dangerous idea. Rev Soc Bras Med Trop. 2020;53: e20200481. doi:https://dx.doi.org/10.1590/0037-8682-0481-2020
823. Martinez EZ, Aragon DC, Nunes AA. Short-term forecasting of daily COVID-19 cases in Brazil by using the Holt's model. Rev Soc Bras Med Trop. 2020;53: e20200283. doi:https://dx.doi.org/10.1590/0037-8682-0283-2020
824. Martínez L, Valencia I, Trofimoff V. Subjective wellbeing and mental health during the COVID-19 pandemic: Data from three population groups in Colombia. Data Br. 2020;32: 106287. doi:10.1016/j.dib.2020.106287
825. Martinez-Fierro ML, Rios-Jasso J, Garza-Veloz I, Reyes-Veyna L, Cerda-Luna RM, Duque-Jara I, et al. The role of close contacts of COVID-19 patients in the SARS-CoV-2 transmission: an emphasis on the percentage of nonevaluated positivity in Mexico. Am J Infect Control. 2020. doi:http://dx.doi.org/10.1016/j.ajic.2020.10.002
826. Martins CM, Gomes RZ, Muller EV, Borges PK de O, Coradassi CE, Montiel EM da S. Predictive model for covid-19 incidence in a medium-sized municipality in brazil (ponta grossa, paraná). Texto & Context enferm. 2020;29: e20200154–e20200154.
827. Martins LD, da Silva I, Batista W V, Andrade MDF, Freitas EDD, Martins JA. How socio-economic and atmospheric variables impact COVID-19 and influenza outbreaks in tropical and subtropical regions of Brazil. Environ Res. 2020;191: 110184. doi:http://dx.doi.org/10.1016/j.envres.2020.110184
828. Martins-Filho PR, de Souza Araujo AA, Quintans-Junior LJ, Santos VS. COVID-19 fatality rates related to social inequality in Northeast Brazil: a neighborhood-level analysis. J Travel Med. 2020. doi:http://dx.doi.org/10.1093/jtm/taaa128
829. Martins-Filho PR, Souza Araujo AA, Quintans-Junior LJ, Santos VS. Digital public interest on coronavirus information and social distancing: a Brazilian nationwide analysis using an infodemiology approach. J Travel Med. 2020. doi:http://dx.doi.org/10.1093/jtm/taaa154
830. Maskin LP, Olarte GL, Palizas Jr. F, Velo AE, Lurbet MF, Bonelli I, et al. High dose dexamethasone treatment for Acute Respiratory Distress Syndrome secondary to COVID-19: a structured summary of a study protocol for a randomised controlled trial. Trials. 2020;21: 743. doi:https://dx.doi.org/10.1186/s13063-020-04646-y
831. Massuda A, Tasca R, Malik AM. Use of private hospital beds by public health systems in response to COVID-19. 2020.
832. Matheus TB, Fernando ML-T, Marco Tulio PC, Ricardo D, Jose Alexandre FD-F. Social distancing and movement constraint as the most likely factors for COVID-19 outbreak control in Brazil. medRxiv. 2020. doi:10.1101/2020.05.02.20088013
833. Matos A da R, Motta FC, Caetano BC, Ogrzewalska M, Garcia CC, Lopes JCO, et al. Identification of SARS-CoV-2 and additional respiratory pathogens cases under the investigation of COVID-19 initial phase in a Brazilian reference laboratory. Mem Inst Oswaldo Cruz. 2020;115: e200232. doi:https://dx.doi.org/10.1590/0074-02760200232

868. Miñan-Tapia A, Conde-Escobar A, Calderon-Arce D, Cáceres-Olazo D, Peña-Rios AJ, Donoso-Romero RC. Associated factors to self-medication with drugs related to COVID-19 in health science students from a peruvian city. 2020.
869. Miot HA, Ianhez M, Ramos PM. Self-reported cutaneous manifestations in 1,429 Brazilian COVID-19 patients. J Eur Acad Dermatol Venereol. 2020. doi:http://dx.doi.org/10.1111/jdv.17024
870. Mirleide C dos S, Edivaldo Costa Jr. S, Jessylene AF, Sandro PS, Michel PCS, Jedson FC, et al. MOLECULAR EPIDEMIOLOGY TO UNDERSTAND THE SARS-CoV-2 EMERGENCE IN THE BRAZILIAN AMAZON REGION. medRxiv. 2020. doi:10.1101/2020.09.04.20184523
871. Mirza N, Hasnaoui JA, Naqvi B, Rizvi SKA. The impact of human capital efficiency on Latin American mutual funds during Covid-19 outbreak. Swiss J Econ Stat. 2020;156: 16. doi:10.1186/s41937-020-00066-6
872. Moiseis Santos C, Geraldo LD, Eveliny Barroso S. Using the infection fatality rate to predict the evolution of Covid-19 in Brazil. medRxiv. 2020. doi:10.1101/2020.07.01.20144279
873. Mondal MRH, Bharati S, Podder P. Data analytics for novel coronavirus disease. Informatics Med Unlocked. 2020;20: 100374. doi:http://dx.doi.org/10.1016/j.imu.2020.100374
874. Monica Santos de Sr. M, Lysandro Pinto Sr. B, Daniela Raguer Valadao Sr. S, Aline Fagundes Sr. M, Jose Melquiades De Rezende Sr. N, Anderson Alves Sr. R, et al. Anti-SARS-CoV-2 IgM and IgG antibodies in health workers in Sergipe, Brazil. medRxiv. 2020. doi:10.1101/2020.09.24.20200873
875. Montanheiro L, Dartora C. Characterizing COVID-19 epidemics dissemination and previsions for Curitiba,Brazil using a modified SIR model. 2020.
876. Montano W, Gushiken E. Lima soundscape before confinement and during curfew. Airplane flights suppressions because of Peruvian lockdown. J Acoust Soc Am. 2020;148: 1824. doi:http://dx.doi.org/10.1121/10.0002112
877. Monteiro de Oliveira M, Fuller TL, Brasil P, Gabaglia CR, Nielsen-Saines K. Controlling the COVID-19 pandemic in Brazil: a challenge of continental proportions. Nat Med. 2020;26: 1505–1506. doi:http://dx.doi.org/10.1038/s41591-020-1071-5
878. Monterrosa-Castro Á, Dávila-Ruiz R, Mejía-Mantilla A, Contreras-Saldarriaga J, Mercado-Lara M, Flores-Monterrosa C. Occupational Stress, Anxiety and Fear of COVID-19 in Colombian Physicians. MedUNAB. 2020;23: 214–232.
879. Monterrosa-Castro A, Redondo-Mendoza V, Mercado-Lara M. Psychosocial factors associated with symptoms of generalized anxiety disorder in general practitioners during the COVID-19 pandemic. J Investig Med. 2020;68: 1228–1234. doi:https://dx.doi.org/10.1136/jim-2020-001456
880. Monterrubio-Flores E, Ramírez-Villalobos M, Espinoza-Montero J, Hernandez B, Barquera S, Villalobos-Daniel V, et al. Impact of a Double Epidemic in Mexico: Non-Communicable Diseases Increase the Case Fatality Rate with Covid-19. ResearchSquare. 2020. doi:10.21203/rs.3.rs-80669/v1
881. Moquillaza-Alcántara V, Romero-Cerdán A, Munares-García O, Merellano-Navarro E. Variation in the epidemiological indicators of COVID-19 a part of the emergency policies adopted in south american countries. 2020.
882. Moraes RF de. Determinants of physical distancing during the covid-19 epidemic in Brazil: effects from mandatory rules, numbers of cases and duration of rules. Cien Saude Colet. 2020;25: 3393–3400. doi:https://dx.doi.org/10.1590/1413-81232020259.21892020
883. Mora-Magana I, Lee SA, Maldonado-Castellanos I, Jimenez-Gutierrez C, Mendez-Venegas J, Maya-Del-Moral A, et al. Coronaphobia among healthcare professionals in Mexico: A psychometric analysis. Death Stud. 2020; 1–10. doi:http://dx.doi.org/10.1080/07481187.2020.1808762
884. Moreau VH. Forecast predictions for the COVID-19 pandemic in Brazil by statistical modeling using the Weibull distribution for daily new cases and deaths. Braz J Microbiol. 2020. doi:10.1007/s42770-020-00331-z

Cesar VM, Amna T, Gabriela GS-C, Mary FR, Andree V, Leonardo R-M, et al. Early transmission dynamics and control of COVID-19 in a southern hemisphere setting: Lima-Peru, February 29th-March 30th, 2020. medRxiv. 2020. doi:10.1101/2020.04.30.20077594
893. Munguía-López A, Ponce-Ortega J. Fair allocation of potential COVID-19 vaccines using an optimization-based strategy. ResearchSquare. 2020. doi:10.21203/rs.3.rs-83772/v1
894. Muñoz L, Pífano M, Bolzán A, Varela T, Comes Y, Specogna M, et al. Surveillance and Seroprevalence: Evaluation of IgG antibodies for SARS-Cov2 by ELISA in the popular neighborhood Villa Azul, Quilmes, province of Buenos Aires, Argentina. 2020.
895. Murillo-Zamora E, Guzman-Esquivel J, Sanchez-Pina RA, Cedeno-Laurent G, Delgado-Enciso I, Mendoza-Cano O. Physical distancing reduced the incidence of influenza and supports a favorable impact on SARS-CoV-2 spread in Mexico. J Infect Dev Ctries. 2020;14: 953–956. doi:http://dx.doi.org/10.3855/JIDC.13250
896. Murillo-Zamora E, Hernandez-Suarez C. Performance of the case definition of suspected influenza before and during the COVID-19 pandemic. Rev Clin Esp. 2020. doi:http://dx.doi.org/10.1016/j.rce.2020.09.001

Efren M-Z, Carlos MH-S. Performance of suspected influenza case definitionbefore and during the COVID-19 pandemic. medRxiv. 2020. doi:10.1101/2020.06.01.20119446
897. Murillo-Zamora E, Trujillo X, Huerta M, Rios-Silva M, Mendoza-Cano O. Male gender and kidney illness are associated with an increased risk of severe laboratory-confirmed coronavirus disease. BMC Infect Dis. 2020;20: 674. doi:https://dx.doi.org/10.1186/s12879-020-05408-6

Efren M-Z, Xochitl T, Miguel H, Monica R-S, Oliver M-C. Male gender and kidney illness associated with an increased risk of severe laboratory-confirmed coronavirus disease. medRxiv. 2020. doi:10.1101/2020.06.29.20142562

898. Nabi KN. Forecasting COVID-19 pandemic: A data-driven analysis. Chaos Solitons Fractals. 2020;139: 110046. doi:10.1016/j.chaos.2020.110046

Khondoker Nazmoon N. FORECASTING COVID-19 PANDEMIC: A DATA-DRIVEN ANALYSIS. medRxiv. 2020. doi:10.1101/2020.05.12.20099192

899. Nadim SS, Chattopadhyay J. Occurrence of backward bifurcation and prediction of disease transmission with imperfect lockdown: A case study on COVID-19. Chaos Solitons Fractals. 2020;140: 110163. doi:10.1016/j.chaos.2020.110163

900. Nagata Cavalheiro M, Fonseca VR, Zeigelboim BS, Costa DF, de Lima L V, Bozzo MK, et al. Evaluating the Quality of Rigid Optic Videolaryngoscopy Image Taken Through Dental Protection Cap and Its Feasibility as Additional Barrier Method Against COVID-19. J Voice. 2020. doi:10.1016/j.jvoice.2020.10.014

901. Nakada LYK, Urban RC. COVID-19 pandemic: environmental and social factors influencing the spread of SARS-CoV-2 in Sao Paulo, Brazil. Environ Sci Pollut Res Int. 2020. doi:http://dx.doi.org/10.1007/s11356-020-10930-w

Nakada L, Urban R. COVID-19 pandemic: environmental and social factors influencing the spread of SARS-CoV-2 in the expanded metropolitan area of São Paulo, Brazil. ResearchSquare. 2020. doi:10.21203/RS.3.RS-34613/V1

909. Nascimento VF do, Hattori TY, Terças-Trette ACP. Difficulties and fears of nurses facing the COVID-19 pandemic in Brazil. Humanidad med. 2020;20.

910. Nascimento VF do, Hattori TY, Trettel ACPT. Personal needs of nurses during the COVID-19 pandemic in Mato Grosso. Enferm foco. 2020;11.

José MO-A. El nuevo coronavirus que llegó de Oriente: análisis de la epidemia inicial en México. Gac Med Mex. 2020;156. doi:10.24875/GMM.20000165

960. Paesano N, Santomil F, Tobia I. Impact of COVID-19 Pandemic on Ibero-American Urology Residents: Perspective of American Confederation of Urology (CAU). Int Braz J Urol. 2020;46. doi:https://dx.doi.org/10.1590/S1677-5538.IBJU.2020.S120
961. Pagotto VPF, Abbas L, Goldenberg DC, Lobato RC, do Nascimento BB, Monteiro GGR, et al. The impact of COVID-19 on the plastic surgery activity in a high-complexity university hospital in Brazil: the importance of reconstructive plastic surgery during the pandemic. Eur J Plast Surg. 2020. doi:http://dx.doi.org/10.1007/s00238-020-01729-6
962. Paixão B, Pedroso M, Baroni L, Salles R, Escobar L, Sousa C, et al. Estimation of COVID-19 under-reporting in the Brazilian States through SARI. ResearchSquare. 2020. doi:10.21203/rs.3.rs-50075/v1
963. Palacios R, Patino EG, de Oliveira Piorelli R, Conde MTRP, Batista AP, Zeng G, et al. Double-Blind, Randomized, Placebo-Controlled Phase III Clinical Trial to Evaluate the Efficacy and Safety of treating Healthcare Professionals with the Adsorbed COVID-19 (Inactivated) Vaccine Manufactured by Sinovac - PROFISCOV: A structured summary of a . Trials. 2020;21: 853. doi:http://dx.doi.org/10.1186/s13063-020-04775-4
964. Palamim CVC, Marson FAL. COVID-19 - The Availability of ICU Beds in Brazil during the Onset of Pandemic. Ann Glob Heal. 2020;86: 100. doi:https://dx.doi.org/10.5334/aogh.3025
965. Pan American Health O. Alcohol Use during the COVID-19 Pandemic in Latin America and the Caribbean, 8 September 2020. 2020.
966. Pan American Health O. Summary of the Status of National Immunization Programs during the COVID-19 Pandemic, July 2020. 2020.
967. Paola Cristina R, Edson D, Tiago G, Daiana M, Fernando CM, Luciana A, et al. Genomic surveillance of SARS-CoV-2 reveals community transmission of a major lineage during the early pandemic phase in Brazil. bioRxiv. 2020. doi:10.1101/2020.06.17.158006
968. Paroli M, Sirinian MI. Predicting SARS-CoV-2 infection trend using technical analysis indicators. Disaster Med Public Health Prep. 2020; 1–9. doi:10.1017/dmp.2020.254
969. Parra-Bracamonte GM, Lopez-Villalobos N, Parra-Bracamonte FE. Clinical characteristics and risk factors for mortality of patients with COVID-19 in a large dataset from Mexico. Ann Epidemiol. 2020. doi:10.1016/j.annepidem.2020.08.005
970. Parra-Saavedra M, Villa-Villa I, Pérez-Olivo J, Guzman-Polania L, Galvis-Centurion P, Cumplido-Romero Á, et al. Attitudes and collateral psychological effects of COVID-19 in pregnant women in Colombia. Int J Gynaecol Obstet. 2020. doi:10.1002/ijgo.13348
971. Parro V, Lafetá M, Ipolito F, Toporcov T. Predicting COVID-19 in very large countries: the case of Brazil. ResearchSquare. 2020. doi:10.21203/rs.3.rs-79708/v1
972. Passos L, Prazeres F, Teixeira A, Martins C. Impact on Mental Health Due to COVID-19 Pandemic: Cross-Sectional Study in Portugal and Brazil. Int J Environ Res Public Health. 2020;17. doi:https://dx.doi.org/10.3390/ijerph17186794
973. Patricia T, Fernanda MA-C, Renata K, Marina AMS, Matheus MT, Bianca QRCA, et al. Medical students perceptions and motivations in time of COVID-19 pandemic. medRxiv. 2020. doi:10.1101/2020.05.28.20115956
974. Patricio S, Hiram C. COVID-19 Fatality and Comorbidity Risk Factors among Confirmed Patients in Mexico. medRxiv. 2020. doi:10.1101/2020.04.21.20074591
975. Patricio V, Sebastian A, Eugenio EV, Sigismund K. Rapid characterization of the propagation of COVID-19 in different countries. medRxiv. 2020. doi:10.1101/2020.06.09.20126631
976. Patrick EB, Zoë RG, Luis Ernesto S, Gabriel Aguirre M, Leslie N, Peter SR, et al. Mortality from COVID in Colombia and Peru: Analyses of Mortality Data and Statistical Forecasts. medRxiv. 2020. doi:10.1101/2020.08.24.20181016
977. Patrick H. Estimating the Impact of COVID-19 on the Individual Lifespan:A Conceptual Detour and an Empirical Shortcut. medRxiv. 2020. doi:10.1101/2020.08.09.20171264

978. Paulino-Ramirez R, Baez AA, Vallejo Degaudenzi A, Tapia L. Seroprevalence of Specific Antibodies against SARS-CoV-2 from Hotspot Communities in the Dominican Republic. Am J Trop Med Hyg. 2020. doi:http://dx.doi.org/10.4269/ajtmh.20-0907
979. Paulo Cardoso Lins F, Jaciel Leandro de Melo F, Millena Mirella Silva de A, Thuanny Silva de M, Andressa Kelly Alves F, Maria Cecilia Freire de M, et al. Predicting social distancing index during COVID-19 outbreak through online search engines trends. medRxiv. 2020. doi:10.1101/2020.05.28.20115816
980. Paulo Cardoso Lins F, Thuanny Silva de M, Andressa Kelly Alves F, Maria Cecilia Freire de M, Millena Mirella Silva de A, Jaciel Leandro de Melo F, et al. Assessing the quality, readability and reliability of online information on COVID-19: aninfoveillance observational study. medRxiv. 2020. doi:10.1101/2020.05.30.20117614
981. Paulo JSS, Tiago P, Luis Gustavo N. Robot dance: a city-wise automatic control of Covid-19 mitigation levels. medRxiv. 2020. doi:10.1101/2020.05.11.20098541
982. Paz C, Mascialino G, Adana-Díaz L, Rodríguez-Lorenzana A, Simbaña-Rivera K, Gómez-Barreno L, et al. Anxiety and depression in patients with confirmed and suspected COVID-19 in Ecuador. Psychiatry Clin Neurosci. 2020. doi:10.1111/pcn.13106
983. Paz C, Mascialino G, Adana-Diaz L, Rodriguez-Lorenzana A, Simbana-Rivera K, Gomez-Barreno L, et al. Behavioral and sociodemographic predictors of anxiety and depression in patients under epidemiological surveillance for COVID-19 in Ecuador. PLoS One. 2020;15: e0240008. doi:https://dx.doi.org/10.1371/journal.pone.0240008
984. Pedro de Lemos M, David MG, Vitor EV. BRAZIL IS PROJECTED TO BE THE NEXT GLOBAL COVID-19 PANDEMIC EPICENTER. medRxiv. 2020. doi:10.1101/2020.04.28.20083675
985. Pedro Emanuel F, Jorge AP, Mario IS, Carlos V, Ruben OC, Alejandro JK, et al. Understanding the value of clinical symptoms of COVID-19. A logistic regression model. medRxiv. 2020. doi:10.1101/2020.10.07.20207019
986. Pedro Henrique dos Santos S, Sara Sabrina Vieira C, Lorena Sousa S, Franciele Basso Fernandes S. Déficit e ocupação de leitos de unidade de terapia intensiva adulto do Sistema Único de Saúde no estado do Piauí sob a ótica da COVID-19. 2020.
987. Pedro CH, Fernando PH, Bernardo Lessa H, Mariangela FS, Claudio JS, Luís Paulo V, et al. Remarkable Variability in SARS-CoV-2 Antibodies across Brazilian Regions: Report on Two Successive Nationwide Serological Household Surveys. SSRN. 2020. doi:10.2139/ssrn.3640428

Pedro H, Fernando H, Bernardo H, Gabriel DV, Mariangela S, Claudio S, et al. Remarkable variability in SARS-CoV-2 antibodies across Brazilian regions: nationwide serological household survey in 27 states. medRxiv. 2020. doi:10.1101/2020.05.30.20117531
988. Pedro MCLP, Marcelo AS, Pedro VS. COVID-19 DYNAMICS CONSIDERING THE INFLUENCE OF HOSPITAL INFRASTRUCTURE: AN INVESTIGATION OF BRAZILIAN SCENARIOS. medRxiv. 2020. doi:10.1101/2020.06.03.20121608
989. Pedro SP, Diego RM, Cláudia MP, Lucas Q, Rafael G, Afonso D, et al. Potential dissemination of epidemics based on Brazilian mobile geolocation data. Part I: Population dynamics and future spreading of infection in the states of Sao Paulo and Rio de Janeiro during the pandemic of COVID-19. medRxiv. 2020. doi:10.1101/2020.04.07.20056739
990. Pedrosa NL, Albuquerque NLS de. Spatial Analysis of COVID-19 cases and intensive care beds in the State of Ceara, Brazil. Cien Saude Colet. 2020;25: 2461–2468. doi:https://dx.doi.org/10.1590/1413-81232020256.1.10952020
991. Pedrozo-Pupo JC, Campo-Arias A. Depression, perceived stress related to COVID, post-traumatic stress, and insomnia among asthma and COPD patients during the COVID-19 pandemic. Chron Respir Dis. 2020;17. doi:http://dx.doi.org/10.1177/1479973120962800

992. Pedrozo-Pupo JC, Pedrozo-Cortes MJ, Campo-Arias A. Perceived stress associated with COVID-19 epidemic in Colombia: an online survey. Cad Saude Publica. 2020;36: e00090520. doi:https://dx.doi.org/10.1590/0102-311x00090520
993. Peixoto PS, Marcondes D, Peixoto C, Oliva SM. Modeling future spread of infections via mobile geolocation data and population dynamics. An application to COVID-19 in Brazil. PLoS One. 2020;15: e0235732. doi:10.1371/journal.pone.0235732
994. Pelayo-Nieto M, Linden-Castro E, Gomez-Alvarado MO, Bravo-Castro EI, Rodriguez-Covarrubias FT. Has the COVID-19 pandemic impacted the practice of urology in Mexico? Rev Mex Urol. 2020;80.
995. Peloso RM, Ferruzzi F, Mori AA, Camacho DP, Franzin LC da S, Margioto Teston AP, et al. Notes from the Field: Concerns of Health-Related Higher Education Students in Brazil Pertaining to Distance Learning During the Coronavirus Pandemic. Eval Health Prof. 2020;43: 201–203. doi:https://dx.doi.org/10.1177/0163278720939302
996. Peña E, Amin S, Bustos V, Muñoz S, Martinez E, Gomez J, et al. Experience of Clinical Screening for Elective Orthopedic Surgery in The Midst of The COVID-19 Pandemic: An Alternative Proposal. ResearchSquare. 2020. doi:10.21203/rs.3.rs-91961/v1
997. Pena VH, Espinosa A. Predictive modeling to estimate the demand for intensive care hospital beds nationwide in the context of the COVID-19 pandemic. Medwave. 2020;20: e8039. doi:http://dx.doi.org/10.5867/medwave.2020.09.8039
998. Pequeno P, Mendel B, Rosa C, Bosholn M, Souza JL, Baccaro F, et al. Air transportation, population density and temperature predict the spread of COVID-19 in Brazil. PeerJ. 2020;2020: e9322. doi:http://dx.doi.org/10.7717/peerj.9322
999. Peralta EA, Taveras M. Effectiveness of teleconsultation use in access to mental health services during the coronavirus disease 2019 pandemic in the Dominican Republic. Indian J Psychiatry. 2020;62: S492–S494. doi:http://dx.doi.org/10.4103/psychiatry.IndianJPsychiatry_1047_20
1000. Peralta O, Ortínez-Alvarez A, Torres-Jardón R, Suárez-Lastra M, Castro T, Ruíz-Suárez LG. Ozone over Mexico City during the COVID-19 pandemic. Sci Total Environ. 2020; 143183. doi:10.1016/j.scitotenv.2020.143183
1001. Percy Sr. S-B, Carlos C, Yamilee H-R, Roger VA-C. Real-World Effectiveness of hydroxychloroquine, azithromycin, and ivermectin among hospitalized COVID-19 patients: Results of a target trial emulation using observational data from a nationwide Healthcare System in Peru. medRxiv. 2020. doi:10.1101/2020.10.06.20208066
1002. Pereda R, González D, Rivero HB, Rivero JC, Pérez A, López LDR, et al. Therapeutic Effectiveness of Interferon-α2b Against COVID-19: The Cuban Experience. J Interferon Cytokine Res. 2020;40: 438–442. doi:10.1089/jir.2020.0124
1003. Pereira LA, Mansano Soares LC, Nascimento PA, Netto Cirillo LR, Sakuma HT, Da Veiga GL, et al. Retinal findings in hospitalised patients with severe COVID-19. Br J Ophthalmol. 2020; 317576. doi:http://dx.doi.org/10.1136/bjophthalmol-2020-317576
1004. Pereira-Ávila F, Botelho E, Góes F, Gir E, Sousa L, Caldeira N, et al. Public Utilization of Face Masks in Brazil During COVID-19 Pandemic Outbreak: Temporal Trend Analysis. ResearchSquare. 2020. doi:10.21203/rs.3.rs-48087/v1
1005. Pereira-Avila FMV, Lam SC, Goes FGB, Gir E, Pereira-Caldeira NMV, Teles SA, et al. Factors associated with the use and reuse of face masks among Brazilian individuals during the COVID-19 pandemic. Rev Lat Am Enfermagem. 2020;28: e3360. doi:https://dx.doi.org/10.1590/1518-8345.4604.3360
1006. Pérez Milian JM, Berrio Aguila JE, Ojeda YP, Rodríguez Quesada NY, Fernández Pérez SE, Aranguren Torriente D. Evaluación de la satisfacción de pacientes atendidos por la COVID-19 en el Hospital Universitario Clínico Quirúrgico Cmdte. Manuel Fajardo Rivero. Medicentro (Villa Clara). 2020;24: 630–641.

1007. Pérez Milian JM, Berrio Aguila JE, Ojeda YP, Rodríguez Quesada NY, Fernández Pérez SE, Aranguren Torriente D. Evaluation of satisfaction of patients seen at ‟ Cmdte. Manuel Fajardo Rivero" Clinico -Surgical University Hospital due to COVID-19. Medicentro (Villa Clara). 2020;24: 630–641.
1008. Perez-Cano HJ, Moreno-Murguia MB, Morales-Lopez O, Crow-Buchanan O, English JA, Lozano-Alcazar J, et al. Anxiety, depression, and stress in response to the coronavirus disease-19 pandemic. Cir y Cir (English Ed. 2020;88: 562–568. doi:http://dx.doi.org/10.24875/CIRU.20000561
1009. Pescarini J, Silveira I, Souza-Filho J, Aquino R, Barreto M, Aquino E. Covid-19 in Latin America countries: Course of the pandemic and the different responses towards control. ResearchSquare. 2020. doi:10.21203/rs.3.rs-56504/v1
1010. Picon R V, Carreno I, da Silva AA, Mossmann M, Laste G, Domingues GDC, et al. Coronavirus disease 2019 population-based prevalence, risk factors, hospitalization, and fatality rates in southern Brazil. Int J Infect Dis. 2020;100: 402–410. doi:http://dx.doi.org/10.1016/j.ijid.2020.09.028
1011. Pierrotti LC, Reusing Junior JO, Freire MP, Barros Machado DJ, Megale Moreira R, Ventura CG, et al. COVID-19 among kidney transplant recipients requiring hospitalization: preliminary data and outcomes from a single-center in Brazil. Transpl Int. 2020. doi:http://dx.doi.org/10.1111/tri.13745
1012. Pimentel FF, Borba JMC, Rodrigues TCGF, Peruchi KPI, Carrara HHA, de Andrade JM, et al. Chemotherapy prescriptions and emergency medical care utilization by patients in neo/adjuvant breast cancer treatment during COVID-19 pandemic: Patterns in a Brazilian University Hospital. Ann Oncol. 2020;31: S1020. doi:http://dx.doi.org/10.1016/j.annonc.2020.08.1811
1013. Pineda-Sic RA, Galarza-Delgado DA, Serna-Pena G, Castillo-Torres SA, Flores-Alvarado DE, Esquivel-Valerio JA, et al. Treatment adherence behaviours in rheumatic diseases during COVID-19 pandemic: a Latin American experience. Ann Rheum Dis. 2020. doi:https://dx.doi.org/10.1136/annrheumdis-2020-218198
1014. Pintado JF, Gibaja W, Vallejos RA, Rosas W, Guerra-Farfan E, Nunez JH. How COVID-19 has affected emergent visits to a Latin-American trauma department: Experience at a Peruvian national trauma referral center. Injury. 2020. doi:http://dx.doi.org/10.1016/j.injury.2020.11.005
1015. Pinto Neto O, Pinto I, Pinto O. Lightning during the COVID-19 pandemic in Brazil. J Atmos solar-terrestrial Phys. 2020;211: 105463. doi:10.1016/j.jastp.2020.105463
1016. Pinto A de S, Santos Junior EG Dos, Rodrigues CA, Nunes PCM, Cruz LA da, Costa MGR, et al. Covid-19 growth rate analysis: application of a low-complexity tool for understanding and comparing epidemic curves. Rev Soc Bras Med Trop. 2020;53: e20200331. doi:https://dx.doi.org/10.1590/0037-8682-0331-2020
1017. Pinzón M, Arango D, Betancur J, Holguín H, Arias C, Muñoz B, et al. Clinical Outcome of Patients with COVID-19 Pneumonia Treated with Corticosteroids and Colchicine in Colombia. ResearchSquare. 2020. doi:10.21203/rs.3.rs-94922/v1
1018. Pio-Abreu A, do Nascimento MM, Vieira MA, de Menezes Neves PDM, Lugon JR, Sesso R. High mortality of CKD patients on hemodialysis with Covid-19 in Brazil. J Nephrol. 2020. doi:10.1007/s40620-020-00823-z
1019. Pitts WJ, Inkpen CS. Assessing the effects of covid-19 in prisons in the northern triangle of central america. Vict Offender. 2020. doi:10.1080/15564886.2020.1828211
1020. Platt VB, Guedert JM, Coelho EBS. VIOLENCE AGAINST CHILDREN AND ADOLESCENTS: NOTIFICATION AND ALERT IN TIMES OF PANDEMIC. Rev Paul Pediatr. 2020;39: e2020267. doi:http://dx.doi.org/10.1590/1984-0462/2021/39/2020267
1021. Plotnikow GA, Matesa A, Nadur JM, Alonso M, Nunez II, Vergara G, et al. Characteristics and outcomes of patients infected with nCoV19 requiring invasive mechanical ventilation in Argentina. Rev Bras Ter intensiva. 2020;32: 348–353. doi:http://dx.doi.org/10.5935/0103-507X.20200062
1022. Plotnikow GA, Matesa A, Nadur JM, Alonso M, Nuñez I I, Vergara G, et al. Características y resultados de los pacientes infectados con nCoV19 con requerimiento de ventilación mecánica invasiva en la Argentina. Rev Bras Ter intensiva. 2020;32: 348–353. doi:10.5935/0103-507x.20200062

1023. Poblete Umanzor R, Saldías Peñafiel F, Sabatini Ugarte N, Vite Valverde A, Ceriani Bravo A, Schaffeld Pernas S, et al. Infección respiratoria aguda por coronavirus Sars-CoV-2 en personal de salud. Implementación de un programa de detección precoz y seguimiento de casos en un hospital universitario. Rev Med Chil. 2020;148: 724–733.
1024. Poterico JA, Mestanza O. Genetic variants and source of introduction of SARS-CoV-2 in South America. J Med Virol. 2020. doi:https://dx.doi.org/10.1002/jmv.26001
1025. Prado MFD, Antunes BBP, Bastos L, Peres IT, Silva A, Dantas LF, et al. Analysis of COVID-19 under-reporting in Brazil. Rev Bras Ter intensiva. 2020;32: 224–228. doi:10.5935/0103-507X.20200030
1026. Prado T, Fumian TM, Mannarino CF, Maranhão AG, Siqueira MM, Miagostovich MP. Preliminary results of SARS-CoV-2 detection in sewerage system in Niterói municipality, Rio de Janeiro, Brazil. Mem Inst Oswaldo Cruz. 2020;115. doi:10.1590/0074-02760200196
1027. Prado-Galbarro FJ, Sanchez-Piedra C, Gamino-Arroyo AE, Cruz-Cruz C. Determinants of survival after severe acute respiratory syndrome coronavirus 2 infection in Mexican outpatients and hospitalised patients. Public Health. 2020;189: 66–72. doi:http://dx.doi.org/10.1016/j.puhe.2020.09.014
1028. Prado-Gascó V, Gómez-Domínguez MT, Soto-Rubio A, Díaz-Rodríguez L, Navarro-Mateu D. Stay at Home and Teach: A Comparative Study of Psychosocial Risks Between Spain and Mexico During the Pandemic. Front Psychol. 2020;11: 566900. doi:10.3389/fpsyg.2020.566900
1029. Prata DN, Rodrigues W, Bermejo PH. Temperature significantly changes COVID-19 transmission in (sub)tropical cities of Brazil. Sci Total Environ. 2020;729: 138862. doi:https://dx.doi.org/10.1016/j.scitotenv.2020.138862
1030. Prata-Barbosa A, Lima-Setta F, Santos GRD, Lanziotti VS, de Castro RE V, de Souza DC, et al. Pediatric patients with COVID-19 admitted to intensive care units in Brazil: a prospective multicenter study. J Pediatr (Rio J). 2020. doi:10.1016/j.jped.2020.07.002
1031. Priolo Filho SR, Goldfarb D, Zibetti MR, Aznar-Blefari C. Brazilian Child Protection Professionals' Resilient Behavior during the COVID-19 Pandemic. Child Abus Negl. 2020; 104701. doi:http://dx.doi.org/10.1016/j.chiabu.2020.104701
1032. Pulido L, Solis-Aramayo MA, Ibarrola M, Heres M, Falco J, Tomaszuk G, et al. Initial experience in the attention of patients with COVID-19 in a private third-level hospital in Buenos Aires City. Medicina (B Aires). 2020;80: 433–438.
1033. Quevedo-Ramírez A, Al-Kassab-Córdova A, Mendez-Guerra C, Cornejo-Venegas G, Alva-Chavez KP. Altitude and excess mortality during COVID-19 pandemic in Peru. Respir Physiol Neurobiol. 2020;281: 103512. doi:10.1016/j.resp.2020.103512
1034. Quintana-Salcedo A, Vargas-Ortiz L, Severiche-Sierra C, Castro-Bocanegra V, Garcia-Moreno A, Vidal-Tovar C, et al. Knowledge and attitudes of health workers for the care of patients covid-19 in some cities of the Colombian Caribbean. Indian J Public Heal Res Dev. 2020;11: 219–229.
1035. Rafael Gonzalez G, Francisco M, Pablo SM, Miguel K. Is a COVID19 Quarantine Justified in Chile or USA Right Now? medRxiv. 2020. doi:10.1101/2020.03.23.20042002
1036. Rafael Marques Da S. Modeling the COVID-19 dissemination in the South Region of Brazil and testing gradual mitigation strategies. medRxiv. 2020. doi:10.1101/2020.07.02.20145136
1037. Rafael Marques Da S, Carlos Fabio de Oliveira M, Cesar M. Scrutinizing the heterogeneous spreading of COVID-19 outbreak in Brazilian territory. medRxiv. 2020. doi:10.1101/2020.06.05.20123604
1038. Rafael Santos Rodrigues V, Erisson Linhares de A, Helmar Abreu Rocha V, Nara Michelle de Araujo E, Marcelo O, Sergio Antonio Bastos S. Management and Clinical Characteristics in Children with SARS-CoV-2 Infection: Experience in a highly complex public hospital in the city of Sao Paulo. medRxiv. 2020. doi:10.1101/2020.06.22.20136994
1039. Rafael HMP, Carlos Kaue Vieira B, Luciana Mendes S, Bernardo S, Pedro A, Nelson G, et al. Geographic access to COVID-19 healthcare in Brazil using a balanced float catchment area approach. medRxiv. 2020. doi:10.1101/2020.07.17.20156505

1040. Rafael RM, Marcos BC, Ana BQ, Andrea D, Joao PL, Tatiana P-C, et al. COVID-19 challenges to dentistry in the new pandemic epicenter: Brazil. medRxiv. 2020. doi:10.1101/2020.06.11.20128744
1041. Rafael R de MR, Neto M, Depret DG, Gil AC, Fonseca MHS, Souza-Santos R. Effect of income on the cumulative incidence of COVID-19: an ecological study. Rev latinoam enferm. 2020;28: e3344–e3344.
1042. Rahul Kalippurayil M. Role of Weather Factors in COVID-19 Deaths in Tropical Climate: A Data-Driven Study Focused on Brazil. medRxiv. 2020. doi:10.1101/2020.09.13.20193532
1043. Raigam Jafet M-P, Alexadros S, Johnatan T-T, Charzakis C, Ameth H-V, Jose Rafael V-B, et al. Risk factors for mortality in pregnant women with SARS-CoV-2 infection. medRxiv. 2020. doi:10.1101/2020.05.31.20107276
1044. Rainisch G, Undurraga EA, Chowell G. A dynamic modeling tool for estimating healthcare demand from the COVID19 epidemic and evaluating population-wide interventions. Int J Infect Dis. 2020;96: 376–383. doi:https://dx.doi.org/10.1016/j.ijid.2020.05.043
1045. Rajneesh B. A Predictive Model for the Evolution of COVID-19. medRxiv. 2020. doi:10.1101/2020.04.13.20063271
1046. Ramirez-Sandoval JC, Gaytan-Arocha JE, Xolalpa-Chavez P, Mejia-Vilet JM, Arvizu-Hernandez M, Rivero-Sigarroa E, et al. Prolonged Intermittent Renal Replacement Therapy for Acute Kidney Injury in COVID-19 Patients with Acute Respiratory Distress Syndrome. Blood Purif. 2020. doi:http://dx.doi.org/10.1159/000510996
1047. Ramsingh RAE, Duval JL, Rahaman NC, Rampersad RD, Angelini GD, Teodori G. Adult cardiac surgery in Trinidad and Tobago during the COVID-19 pandemic: Lessons from a developing country. J Card Surg. 2020. doi:http://dx.doi.org/10.1111/jocs.14975
1048. Raphael B, Alexsandros C, Luis M. Filling the Void? Organized Crime and COVID-19 in Rio De Janeiro. SSRN. 2020. doi:10.2139/ssrn.3678840
1049. Raphael B, Sergio F, Michael F, Luis M. Racial Inequality in Health Care During a Pandemic. SSRN. 2020. doi:10.2139/ssrn.3691313
1050. Rasa Z, Renne V, Daniel B, Adrian B, Danilo M, Rafael Z, et al. Drastic Improvements in Air Quality in Ecuador during the COVID-19 Outbreak. Aerosol Air Qual Res. 2020;20. doi:10.4209/AAQR.2020.05.0254
1051. Ravelo-Hernández J, Cáceres-Bernaola U, Becerra-Nuñez C, Mendívil-Tuchia S, Vásquez-Rivas ME, Aguirre-Quispe W, et al. EARLY PREDICTORS OF HOSPITAL MORTALITY IN PATIENTS WITH COVID-19 PNEUMONIA AT A LEVEL III HOSPITAL. LIMA, PERU. 2020. doi:10.1590/SciELOPreprints.1314
1052. Reina Ortiz M, Grijalva MJ, Turell MJ, Waters WF, Montalvo AC, Mathias D, et al. Biosafety at Home: How to Translate Biomedical Laboratory Safety Precautions for Everyday Use in the Context of COVID-19. Am J Trop Med Hyg. 2020. doi:https://dx.doi.org/10.4269/ajtmh.20-0677
1053. Reis RF, de Melo Quintela B, de Oliveira Campos J, Gomes JM, Rocha BM, Lobosco M, et al. Characterization of the COVID-19 pandemic and the impact of uncertainties, mitigation strategies, and underreporting of cases in South Korea, Italy, and Brazil. Chaos Solitons Fractals. 2020; 109888. doi:https://dx.doi.org/10.1016/j.chaos.2020.109888
1054. Remes-Troche J, Coss-Adame E, Amieva-Balmori M, Velasco J, Gómez-Castaños P, Flores-Rendón R, et al. Incidence of “new-onset” constipation and associated factors during lockdown due to the coronavirus-19 pandemic. ResearchSquare. 2020. doi:10.21203/rs.3.rs-77783/v1
1055. Remes-Troche JM, Ramos-de-la-Medina A, Manriquez-Reyes M, Martinez-Perez-Maldonado L, Lara EL, Solis-Gonzalez MA. Initial Gastrointestinal Manifestations in Patients with SARS-CoV-2 in 112 patients from Veracruz (Southeastern Mexico). Gastroenterology. 2020. doi:https://dx.doi.org/10.1053/j.gastro.2020.05.055
1056. Renata Maurício S. Teaching and literacy practices in COVID-19 pandemic times. Res Soc Dev. 2020;9. doi:10.33448/RSD-V9I7.4430

1057. Renato Rodrigues S, Wisley Donizetti V, Wanderson da Silva M, Carlos Augusto Goncalves T. A Bayesian analysis of the total number of cases of the COVID 19 when only a few data is available. A case study in the state of Goias, Brazil. medRxiv. 2020. doi:10.1101/2020.04.19.20071852
1058. Renzo JCC-A, Jay SK. Impact of COVID-19 Lockdown Policy on Homicide, Suicide, and Motor Vehicle Deaths in Peru. medRxiv. 2020. doi:10.1101/2020.07.11.20150193
1059. Requena D, Medico A, Chacon RD, Ramirez M, Marin-Sanchez O. Identification of Novel Candidate Epitopes on SARS-CoV-2 Proteins for South America: A Review of HLA Frequencies by Country. Front Immunol. 2020;11: 2008. doi:http://dx.doi.org/10.3389/fimmu.2020.02008
1060. Requia WJ, Kondo EK, Adams MD, Gold DR, Struchiner CJ. Risk of the Brazilian health care system over 5572 municipalities to exceed health care capacity due to the 2019 novel coronavirus (COVID-19). Sci Total Environ. 2020;730: 139144. doi:https://dx.doi.org/10.1016/j.scitotenv.2020.139144
1061. Rex FE, Borges CA de S, Kafer PS. Spatial analysis of the COVID-19 distribution pattern in Sao Paulo State, Brazil. Cien Saude Colet. 2020;25: 3377–3384. doi:https://dx.doi.org/10.1590/1413-81232020259.17082020
1062. Reyes-Olavarría D, Latorre-Román PÁ, Guzmán-Guzmán IP, Jerez-Mayorga D, Caamaño-Navarrete F, Delgado-Floody P. Positive and Negative Changes in Food Habits, Physical Activity Patterns, and Weight Status during COVID-19 Confinement: Associated Factors in the Chilean Population. Int J Environ Res Public Health. 2020;17: 1–14. doi:10.3390/ijerph17155431
1063. Rezende LFM, Thome B, Schveitzer MC, Souza-Junior PRB de, Szwarcwald CL. Adults at high-risk of severe coronavirus disease-2019 (Covid-19) in Brazil. Rev Saude Publica. 2020;54: 50. doi:https://dx.doi.org/10.11606/s1518-8787.2020054002596
1064. Ribeiro H V, Sunahara AS, Sutton J, Perc M, Hanley QS. City size and the spreading of COVID-19 in Brazil. PLoS One. 2020;15: e0239699. doi:https://dx.doi.org/10.1371/journal.pone.0239699
1065. Ribeiro MHDM, da Silva RG, Mariani VC, Coelho LDS. Short-term forecasting COVID-19 cumulative confirmed cases: Perspectives for Brazil. Chaos Solitons Fractals. 2020;135: 109853. doi:https://dx.doi.org/10.1016/j.chaos.2020.109853
1066. Ribeiro SP, Castro e Silva A, Dattilo W, Reis AB, Goes-Neto A, Alcantara LC, et al. Severe airport sanitarian control could slow down the spreading of COVID-19 pandemics in Brazil. PeerJ. 2020;2020: e9446. doi:http://dx.doi.org/10.7717/peerj.9446
1067. Ricardo Vicente F, Marcos RM, Rogerio HT, Luiza MA, Michael Richard D, Eric MD. Applying a Prospective Space-Time Scan Statistic to Examine the Evolution of COVID-19 Clusters in the State of Sao Paulo, Brazil. medRxiv. 2020. doi:10.1101/2020.06.04.20122770
1068. Ricardo A, Edgar CC, Anabel SB, Adrian LP, Orlando G, Fernando H, et al. Decision-making on management of ms and nmosd patients during the COVID-19 pandemic: A latin american survey. Mult Scler Relat Disord. 2020;44: 102310. doi:https://dx.doi.org/10.1016/j.msard.2020.102310
1069. Ricardo P, Daniel G, Hubert Blas R, Juan Carlos R, albadio P, Lissette del Rosario L, et al. Therapeutic effectiveness of interferon alpha 2b treatment for COVID-19 patient recovery. medRxiv. 2020. doi:10.1101/2020.07.28.20157974
1070. Ricardo P, Daniel G, Hubert R, Juan R, Albadio P, Lizet del Rosario L, et al. Therapeutic effectiveness of interferon-alpha2b against COVID-19: the Cuban experience. medRxiv. 2020. doi:10.1101/2020.05.29.20109199
1071. Ricardo V, Juan D, Jose F, Jorge S, Fernando P, Dardo R, et al. Convalescent plasma as potential therapy for severe COVID-19 pneumonia. medRxiv. 2020. doi:10.1101/2020.09.01.20184390
1072. Richard Oliveira J, Claudia Rakel Pena P. PANDEMIA DO CORONAVÍRUS (COVID-19): análise da evolução de casos frente à disponibilidade de leitos no estado do Maranhão, Brasil, 2020. 2020;5.
1073. Ridwan K, Jeff C. Mayoral Party Identity and Social Distancing Behaviors in Brazil. SSRN. 2020. doi:10.2139/ssrn.3642710
1074. Rios AFM, Lira LSSP, Reis IM, Silva GA. Atenção Primária à Saúde frente à COVID-19: Relato de experiência de um Centro de Saúde. Enferm foco. 2020;11: 246–251.

1075. Rios AFM, Lira LSSP, Reis IM, Silva GA. Primary health care in front of COVID-19: experience report from a health center. Enferm foco. 2020;11: 246–251.
1076. Rios-González CM. Knowledge, attitudes and practices towards COVID-19 in Paraguayans during outbreaks: a quick online survey. 2020.
1077. Rios-González CM, Palacios JM. Symptoms of Anxiety and depression during the outbreak of COVID-19 in Paraguay. 2020.
1078. Rios-Silva M, Murillo-Zamora E, Mendoza-Cano O, Trujillo X, Huerta M. COVID-19 mortality among pregnant women in Mexico: A retrospective cohort study. J Glob Health. 2020;10: 20512. doi:http://dx.doi.org/10.7189/jogh.10.020512
1079. Risi LR, Oliveira AB de, Bernardes MMR, Ramos R de S, Moutinho ECV da S, Passos AC. Desenvolvimento de máscaras de tecido e celulose para resposta emergencial à pandemia provocada pelo SARS-CoV-2. Rev enferm UERJ. 2020;28: e51476–e51476.
1080. Rivera B, Leyva A, Portela MM, Moratorio G, Moreno P, Durán R, et al. Quantitative proteomic dataset from oro- and naso-pharyngeal swabs used for COVID-19 diagnosis: Detection of viral proteins and host’s biological processes altered by the infection. Data Br. 2020;32: 106121. doi:10.1016/j.dib.2020.106121
1081. Rivera VM, Gracia F, Diaz A. Therapeutic attitudes towards multiple sclerosis in Central America and the Caribbean facing the SARS-CoV-2 pandemia. Neurologia. 2020. doi:http://dx.doi.org/10.1016/j.nrl.2020.07.009
1082. Rivera-Franco MM, Saldana-Gonzalez ES, Leon-Rodriguez E. Changes within the care of hematopoietic cell transplantation patients after the shift of a Mexican institution to a COVID-19-only center. Hematol Transfus cell Ther. 2020;42: 313–315. doi:http://dx.doi.org/10.1016/j.htct.2020.08.001
1083. Rizo-Téllez SA, Méndez-García LA, Flores-Rebollo C, Alba-Flores F, Alcántara-Suárez R, Manjarrez-Reyna AN, et al. The Neutrophil-to-Monocyte Ratio and Lymphocyte-to-Neutrophil Ratio at Admission Predict In-Hospital Mortality in Mexican Patients with Severe SARS-CoV-2 Infection (Covid-19). Microorganisms. 2020;8. doi:10.3390/microorganisms8101560
1084. Roberto E, Rodrigo Q. Estimate of the actual number of COVID-19 cases from the analysis of deaths. medRxiv. 2020. doi:10.1101/2020.09.21.20198416
1085. Rocha HAL, Alcantara ACC, Netto FCB, Ibiapina FLP, Lopes LA, Rocha SGMO, et al. Dealing with the impact of the COVID-19 pandemic on a Rapid Response Team operation in Brazil: Quality in practice. Int J Qual Heal care J Int Soc Qual Heal Care. 2020. doi:http://dx.doi.org/10.1093/intqhc/mzaa114
1086. Rodrigues Aguilar AC, Frange C, Huebra L, Dias Gomes AC, Tufik S, Santos Coelho FM. The effects of the COVID-19 pandemic on patients with narcolepsy. J Clin Sleep Med. 2020. doi:http://dx.doi.org/10.5664/jcsm.8952
1087. Rodrigues NCP, Andrade MKN, Monteiro DLM, Lino VTS, Reis IDN, Frossard VC, et al. COVID-19 morbidity and mortality in 2020: the case of the city of Rio de Janeiro. J Bras Pneumol. 2020;46: e20200341. doi:10.36416/1806-3756/e20200341
1088. Rodrigues NH, Silva LGA da. Management of the coronavirus pandemic in a hospital: professional experience report. J nurs Heal. 2020;10: 20104004.
1089. Rodríguez Monteagudo MA, Avello Martínez R, Morejón Giraldoni A, Rodríguez Monteagudo P, Avello Rodríguez A. A weekly analysis of cases confirmed to COVID-19 in Cuba: first 70 days. Medisur. 2020;18: 485–491.
1090. Rodriguez J, Fletcher A, Heredia F, Fernandez R, Ramirez Salazar H, Sanabria D, et al. Alternative management for gynecological cancer care during the COVID-2019 pandemic: A Latin American survey. Int J Gynaecol Obstet. 2020. doi:https://dx.doi.org/10.1002/ijgo.13272
1091. Rodriguez M, Peters A, Perez I, Spencer M, Barbe M, Porte L, et al. Covid-19 in South America: Clinical and Epidemiological Characteristics Among 381 Patients During the Early Phase of the Pandemic in Santiago, Chile. ResearchSquare. 2020. doi:10.21203/rs.3.rs-64458/v1

1092. Rodriguez R, Mondeja B, Valdes O, Resik S, Vizcaino A, Acosta E, et al. SARS-CoV-2: Enhancement and Segmentation of High-ResolutionMicroscopy Images. Part I. ResearchSquare. 2020. doi:10.21203/rs.3.rs-65818/v1
1093. Rodriguez-Benito C V, Navarro G, Caballero I. Using Copernicus Sentinel-2 and Sentinel-3 data to monitor harmful algal blooms in Southern Chile during the COVID-19 lockdown. Mar Pollut Bull. 2020;161: 111722. doi:http://dx.doi.org/10.1016/j.marpolbul.2020.111722
1094. Rogrigues P, Reis EC, Bianchi L, Palma A. Factors associated with physical activity during the COVID-19 pandemic in Rio de Janeiro, Brazil. Rev bras ativ fís saúde. 2020;25: 1–9.
1095. Romero P, Sánchez-Yupari M, Montero S, Tsukayama P. Use of complete SARS-CoV-2 genomes to estimate the Rt in Peru during March - April 2020. 2020.
1096. Romero-Lopez JP, Carnalla-Cortes M, Pacheco-Olvera DL, Ocampo-Godinez JM, Oliva-Ramirez J, Moreno-Manjon J, et al. A bioinformatic prediction of antigen presentation from SARS-CoV-2 spike protein revealed a theoretical correlation of HLA-DRB1*01 with COVID-19 fatality in Mexican population: An ecological approach. J Med Virol. 2020. doi:http://dx.doi.org/10.1002/jmv.26561
1097. Rosa MFP, Silva WNT da, Carvalho WRG de, Oliveira SV de. Epidemiology of COVID-19 in Uberlândia (MG): preliminary analysis of the impact of commercial activities's openness. J Heal NPEPS. 2020;5.
1098. Rosalinda P, Carlos A, Jose N, Wilneg Carolina R, Nathalia P, Emily L, et al. PREDICTIVE VALUE OF SMELL AND TASTE TEST VS PCR-RT SARS-COV-2 AND RAPID DIAGNOSTIC TESTS IN THE DIAGNOSIS OF INFECTION BY COVID-19. A PROSPECTIVE MULTI-CENTRIC STUDY. medRxiv. 2020. doi:10.1101/2020.08.31.20185298
1099. Rosana Moraes P, Paula Fernanda Ribeiro L, Keulle Oliveira da S, Amanda Alves F, Claudio Alberto Gellis de Mattos D, Sylvia Maria T, et al. Covid-19 and the current scenario of the City of Castanhal – PA, Brazil. Res Soc Dev. 2020;9. doi:10.33448/RSD-V9I8.5717
1100. Rosario DKA, Mutz YS, Bernardes PC, Conte-Junior CA. Relationship between COVID-19 and weather: Case study in a tropical country. Int J Hyg Environ Health. 2020;229: 113587. doi:https://dx.doi.org/10.1016/j.ijheh.2020.113587
1101. Rosse VP, Pereira JN, Boari A, Costa G V, Ribeiro JPC, Vieira-Filho M. São Paulo's atmospheric pollution reduction and its social isolation effect, Brazil. Air Qual Atmos Health. 2020; 1–10. doi:10.1007/s11869-020-00959-8
1102. Roy KH, Johns D. Transcervical Fibroid Ablation (TFA) in an Ambulatory Surgical Center Setting: Utility during the COVID-19 Pandemic. J Minim Invasive Gynecol. 2020;27: S113. doi:http://dx.doi.org/10.1016/j.jmig.2020.08.178
1103. Rubens Lichtenthaler Filho V, Daniel Gomes Sr. L. A dynamic model for Covid-19 in Brazil. medRxiv. 2020. doi:10.1101/2020.05.10.20097550
1104. Ruiz Brunner M de LM, Escobar Zuluaga LJ, Cieri ME, Condinanzi AL, Cuestas E. [COVID-19 and the reality of people with disabilities in Argentina: possibilities of complying with the considerations of the World Health Organization]. COVID-19 y la Real las Pers con Discapac en Argentina posibilidades del cumplimiento las consideraciones planteadas por la Organ Mund la Salud. 2020;77: 168–175. doi:https://dx.doi.org/10.31053/1853.0605.v77.n3.28767
1105. Ruiz Mamani PG, Morales-Garcia WC, White M, Marquez-Ruiz MS. Properties of a scale of concern for COVID-19: Exploratory analysis in a Peruvian sample. Med Clin (Barc). 2020. doi:http://dx.doi.org/10.1016/j.medcli.2020.06.022
1106. Ruiz-Manriquez J, Leon-Lara X, Campos-Murguia A, Solis-Ortega AA, Perez-Gonzalez B, Uscanga LF, et al. Knowledge of Latin American gastroenterologists and endoscopists regarding SARS-CoV-2 infection. Rev Gastroenterol Mex. 2020. doi:https://dx.doi.org/10.1016/j.rgmx.2020.04.003
1107. Ruiz-Patino A, Arrieta O, Pino LE, Rolfo C, Ricaurte L, Recondo G, et al. Mortality and Advanced Support Requirement for Patients With Cancer With COVID-19: A Mathematical Dynamic Model for Latin America. JCO Glob Oncol. 2020;6: 752–760. doi:https://dx.doi.org/10.1200/GO.20.00156

1108. Ruíz-Roso MB, de Carvalho Padilha P, Matilla-Escalante DC, Brun P, Ulloa N, Acevedo-Correa D, et al. Changes of Physical Activity and Ultra-Processed Food Consumption in Adolescents from Different Countries during Covid-19 Pandemic: An Observational Study. Nutrients. 2020;12: 1–13. doi:10.3390/nu12082289
1109. Ruiz-Roso MB, de Carvalho Padilha P, Mantilla-Escalante DC, Ulloa N, Brun P, Acevedo-Correa D, et al. Covid-19 Confinement and Changes of Adolescent's Dietary Trends in Italy, Spain, Chile, Colombia and Brazil. Nutrients. 2020;12. doi:https://dx.doi.org/10.3390/nu12061807
1110. Saavedra D, Añé-Kourí A, Sánchez N, Filgueira L, Betancourt J, Herrera C, et al. An Anti-CD6 Monoclonal Antibody (Itolizumab) Reduces Circulating IL-6 in Severe Covid-19 Elderly Patients. ResearchSquare. 2020. doi:10.21203/rs.3.rs-32335/v1
1111. Sakellariou D, Malfitano APS, Rotarou ES. Disability inclusiveness of government responses to COVID-19 in South America: a framework analysis study. Int J Equity Health. 2020;19: 131. doi:10.1186/s12939-020-01244-x
1112. Salama M, Ataman-Millhouse L, Braham M, Berjeb K, Khrouf M, Rodrigues JK, et al. Installing oncofertility programs for common cancers in limited resource settings (Repro-Can-OPEN Study): An extrapolation during the global crisis of Coronavirus (COVID-19) pandemic. J Assist Reprod Genet. 2020. doi:https://dx.doi.org/10.1007/s10815-020-01821-7
1113. Saldana F, Flores-Arguedas H, Camacho-Gutierrez JA, Barradas I. Modeling the transmission dynamics and the impact of the control interventions for the COVID-19 epidemic outbreak. Math Biosci Eng. 2020;17: 4165–4183. doi:https://dx.doi.org/10.3934/mbe.2020231
1114. Salgotra R, Gandomi M, Gandomi AH. Evolutionary modelling of the COVID-19 pandemic in fifteen most affected countries. Chaos Solitons Fractals. 2020;140: 110118. doi:10.1016/j.chaos.2020.110118
1115. Salinas-Escudero G, Carrillo-Vega MF, Granados-Garcia V, Martinez-Valverde S, Toledano-Toledano F, Garduno-Espinosa J. A survival analysis of COVID-19 in the Mexican population. BMC Public Health. 2020;20: 1616. doi:http://dx.doi.org/10.1186/s12889-020-09721-2
1116. Samaniego A, Urzúa A, Buenahora M, Vera-Villarroel P. Sintomatología asociada a trastornos de salud mental en trabajadores sanitarios en paraguay: Efecto COVID-19. Rev Interam Psicol. 2020;54. doi:10.30849/ripijp.v54i1.1298
1117. Samara Avelino de Souza F, Angelo Cezar Pinho T. A COVID-19 e os desafios da urbanização e habitabilidade nas cidades amazônicas: estudo de caso em Belém do Pará. 2020;29. doi:10.18542/PAPERSNAEA.V29I1.8861
1118. Samy D, Alexandre BS, Bruno AF, Rodrigo NR, Leandro Lane da Costa V, Jose G-N. Brazilian Modeling of COVID-19 (BRAM-COD): a Bayesian Monte Carlo approach for COVID-19 spread in a limited data set context. medRxiv. 2020. doi:10.1101/2020.04.29.20081174
1119. Sánchez Barajas DA, Maita Blanco EC, Garrido Urdaneta F, García Rojas JM. Epidemiological analysis of COVID-19. Bolivarian Republic of Venezuela. Obs del Conoc Rev Espec en Gestión Soc del Conoc. 2020;5: 32–52.
1120. Sánchez Hernández E, Cardona Gordo O, Ferrer Castro JE, Pérez Fouces F, Despaigne Bicet A. Variability of the evolutionary PCR results in patients with COVID -19. Medisan. 2020;24: 565–577.
1121. Sandes S, Freitas A dos S. SIR model with exposure rate for the study of the projection of COVID-19 cases in Sergipe. 2020.
1122. Santamaría-Holek I, Castaño V. Possible fates of the spread of SARS-CoV-2 in the Mexican context. R Soc open Sci. 2020;7: 200886. doi:10.1098/rsos.200886

Ivan S-H, Victor C. Possible fates of the dispersion of SARS-COV-2 in the Mexican context. medRxiv. 2020. doi:10.1101/2020.07.15.20154526
1123. Santana F, Lopes J, Perez M, Campana G, Levi J, Lopes F, et al. Seroconversion for SARS-CoV-2 in rheumatic patients on synthetic and biologics Disease Modifying Anti-Rheumatic Drugs in São Paulo, Brazil. ResearchSquare. 2020. doi:10.21203/rs.3.rs-97191/v1

1124. Santana-Cibrian M, Acuna-Zegarra MA, Velasco-Hernandez JX. Lifting mobility restrictions and the effect of superspreading events on the short-term dynamics of COVID-19. Math Biosci Eng. 2020;17: 6240–6258. doi:http://dx.doi.org/10.3934/mbe.2020330

Mario S-C, Manuel AA-Z, Jorge XV-H. Lifting mobility restrictions and the induced short-term dynamics of COVID-19. medRxiv. 2020. doi:10.1101/2020.07.23.20161026

1125. Santos DS, Menezes MO, Andreucci CB, Nakamura-Pereira M, Knobel R, Katz L, et al. Disproportionate impact of COVID-19 among pregnant and postpartum Black Women in Brazil through structural racism lens. Clin Infect Dis. 2020. doi:10.1093/cid/ciaa1066

1126. Santos HLPCD, Maciel FBM, Santos KR, Conceicao CDVSD, Oliveira RS, Silva NRFD, et al. Necropolitics and the impact of COVID-19 on the Black community in Brazil: a literature review and a document analysis. Cien Saude Colet. 2020;25: 4211–4224. doi:http://dx.doi.org/10.1590/1413-812320202510.2.25482020

1127. Santos I, Almeida G, Moura F. Adaptive SIR model for propagation of SARS-CoV-2 in Brazil. ResearchSquare. 2020. doi:10.21203/rs.3.rs-40116/v1

1128. Santos JPC Dos, Siqueira ASP, Praca HLF, Albuquerque HG. Vulnerability to severe forms of COVID-19: an intra-municipal analysis in the city of Rio de Janeiro, Brazil. Cad Saude Publica. 2020;36: e00075720. doi:https://dx.doi.org/10.1590/0102-311x00075720

1129. Santos JF, Dahmer Santos D. Hierarchy of Flu-like symptoms related to COVID-19 according to sex and color or race in reports of patients with Severe Acute Respiratory Syndrome in Brazil. 2020. doi:10.1590/SciELOPreprints.1161

1130. Santos LSE, Nunes LMM, Rossi BA, Taets G. Impacts of the COVID-19 pandemic on violence against women: reflections from the theory of human motivation from Abraham Maslow. 2020.

1131. Santos MM, Lucena EES, Lima KC, Brito AAC, Bay MB, Bonfada D. Survival and predictors of deaths of patients hospitalised due to COVID-19 from a retrospective and multicentre cohort study in Brazil. Epidemiol Infect. 2020;148: e198. doi:https://dx.doi.org/10.1017/S0950268820002034

1132. Santos VS, Quintans-Júnior LJ, de Souza Barboza W, Antunes de Souza Araújo A, Ricardo Martins-Filho P. Clinical Characteristics and Outcomes in Patients With COVID-2019 and Leprosy. J Eur Acad Dermatol Venereol. 2020. doi:10.1111/jdv.16899

1133. Santos VS, Souza Araujo AA, de Oliveira JR, Quintans-Junior LJ, Martins-Filho PR. COVID-19 mortality among Indigenous people in Brazil: a nationwide register-based study. J Public Health (Oxf). 2020. doi:http://dx.doi.org/10.1093/pubmed/fdaa176

1134. São Paulo Secretaria da S. Serological survey for SARS-COV-2: prevalence of infection in students from public and private schools in the city of São Paulo: phases 1 to 3. 2020.

1135. Sarabia-Cobo C, Pérez V, De Lorena P, Hermosilla-Grijalbo C, Sáenz-Jalón M, Fernández-Rodríguez A, et al. Experiences of geriatric nurses in nursing home settings across four countries in the face of the COVID-19 pandemic. J Adv Nurs. 2020. doi:10.1111/jan.14626

1136. Saravia-Bartra MM, Cazorla-Saravia P, Cedillo-Ramirez L. Anxiety level of first-year medicine students at a private university in peru in times of covid-19. Rev la Fac Med Humana. 2020;20: 568–573. doi:http://dx.doi.org/10.25176/RFMH.V20I4.3198

1137. Saueressig MG, Hackmann CL, Silva CES da, Ferreira J. Estimation of patients hospitalized for COVID-19 in an intensive care unit at the peak of the pandemic in Porto Alegre: Study with epidemiological model SEIHDR. 2020.

1138. Saulo BB, Marcelo MM, Daniel OC anda JEN-R. The COVID-19 (SARS-CoV-2) Uncertainty Tripod in Brazil: Assessments on model-based predictions with large under-reporting. arXiv Popul Evol. 2020.

1139. Savi P V, Savi MA, Borges B. A Mathematical Description of the Dynamics of Coronavirus Disease 2019 (COVID-19): A Case Study of Brazil. Comput Math Methods Med. 2020;2020: 9017157. doi:http://dx.doi.org/10.1155/2020/9017157

Marcelo AS, Beatriz B, Pedro VS. A Mathematical Description of the Dynamics of Coronavirus Disease (COVID-19): A Case Study of Brazil. arXiv Popul Evol. 2020.
1140. Schinkothe T, Gabri MR, Mitterer M, Gouveia P, Heinemann V, Harbeck N, et al. A Web- and App-Based Connected Care Solution for COVID-19 In- and Outpatient Care: Qualitative Study and Application Development. JMIR public Heal Surveill. 2020;6: e19033. doi:https://dx.doi.org/10.2196/19033
1141. Schmulson M, Gudino-Zayas M, Hani A. The Impact of COVID-19 Pandemic on Neurogastroenterologists in Latin America: Results of an Online Survey. J Clin Gastroenterol. 2020. doi:http://dx.doi.org/10.1097/MCG.0000000000001413
1142. Schuch FB, Bulzing RA, Meyer J, Vancampfort D, Firth J, Stubbs B, et al. Associations of moderate to vigorous physical activity and sedentary behavior with depressive and anxiety symptoms in self-isolating people during the COVID-19 pandemic: A cross-sectional survey in Brazil. Psychiatry Res. 2020;292: 113339. doi:10.1016/j.psychres.2020.113339
1143. Sebastian Iglesias O, Johnny Leandro Saavedra C. Aplicaciones de búsqueda de Google para la comunicación de riesgos en el manejo de la COVID-19. 2020;24.
1144. Seclen SN, Nunez-Robles E, Yovera-Aldana M, Arias- Chumpitaz A. Incidence of COVID-19 infection and prevalence of diabetes, obesity and hypertension according to altitude in Peruvian population. Diabetes Res Clin Pract. 2020;169: 108463. doi:http://dx.doi.org/10.1016/j.diabres.2020.108463
1145. Segovia-Juarez J, Castagnetto JM, Gonzales GF. High altitude reduces infection rate of COVID-19 but not case-fatality rate. Respir Physiol Neurobiol. 2020;281: 103494. doi:https://dx.doi.org/10.1016/j.resp.2020.103494
1146. Seligmann H, Iggui S, Rachdi M, Vuillerme N, Demongeot J. Inverted Covariate Effects for First versus Mutated Second Wave Covid-19: High Temperature Spread Biased for Young. Biology (Basel). 2020;9. doi:10.3390/biology9080226
1147. Sergio C. Modeling COVID-19 as a National Dynamics with a SARS-CoV-2 Prevalent Variant: Brazil - A Study Case. medRxiv. 2020. doi:10.1101/2020.09.25.20201558
1148. Sergio C. Quantifying Effects, Forecasting Releases, and Herd Immunity of the Covid-19 Epidemic in S. Paulo, Brazil. medRxiv. 2020. doi:10.1101/2020.05.20.20107912
1149. Serpa Franco A, de Mendonça Henrique D, Ferreira de Almeida L, Moreira do Prado L, Schwenck Gomes RV, Lyra da Silva RC. Segurança na intubação de sequência rápida recomendada no COVID-19: Relato de experiência. Rev Cuid. 2020;11: 1–11. doi:10.15649/cuidarte.1260
1150. Servin-Caamano A, Reyes-Herrera D, Flores-Lopez A, Robiou-Vivero EJA, Martinez-Rivera F, Galindo-Hernandez V, et al. Impact of liver enzymes on SARSCoV-2 infection and on the severity of clinical disease. Ann Hepatol. 2020;19: 20. doi:http://dx.doi.org/10.1016/j.aohep.2020.08.044
1151. Shanbhag NM, Duncan A, Santos EGD, Yazigi H, Grant-Tate M, Girgis NFF, et al. Results of the Survey Conducted Among Caribbean Physicians on a Zoom Meeting Discussing the Article "A Practical Approach to the Management of Cancer Patients During the Novel Coronavirus Disease 2019 (COVID-19) Pandemic: An International Collaborative Gr. Oncologist. 2020. doi:http://dx.doi.org/10.1002/onco.13556
1152. Shen X, Cai C, Li H. Socioeconomic restrictions slowdown COVID-19 far more effectively than favorable weather-evidence from the satellite. Sci Total Environ. 2020;748: 141401. doi:10.1016/j.scitotenv.2020.141401
1153. Shima S, Soheil N, Jiang L, Ayesha K, Durgesh C, Ghassem F, et al. Risk of Cerebrovascular Events in Hospitalized Patients with SARS-CoV-2 Infection. SSRN. 2020. doi:10.2139/ssrn.3605289
1154. Shunxiang H, Lin W, Li X, Aihong Z, Li S, Feng L, et al. Transmission Dynamics of Coronavirus Disease 2019 (COVID-19) in the World: The Roles of Intervention and Seasonality. medRxiv. 2020. doi:10.1101/2020.07.17.20156430

1155. Siciliano B, Carvalho G, da Silva CM, Arbilla G. The Impact of COVID-19 Partial Lockdown on Primary Pollutant Concentrations in the Atmosphere of Rio de Janeiro and Sao Paulo Megacities (Brazil). Bull Environ Contam Toxicol. 2020. doi:https://dx.doi.org/10.1007/s00128-020-02907-9
1156. Siciliano B, Dantas G, da Silva CM, Arbilla G. Increased ozone levels during the COVID-19 lockdown: Analysis for the city of Rio de Janeiro, Brazil. Sci Total Environ. 2020;737: 139765. doi:https://dx.doi.org/10.1016/j.scitotenv.2020.139765
1157. Siewe Fodjo JN, Faria de Moura Villela E, Van Hees S, Tibério Dos Santos T, Vanholder P, Vanholder P, et al. Impact of the COVID-19 pandemic on the medical follow-up and psychosocial well-being of people living with HIV: A cross-sectional survey. J Acquir Immune Defic Syndr. 2020. doi:10.1097/QAI.0000000000002468
1158. Silberman P, López E, Medina A, Díaz Bazán JM, Gómez Marquisio MD, López GA. Tele-revistas en unidades de terapia intensiva: Coordinación asistencial y aprendizaje en el marco de la pandemia. Rev argent salud publica. 2020;12: 1–5.
1159. Silva APSC, Maia LTS, Souza W V. Severe Acute Respiratory Syndrome in Pernambuco: comparison of patterns before and during the COVID-19 pandemic. Cien Saude Colet. 2020;25: 4141–4150. doi:http://dx.doi.org/10.1590/1413-812320202510.2.29452020
1160. Silva ARA da, Leal IA, Gonçalves FM, Setti RF, Tavares MC, Souza CV, et al. INFLUENCE OF SOCIAL DISTANCING DUE TO COVID-19 PANDEMIC IN EMERGENCY ATTENDANCE AND HOSPITALIZATIONS IN PEDIATRICS. 2020. doi:10.1590/SciELOPreprints.364
1161. Silva CPG da, Silva VC da, Britto PF, Jesus D de, Nonato VWC, Vilella RR. Educational activities for proper use of personal protection equipment in a federal reference hospital. Enferm foco. 2020;11: 228–233.
1162. Silva DF da, Oliveira MLC de. Epidemiologia da COVID-19: comparação entre boletins epidemiológicos. Comun ciênc saúde. 2020;31.
1163. Silva GCDA, Abe DK, Pedrenho R, Vilares RN, Cordeiro MD, Coelho RF, et al. Evaluation of uro-oncological surgical treatment during the Sars-CoV-2 pandemic in a Brazilian tertiary oncology institution, the new world epicenter. Int Braz J Urol. 2020;47. doi:http://dx.doi.org/10.1590/S1677-5538.IBJU.2020.0479
1164. Silva GAE, Jardim BC, Santos CVB Dos. Excess mortality in Brazil in times of Covid-19. Excesso mortalidade no Bras em tempos COVID-19. 2020;25: 3345–3354. doi:https://dx.doi.org/10.1590/1413-81232020259.23642020
1165. Silva JHD, Oliveira EC, Hattori TY, Lemos ERS, Terças-Trettel ACP. Description of COVID-19 cluster: isolation and testing in asymptomatic individuals as strategies to prevent local dissemination in Mato Grosso state, Brazil, 2020. Epidemiol e Serv saude Rev do Sist Unico Saude do Bras. 2020;29: e2020264. doi:10.5123/s1679-49742020000400005
1166. Silva L, Figueiredo Filho D. Using Benford's law to assess the quality of COVID-19 register data in Brazil. J Public Health (Oxf). 2020. doi:http://dx.doi.org/10.1093/pubmed/fdaa193
1167. Silva L, Figueiredo Filho D, Fernandes A. The effect of lockdown on the COVID-19 epidemic in Brazil: evidence from an interrupted time series design. Cad Saude Publica. 2020;36: e00213920. doi:http://dx.doi.org/10.1590/0102-311X00213920
1168. Silva LLS da, Lima AFR, Polli DA, Razia PFS, Pavao LFA, Cavalcanti MAF de H, et al. Social distancing measures in the fight against COVID-19 in Brazil: description and epidemiological analysis by state. Medidas distanciamento Soc para o enfrentamento da COVID-19 no Bras Caracter e Anal Epidemiol por estado. 2020;36: e00185020. doi:https://dx.doi.org/10.1590/0102-311X00185020
1169. Silva MT, Quintanilha G, Giesel L, Soldati AB, Jabarra C, Pereira C de A, et al. The Impact of the COVID-19 pandemic on a stroke center in Latin America. Int J Stroke. 2020; 1747493020941637. doi:https://dx.doi.org/10.1177/1747493020941637
1170. Silva RM da. Using the SIRD model to characterize the COVID-19 spreading in the states of Paraná, Rio Grande do Sul, and Santa Catarina. 2020.

1171. Silva R da. Estimates of effective reproduction number of SARS-CoV-2 in Brazil using phylodynamic analysis. 2020.
1172. Silva RR da, Santos MB dos, Santos AD dos, Tavares D dos S, Santos PL dos. Coronavirus disease and basic sanitation: too early to be worried? Rev Soc Bras Med Trop. 2020;53. doi:10.1590/0037-8682-0345-2020
1173. Silva RJ, Silva K, Mattos J. Spatial analysis on the dispersion of covid-19 in the state of Bahia. 2020.
1174. Silva R, Pessanha C, Meireles I, Coura C, Silva C, Peregrino A. Budgetarry impact of the incorporation of claritromycin in the treatment of infection by new coronavirus. 2020. doi:10.1590/SciELOPreprints.1026
1175. Silva R, Silva C, Machado D, Peregrino A, Marta C, Pestana L, et al. Lost years of life adjusted for disability (DALY) among nursing professionals due to COVID-19 infection in Brazil. 2020.
1176. Silva-Júnior MJ da, Mendonça KS, Lima CA de, Pires PLS, Calegari T, Oliveira SV de. Analysis of the spatial-temporal dynamics of incidence, mortality and test rates (rapid and RT-PCR) of COVID-19 in the state of Minas Gerais, Brazil. 2020.
1177. Silva-Malta MCF, Rodrigues DDOW, Chaves DG, Magalhaes NNS, Ribeiro MA, Cioffi JGM, et al. Impact of COVID-19 in the attendance of blood donors and production on a Brazilian Blood Centres. Transfus Med. 2020. doi:http://dx.doi.org/10.1111/tme.12733
1178. Silvana F, Vanina P, Lorena L, Julieta S, Magdalena Wagner M, Alicia M, et al. Community-level SARS-CoV-2 Seroprevalence Survey in urban slum dwellers of Buenos Aires City, Argentina: a participatory research. medRxiv. 2020. doi:10.1101/2020.07.14.20153858
1179. Silvano B de O, Victor Bertollo Gomes P, Fabiana G, Fabio Macedo M, Maria A, Wanderson Kleber de O, et al. Monitoring social distancing and SARS-CoV-2 transmission in Brazil using cell phone mobility data. medRxiv. 2020. doi:10.1101/2020.04.30.20082172
1180. Silva-Rodriguez EA, Galvez N, Swan GJF, Cusack JJ, Moreira-Arce D. Urban wildlife in times of COVID-19: What can we infer from novel carnivore records in urban areas? Sci Total Environ. 2020; 142713. doi:http://dx.doi.org/10.1016/j.scitotenv.2020.142713
1181. Silva-Tinoco R, GonzÁLez-CantÚ A, De La Torre-SaldaÑA V, GuzmÁN-Olvera E, Cuatecontzi-Xochitiotzi T, Castillo-MartÍNez L, et al. Effect in self-care behavior and difficulties in coping with diabetes during the COVID-19 pandemic. Rev Mex Endocrinol Metab y Nutr. 2020;7: 1–7. doi:10.24875/RME.20000063
1182. Silveira MF, Barros AJD, Horta BL, Pellanda LC, Victora GD, Dellagostin OA, et al. Population-based surveys of antibodies against SARS-CoV-2 in Southern Brazil. Nat Med. 2020. doi:10.1038/s41591-020-0992-3
1183. Silvina R, Hector C, Javier P, Sandra Di L, Maria Jose C, Fernando Adrian T, et al. Children hospitalized for COVID-19 during first winter of the pandemic in Buenos Aires, Argentina. medRxiv. 2020. doi:10.1101/2020.11.05.20225300
1184. Simian M ME, Cifuentes D M. Epidemiological characterization of patients with Covid-19 in the Hospital Clínico Universidad de Chile. Rev Hosp Clin Univ Chile. 2020;31: 103–108.
1185. Singh RK, Rani M, Bhagavathula AS, Sah R, Rodriguez-Morales AJ, Kalita H, et al. Prediction of the COVID-19 Pandemic for the Top 15 Affected Countries: Advanced Autoregressive Integrated Moving Average (ARIMA) Model. JMIR public Heal Surveill. 2020;6: e19115. doi:https://dx.doi.org/10.2196/19115
1186. Sippy R, Prado EO, Pizarro Fajardo F, Hidalgo I, Aguilar G V, Bonville CA, et al. Medically Attended Outpatient Coronavirus Infections in Ecuadorean Children During the 20 Months Preceding Countrywide Lockdown Related to the SARS-CoV-2 Pandemic of 2020. Pediatr Infect Dis J. 2020. doi:10.1097/INF.0000000000002840
1187. Skums P, Kirpich A, Icer Baykal P, Zelikovsky A, Chowell G. Global transmission network of SARS-CoV-2: from outbreak to pandemic. medRxiv Prepr Serv Heal Sci. 2020. doi:https://dx.doi.org/10.1101/2020.03.22.20041145

1188. Soares RCM, Mattos LR, Raposo LM. Risk Factors for Hospitalization and Mortality due to COVID-19 in Espírito Santo State, Brazil. Am J Trop Med Hyg. 2020. doi:10.4269/ajtmh.20-0483
1189. Sola A, Rodríguez S, Cardetti M, Dávila C. [Perinatal COVID-19 in Latin America]. Rev Panam Salud Publica. 2020;44: e47. doi:10.26633/RPSP.2020.47
1190. Sorbara M, Graviotto HG, Lage-Ruiz GM, Turizo-Rodriguez CM, Sotelo-Lopez LA, Serra A, et al. COVID-19 and the forgotten pandemic: follow-up of neurocognitive disorders during lockdown in Argentina. Neurologia. 2020. doi:http://dx.doi.org/10.1016/j.nrl.2020.07.015
1191. Soriano Sanchez JA, Perilla Cepeda TA, Zenteno M, Campero A, Yampolsky C, Varela ML, et al. Early Report on the Impact of COVID-19 Outbreak in Neurosurgical Practice Among Members of the Latin American Federation of Neurosurgical Societies. World Neurosurg. 2020. doi:https://dx.doi.org/10.1016/j.wneu.2020.04.226
1192. Soto R N, Cortés M C, Muñoz P M, Herrera G ME, Postigo V J, Suquilanda J A, et al. Clinical and epidemiological characterization of patients with Covid-19 treated in the Emergency Service of the Hospital Clínico Universidad de Chile between March and May 2020. Rev Hosp Clin Univ Chile. 2020;31: 97–102.
1193. Sousa AR de, Carvalho ES de S, Santana T da S, Sousa AFL, Figueiredo TFG, Escobar OJV, et al. Men's feelings and emotions in the Covid-19 framing. Sentim e emocoes homens no enquadramento da doenca Covid-19. 2020;25: 3481–3491. doi:https://dx.doi.org/10.1590/1413-81232020259.18772020
1194. Sousa AR de, Santana TDS, Palma EMS, Sousa AF de, Moreira WC, Rezende MF, et al. SARS-CoV-2 in Brazil and the psychosocial repercussions on male health: a socio-historical study. 2020.
1195. Sousa GJB, Garces TS, Cestari VRF, Florencio RS, Moreira TMM, Pereira MLD. Mortality and survival of COVID-19. Epidemiol Infect. 2020;148: e123. doi:https://dx.doi.org/10.1017/S0950268820001405
1196. Sousa GJB, Garces TS, Cestari VRF, Moreira TMM, Florêncio RS, Pereira MLD. Estimation and prediction of COVID-19 cases in Brazilian metropolises. Rev latinoam enferm. 2020;28: e3345–e3345.
1197. Souza A, Mota C, Rosa A, Figueiredo C, Candeias A. An Environmental GIS-based Variable Analysis on SARS-CoV-2 in the City of Recife, Brazil. ResearchSquare. 2020. doi:10.21203/rs.3.rs-95748/v1
1198. Souza CT V, Santana CS, Ferreira P, Nunes JA, Teixeira MLB, Gouvêa M. Caring in the age of COVID-19: lessons from science and society. Cad Saude Publica. 2020;36: e00115020. doi:10.1590/0102-311X00115020
1199. Souza CDF de, Leal TC, Santos LG. Circulatory System Diseases in Patients with COVID-19: Description of Clinical and Epidemiological Profile of 197 Deaths. Doencas do Aparel Circ em Individuos com COVID-19 Descricao do Perf Clin e Epidemiol 197 Obitos. 2020;115: 281–283. doi:https://dx.doi.org/10.36660/abc.20200453
1200. Souza CDF de, Paiva JPS de, Leal TC, Silva LF da, Santos LG. Spatiotemporal evolution of case fatality rates of COVID-19 in Brazil, 2020. J Bras Pneumol. 2020;46: e20200208. doi:https://dx.doi.org/10.36416/1806-3756/e20200208
1201. Souza JB de, Conceição VM da, Araújo JS, Bitencourt JV de OV, Silva Filho CC da, Rossetto M. Cancer in time of COVID-19: repercussions in the lives of women undergoing oncological treatment. Rev enferm UERJ. 2020;28: e51821–e51821.
1202. Souza L, Lima F, Roca T, Santos A, Naveca F, Maia A, et al. SARS-COV-2 genomic surveillance in the Brazilian Western Amazon region: evolutionary history of epidemic dissemination and genetic signature in Rondônia, Brazil. ResearchSquare. 2020. doi:10.21203/rs.3.rs-72476/v1
1203. Souza LG, Randow R, Lima PC. Reflections about the COVID-19: differentials by sex and age. Comun ciênc saúde. 2020;31.
1204. Souza LF de. Informal app workers and the impact of disease by the new Coronavirus: a theoretical reflection. J nurs Heal. 2020;10: 20104011.
1205. Srijit B, Md Moinul I, Alokkumar DE. Search for trends of Covid-19 infection in India, China, Denmark, Brazil, France. Germany and the USA on the basis of power law scaling. medRxiv. 2020. doi:10.1101/2020.04.03.20052878

1206. Steele EM, Rauber F, Costa CDS, Leite MA, Gabe KT, Louzada ML da C, et al. Dietary changes in the NutriNet Brasil cohort during the covid-19 pandemic. Rev Saude Publica. 2020;54: 91. doi:https://dx.doi.org/10.11606/s1518-8787.2020054002950
1207. Stephen XZ, Jiyao C, Asghar Afshar J, Aldo A-R, Huiyang D, Jizhen L, et al. Succumbing to the COVID-19 Pandemic: Healthcare Workers not Satisfied and Intend to Leave Their Jobs. medRxiv. 2020. doi:10.1101/2020.05.22.20110809
1208. Stephen XZ, Yifei W, Asghar Afshar J, Jianfeng J, Valentina Gomes Haensel S. First study on mental distress in Brazil during the COVID-19 crisis. medRxiv. 2020. doi:10.1101/2020.04.18.20070896
1209. Studart-Neto A, Guedes BF, Tuma RLE, Camelo Filho AE, Kubota GT, Iepsen BD, et al. Neurological consultations and diagnoses in a large, dedicated COVID-19 university hospital. Arq Neuropsiquiatr. 2020. doi:10.1590/0004-282x20200089
1210. Suarez V, Suarez Quezada M, Oros Ruiz S, Ronquillo De Jesus E. Epidemiology of COVID-19 in Mexico: from the 27th of February to the 30th of April 2020. Rev Clin Esp. 2020. doi:https://dx.doi.org/10.1016/j.rce.2020.05.007
1211. Sun SY, Guazzelli CAF, de Morais LR, Dittmer FP, Augusto MN, Soares AC, et al. Effect of delayed obstetric labor care during the COVID-19 pandemic on perinatal outcomes. Int J Gynaecol Obstet. 2020;151: 287–289. doi:10.1002/ijgo.13357
1212. Szente Fonseca SN, de Queiroz Sousa A, Wolkoff AG, Moreira MS, Pinto BC, Valente Takeda CF, et al. Risk of hospitalization for Covid-19 outpatients treated with various drug regimens in Brazil: Comparative analysis. Travel Med Infect Dis. 2020;38: 101906. doi:http://dx.doi.org/10.1016/j.tmaid.2020.101906
1213. Szwarcwald CL, Souza PRB de, Malta DC, Barros MB de A, Magalhães M de AFM, Xavier DR, et al. Adherence to physical contact restriction measures and the spread of COVID-19 in Brazil. Epidemiol e Serviços Saúde. 2020;29.
1214. Taboada B, Vazquez-Perez JA, Muñoz Medina JE, Ramos Cervantes P, Escalera-Zamudio M, Boukadida C, et al. Genomic Analysis of Early SARS-CoV-2 Variants Introduced in Mexico. J Virol. 2020. doi:10.1128/JVI.01056-20

Blanca T, Joel AV-P, Jose Esteban M-M, Pilar R-C, Marina E-Z, Celia B, et al. Genomic analysis of early SARS-CoV-2 strains introduced in Mexico. bioRxiv. 2020. doi:10.1101/2020.05.27.120402
1215. Tagliazucchi E, Balenzuela P, Travizano M, Mindlin GB, Mininni PD. Lessons from being challenged by COVID-19. Chaos Solitons Fractals. 2020;137: 109923. doi:https://dx.doi.org/10.1016/j.chaos.2020.109923
1216. Takemoto MLS, Menezes MO, Andreucci CB, Knobel R, Sousa LAR, Katz L, et al. Maternal mortality and COVID-19. J Matern Fetal Neonatal Med. 2020; 1–7. doi:10.1080/14767058.2020.1786056
1217. Takemoto MLS, Menezes MO, Andreucci CB, Knobel R, Sousa LAR, Katz L, et al. Clinical characteristics and risk factors for mortality in obstetric patients with severe COVID-19 in Brazil: a surveillance database analysis. BJOG. 2020. doi:10.1111/1471-0528.16470
1218. Tang Y, Serdan TDA, Masi LN, Tang S, Gorjao R, Hirabara SM. Epidemiology of COVID-19 in Brazil: using a mathematical model to estimate the outbreak peak and temporal evolution. Emerg Microbes Infect. 2020; 1–11. doi:https://dx.doi.org/10.1080/22221751.2020.1785337
1219. Tarcisio MRF, Fabiana SG dos S, Victor BG, Thiago AHR, Julio HRC, Walter MR, et al. Expected impact of COVID-19 outbreak in a major metropolitan area in Brazil. medRxiv. 2020. doi:10.1101/2020.03.14.20035873
1220. Tatiane Cristina Moraes de S, Natalia de Paula M, Jose K, Isabel Seelaender CR, Marcela Z, Maria Amelia SMV, et al. Socioeconomic Vulnerabilities and the Intensity of RT-PCR SARS-CoV-2 Testing Efforts in the Public Health System in Sao Paulo State. medRxiv. 2020. doi:10.1101/2020.10.29.20221960
1221. Tavares DM dos S, Oliveira NGN, Marchiori GF, Guimarães MSF, Santana LPM. Elderly individuals living by themselves: knowledge and measures to prevent the novel coronavirus. Rev Lat Am Enfermagem. 2020;28. doi:10.1590/1518-8345.4675.3383

1222. Teich VD, Klajner S, Almeida FAS, Dantas ACB, Laselva CR, Torritesi MG, et al. Epidemiologic and clinical features of patients with COVID-19 in Brazil. Einstein (Sao Paulo). 2020;18: eAO6022. doi:10.31744/einstein_journal/2020ao6022
1223. Teixeira L, Freitas RL de, Abad A, Silva JA da, Antonelli-Ponti M, Bastos S, et al. Anxiety-related psychological impacts in the COVID-19 pandemic on cardiovascular diseases and diabetes. 2020. doi:10.1590/SciELOPreprints.1010
1224. Teixeira SC. Circumnavigating the challenges of COVID-19 for Indigenous people: perspectives for public health. Public Health. 2020;186: 127–128. doi:http://dx.doi.org/10.1016/j.puhe.2020.07.021
1225. Tenorio-Mucha J, Lazo-Porras M, Monroy-Hidalgo A, Malaga G, Cardenas MK. Prices of essential drugs for management and treatment of COVID-19 in public and private Peruvian pharmacies. Acta Medica Peru. 2020;37: 267–277. doi:http://dx.doi.org/10.35663/amp.2020.373.1560
1226. Teoh JY-C, Ong WLK, Gonzalez-Padilla D, Castellani D, Dubin JM, Esperto F, et al. A Global Survey on the Impact of COVID-19 on Urological Services. Eur Urol. 2020. doi:https://dx.doi.org/10.1016/j.eururo.2020.05.025
1227. Tercio D-C, Daniel Souza L, Antonio Jr. CM, Bruno MP, Marcelo Augusto Fontenelle Ribeiro J. The trauma and acute care surgeon in the covid-19 pandemic era/ O cirurgião de trauma e emergência na era da pandemia de covid-19. 2020;1.
1228. The Gibraltar C-RGHS, Daniel G. Oxygen and mortality in COVID-19 pneumonia: a comparative analysis of supplemental oxygen policies and health outcomes across 26 countries. medRxiv. 2020. doi:10.1101/2020.07.03.20145763
1229. Thiago Barbosa S, Thais de Rezende Bessa G, Larissa Gomes da S, Cintia Braga da S, Larissa Antônio da S, Amanda Felix A. The psychological health of Nursing professionals during direct coping with COVID-19 in Brazil. Res Soc Dev. 2020;9. doi:10.33448/RSD-V9I8.5913
1230. Thiago Christiano S, Leandro A, Liang Z. Quantitative Analysis of the Effectiveness of Public Health Measures on COVID-19 Transmission. medRxiv. 2020. doi:10.1101/2020.05.15.20102988
1231. Thiago Gomes H, Matias Nunes F, Carlos Henrique Ramires F, Mirna Stela L, Marilia Arndt M, Giovano Pereira B, et al. Insufficient social distancing may be related to a future COVID-19 outbreak in Ijui-Brazil: Predictions of further social interventions. medRxiv. 2020. doi:10.1101/2020.06.22.20132910
1232. Thiago Henrique Evangelista A, Tafarel Andrade de S, Samyla de Almeida S, Nayani Alves R, Stefan Vilges de O. Underreporting of death by COVID-19 in Brazil's second most populous state. medRxiv. 2020. doi:10.1101/2020.05.20.20108415
1233. Thomas AM, Henrique HH, Swapnil M, Charlie W, Ricardo PS, Axel G, et al. Report 21: Estimating COVID-19 cases and reproduction number in Brazil. medRxiv. 2020. doi:10.1101/2020.05.09.20096701
1234. Thomas P, Tara P, Djane Clarys B-S, Vanderson S, Wuelton Marcelo M, James W, et al. INFERRED RESOLUTION THROUGH HERD IMMMUNITY OF FIRST COVID-19 WAVE IN MANAUS, BRAZILIAN AMAZON. medRxiv. 2020. doi:10.1101/2020.09.25.20201939
1235. Thomaz Ugliara Barone M, Bega Harnik S, Vieira de Luca P, Letícia de Souza Lima B, José Pineda Wieselberg R, Ngongo B, et al. The Impact of COVID-19 on People with Diabetes in Brazil. Diabetes Res Clin Pract. 2020;166: 108304. doi:10.1016/j.diabres.2020.108304
1236. Thome B, Rezende LFM, Schveitzer MC, Monteiro CN, Goldbaum M. Differences in the prevalence of risk factors for severe COVID-19 across regions of Sao Paulo City. Rev Bras Epidemiol. 2020;23: e200087. doi:https://dx.doi.org/10.1590/1980-549720200087
1237. Timothy MT, Fresia C, Harold Andre G, Rómulo F-M, Francisco CV, Claudia M. Altitude as a protective factor from COVID-19. medRxiv. 2020. doi:10.1101/2020.08.03.20167262
1238. Tomas de-C-B. A modified SEIR Model with Confinement and Lockdown of COVID-19 for Costa Rica. medRxiv. 2020. doi:10.1101/2020.05.19.20106492
1239. Tomazini BM, Maia IS, Cavalcanti AB, Berwanger O, Rosa RG, Veiga VC, et al. Effect of Dexamethasone on Days Alive and Ventilator-Free in Patients with Moderate or Severe Acute Respiratory

Distress Syndrome and COVID-19: The CoDEX Randomized Clinical Trial. JAMA - J Am Med Assoc. 2020;324: 1307–1316. doi:http://dx.doi.org/10.1001/jama.2020.17021

Bruno Martins T, Israel SM, Flavia RB, Maria Vitoria AOS, Franca PB, Eduardo Leite VC, et al. COVID-19-associated ARDS treated with DEXamethasone (CoDEX): Study design and rationale for a randomized trial. medRxiv. 2020. doi:10.1101/2020.06.24.20139303

1240. Toro L, Parra A, Alvo M. [COVID-19 epidemic in chile: impact on emergency services care and specific pathologies]. Epidemia COVID-19 en Chile impacto en atenciones Serv Urgenc y Patol Especificas. 2020;148: 558–560. doi:https://dx.doi.org/10.4067/s0034-98872020000400558

1241. Torre D, Pibaque R, Veloz T, Beltrán P, Baldeón L. In-House Development, Standardization, and Validation of a RT-qPCR Assay for the Detection of SARS-CoV-2 Virus in Ecuador. ResearchSquare. 2020. doi:10.21203/rs.3.rs-91429/v1

1242. Torrealba-Rodriguez O, Conde-Gutiérrez RA, Hernández-Javier AL. Modeling and prediction of COVID-19 in Mexico applying mathematical and computational models. Chaos Solitons Fractals. 2020;138: 109946. doi:10.1016/j.chaos.2020.109946

1243. Torres JP, Izquierdo G, Acuna M, Pavez D, Reyes F, Fritis A, et al. Multisystem inflammatory syndrome in children (MIS-C): Report of the clinical and epidemiological characteristics of cases in Santiago de Chile during the SARS-CoV-2 pandemic. Int J Infect Dis. 2020;100: 75–81. doi:http://dx.doi.org/10.1016/j.ijid.2020.08.062

1244. Torres JP, Piñera C, De La Maza V, Lagomarcino AJ, Simian D, Torres B, et al. SARS-CoV-2 antibody prevalence in blood in a large school community subject to a Covid-19 outbreak: a cross-sectional study. Clin Infect Dis. 2020. doi:10.1093/cid/ciaa955

1245. Torres R, Soares W, Ohashi O, Pessin G. The quest for better machine learning models to forecast COVID-19-related infections: A case study in the state of Pará-Brazil. ResearchSquare. 2020. doi:10.21203/rs.3.rs-37547/v1

1246. Torres TS, Hoagland B, Bezerra DRB, Garner A, Jalil EM, Coelho LE, et al. Impact of COVID-19 Pandemic on Sexual Minority Populations in Brazil: An Analysis of Social/Racial Disparities in Maintaining Social Distancing and a Description of Sexual Behavior. AIDS Behav. 2020. doi:10.1007/s10461-020-02984-1

1247. Torres-Roman JS, Kobiak IC, Valcarcel B, Diaz-Velez C, La Vecchia C. The reproductive number R0 of COVID-19 in Peru: An opportunity for effective changes. Travel Med Infect Dis. 2020; 101689. doi:https://dx.doi.org/10.1016/j.tmaid.2020.101689

1248. Trudeau JM, Alicea-Planas J, Vasquez WF. The value of COVID-19 tests in Latin America. Econ Hum Biol. 2020;39: 100931. doi:http://dx.doi.org/10.1016/j.ehb.2020.100931

1249. Trujillo A, Salazar F, Torres R, Arias P, Sotomayor K. An integer programming model to assign patients based on mental health impact for tele-psychotherapy intervention during the Covid–19 emergency. ResearchSquare. 2020. doi:10.21203/rs.3.rs-53836/v1

1250. Turner S, Niño N. Qualitative Analysis of the Coordination of Major System Change Within the Colombian Health System in Response to COVID-19: Study Protocol. ResearchSquare. 2020. doi:10.21203/rs.3.rs-46942/v1

1251. Ugas-Charcape C, Ucar M, Almanza-Aranda J, Rizo-Patrón E, Rantes C, Caro-Domínguez P, et al. Pulmonary Imaging Findings in Coronavirus Disease 2019 (COVID-19): A Series of 140 Latin American Children. ResearchSquare. 2020. doi:10.21203/rs.3.rs-94820/v1

1252. Ugliara Barone MT, Harnik SB, Chaluppe M, Vieira de Luca P, Ngongo B, Pedrosa HC, et al. Decentralized COVID-19 measures in Brazil were ineffective to protect people with diabetes. Diabetes Metab Syndr Clin Res Rev. 2020;14: 1973–1978. doi:http://dx.doi.org/10.1016/j.dsx.2020.10.005

1253. Urban RC, Nakada LYK. COVID-19 pandemic: Solid waste and environmental impacts in Brazil. Sci Total Environ. 2021;755: 142471. doi:http://dx.doi.org/10.1016/j.scitotenv.2020.142471

1254. valderilio F azevedo, leiticia chaffin Barbosa P, Gabriel mondin N, faissal Nemer hajar, guilherme kyuzaemon Osako N, leonardo krieger rafael, et al. STRATEGY FOR THE CONTAINMENT, MITIGATION, AND SUPPRESSION OF THE COVID-19 PANDEMIC IN FRAGILIZED COMMUNITIES ON THE PERIPHERY OF A LARGE BRAZILIAN CITY. medRxiv. 2020. doi:10.1101/2020.09.28.20203174
1255. Valente TM, Ferreira LPS, Silva RAD, Leite JMRS, Tiraboschi FA, Barboza MCC. Brazil Covid-19: Change of hospitalizations and deaths due to burn injury? Burns. 2020. doi:http://dx.doi.org/10.1016/j.burns.2020.10.009
1256. Valenti VE, Menezes P de L, Abreu ACG de, Vieira GNA, Garner DM. Social distancing measures could have reduced estimated deaths related to COVID-19 in Brazil. J Hum Growth Dev. 2020;30: 164–169.
1257. Valentina V, Milva G, Juan C, Ezequiel J. Incubation period and serial interval of Covid-19 in a chain of infections in Bahia Blanca (Argentina). medRxiv. 2020. doi:10.1101/2020.06.18.20134825
1258. Valenzuela-Rodriguez G, Zambrano LI, Munoz-Lara F, Pecho-Silva S, Arteaga-Livias K, Rodriguez-Morales AJ. Intranational differences in the case fatality rates for COVID-19 among Peruvian physicians. Int J Infect Dis. 2020;101: 226–227. doi:http://dx.doi.org/10.1016/j.ijid.2020.09.018
1259. Valeria Oliveira S, Elaine Lopes de O, Marcia Jorge C, Rosemeire Y, Cintia Mayumi A, Giselle Ibette L-L, et al. PREVALENCE OF ANTIBODIES AGAINST SARS-CoV-2 IN PROFESSIONALS OF A PUBLIC HEALTH LABORATORY AT SAO PAULO, SP, BRAZIL. medRxiv. 2020. doi:10.1101/2020.10.19.20213421
1260. Valerio M, Miguel Q. An estimate of the COVID-19 infection fatality rate in Brazil based on a seroprevalence survey. medRxiv. 2020. doi:10.1101/2020.08.18.20177626
1261. Valle JAM. Predicting the number of total COVID-19 cases and deaths in Brazil by the Gompertz model. Nonlinear Dyn. 2020; 1–7. doi:10.1007/s11071-020-06056-w
1262. Vancini RL, Camargo-Neto L, de Lira CAB, Andrade MS, Viana RB, Nikolaidis PT, et al. Physical Activity and Sociodemographic Profile of Brazilian People during COVID-19 Outbreak: An Online and Cross-Sectional Survey. Int J Environ Res Public Health. 2020;17. doi:http://dx.doi.org/10.3390/ijerph17217964
1263. Vandormael A, Adam M, Greuel M, Barnighausen T. A short, animated video to improve good COVID-19 hygiene practices: a structured summary of a study protocol for a randomized controlled trial. Trials. 2020;21: 469. doi:https://dx.doi.org/10.1186/s13063-020-04449-1
1264. Varotsos CA, Krapivin VF. A new model for the spread of COVID-19 and the improvement of safety. Saf Sci. 2020; 104962. doi:10.1016/j.ssci.2020.104962
1265. Vasconcelos GL, Brum AA, Duarte-Filho GC, Ospina R, Almeida FAG, Macêdo AMS. OVERVIEW OF THE COVID-19 IN NORTHEAST BRAZIL: ANALYSES AND FORECASTS VIA GROWTH MODELS. 2020.
1266. Vasconcelos GL, Duarte-Filho GC, Brum AA, Ospina R, Almeida FAG, Macêdo AMS. Analysis of Covid-19 epidemic curves via generalized growth models: Case study for the cities of Recife and Teresina. 2020.
1267. Vasquez L, Sampor C, Villanueva G, Maradiegue E, Garcia-Lombardi M, Gomez-Garcia W, et al. Early impact of the COVID-19 pandemic on paediatric cancer care in Latin America. Lancet Oncol. 2020;21: 753–755. doi:https://dx.doi.org/10.1016/S1470-2045(20)30280-1
1268. Vasquez-Apestegui V, Parras-Garrido E, Tapia V, Paz-Aparicio VM, Rojas JP, Sánchez-Ccoyllo OR, et al. Association Between Air Pollution in Lima and the High Incidence of COVID-19: Findings from a Post Hoc Analysis. Res Sq. 2020. doi:10.21203/rs.3.rs-39404/v1
1269. Vasquez-Hoyos P, Diaz-Rubio F, Monteverde-Fernandez N, Jaramillo-Bustamante JC, Carvajal C, Serra A, et al. Reduced PICU respiratory admissions during COVID-19. Arch Dis Child. 2020; archdischild-2020. doi:http://dx.doi.org/10.1136/archdischild-2020-320469

1270. Vaz SN, Santana DSD, Netto EM, Pedroso C, Wang WK, Santos FDA, et al. Saliva is a reliable, non-invasive specimen for SARS-CoV-2 detection. Brazilian J Infect Dis. 2020;24: 422–427. doi:http://dx.doi.org/10.1016/j.bjid.2020.08.001
1271. Vecchio G, Zapico V, Catanzariti A, Bisso IC, Heras ML. Adverse effects of lopinavir/ritonavir in critically ill patients with COVID-19. Med. 2020;80: 439–441.
1272. Veiga E Silva L, de Andrade Abi Harb MDP, Teixeira Barbosa Dos Santos AM, de Mattos Teixeira CA, Macedo Gomes VH, Silva Cardoso EH, et al. COVID-19 Mortality Underreporting in Brazil: Analysis of Data From Government Internet Portals. J Med Internet Res. 2020;22: e21413. doi:https://dx.doi.org/10.2196/21413
1273. Veiga ESL, de Andrade Abi Harb MDP, Teixeira Barbosa Dos Santos AM, de Mattos Teixeira CA, Macedo Gomes VH, Silva Cardoso EH, et al. An analysis of COVID-19 mortality underreporting based on data available from official Brazilian government internet portals. J Med Internet Res. 2020;22: e21413. doi:10.2196/21413
1274. Vejar-Aguirre T, Jáuregui-Ulloa E, Gallo-Sánchez K, Mejía-García JA, Zavala-Hernández R, Arredondo A. [Evidences and trends for decision making about measures of containment and mitigation of Covid-19 in Jalisco, Mexico]. Salud Publica Mex. 2020. doi:10.21149/11724
1275. Vera-Hernandez CJ, Mendias-Alarcon A, Castillo-Olea C. 81 Implementing a Telehealth System in Baja California, Mexico to Address COVID-19 Pandemic. Ann Emerg Med. 2020;76: S32. doi:http://dx.doi.org/10.1016/j.annemergmed.2020.09.091
1276. Vera-Ponce VJ, Torres-Malca JR, Tello-Quispe EK, Orihuela-Manrique EJ, De La Cruz-Vargas JA. Validation of scale of changes in lifestyles during the quarantine period in a population of university students from lima, peru. Rev la Fac Med Humana. 2020;20: 614–623. doi:http://dx.doi.org/10.25176/RFMH.V20I4.3193
1277. Vicente CR, Sant'Ana MCS, Reis AP, Costa CF, Cerutti Junior C, Lanes CM, et al. Increasing Demand for Chest CT Due to COVID-19 in Brazil. Rev Soc Bras Med Trop. 2020;53: e20200608. doi:http://dx.doi.org/10.1590/0037-8682-0608-2020
1278. Victor D, Abayuba P, Enrique S, Alvaro N, Natalia P, Alejandro C, et al. Impact of COVID-19 Outbreak on Quality of Care for STEMI in Uruguay: A Whole Country Population Retrospective Study. SSRN. 2020. doi:10.2139/ssrn.3631297
1279. Victor JS, Diana F-L, Brian RZ, Eduardo R. Environmental and climatic impact on the infection and mortality of SARS-CoV-2 in Peru. medRxiv. 2020. doi:10.1101/2020.09.16.20196170
1280. Víctor SP. Preparados para un virus nocturno: coronavirus, decisiones de política pública y vulneración de derechos en cinco municipios de Sonora, México. Región y Soc. 2020;32. doi:10.22198/RYS2020/32/1324
1281. Victoria Izabela C, Carlos M. Percepción de riesgo y consumo mediático durante el inicio de la pandemia de COVID-19 en México. 2020.
1282. Victoria E, Gordon WH, Batsirai M, Sabine S, Paul Z, Christian M, et al. SARS-CoV-2 genomic characterization and clinical manifestation of the COVID-19 outbreak in Uruguay. medRxiv. 2020. doi:10.1101/2020.10.08.20208546
1283. Vieira MA da CES, Vieira CP de B, Borba A de S, Melo MC de C, Oliveira M de S, Melo RM, et al. Sequential serological surveys in the early stages of the coronavirus disease epidemic: limitations and perspectives. Rev Soc Bras Med Trop. 2020;53: e20200351. doi:https://dx.doi.org/10.1590/0037-8682-0351-2020
1284. Vieira-Meyer APGF, Coutinho MB, Guerra HP, Saintrain M V, Candeiro GTM. Brazilian primary and secondary public oral health attention: are dentists ready to face COVID-19 Pandemic? Disaster Med Public Health Prep. 2020; 1–22. doi:http://dx.doi.org/10.1017/dmp.2020.342
1285. Vilar-Compte M, Perez V, Teruel G, Alonso A, Perez-Escamilla R. Costing of actions to safeguard vulnerable Mexican households with young children from the consequences of COVID-19 social distancing measures. Int J Equity Health. 2020;19: 70. doi:https://dx.doi.org/10.1186/s12939-020-01187-3

1286. Villarreyes E, Luna A, Soriano A. Estimating the number of deaths due to COVID-19 in Lima and Peru during March and April 2020 using ARIMA time series and modeling. ResearchSquare. 2020. doi:10.21203/rs.3.rs-62317/v1
1287. Villela D. How limitations in data of health surveillance impact decision making in the COVID-19 epidemic. 2020.
1288. Villela E, Lopez R, Sato A, Oliveira F, Waldman E, Bergh R, et al. COVID-19 outbreak in Brazil: Adherence to national preventive measures and impact on people's lives. ResearchSquare. 2020. doi:10.21203/rs.3.rs-40186/v1
1289. Vinícius Carvalho J, Marcos Silveira B. Análise sistêmica do município de São Paulo e suas implicações para o avanço dos casos de Covid-19. 2020;34. doi:10.1590/S0103-4014.2020.3499.010
1290. Vinueza-Veloz AF, Aldaz-Pachacama NR, Mera-Segovia CM, Pino-Vaca DP, Tapia-Veloz EC, Vinueza-Veloz MF. Burnout syndrome among Ecuadorian medical doctors and nurses during COVID-19 pandemic. 2020.
1291. Viviana Guzzo L, Maria Sanali Moura P, Giordana Zeferino M, Marcelo Antônio Cartaxo Queiroga L, Ricardo Alves da C, Gláucia Maria Moraes de O. Brazilian Cardiology Interventional Registry during COVID-19 pandemic (RBCI-COVID19). 2020;28. doi:10.31160/JOTCI202028A202010
1292. Viviane Alves Dos Santos B, Roseane Christhina da Nova S-S, Maria Da Penha De Lima C, Emerson Araújo Do B, Maria Edna Silva De A. Representations and social anchorages of the new coronavirus and the COVID-19 treatment by Brazilians/ Representações e ancoragens sociais do novo coronavírus e do tratamento da COVID-19 por brasileiros. Estud Psicol. 2020.
1293. Vojvodic Hernández I, Ortega Checa D, Pinares Carrillo D, Alemán López J, Aburto Loroña V. Operaciones abdominopélvicas de emergencia en pacientes con COVID-19 en el Hospital Edgardo Rebagliati Martins - EsSalud, Lima, Perú. Rev colomb cir. 2020;35: 414–421.
1294. von Humboldt S, Mendoza-Ruvalcaba NM, Arias-Merino ED, Costa A, Cabras E, Low G, et al. Smart technology and the meaning in life of older adults during the Covid-19 public health emergency period: a cross-cultural qualitative study. Int Rev Psychiatry. 2020. doi:http://dx.doi.org/10.1080/09540261.2020.1810643
1295. Wachholz PA, Moreira VG, Oliveira D, Watanabe HAW, Villas Boas PJF. OCORRÊNCIA DE INFECÇÃO E MORTALIDADE POR COVID-19 EM RESIDENCIAIS PARA IDOSOS NO BRASIL. 2020.
1296. Wainer P, Saavedra F, Tagliapietra V, Abeledo D, Migliore D, Lapadula P, et al. [COVID-19 experience in a private institution in Buenos Aires during the first month of the pandemic: 26 cases]. Medicina (B Aires). 2020;80: 193–196.
1297. Waldecy Sr. R, David Nadler Sr. P, Wainesten Sr. C. REGIONAL DETERMINANTS OF THE EXPANSION OF COVID-19 IN BRAZIL. medRxiv. 2020. doi:10.1101/2020.04.13.20063925
1298. Wang P, Zheng X, Ai G, Liu D, Zhu B. Time series prediction for the epidemic trends of COVID-19 using the improved LSTM deep learning method: Case studies in Russia, Peru and Iran. Chaos Solitons Fractals. 2020;140: 110214. doi:10.1016/j.chaos.2020.110214
1299. Wang P, Zheng X, Li J, Zhu B. Prediction of epidemic trends in COVID-19 with logistic model and machine learning technics. Chaos Solitons Fractals. 2020;139: 110058. doi:10.1016/j.chaos.2020.110058
1300. Wang Y, Xu C, Yao S, Zhao Y. Forecasting the epidemiological trends of COVID-19 prevalence and mortality using the advanced alpha-Sutte Indicator. Epidemiol Infect. 2020;148: e236. doi:http://dx.doi.org/10.1017/S095026882000237X
1301. Wassenaar TM, Buzard GS, Newman DJ. BCG vaccination early in life does not improve COVID-19 outcome of elderly populations, based on nationally reported data. Lett Appl Microbiol. 2020. doi:10.1111/lam.13365
1302. Wendel S, Kutner JM, Machado R, Fontao-Wendel R, Bub C, Fachini R, et al. Screening for SARS-CoV-2 antibodies in convalescent plasma in Brazil: Preliminary lessons from a voluntary convalescent donor program. Transfusion. 2020. doi:http://dx.doi.org/10.1111/trf.16065

1303. Werberich GM, Marchiori E, Barreto MM, Rodrigues RS. Computed tomography findings in a Brazilian cohort of 48 patients with pneumonia due to coronavirus disease. Rev Soc Bras Med Trop. 2020;53. doi:10.1590/0037-8682-0405-2020
1304. Werneck AO, Silva DRD, Malta DC, Souza-Junior PRB, Azevedo LO, Barros MBA, et al. Lifestyle behaviors changes during the COVID-19 pandemic quarantine among 6,881 Brazilian adults with depression and 35,143 without depression. Cien Saude Colet. 2020;25: 4151–4156. doi:http://dx.doi.org/10.1590/1413-812320202510.2.27862020
1305. Werneck AO, Silva DR, Malta DC, Gomes CS, Souza-Junior PRB, Azevedo LO, et al. Associations of sedentary behaviors and incidence of unhealthy diet during the COVID-19 quarantine in Brazil. Public Health Nutr. 2020; 1–12. doi:http://dx.doi.org/10.1017/S1368980020004188
1306. Werneck AO, Silva DR, Malta DC, Lima MG, Souza-Junior PRB, Azevedo LO, et al. The mediation role of sleep quality in the association between the incidence of unhealthy movement behaviors during the COVID-19 quarantine and mental health. Sleep Med. 2020;76: 10–15. doi:http://dx.doi.org/10.1016/j.sleep.2020.09.021
1307. Werneck AO, Silva DR, Malta DC, Souza-Junior PRB, Azevedo LO, Barros MBA, et al. Changes in the clustering of unhealthy movement behaviors during the COVID-19 quarantine and the association with mental health indicators among Brazilian adults. Transl Behav Med. 2020. doi:http://dx.doi.org/10.1093/tbm/ibaa095
1308. Werneck AO, Silva DR, Malta DC, Souza-Júnior PRB, Azevedo LO, Barros MBA, et al. Physical inactivity and elevated TV-viewing reported changes during the COVID-19 pandemic are associated with mental health: A survey with 43,995 Brazilian adults. J Psychosom Res. 2021;140: 110292.
1309. Wesley Sr. D, Alcides Castro e S, Roger Sr. G, Ian M-F, Servio Pontes R. COVID-19 most vulnerable Mexican cities lack the public health infrastructure to face the pandemic: a new temporally-explicit model. medRxiv. 2020. doi:10.1101/2020.04.10.20061192
1310. Wilches Visbal JH, Castillo Pedraza MC. [Mathematical approach of the SIR epidemiological model for the comprehension of the containment measures against the Covid-19.]. Aproximacion Mat del Model Epidemiol SIR para la comprension las medidas contencion contra la COVID-19. 2020;94.
1311. William JA-B, Aveiro R, Telmo R, Fernández, María F, Diana C-P, et al. Progresión de casos de Coronavirus en Latinoamérica: Análisis comparativo a una semana de iniciada la pandemia en cada país. Kasmera. 2020;48.
1312. Willy R, Juan A, Jhony ADLC-V, Luis O, Maria V, Yovanna S, et al. Epidemiological characteristics of deaths from COVID-19: an analysis at almost three months of the first confirmed case in Peru. medRxiv. 2020. doi:10.1101/2020.11.05.20226639
1313. Wollenstein-Betech S, Silva AAB, Fleck JL, Cassandras CG, Paschalidis IC. Physiological and socioeconomic characteristics predict COVID-19 mortality and resource utilization in Brazil. PLoS One. 2020;15: e0240346. doi:http://dx.doi.org/10.1371/journal.pone.0240346
1314. Wollenstein-Betech S, Cassandras CG, Paschalidis IC. Personalized Predictive Models for Symptomatic COVID-19 Patients Using Basic Preconditions: Hospitalizations, Mortality, and the Need for an ICU or Ventilator. medRxiv Prepr Serv Heal Sci. 2020. doi:https://dx.doi.org/10.1101/2020.05.03.20089813
1315. Ximenes Neto FRG, Araújo CR de C e, Silva RCC da, Ribeiro MA, Sousa LA de, Serafim TF, et al. Coordination of care, surveillance and monitoring of cases of COVID-19 in primary health care. Enferm foco. 2020;11: 239–245.
1316. Xu C, Dong Y, Yu X, Wang H, Tsamlag L, Zhang S, et al. Estimation of reproduction numbers of COVID-19 in typical countries and epidemic trends under different prevention and control scenarios. Front Med. 2020. doi:https://dx.doi.org/10.1007/s11684-020-0787-4
1317. Yáñez JA, Afshar Jahanshahi A, Alvarez-Risco A, Li J, Zhang SX. Anxiety, Distress, and Turnover Intention of Healthcare Workers in Peru by Their Distance to the Epicenter during the COVID-19 Crisis. Am J Trop Med Hyg. 2020. doi:10.4269/ajtmh.20-0800

1318. Yang HM, Lombardi Junior LP, Castro FFM, Yang AC. Mathematical model describing CoViD-19 in Sao Paulo, Brazil - evaluating isolation as control mechanism and forecasting epidemiological scenarios of release. Epidemiol Infect. 2020;148: e155. doi:https://dx.doi.org/10.1017/S0950268820001600

Hyun Mo Y, Luis Pedro Lombardi J, Fabio Fernandes Morato C, Ariana Campos Y. Mathematical model describing CoViD-19 in Sao Paulo State, Brazil - Evaluating isolation as control mechanism and forecasting epidemiological scenarios of release. medRxiv. 2020. doi:10.1101/2020.04.29.20084830

1319. Yaranay L-A, Javier M-N, Fabiola S-D, Rodolfo Á-J. La Inteligencia emocional en apoderados chilenos confinados por la COVID-19. 2020;9. doi:10.33210/CA.V9I2.320

1320. Yasmany Damián G-R, María Soledad S-M, Belizario Amador Z-T, Marlon Agusto C-R. Relación entre las restricciones del tránsito vehicular y las tendencias del COVID-19: un caso de estudio ecuatoriano. 2020;9. doi:10.33210/CA.V9I2.308

1321. Yige L, Eduardo AU, Jose Ramon Z. Efficacy of Localized Lockdowns in the SARS-CoV-2 Pandemic. medRxiv. 2020. doi:10.1101/2020.08.25.20182071

1322. Yomayusa N, Acevedo K, Reina A, Rincón K, Toloza C, Gomez O, et al. Clinical course, biomarkers, management and outcomes of patients hospitalised due to COVID-19 in Colombia. ResearchSquare. 2020. doi:10.21203/rs.3.rs-57978/v1

1323. You C, Yubo F, Chao Y, Xinmeng Z, Cheng G. Modeling COVID-19 Growing Trends to Reveal the Differences in the Effectiveness of Non-Pharmaceutical Interventions among Countries in the World. medRxiv. 2020. doi:10.1101/2020.04.22.20075846

1324. Yunier B-P, Zoylen F-F, Elizabeth J-P, Evelyn F-C, Boris CR-M. Gender and Fear of COVID-19 in a Cuban Population Sample. Int J Ment Health Addict. 2020. doi:10.1007/S11469-020-00343-8

1325. Yuri Castro R, Olenka Valenzuela T. Repercusiones de la pandemia de COVID 19 en la atención odontológica, una perspectiva de los odontólogos clínicos. 2020;19.

1326. Zambrano-Monserrate MA, Ruano MA. Has air quality improved in Ecuador during the COVID-19 pandemic? A parametric analysis. Air Qual Atmos Health. 2020; 1–10. doi:10.1007/s11869-020-00866-y

1327. Zavala-Flores E, Salcedo-Matienzo J. Pre-hospitalary medication in COVID-19 patients from a public hospital in Lima-Peru. Acta Medica Peru. 2020;37: 393–395. doi:http://dx.doi.org/10.35663/amp.2020.373.1277

1328. Zavarce Castillo C, Zavarce Castillo F. Modeling the trend and seasonal components of the SARS-COV2 in the Bolivarian Republic of Venezuela. Obs del Conoc Rev Espec Gestión Soc del Conoc. 2020;5: 46–56.

1329. Zavarce Castillo C, Zavarce Castillo F. Stochastic behavior of covid-19 in the Bolivarian Republic of Venezuela: persistence or antipersistence in infections? Obs del Conoc Rev Espec en Gestión Soc del Conoc. 2020;5: 91–110.

1330. Zhang SX, Sun S, Afshar Jahanshahi A, Alvarez-Risco A, Ibarra VG, Li J, et al. Developing and testing a measure of COVID-19 organizational support of healthcare workers - results from Peru, Ecuador, and Bolivia. Psychiatry Res. 2020;291: 113174. doi:https://dx.doi.org/10.1016/j.psychres.2020.113174

1331. Zhiwei D, Feng S, Bingyu L, Jing K, Yi Z, Yip P, et al. Predicting the Impact of the COVID-19 Pandemic for the Low- and Middle-Income Countries. medRxiv. 2020. doi:10.1101/2020.08.13.20167064

1332. Zhu D, Mishra SR, Han X, Santo K. Social distancing in Latin America during the COVID-19 pandemic: an analysis using the Stringency Index and Google Community Mobility Reports. J Travel Med. 2020. doi:10.1093/jtm/taaa125

1333. Zhu L, Liu X, Huang H, Avellan-Llaguno RD, Lazo MML, Gaggero A, et al. Meteorological impact on the COVID-19 pandemic: A study across eight severely affected regions in South America. Sci Total Environ. 2020;744: 140881. doi:https://dx.doi.org/10.1016/j.scitotenv.2020.140881

1334. Zimman S, Cura MJ, Luna PC, Echeverria CM, Mazzuoccolo LD. Impact of the COVID-19 Pandemic on Immunomodulatory and Immunosuppressive Therapies in Dermatology: Patient and Physician Attitudes in Argentina. Actas Dermosifiliogr. 2020. doi:https://dx.doi.org/10.1016/j.ad.2020.04.004

1335. Zimmerer KS, de Haan S. Informal food chains and agrobiodiversity need strengthening-not weakening-to address food security amidst the COVID-19 crisis in South America. Food Secur. 2020; 1–4. doi:10.1007/s12571-020-01088-x
1336. Zimmermann IR, Sanchez M, Brant J, Alves D. Demand for ICU beds by COVID-19 in the Federal District, Brazil: an analysis of the impact of social distance measures with Monte Carlo simulations. 2020.
1337. Zoilo Emilio Sr. G-B, Kiero Sr. G-P, Vahid Vahid Nouri K, Maria Isabel M, Luis Eduardo Sr. G, Luisa Marilia C-G, et al. COVID-19 pandemic and health worker stress: The mediating effect of emotional regulation. medRxiv. 2020. doi:10.1101/2020.06.19.20135574
1338. Zuniga-Moya JC, Norwood DA, Romero Reyes LE, Barrueto Saavedra E, Diaz R, Fajardo WC, et al. Epidemiology, outcomes and associated factors of COVID-19 RT-PCR confirmed cases in the San Pedro Sula Metropolitan Area, Honduras. Clin Infect Dis. 2020. doi:10.1093/cid/ciaa1188
1339. Zylberman V, Sanguineti S, Pontoriero A V, Higa S V, Cerutti ML, Morrone Seijo SM, et al. Development of a hyperimmune equine serum therapy for COVID-19 in Argentina. Desarro un suero equino hiperinmune para el Trat COVID-19 en Argentina. 2020;80 Suppl 3: 1–6.

#### Protocol registry of included studies

175. Lung Function, Exercise Capacity and Health-Related Quality of Life After Severe COVID-19. 2020. Available in https://clinicaltrials.gov/show/NCT04410107
176. M5049 Study in Participants With Coronavirus Disease 2019 (COVID-19) Pneumonia. 2020. Available in https://clinicaltrials.gov/show/NCT04448756
177. Mechanical Ventilation Strategy for Coronavirus Disease 2019 (COVID-19). 2020. Available in https://clinicaltrials.gov/show/NCT04497454
178. Melatonin Agonist on Hospitalized Patients With Confirmed or Suspected COVID-19. 2020. Available in https://clinicaltrials.gov/ct2/show/NCT04470297
179. Mental health affected by COVID-19. 2020. Available in http://www.ctri.nic.in/Clinicaltrials/pmaindet2.php?trialid=45764
180. Mental Health and Academic Performance in COVID-19. 2020. Available in https://clinicaltrials.gov/show/NCT04420416
181. Mental Health Impact of the COVID-19 Pandemic Among Migrants in Chile. 2020. Available in https://clinicaltrials.gov/ct2/show/NCT04497636
182. Mesenchymal Stem Cell for Acute Respiratory Distress Syndrome Due for COVID-19. 2020. Available in https://clinicaltrials.gov/show/NCT04416139
183. Methylprednisolone in the Treatment of Patients With Signs of Severe Acute Respiratory Syndrome in Covid-19. 2020. Available in https://clinicaltrials.gov/show/NCT04343729
184. Microvascular Flow and Reactivity in Patients Presenting in the Acute Phase of COVID-19.. 2020. Available in https://clinicaltrials.gov/show/NCT04406545
185. Muscle Mass and Strength as Predictors of Time to Discharge in Patients With COVID-19. 2020. Available in https://clinicaltrials.gov/show/NCT04406233
186. Needs, Perceptions and Acute Stress of Healtcare Workers Caring for COVID-19 Patients in South America. 2020. Available in https://clinicaltrials.gov/ct2/show/NCT04486404
187. NestaCell® Mesenchymal Stem Cell to Treat Patients With Severe COVID-19 Pneumonia (HOPE). 2020. Available in https://clinicaltrials.gov/ct2/show/NCT04315987
188. Northeast COVID-19 and Pregnancy Study Group. 2020. Available in https://regroup-https://clinicaltrials.gov/ct2/keydates/NCT04462367
189. Novel Coronavirus infection among health workers. 2020. Available in http://www.ensaiosclinicos.gov.br/rg/RBR-7nky44/
190. Oncological Surgery in Times of COVID-19: Effectiveness of Preoperative Screening for Sars-Cov-2. 2020. Available in https://clinicaltrials.gov/show/NCT04434261
191. One year follow-ups of Chilean patients with COVID-19. 2020. Available in 10.1186/ISRCTN16865246
192. Online meditation as an aid in reducing symptoms of Depression, Anxiety and Psychological Stress in Resident Physicians during COVID-19 pandemic in Brazil. 2020. Available in https://ensaiosclinicos.gov.br/rg/RBR-2w7ghc/
193. Online Positive Psychology Intervention for Depression and Anxiety Due to COVID-19. 2020. Available in https://clinicaltrials.gov/ct2/show/NCT04468893
194. Opaganib, a Sphingosine Kinase-2 (SK2) Inhibitor in COVID-19 Pneumonia. 2020. Available in https://clinicaltrials.gov/ct2/show/NCT04467840
195. Outcomes Of Cancer Surgery During COVID-19 Pandemic. 2020. Available in http://www.ctri.nic.in/Clinicaltrials/pmaindet2.php?trialid=43628
196. Outcomes of Patients With COVID-19 in the Intensive Care Unit. 2020. Available in https://clinicaltrials.gov/show/NCT04336345
197. Outpatient Treatment With CoVid-19 With Prexablu. 2020. Available in https://clinicaltrials.gov/ct2/show/NCT04619290
198. P2Et Extract in the Symptomatic Treatment of Subjects With COVID-19. 2020. Available in https://clinicaltrials.gov/ct2/show/NCT04410510
199. Pegylated Interferon - a2b With SARSCoV- 2 (COVID-19). 2020. Available in https://clinicaltrials.gov/show/NCT04480138
200. Performance Evaluation of BCG vs COVID-19. 2020. Available in https://clinicaltrials.gov/ct2/show/NCT04362124

### 5. Excluded studies

#### Articles excluded with their reason

| Reference | Title | Exclusion reason |
|---|---|---|
| A 2020 | Diffusion processes in multilayer transportation networks: the flight of the Coronavirus | Not focused in LAC countries |
| Ã‘amendys-Silva 2020 | Health care workers with COVID-19 in Mexico | Not empirical primary research |
| AbrahÃ£odeCastro 2020 | Social protection in times of Covid-19 | Not empirical primary research |
| Abrantes 2020 | Oral cancer diagnosis during the COVID-19 pandemic in an oral pathology laboratory in Rio de Janeiro, Brazil | Not empirical primary research |
| Abu-Rayash 2020 | Analysis of mobility trends during the COVID-19 coronavirus pandemic: Exploring the impacts on global aviation and travel in selected cities | Not focused in LAC countries |
| Accinelli 2020 | At High Altitude COVID-19 Is Less Frequent: The Experience of Peru | Not empirical primary research |
| Adam 2020 | Networks in a World Unknown: Public WhatsApp Groups in the Venezuelan Refugee Crisis | Not produced in response to the COVID-19 pandemic |
| Aguilar-Guerra 2020 | Mobilizing Primary Health Care:Cuba's Powerful Weapon against COVID-19. | Not empirical primary research |
| Aisa-Alvarez 2020 | A Randomized clinical trial of antioxidant therapy in patients with septic shock. Reference study to propose adjuvant therapy in patients with critical organic damage by COVID-19 | Not produced in response to the COVID-19 pandemic |
| Albuquerque 2020 | Overview of confirmed cases of COVID-19 in five countries facing community transmission | Not focused in LAC countries |
| Albuquerque 2020 | Planejamento operacional durante a pandemia de Covid-19: comparaÃ§Ã£o entre recomendaÃ§Ãµes da OrganizaÃ§Ã£o Mundial de SaÃºde e o Plano de de ContigÃªncia Nacional | Not empirical primary research |
| AlcidesEduardodosReis 2020 | Viral Surveillance: Governing Social Isolation in SÃ£o Paulo, Brazil, During the COVID-19 Pandemic | Not empirical primary research |
| Alexandre 2020 | A COVID-19 na Baixada Fluminense: Colapso e apreensÃ£o a partir da periferia metropolitana do Rio de Janeiro | Not focused in LAC countries |
| Almeida-Espinosa 2020 | COVID-19: Implications of SARS-CoV-2 in Colombia | Not empirical primary research |
| Alonso 2020 | Recurrence of SARS-CoV-2 infection with a more severe case after mild COVID-19, reversion of RT-qPCR for positive and late antibody response: case report | Not empirical primary research |
| Alvarado-Moreno 2020 | COVID-19 and Dysfunctional Endothelium: The Mexican Scenario. | Not empirical primary research |
| Andrew 2020 | Rhythmic components of COVID-19 daily cases in various countries | Not focused in LAC countries |
| Angus 2020 | The Randomized Embedded Multifactorial Adaptive Platform for Community-acquired | Not focused in LAC countries |

| | | |
|---|---|---|
| | Pneumonia (REMAP-CAP) Study: rationale and Design | |
| Ankarali 2020 | Modeling and short-term forecasts of indicators for COVID-19 outbreak in 25 countries at the end of march | Not focused in LAC countries |
| Anonymous 2020 | COVID-19 in Argentina, what is the current epidemiological situation? | Not empirical primary research |
| AntÃºnez-Montes 2020 | Feasibility of Lung Point-of-Care Ultrasound for Patients With COVID-19 in Air Medical Transport: Triage of 2 Initially Suspected Cases on Mexico's Front Line | Not empirical primary research |
| Aquino 2020 | Social distancing measures to control the COVID-19 pandemic: potential impacts and challenges in Brazil. | Not empirical primary research |
| Araujo 2020 | Factors associated with Severe Acute Respiratory Syndrome in a Brazilian central region | Not produced in response to the COVID-19 pandemic |
| Araujo 2020 | In defense of the Unified Health System in the context of SARS-CoV-2 pandemic | Not empirical primary research |
| Araujo 2020 | Nursing management in an accredited public general hospital in the response to the COVID-19 pandemic | Not empirical primary research |
| Araujo-Dos-Santos 2020 | Nursing Comiittee to Coping with COVID-19 in Bahia | Not empirical primary research |
| Armandoda 2020 | Regression Polynomial Analysis of the COVID-19 Epidemics: Some Initial Findings | Not focused in LAC countries |
| Artaza 2020 | COVID-19: Lessons for Chile | Not empirical primary research |
| Atique 2020 | A nursing informatics response to COVID-19: perspectives from five regions of the world. | Not empirical primary research |
| Atkinson 2020 | SARS-CoV-2 shedding and infectivity | Not empirical primary research |
| Avancini 2020 | Absence of specific cutaneous manifestations of SARS-Cov-2 in a reference center in Brazil | Not empirical primary research |
| Avaneesh 2020 | SEIHCRD Model for COVID-19 spread scenarios, disease predictions and estimates the basic reproduction number, case fatality rate, hospital, and ICU beds requirement | Not focused in LAC countries |
| Aveiro-Robalo 2020 | [Learning at home during the COVID-19 pandemic: An initiative of Latin American students]. | Not empirical primary research |
| Avila-Castro 2020 | Proposal for the management of COVID-19-associated coagulopathy in children | Not focused in LAC countries |
| Ayala 2020 | Pregnant woman with moderate COVID 19 and care process in the newborn | Not empirical primary research |
| Azarpazhooh 2020 | COVID-19 pandemic and burden of non-communicable diseases: An ecological study on data of 185 countries | Not focused in LAC countries |
| Balilla 2020 | Characterizing the Impact of COVID-19 on the Global Stock Indices through Comparative Analysis | Not focused in LAC countries |

| | | |
|---|---|---|
| Baptista 2020 | Contributions of residents from multiple specializations in managing the COVID-19 pandemic in the largest public hospital Brazil | Not empirical primary research |
| Barajas-Ochoa 2020 | Retos para la educacion medica en Mexico en los tiempos del COVID-19, Challenges for medical education in Mexico in the time of COVID-19 | Not empirical primary research |
| Barberia 2020 | Political and institutional perils of Brazil's COVID-19 crisis | Not empirical primary research |
| Barbero 2020 | SARS-CoV-2 in pregnancy: characteristics and outcomes of hospitalized and non-hospitalized women due to COVID-19 | Not focused in LAC countries |
| Barrios 2020 | Volver a empezar: cirugÃa electiva durante la pandemia del SARS-CoV2. Recomendaciones desde la AsociaciÃ³n Colombiana de CirugÃa | Not empirical primary research |
| Barros 2020 | Brazilian Nursing Process Research Network contributions for assistance in the COVID-19 pandemic | Not empirical primary research |
| BarrosdeSouza 2020 | ESTRATÃ‰GIA MUSICAL PARA CUIDAR DE DISCENTES DE ENFERMAGEM: EXPERIÃŠNCIA NO ENFRENTAMENTO DA COVID-19 | Not empirical primary research |
| Bautista-Molano 2020 | Exploring the Impact of COVID-19 in Latin America | Not empirical primary research |
| Behrens 2020 | COVID-19: IFSO LAC Recommendations for the Resumption of Elective Bariatric Surgery | Not empirical primary research |
| Belarmino 2020 | Collaborative practices from health care teams to face the covid-19 pandemic | Not empirical primary research |
| BelizÃ¡n 2020 | [Reflections on translational health research and the COVID-19 case in Argentina] | Not empirical primary research |
| Ben-HurFrancisco 2020 | Universal scaling law for COVID-19 propagation in urban centers | Not focused in LAC countries |
| Betonico 2020 | Challenges in COVID-19 medical response: A nephrology perspective | Not empirical primary research |
| BispoJunior 2020 | Community participation in the fight against COVID-19: between utilitarianism and social justice | Not empirical primary research |
| Bitencourt 2020 | Nurse's protagonism in structuring and managing a specific unit for covid-19 | Not empirical primary research |
| Blanc 2020 | What the world could learn from the Haitian resilience while managing COVID-19 | Not empirical primary research |
| Blukacz 2020 | COVID-19: leaving no one behind in Latin America | Not empirical primary research |
| Boccalatte 2020 | Recomendaciones para prevenir la infecciÃ³n por COVID-19 en cirujanos de cabeza y cuello: el comienzo de la pandemia en Argentina | Not empirical primary research |
| BomfimRibeiro 2020 | The Brazilian ongoing intervention studies for COVID-19: Clinical trials protocols | Not focused in LAC countries |
| Boni 2020 | Websurveys nos tempos de COVID-19, Web surveys in the time of COVID-19 | Not empirical primary research |
| Bonifaz 2020 | Of viruses, coronaviruses and imagination | Not empirical primary research |

| | | |
|---|---|---|
| Bonilla-Aldana 2020 | Importance of the One Health approach to study the SARS-CoV-2 in Latin America | Not empirical primary research |
| Bonilla-Aldana 2020 | Una nueva zoonosis viral de preocupaciÃ³n global: COVID-19, enfermedad por coronavirus 2019 | Not empirical primary research |
| Borges 2020 | Military Nursing in "Operation Return to Brazil": aeromedical evacuation in the coronavirus pandemic | Not empirical primary research |
| Branco 2020 | Sus hospital emergency service: service flows for suspected or confirmed patients for COVID-19 | Not empirical primary research |
| BrandÃ£o 2020 | Clinical simulation strategies for knowledge integration relating to initial critical recognition and management of COVID-19 for use within continuing education and health-related academia in Brazil: a descriptive study | Not empirical primary research |
| Brasil 2020 | AtenÃ§Ã£o Ã saÃºde do recÃ©m-nascido no contexto da infecÃ§Ã£o do novo CoronavÃrus (SARS-CoV-2) | Not empirical primary research |
| Brasil 2020 | AtencÃŒÂ§aÃŒÂƒo aÃŒÂ€ sauÃŒÂ• de do receÃŒÂ• m-nascido no contexto da infeccÃŒÂ§aÃŒÂƒo pelo novo coronaviÃŒÂ• rus (SARS-CoV-2) | Not empirical primary research |
| Brasil 2020 | Cabine de proteÃ§Ã£o para o tratamento de pacientes com COVID-19 | Not focused in LAC countries |
| Brasil 2020 | Como registrar no e-SUS APS quando for realizado uma teleconsulta | Not empirical primary research |
| Brasil 2020 | Condutas para a doaÃ§Ã£o de leite materno aos bancos de leite humano e postos de coleta de leite humano no contexto da infecÃ§Ã£o Covid 19 causada pelo novo CoronavÃrus (SARS-CoV-2) | Not empirical primary research |
| Brasil 2020 | Condutas para a doaÃ§Ã£o de leite materno aos bancos de leite humano e postos de coleta de leite humano no contexto da infecÃ§Ã£o pelo CoronavÃrus (SARS-CoV-2) | Not empirical primary research |
| Brasil 2020 | CoronavÃrus CoViD-19: Atendimento odontolÃ³gico no SUS | Not empirical primary research |
| Brasil 2020 | CoronavÃrus Covid-19: Fast-track de teleatendimento para a atenÃ§Ã£o primÃ¡ria: fluxo rÃ¡pido - versÃ£o 6 | Not empirical primary research |
| Brasil 2020 | CoronavÃrus Covid-19: Fast-track de teleatendimento para a atenÃ§Ã£o primÃ¡ria: fluxo rÃ¡pido - versÃ£o 8 | Not empirical primary research |
| Brasil 2020 | CoronavÃrus Covid-19: Fast-track de teleatendimento para a atenÃ§Ã£o primÃ¡ria: fluxo rÃ¡pido - versÃ£o 9 | Not empirical primary research |
| Brasil 2020 | CoronavÃrus CoViD-19: Fast-Track para a atenÃ§Ã£o primÃ¡ria em locais com transmissÃ£o comunitÃ¡ria fluxo rÃ¡pido - versÃ£o 8 | Not empirical primary research |
| Brasil 2020 | CoronavÃrus CoViD-19: Fast-Track para a atenÃ§Ã£o primÃ¡ria em locais com | Not empirical primary research |

| | | |
|---|---|---|
| | transmissÃ£o comunitÃ¡ria fluxo rÃ¡pido - versÃ£o 9 | |
| Brasil 2020 | CoronavÃrus CoViD-19: Fast-track para a atenÃ§Ã£o primÃ¡ria em locais com transmissÃ£o comunitÃ¡ria fluxo rÃ¡pido: versÃ£o 6 | Not empirical primary research |
| Brasil 2020 | CoronavÃrus CoViD-19: Fluxo de manejo clÃ-nico de gestantes na atenÃ§Ã£o especializada | Not empirical primary research |
| Brasil 2020 | CoronavÃrus Covid-19: Fluxo de manejo clÃnico na atenÃ§Ã£o primÃ¡ria Ã saÃºde em transmissÃ£o comunitÃ¡ria: versÃ£o 9 | Not empirical primary research |
| Brasil 2020 | CoronavÃrus CoViD-19: Fluxo de manejo clÃ-nico pediÃ¡trico na atenÃ§Ã£o especializada | Not empirical primary research |
| Brasil 2020 | CoronavÃrus COVID-19: Nota TÃ©cnica: saÃºde mental e apoio e psicossocial na atenÃ§Ã£o especializada | Not empirical primary research |
| Brasil 2020 | CoronavÃrus Covid-19: OrientaÃ§Ãµes sobre amamentaÃ§Ã£o | Not empirical primary research |
| Brasil 2020 | CoronavÃrus Covid-19: RecomendaÃ§Ãµes Programa Academia da SaÃºde | Not empirical primary research |
| Brasil 2020 | Evitar transmissÃ£o intradomiciliar da doenÃ§a pelo CoronavÃrus (COVID-19) | Not empirical primary research |
| Brasil 2020 | Fluxo de atendimento no hospital de referÃªncia para paciente referenciado de outros serviÃ§os de saÃºde | Not empirical primary research |
| Brasil 2020 | Fluxo de pacientes com sintomas respiratÃ³rios em unidades de urgÃªncia nÃ£o hospitalares | Not empirical primary research |
| Brasil 2020 | Fluxograma para atendimento e detecÃ§Ã£o precoce de COVID-19 em hospital de referÃªncia para indivÃduos por demanda espontÃ¢nea | Not empirical primary research |
| Brasil 2020 | Fluxograma para atendimento e detecÃ§Ã£o precoce de COVID-19 em pronto atendimento UPA 24 horas e unidade hospitalar nÃ£o definida como referÃªncia | Not empirical primary research |
| Brasil 2020 | InfecÃ§Ã£o COVID-19 e os riscos Ã s mulheres no ciclo gravÃdico-puerperal | Not empirical primary research |
| Brasil 2020 | Novo CoronavÃrus: atendimento a pessoas com suspeita de infecÃ§Ã£o pelo novo coronavÃrus (2019-nCoV) na AtenÃ§Ã£o PrimÃ¡ria Ã SaÃºde | Not empirical primary research |
| Brasil 2020 | OrientaÃ§Ãµes do MinistÃ©rio da SaÃºde para manuseio medicamentoso precoce de pacientes com diagnÃ³stico da COVID-19 | Not empirical primary research |
| Brasil 2020 | OrientaÃ§Ãµes gerais sobre a atuaÃ§Ã£o do ACS frente Ã pandemia de COVID-19 e os registros a serem realizados no e-SUS APS | Not empirical primary research |
| Brasil 2020 | OrientaÃ§Ãµes para o registro do FAST-TRACK da abordagem sindrÃ´mica de SÃndrome Gripal nos atendimentos aos cidadÃ£os com suspeita de H1N1 e COVID-19 no ProntuÃ¡rio EletrÃ´nico do CidadÃ£o (PEC) do e-SUS APS | Not empirical primary research |

| | | |
|---|---|---|
| Brasil 2020 | PrevenÃ§Ã£o ao Covid-19 no Ã¢mbito das equipes de consultÃ³rios na rua | Not empirical primary research |
| Brasil 2020 | RecomendaÃ§Ãµes aos Agentes de Combate a Endemias (ACE) para adequaÃ§Ã£o das aÃ§Ãµes de vigilÃ¢ncia e controle de zoonoses frente Ã atual situaÃ§Ã£o epidemiolÃ³gica referente ao CoronavÃrus (COVID-19) | Not empirical primary research |
| Brasil 2020 | RecomendaÃ§Ãµes para preveÃ§Ã£o, manejo e controle de casos de COVID-19 no Ã¢mbito do sistema socioeducativo | Not empirical primary research |
| Brasil 2020 | SaÃºde mental e atenÃ§Ã£o psicossocial na pandemia Covid-19: recomendaÃ§Ãµes gerais | Not empirical primary research |
| Brasil 2020 | SaÃºde mental e atenÃ§Ã£o psicossocial na pandemia Covid-19: recomendaÃ§Ãµes para gestores | Not empirical primary research |
| Brasil 2020 | SuspensÃ£o temporÃ¡ria das consultas eletivas e atividades nos centros de referÃªncia da pessoa idosa em razÃ£o da contenÃ§Ã£o de aglomeraÃ§Ã£o em populaÃ§Ã£o de risco para o CoronavÃrus (SARS-CoV-2) | Not empirical primary research |
| Brasil 2020 | UtilizaÃ§Ã£o do gÃ¡s ozÃ´nio e da ozonioterapia no combate Ã disseminaÃ§Ã£o da COVID-19 submetidas pela Sociedade Brasileira de Ozonioterapia MÃ©dica (SOBOM) | Not focused in LAC countries |
| Bravo 2020 | Mortality from respiratory infections and chronic non-communicable diseases before the covid-19 pandemic in cali, colombia | Not produced in response to the COVID-19 pandemic |
| Brito 2020 | COVID-19 in the Americas: Who's Looking After Refugees and Migrants? | Not empirical primary research |
| Brito 2020 | How Brazil Is Dealing with COVID-19 Pandemic Arrival Regarding Elective Gynecological Surgeries. | Not empirical primary research |
| Burki 2020 | COVID-19 in Latin America. | Not empirical primary research |
| Busnardo 2020 | A multidisciplinary approach to prevent and treat pressure sores in proned COVID-19 patients at a quaternary university hospital | Not empirical primary research |
| CÃ³rdova 2020 | Ciencia y TecnologÃa en Clave de Post-Pandemia. Una mirada inacabada | Not empirical primary research |
| CÃ³rdova 2020 | El Coronavirus y la pandemia: una crÃ³nica sociosimbÃ³lica | Not empirical primary research |
| CaÃ±eteRojas 2020 | Social performance of the specialist in Comprehensive General Medicine in the confrontation with COVID-19 | Not empirical primary research |
| Caballero 2020 | Political will and intersectorality to confront COVID-19 in Cuba | Not empirical primary research |
| Cabanillas-Rojas 2020 | BEHAVIOR AND SPREAD OF COVID-19 IN PERU: A FRAMEWORK FOR THE DESIGN BEHAVIORAL PUBLIC HEALTH INTERVENTIONS | Not empirical primary research |

| | | |
|---|---|---|
| Cabrera 2020 | Letter to the Editor: Is COVID-19 the Cause of Delayed Surgical Treatment of Spine Trauma in Latin America?. | Not empirical primary research |
| Caceres-Bernaola 2020 | First deceased by covid-19 in Peru | Not empirical primary research |
| Cadena-Estrada 2020 | Nursing before COVID-19, a key point for the prevention, control and mitigation of the pandemic. | Not empirical primary research |
| Caetano 2020 | Challenges and opportunities for telehealth during the COVID-19 pandemic: ideas on spaces and initiatives in the Brazilian context. | Not focused in LAC countries |
| Caldera-Villalobos 2020 | The Coronavirus Disease (COVID-19) Challenge in Mexico: A Critical and Forced Reflection as Individuals and Society | Not empirical primary research |
| Calderon-Garciduenas 2020 | Environmental Nanoparticles, SARS-CoV-2 Brain Involvement, and Potential Acceleration of Alzheimer's and Parkinson's Diseases in Young Urbanites Exposed to Air Pollution | Not empirical primary research |
| Camarano 2020 | Depending on the income of older adults and the coronavirus: orphans or newly poor? | Not empirical primary research |
| Campos 2020 | Burden of disease from COVID-19 and its acute and chronic complications: reflections on measurement (DALYs) and prospects for the Brazilian Unified National Health System | Not empirical primary research |
| Campos 2020 | Carga de doenÃ§a da COVID-19 e de suas complicaÃ§Ãµes agudas e crÃ´nicas: reflexÃµes sobre a mensuraÃ§Ã£o (DALY) e perspectivas no Sistema Ãšnico de SaÃºde | Not empirical primary research |
| Campos 2020 | Ion torrent-based nasopharyngeal swab metatranscriptomics in COVID-19. | Not empirical primary research |
| Canals 2020 | [Concepts for good decision-making in the COVID-19 pandemic in Chile] | Not empirical primary research |
| CanalsLambarri 2020 | Learning from the COVID-19 pandemic: Concepts for good decision-making | Not empirical primary research |
| CarboniBisso 2020 | COVID-19 in the intensive care unit. Analysis of the experience during the first month of pandemic | Not empirical primary research |
| Carda 2020 | COVID-19 pandemic. What should PRM specialists do? A clinician's perspective. | Not focused in LAC countries |
| Cardoso 2020 | The role of patient safety service in the fight against COVID-19 in a hospital | Not empirical primary research |
| Carlos 2020 | Increased risk for COVID-19 among Migrants from Latin-America, Caribbean, and Sub-Saharan Africa living in Spain | Not focused in LAC countries |
| Carmona 2020 | Transforming operating rooms into intensive care units and the versatility of the physician anesthesiologist during the COVID-19 crisis. | Not empirical primary research |
| Carneiro 2020 | Impact of the COVID-19 Pandemic on the Urologist's clinical practice in Brazil: a management guideline proposal for low- and middle-income countries during the crisis period. | Not empirical primary research |
| Carnut 2020 | Coronavirus, Capitalism in Crisis and the Perversity of Public Health in Bolsonaro's Brazil | Not empirical primary research |

| | | |
|---|---|---|
| Carolina 2020 | Chatting with my theoretical frameworks: Live sessions on Instagram as an opportunity to reflect in times of coronavirus | Not empirical primary research |
| Carrer 2020 | A COVID-19 na AmÃ©rica Latina e suas repercussÃµes para a odontologia | Not empirical primary research |
| CarvajalBaÃ±ados 2020 | Small is not beautiful: minimalizaciÃ³n de la salud pÃºblica enfrente de coronavirus | Not empirical primary research |
| Carvalho 2020 | Ciencia em tempos de pandemia, Science during the pandemic | Not empirical primary research |
| Carvalho 2020 | Fast-track COVID-19 em CSP, Fast-track for COVID-19 at CSP | Not empirical primary research |
| Carvalho 2020 | HOW IS BRAZIL FACING THE CRISIS OF FOOD AND NUTRITION SECURITY DURING THE COVID-19 PANDEMIC? | Not empirical primary research |
| Carvalho 2020 | Potential of Solar UV Radiation for Inactivation of Coronaviridae Family Estimated from Satellite Data | Not focused in LAC countries |
| Carvalho 2020 | Use of telemedicine to combat the COVID-19 pandemic in Brazil | Not empirical primary research |
| Casadevall 2020 | A Randomized Trial of Convalescent Plasma for COVID-19 - Potentially Hopeful Signals | Not focused in LAC countries |
| Castaneda-Millan 2020 | Organ Donation and Transplantation in the Midst of the COVID-19 Pandemic: Consequences and Challenges for an Emerging Country | Not empirical primary research |
| Castillo-Lopez 2020 | Olfactory Dysfunction in a Mexican Population Outside of COVID-19 Pandemic: Prevalence and Associated Factors (the OLFAMEX Study) | Not produced in response to the COVID-19 pandemic |
| Cavalcante 2020 | Como a PolÃtica Nacional de Controle do Tabaco Pode Ajudar no Enfrentamento da Pandemia da Covid-19? | Not empirical primary research |
| Cayo-Rojas 2020 | La empatÃa en la educaciÃ³n mÃ©dica: una oportunidad despuÃ©s de la crisis por COVID-19 | Not empirical primary research |
| Cenat 2020 | The vulnerability of low-and middle-income countries facing the COVID-19 pandemic: The case of Haiti. | Not empirical primary research |
| ChacÃ³n 2020 | El SARS-CoV-2 desde una perspectiva ecolÃ³gica | Not empirical primary research |
| Chahuan 2020 | Management plan for breast cancer during the COVID-19 pandemic. A single-institution alternative to treat early breast cancer patients in a short time | Not produced in response to the COVID-19 pandemic |
| Charlier 2020 | Is COVID-19 being used as a weapon against Indigenous Peoples in Brazil? | Not empirical primary research |
| Chavez 2020 | COVID-19: Antiviral treatment in mild cases could be useful at this time | Not empirical primary research |
| Chen 2020 | COVID-19 Pandemic Imperils Weather Forecast | Not focused in LAC countries |
| Chew 2020 | Ethnically diverse mutations in PIEZO1 associate with SARS-CoV-2 positivity | Not focused in LAC countries |
| Chiaravalloti 2020 | The emotional impact of the COVID-19 pandemic on individuals with progressive multiple sclerosis | Not focused in LAC countries |

| | | |
|---|---|---|
| Chiotos 2020 | Multicenter initial guidance on use of antivirals for children with COVID-19/SARS-CoV-2. | Not focused in LAC countries |
| Chirinos 2020 | Regarding the MINSA recommendations on the pharmacological treatment of COVID-19 in Peru | Not empirical primary research |
| Christoffel 2020 | Children's (in)visibility in social vulnerability and the impact of the novel coronavirus (COVID-19) | Not empirical primary research |
| Christoffel 2020 | Children's (in)visibility in social vulnerability and the impact of the novel coronavirus (COVID-19) | Not empirical primary research |
| Christopher 2020 | Puerto Rico Health System Resilience After Hurricane Maria: Implications for Disaster Preparedness in the COVID-19 Era | Not produced in response to the COVID-19 pandemic |
| ChristovamReisdosSantos 2020 | IGREJA UNIVERSAL E O CORONAVIRUS: a dimensÃ£o geossimbÃ³lica e negaÃ§Ã£o do isolamento social | Not empirical primary research |
| Cimerman 2020 | Deep impact of COVID-19 in the healthcare of Latin America: the case of Brazil. | Not empirical primary research |
| Cipitelli 2020 | SARS-CoV-2 diagnostic diary: from rumors to the first case. Early reports of molecular tests from the military research and diagnostic institute of Rio de Janeiro | Not empirical primary research |
| Claudio 2020 | Group testing for SARS-CoV-2 allows for up to 10-fold efficiency increase across realistic scenarios and testing strategies | Not empirical primary research |
| Claure-DelGranado 2020 | Renal Replacement Therapy for Acute Kidney Injury in COVID-19 Patients in Latin America | Not focused in LAC countries |
| Cleemput 2020 | Genome Detective Coronavirus Typing Tool for rapid identification and characterization of novel coronavirus genomes. | Not focused in LAC countries |
| Clososki 2020 | Tenofovir Disoproxil Fumarate: New Chemical Developments and Encouraging in vitro Biological Results for SARS-CoV-2 | Not focused in LAC countries |
| CoelhodaMaia 2020 | Lack of Personal Protective Equipment in Brazil | Not empirical primary research |
| Collado-Borrell 2020 | Features and functionalities of smartphone apps related to COVID-19 | Not focused in LAC countries |
| Concha 2020 | Diagnosis and treatment of pediatric patients with covid-19 infectious disease at the emergency room. A guideline for this pandemia | Not empirical primary research |
| ConselhoNacionaldeSecretÃ¡riosde 2020 | Second phase of the research "Evolution of the Prevalence of Infection by Covid-19 in Brazil: Population-Based Study" | Not empirical primary research |
| ConselhoNacionaldeSecretariasMunicipaisde 2020 | Guia orientador para o enfrentamento da pandemia COVID-19 na Rede de AtenÃ§Ã£o Ã SaÃºde | Not empirical primary research |
| Cordero-Escobar 2020 | Differences among the mechanical ventilators and those of anesthesia machine, in patients suffering COVID-19 | Not empirical primary research |
| Cortinez-O'Ryan 2020 | Could severe mobility and park use restrictions during the COVID-19 pandemic aggravate health inequalities? Insights and challenges from Latin America | Not empirical primary research |

| | | |
|---|---|---|
| Crabtree-Ramirez 2020 | Being a Latin American Woman in Science During the COVID-19 Pandemic | Not empirical primary research |
| Croda 2020 | Immediate Health Surveillance Response to COVID-19 Epidemic. | Not empirical primary research |
| Croda 2020 | Resposta imediata da VigilÃ¢ncia em SaÃºde Ã epidemia da COVID-19 | Not empirical primary research |
| Cruz 2020 | Therapeutic alternatives for COVID-19 | Not focused in LAC countries |
| Cunha 2020 | Actions and strategies of schools and nursing departments of federal universities front COVID-19 | Not focused in LAC countries |
| Cupertino 2020 | COVID-19 in Brazil: Epidemiological update and perspectives | Not empirical primary research |
| Curioso 2020 | Collaboration in times of COVID-19: the urgent need for open-data sharing in Latin America | Not empirical primary research |
| Curioso 2020 | The role of telehealth in the fight against COVID-19 and the evolution of the Peruvian regulatory framework | Not empirical primary research |
| DÃaz 2020 | Manifestaciones gastrointestinales de pacientes infectados con el nuevo Coronavirus SARS-CoV-2 | Not empirical primary research |
| DÃazCastrillÃ³n 2020 | La incertidumbre en la apertura de los servicios de cirugÃa ambulatoria en Colombia | Not empirical primary research |
| DÃazRosario 2020 | GestiÃ³n de riesgos y condicionalidad polÃtica de la cooperaciÃ³n internacional en tiempos de Coronavirus en Venezuela | Not empirical primary research |
| DÃazRosario 2020 | Risk management and political conditionality of international cooperation in Venezuela during COVID-19 | Not empirical primary research |
| daCostaBelarmino 2020 | SaÃºde ocupacional da equipe de enfermagem obstÃ©trica intensiva durante a pandemia da Covid-19 | Not empirical primary research |
| Dammert-Elejalde 2020 | Covid-19: Socio-cultural influences and adherence to governmental strategies for social isolation in peru | Not empirical primary research |
| Dantas 2020 | Implications of social distancing in Brazil in the COVID-19 pandemic. | Not empirical primary research |
| Dantas 2020 | Implications of social distancing in Brazil in the pandemic period of COVID-19 | Not empirical primary research |
| daSilvaNoal 2020 | National Emergency Training in Mental Health and Psychosocial Care at COVID-19: an experience report | Not empirical primary research |
| Daumas 2020 | The role of primary care in the Brazilian healthcare system: limits and possibilities for fighting COVID-19 | Not empirical primary research |
| Davalos 2020 | Pandemics' historical role in creating inequality | Not empirical primary research |
| David 2020 | A student made MOOC for medical students during the SARS-CoV-2 pandemic | Not empirical primary research |
| David 2020 | Pandemics, crisis conjunctures, and professional practices: what is the role of nursing with regard to Covid-19? | Not empirical primary research |

| | | |
|---|---|---|
| David 2020 | Vulnerability After COVID-19 and the Response of a Developing City: The Case of Bogota, Colombia | Not empirical primary research |
| deAlmeida-Leite 2020 | Is hydroxychloroquine a safe and effective choice for COVID-19 prophylaxis | Not empirical primary research |
| deArruda 2020 | The double-edged sword of SARS-CoV-2 vs. market economy: a perspective of Brazilian dental providers | Not empirical primary research |
| deFreitasESilva 2020 | What are the factors influencing the COVID-19 outbreak in Latin America?. | Not empirical primary research |
| delaCruz-Vargas 2020 | PROTECTING HEALTH PERSONNEL in the COVID-19 PANDEMIC | Not empirical primary research |
| DelCastillo 2020 | Low-dose radiotherapy for COVID-19 pneumonia treatment: case report, procedure, and literature review | Not empirical primary research |
| deMouraVillela 2020 | Student engagement in a public health initiative in response to COVID-19 | Not empirical primary research |
| Dentzer 2020 | Disruptive Innovation in rural cancer care: Opening the doors amid the pandemic | Not empirical primary research |
| deOliveira 2020 | Neighborhood Effects and Urban Inequalities: The Impact of Covid-19 on the Periphery of Salvador, Brazil. | Not empirical primary research |
| deOliveiraAndrade 2020 | Covid-19 is causing the collapse of Brazil's national health service | Not empirical primary research |
| deOliveiraAndrade 2020 | Covid-19: Prisons exposed in Brazil's crisis | Not empirical primary research |
| deSandes-Freitas 2020 | Lessons from SARS-CoV-2 screening in a Brazilian Organ Transplant Unit. | Not empirical primary research |
| Devakumar 2020 | COVID-19: the great unequaliser | Not focused in LAC countries |
| Dias 2020 | Respiratory diseases in the TriÃ¢ngulo Mineiro: Epidemiological analysis and projective with the COVID-19 pandemic | Not produced in response to the COVID-19 pandemic |
| Diaz-Guio 2020 | Will the Colombian intensive care units collapse due to the COVID-19 pandemic?. | Not empirical primary research |
| Diaz-Quinonez 2020 | Emergence of novel coronavirus SARS-CoV2 in China and the response in Mexico. | Not empirical primary research |
| Dietz 2020 | Obesity and its Implications for COVID-19 Mortality | Not empirical primary research |
| Dimer 2020 | The COVID-19 pandemic and the implementation of telehealth in speech-language and hearing therapy for patients at home: an experience report. | Not empirical primary research |
| Dominguez-Varela 2020 | High mortality among health personnel with COVID-19 in Mexico | Not empirical primary research |
| Dong 2020 | A Call for Consistency in the Official Naming of the Disease Caused by Severe Acute Respiratory Syndrome Coronavirus 2 in Non-English Languages. | Not focused in LAC countries |
| Dongarwar 2020 | COVID-19 Pandemic: Marked Global Disparities in Fatalities According to Geographic Location and Universal Health Care. | Not focused in LAC countries |

| | | |
|---|---|---|
| dosSantos 2020 | Two million COVID-19 cases in Brazil | Not empirical primary research |
| Dourado 2020 | Adapting to the COVID-19 Pandemic: Continuing HIV Prevention Services for Adolescents Through Telemonitoring, Brazil. | Not empirical primary research |
| Duim 2020 | Caring for the Workforce of a Health System During the COVID-19 Epidemic in Brazil: Strategies of Surveillance and Expansion of Access to Care | Not empirical primary research |
| Dyer 2020 | Covid-19: Mexico acknowledges 50000 more deaths than official figures show | Not empirical primary research |
| Echeverria-Ibazeta 2020 | Epidemiological situation of covid-19 in South America | Not empirical primary research |
| EderCano 2020 | Faecal-oral transmission of SARS-CoV-2: a threat to Latin America? | Not empirical primary research |
| EdesaBiotech 2020 | A Randomized, Double-Blind, Placebo-Controlled Study to Evaluate the Safety and Efficacy of EB05 + SOC vs. Placebo + SOC in Adult Hospitalized Patients With Moderate to Severe COVID-19 Pneumonia | Not focused in LAC countries |
| EduardoAmaral 2020 | Input-Output Analysis of COVID-19: Methodology for Assessing the Impacts of Lockdown Measures | Not empirical primary research |
| Effenberger 2020 | Association of the COVID-19 pandemic with Internet Search Volumes: A Google TrendsTM Analysis. | Not focused in LAC countries |
| Elera-Fitzcarrald 2020 | COVID-19 and Its Potential Effect on Patients With Rheumatic Diseases in Latin America. | Not empirical primary research |
| EliseuJosÃ© 2020 | Monitoramento do CoronavÃrus (Covid-19) nos municÃpios do Rio Grande do Sul, Brasil | Not empirical primary research |
| EmanuelleFreitas 2020 | Racial health inequalities and the COVID-19 pandemic Desigualdades raciais em saÃºde e a pandemia da Covid-19/ Racial health inequalities and the COVID-19 pandemic Las desigualdades raciales en la salud y la pandemia del Covid-19 | Not empirical primary research |
| Emparan 2020 | Recomendaciones para el manejo de pacientes que requieren atenciÃ³n oftalmolÃ³gica durante la pandemia de SARS-CoV-2 | Not focused in LAC countries |
| Enoki-Minano 2020 | Danger of major outbreaks of COVID-19 in the Peruvian prison population | Not empirical primary research |
| ErickdaLuz 2020 | The Management (or Lack Thereof) of COVID-19 in Brazil: Implications for Human Rights and Public Health | Not empirical primary research |
| ErnandesGonÃ§alves 2020 | Manejo do cuidado e a educaÃ§Ã£o em saÃºde na atenÃ§Ã£o bÃ¡sica na pandemia do CoronavÃrus | Not empirical primary research |
| Escudero 2020 | La pandemia de Coronavirus SARS-CoV-2 (COVID-19): Situacion actual e implicaciones para Mexico, The SARS-CoV-2 (COVID-19) coronavirus pandemic: current situation and implications for Mexico | Not empirical primary research |

| | | |
|---|---|---|
| Espinel 2020 | Venezuelan migrants in Colombia: COVID-19 and mental health | Not empirical primary research |
| Espinosa-Uribe 2020 | Regarding "The orthopaedic and traumatology scenario during Covid-19 outbreak in Italy: chronicles of a silent war" a Mexican perspective | Not empirical primary research |
| Evaldo 2020 | A discrete-time-evolution model to forecast progress of Covid-19 outbreak | Not focused in LAC countries |
| Ezequiel 2020 | The COVID-19 Pandemic: A Call to Action for Health Systems in Latin America to Strengthen Quality of Care. | Not empirical primary research |
| FalcÃ£o 2020 | Strategy and tactics of the "Cine Luce" project in the pandemic context of Covid-19: leisure in focus | Not empirical primary research |
| Faria 2020 | OSCE 3D: a virtual clinical skills assessment tool for coronavirus pandemic times | Not empirical primary research |
| Farias 2020 | COVID-19 Therapies in Brazil: Should We Be Concerned with the Impacts on Aquatic Wildlife? | Not empirical primary research |
| Faruq 2020 | Effect of preventive actions and health care factors in controlling the outbreaks of COVID-19 pandemic | Not focused in LAC countries |
| Felix-Arellano 2020 | [Quick review: air pollution and morbi-mortality by Covid-19]. | Not focused in LAC countries |
| Ferigato 2020 | The Brazilian Government's mistakes in responding to the COVID-19 pandemic | Not empirical primary research |
| Fernandes 2020 | Covid-19 in Brazil: Learning How to Walk in the Dark Without Leaving Anything Behind | Not empirical primary research |
| Ferran 2020 | Hi Sigma, do I have the Coronavirus?: Call for a New Artificial Intelligence Approach to Support Health Care Professionals Dealing With The COVID-19 Pandemic | Not focused in LAC countries |
| Ferrante 2020 | Brazil's policies condemn Amazonia to a second wave of COVID-19 | Not empirical primary research |
| Ferreira 2020 | Plasma-based COVID-19 treatments in low- and middle-income nations pose a high risk of an HIV epidemic | Not empirical primary research |
| Ferreira-Filho 2020 | Contraception and reproductive planning during the COVID-19 pandemic. | Not focused in LAC countries |
| FigueroaPÃ©rez 2020 | La prÃ¡ctica estomatolÃ³gica en tiempos de coronavirus en Cuba | Not empirical primary research |
| Fischman 2020 | Debate: Mental health, social crisis and the COVID-19 pandemic in Chile | Not empirical primary research |
| Fontanella 2020 | Neurosurgical activity during COVID-19 pandemic: an expert opinion from China, South Korea, Italy, United Stated of America, Colombia and United Kingdom. | Not empirical primary research |
| Forero-Pena 2020 | The first pregnant woman with COVID-19 in Venezuela: Pre-symptomatic transmission. | Not empirical primary research |
| Fosker 2020 | COVID-19 and cancer care in Bermuda | Not empirical primary research |
| Franca 2020 | Deaths due to COVID-19 in Brazil: how many are there and which are being identified?. | Not empirical primary research |

| | | |
|---|---|---|
| Franzoi 2020 | Participation of nursing students in the Covid-19 pandemic | Not empirical primary research |
| Franzotti 2020 | Desempenho dos Ã• ndices de Gravidade na PrediÃ§Ã£o de ComplicaÃ§Ãµes PÃ³s-OperatÃ³rias de RevascularizaÃ§Ã£o MiocÃ¡rdica | Not produced in response to the COVID-19 pandemic |
| Fraser 2020 | COVID-19 strains remote regions of Peru. | Not empirical primary research |
| Freitas 2020 | COVID-19 and orthodontics in Brazil: What should we do? | Not empirical primary research |
| Freudenberg 2020 | Global Impact of COVID-19 on Nuclear Medicine Departments: An International Survey in April 2020 | Not focused in LAC countries |
| Fuentes 2020 | Los valores profesionales de las enfermeras (os) chilenas (os) en tiempos de crisis sanitaria por Covid-19 | Not empirical primary research |
| Fuenzalida 2020 | Recommendations for the implementation of pediatric respiratory support in COVID-19. Intensive kinesiology and respiratory therapy Chilean | Not empirical primary research |
| Gaete 2020 | Coronavirus: cuidar la vida desnuda sin desconocer la vida vivida | Not empirical primary research |
| Gaiarsa 2020 | Structural vulnerability of hospitals, cemeteries, and crematoriums of the city of SÃ£o Paulo to COVID-19 | Not focused in LAC countries |
| GallardoSÃ¡nchez 2020 | The postgraduate course in front of COVID-19 in Granma | Not empirical primary research |
| Gallegos 2020 | COVID-19 in Latin America: A bibliometric analysis of scientific publications in health | Not focused in LAC countries |
| Gaojun 2020 | Predication of Inflection Point and Outbreak Size of COVID-19 in New Epicentres | Not focused in LAC countries |
| GarcÃa-Rey 2020 | Colombia en tiempos de COVID-19 | Not empirical primary research |
| Garcia 2020 | Contextualizing the COVID-19 Era in Puerto Rico: Compounding Disasters and Parallel Pandemics | Not empirical primary research |
| Garcia 2020 | COVID-19 Response in Latin America | Not empirical primary research |
| Garcia 2020 | Infodemic: excess quantity to the detriment of quality of information about COVID-19 | Not empirical primary research |
| Garcia 2020 | IntervenÃ§Ãµes nÃ£o farmacolÃ³gicas para o enfrentamento Ã epidemia da COVID-19 no Brasil | Not empirical primary research |
| Garcia 2020 | Nonpharmaceutical interventions for tackling the COVID-19 epidemic in Brazil. | Not empirical primary research |
| Garcia 2020 | Sex, gender and race dimensions in COVID-19 research | Not empirical primary research |
| Garcia-Bolanos 2020 | COVID-19: It marks a before and after in Mexico, final point or period? | Not empirical primary research |
| Garcia-Silva 2020 | MEXICAN HUMAN LEUKOCYTE ANTIGEN ALLELES MIGHT PREDICT CLINICAL | Not empirical primary research |

| | | |
|---|---|---|
| | OUTCOME IN SARS-COV-2 INFECTED PATIENTS. | |
| Garrido 2020 | Impasses entre dignidade e saÃºde no manejo de cadÃ¡veres da COVID-19: identificar ou reconhecer? | Not empirical primary research |
| Garrido 2020 | RestriÃ§Ã£o de contato social e saÃºde mental na pandemia: possÃveis impactos das condicionantes sociais | Not empirical primary research |
| Garrido 2020 | Restriction of social contact and mental health in pandemic: possible impacts of social constraints | Not focused in LAC countries |
| Garrido 2020 | Stalemates between dignity and health in the handling of COVID-19 corpses: identify or recognize? | Not empirical primary research |
| Gaspari 2020 | The First 60 Days: Physical Therapy in a Neurosurgical Center Converted Into a COVID-19 Center in Brazil | Not empirical primary research |
| Gauna 2020 | Recomendaciones diagnÃ³sticas y terapÃ©uticas ante la Respuesta Inmune TrombÃ³tica Asociada a COVID-19 (RITAC) | Not empirical primary research |
| Gemelli 2020 | Management of COVID-19 Outbreak in Argentina: The Beginning. | Not empirical primary research |
| GerardoRuiz 2020 | Los presupuestos de la resoluciÃ³n del contrato por imposibilidad sobrevenida, en relaciÃ³n con la actual pandemia por COVID-19 en Bolivia (The Requirements for the Termination of Contract by Force Majeure in Relation to the Current Pandemic Due to COVID-19 in Bolivia) | Not empirical primary research |
| Germano 2020 | Coronavirus Disease 2019 (COVID-19) and Neurosurgery: Literature and Neurosurgical Societies Recommendations Update. | Not focused in LAC countries |
| Gilligan 2020 | Delays in the diagnosis of oral cancer due to the quarantine of COVID-19 in Cordoba, Argentina | Not empirical primary research |
| Giovani 2020 | Fighting the coronavirus: the window of opportunity for non-pharmacological interventions is narrow | Not focused in LAC countries |
| Giraldo 2020 | COVID-19 in Peru | Not empirical primary research |
| Godoy 2020 | Recommendations for pediatric chronic liver disease and liver transplant during COVID-19 | Not empirical primary research |
| GoiÃ¡s 2020 | A Hidroxicloroquina eÃŒÂ• eficaz e segura no tratamento da COVID-19? | Not empirical primary research |
| GoiÃ¡s 2020 | Atualiza informaÃ§Ãµes sobre o uso da Cloroquina como terapia adjuvante no tratamento de formas graves do COVID-19 | Not empirical primary research |
| GoiÃ¡s 2020 | COVID-19 em crianÃ§as e reabertura de escolas | Not empirical primary research |
| GoiÃ¡s 2020 | DispÃµe sobre recomendaÃ§Ãµes para investigaÃ§Ã£o e notificaÃ§Ã£o da COVID19, no SINAN, na ficha de acidente de trabalho, por ser considerada doenÃ§a ocupacional | Not empirical primary research |
| GoiÃ¡s 2020 | Manejo ambiental de dengue e situaÃ§Ã£o de pandemia do COVID-19 | Not empirical primary research |

| | | |
|---|---|---|
| GoiÃ¡s 2020 | Multimorbidade e COVID-19 | Not empirical primary research |
| GoiÃ¡s 2020 | OrientaÃ§Ãµes para a utilizaÃ§Ã£o de cloroquina e hidroxicloroquina no tratamento de pacientes com COVID-19 | Not empirical primary research |
| GoiÃ¡s 2020 | OrientaÃ§Ãµes para o atendimento de gestantes e puÃ©rperas diante da pandemia do COVID-19 | Not empirical primary research |
| GoiÃ¡s 2020 | RecomendaÃ§Ãµes dos Tutores Estaduais para a estratÃ©gia de AtenÃ§Ã£o Humanizada ao RecÃ©m-Nascido: MÃ©todo Canguru, frente aos desafios enfrentados diante da pandemia decorrente de COVID-19 | Not empirical primary research |
| GoiÃ¡s 2020 | RecomendaÃ§Ãµes gerais para implantaÃ§Ã£o de medidas de prevenÃ§Ã£o e controle de casos de COVID-19 em empresas no estado de GoiÃ¡s | Not empirical primary research |
| GoiÃ¡s 2020 | Sobre serviÃ§os de somatoconservaÃ§Ã£o, funerÃ¡rias, cemitÃ©rios e crematÃ³rios no manejo de corpos de vÃtimas de casos suspeitos ou confirmados por COVID-19 | Not empirical primary research |
| GoiÃ¡s 2020 | Uso de mÃ¡scaras pela populaÃ§Ã£o em geral | Not empirical primary research |
| Gois-Santos 2020 | Primary Health Care in Brasil in the times of COVID-19: changes, challenges and perspectives | Not empirical primary research |
| Goldani 2020 | Infectious diseases and the COVID-19 scenario in Brazil | Not empirical primary research |
| Goldfarb 2020 | Prevalence and Severity of Coronavirus Disease 2019 (COVID-19) Illness in Symptomatic Pregnant and Postpartum Women Stratified by Hispanic Ethnicity | Not focused in LAC countries |
| Gomes 2020 | Reduced demand for oral diagnosis during COVID-19: a Brazilian center experience | Not empirical primary research |
| Gomez-Marin 2020 | Pandemic COVID-19: A reflection on the impact for preparedness on the control of infectious diseases in Colombia | Not empirical primary research |
| Gomez-Rios 2020 | The effect of uncontrolled travelers and social distancing on the spread of novel coronavirus disease (COVID-19) in Colombia. | Not empirical primary research |
| GovernodoEstadode Santa 2020 | Guia de convÃvio responsÃ¡vel: regras do coronavÃrus | Not empirical primary research |
| Gozzer 2020 | COVID-19 and the need for action to improve the pandemic capabilities of Peru | Not produced in response to the COVID-19 pandemic |
| Grech 2020 | COVID-19 and potential global mortality - Revisited | Not empirical primary research |
| Grech 2020 | Unknown unknowns - COVID-19 and potential global mortality | Not empirical primary research |
| GringsdeOliveirada 2020 | Centro de oncologia e infusÃ£o: desafios da equipe interprofissional frente Ã pandemia CoronavÃrus | Not empirical primary research |
| GrossAlbornoz 2020 | Between dogma and reason: epidemic theories and social representations in the Covid-19 pandemic in Ecuador | Not empirical primary research |

| | | |
|---|---|---|
| Guatemala 2020 | Flujogramas de Vigilancia EpidemiolÃ³gica COVID-19: InvestigaciÃ³n de casos sospechosos comunitarios de COVID-19 y en servicios de salud no habilitados para toma de muestra | Not empirical primary research |
| Guatemala 2020 | Lineamientos para el manejo de los Centros de Operaciones de Emergencia Institucionales | Not empirical primary research |
| Guatemala 2020 | OrganizaciÃ³n de los servicios y preparaciÃ³n de la respuesta hospitalaria para el manejo de pacientes asintomÃ¡ticos leves ante el Nuevo COVID -19 | Not empirical primary research |
| GuimarÃ£esJr 2020 | Vulnerabilidade das microrregiÃµes do Estado de SÃ£o Paulo Ã pandemia do novo coronavÃrus (SARS-CoV-2) | Not empirical primary research |
| Gutierrez-Aguilar 2020 | [Model for a threshold of daily rate reduction of COVID-19 cases to avoid hospital collapse in Chile]. | Not empirical primary research |
| GutierrezCortes 2020 | Role of the Colombian geriatrician in the face of the pandemic by COVID-19 | Not empirical primary research |
| Hallak 2020 | Concise practice recommendations for the provision of andrological services and assisted reproductive technology for male infertility patients during the SARS-CoV-2 in Brazil | Not empirical primary research |
| Hallo 2020 | Perspective from Ecuador, the Second Country with More Confirmed Cases of Coronavirus Disease 2019 in South America: A Review. | Not empirical primary research |
| Hambleton 2020 | COVID-19 in the Caribbean small island developing states: lessons learnt from extreme weather events | Not empirical primary research |
| Haniel 2020 | Would the Lockdown Really Be Necessary for the Control of COVID-19 in Brazil? | Not empirical primary research |
| Hartman 2020 | COVID-19 Respiratory Failure: Targeting Inflammation on VV-ECMO Support. | Not empirical primary research |
| Hawas 2020 | Generated Time-series Prediction Data of COVID-19's Daily Infections in Brazil by Using Recurrent Neural Networks | Not focused in LAC countries |
| Hazarika 2020 | Modelling and forecasting of COVID-19 spread using wavelet-coupled random vector functional link networks | Not focused in LAC countries |
| Hazhir 2020 | Estimating the global spread of COVID-19 | Not focused in LAC countries |
| HenriqueAraÃºjodaSilva 2020 | The fragility of the Brazilian health system and social vulnerability in front of COVID-19 | Not empirical primary research |
| Henriquez-Marquez 2020 | Prevention and early identification of COVID-19 suspected cases at the first level of care in Central America | Not empirical primary research |
| HernÃ¡ndez-GarcÃa 2020 | Role of the medical Science Students at COVID-19: the example of Cuba | Not empirical primary research |
| Hernandez 2020 | Epidemiological Chronicle of the First Recovered Coronavirus Disease Patient From Panama: Evidence of Early Cluster Transmission in a High School of Panama City | Not empirical primary research |

| | | |
|---|---|---|
| Hernandez-Vasquez 2020 | COVID-19 in Peru: the need to pay attention to the high number of police deaths due to the pandemic. | Not empirical primary research |
| Herrera-Anazco 2020 | Medical education during the COVID -19 pandemic: Global initiatives for undergraduate, internship, and medical residency | Not empirical primary research |
| HervÃ© 2020 | Inverted covariate effects for mutated 2nd vs 1st waveCovid-19: high temperature spread biased for young | Not focused in LAC countries |
| Hetrick 2020 | Addressing pandemic-Intensified food insecurity | Not empirical primary research |
| Heuveline 2020 | Beyond Deaths per Capita: Three CoViD-19 Mortality Indicators for Temporal and International Comparisons. | Not focused in LAC countries |
| Hilton 2020 | Estimation of country-level basic reproductive ratios for novel Coronavirus (COVID-19) using synthetic contact matrices | Not focused in LAC countries |
| HipOlito 2020 | Unwrapping the long-term impacts of COVID-19 pandemic on Brazilian academic mothers: the urgency of short, medium, and long-term measures | Not empirical primary research |
| Hisaka 2020 | Global Comparison of Changes in the Number of Test-Positive Cases and Deaths by Coronavirus Infection (COVID-19) in the World. | Not focused in LAC countries |
| Hoagland 2020 | Telemedicine as a tool for PrEP delivery during the COVID-19 pandemic in a large HIV prevention service in Rio de Janeiro-Brazil. | Not empirical primary research |
| Homma 2020 | Vaccines for neglected and emerging diseases in Brazil by 2030: the "valley of death" and opportunities for RD&I in Vaccinology 4.0 | Not empirical primary research |
| Huamani 2020 | Propagation by COVID-19 at high altitude: Cusco case | Not empirical primary research |
| Huarcaya-Victoria 2020 | Mental health considerations about the COVID-19 pandemic | Not empirical primary research |
| Huarcaya-Victoria 2020 | Psychotic symptoms in COVID-19 infection: A case series from Lima, Peru | Not empirical primary research |
| Humerez 2020 | Mental health of brazilian nursing professionals in the context of the Covid-19 pandemic: action by the Nursing Federal Council | Not empirical primary research |
| Husaini 2020 | COVID-19: Belize's Success Story in Containing Community Spread Has Suffered a Setback! | Not empirical primary research |
| Ibanez 2020 | COVID-19 in older people with cognitive impairment in Latin America | Not empirical primary research |
| Ibarra-Nava 2020 | Mexico and the COVID-19 Response | Not empirical primary research |
| Idrovo 2020 | Long but unreal lockdowns in latin america. Comment on chen, y.t.; yen, y.f.; yu, s.h.; su, e.c. an examination on the transmission of covid-19 and the effect of response strategies: A comparative analysis. int. j. environ. res. public health 2020, 17, e5687 | Not empirical primary research |

| | | |
|---|---|---|
| Iglesias-Osores 2020 | COVID-19 in indigenous communities of Peru: Cases and accessibility to health services | Not empirical primary research |
| IgorBrenoBarbosade 2020 | Covid-19 e suas Redes de Conectividades no TerritÃ³rio Maranhense: Covid-19 y sus redes de conectividad en el territorio de Maranhense | Not empirical primary research |
| Ilie 2020 | Forecasting the Spreading of COVID-19 across Nine Countries from Europe, Asia, and the American Continents Using the ARIMA Models | Not focused in LAC countries |
| Ingram 2020 | Characterization of initial North American pediatric surgical response to the COVID-19 pandemic. | Not focused in LAC countries |
| Institut 2020 | COVID-19 Seroprevalence Study in French Guiana | Not focused in LAC countries |
| InstitutodeEfectividadClÃnicay 2020 | Barbijos y protectores faciales en poblaciÃ³n general en la pandemia por COVID-19 | Not focused in LAC countries |
| Isea 2020 | Simulando la dinÃ¡mica de transmisiÃ³n de pacientes coinfectados con la Covid-19 y dengue | Not focused in LAC countries |
| Isea 2020 | Simulating the transmission dynamics of patients coinfected with Covid-19 and Dengue | Not empirical primary research |
| Itiki 2020 | Fast deployment of COVID-19 disinfectant from common ethanol of gas stations in Brazil | Not empirical primary research |
| JÃ©ssicaKarolineAlves 2020 | PercepÃ§Ã£o do impacto emocional da equipe de enfermagem diante da pandemia de COVID-19: relato de experiÃªncia | Not empirical primary research |
| JadsonBelemde 2020 | IMPACTS OF SARS-COV-2 ON BRAZILIAN AGRIBUSINESS | Not empirical primary research |
| Jardim 2020 | COVID-19 experience among Brasil's indigenous people | Not empirical primary research |
| Jimenez-Ruiz 2020 | Manifestaciones neurologicas por COVID-19, Neurologic manifestations of COVID-19 | Not empirical primary research |
| Jimenez-Ruiz 2020 | Neurologic manifestations of COVID-19. | Not empirical primary research |
| Jinyan 2020 | Tax Weapons in the COVID-19 War: A Preliminary Study of Brazil, Canada, Denmark, UK and US | Not empirical primary research |
| Joelle 2020 | States of Emergency | Not empirical primary research |
| Jondison 2020 | Narrativas polÃticas, produÃ§Ã£o de vulnerabilidades e convulsÃ£o social no Brasil e no mundo, no contexto do Novo CoronavÃrus | Not focused in LAC countries |
| JosÃ©Borzacchiellod a 2020 | Pandemia do CoronavÃrus no Brasil: Impactos no TerritÃ³rio Cearense | Not empirical primary research |
| JosÃ©Emilio 2020 | Las nuevas necesidades ante la Covid-19: desafÃos y tendencias en la provincia argentina de CÃ³rdoba | Not empirical primary research |
| JoseAntonio 2020 | IMPACTOS INICIAIS DA COVID-19 NAS MICRO E PEQUENAS EMPRESAS DO ESTADO DO PARANÃ• - BRASIL | Not focused in LAC countries |
| Joseph 2020 | Maternal deaths in Brazil from severe COVID-19 respiratory disease: time for a global commitment to ending health disparities | Not empirical primary research |

| | | |
|---|---|---|
| Josimar 2020 | Analysis of internet trends related to medications for COVID-19in ten countries with the highest number of cases | Not focused in LAC countries |
| JosyLira 2020 | A campanha da influenza 2020 em meio a pandemia do coronavÃrus no estado do amazonas: um relato de experiÃªncia | Not empirical primary research |
| Jung 2020 | External validation of multimodal termination of resuscitation rules for out-of-hospital cardiac arrest patients in the COVID-19 era | Not focused in LAC countries |
| JustichZabala 2020 | Towards a new epidemiological worldview | Not empirical primary research |
| Kabad 2020 | The experience with volunteer and collaborative work in mental health and psychosocial care during the COVID-19 pandemic | Not focused in LAC countries |
| Kaplan 2020 | Voluntary collective isolation as a best response to COVID-19 for indigenous populations? A case study and protocol from the Bolivian Amazon. | Not empirical primary research |
| Karl 2020 | Dark matter, second waves and epidemiological modelling | Not focused in LAC countries |
| Kayral 2020 | Forecasting of COVID-19 infections in E7 countries and proposing some policies based on the Stringency Index | Not focused in LAC countries |
| Khafaie 2020 | Cross-country comparison of case fatality rates of Covid-19/SARS-CoV-2 | Not focused in LAC countries |
| Kluger 2020 | Is there an under-representation of skin of colour images during the COVID-19 outbreak? | Not empirical primary research |
| Koen 2020 | How FinTech Can Help Latin America to Deal with Economic Challenges and the COVID-19 Crisis | Not empirical primary research |
| Koterba 2020 | COVID-19 issues related to pediatric neuropsychology and inpatient rehabilitation - challenges to usual care and solutions during the pandemic | Not empirical primary research |
| Krecl 2020 | Drop in urban air pollution from COVID-19 pandemic: Policy implications for the megacity of Sao Paulo | Not empirical primary research |
| Kushnir 2020 | Plans to Reactivate Gastroenterology Practices Following the COVID-19 Pandemic: A Survey of North American Centers. | Not focused in LAC countries |
| Kyurkchiev 2020 | Another look at a good approximation of data for the distribution of COVID-19 in Cuba | Not empirical primary research |
| LÃ³pez 2020 | EnseÃ±anza virtual en educaciÃ³n fÃsica en primaria en MÃ©xico y la pandemia por COVID-19 | Not empirical primary research |
| LÃ³pez-Feldman 2020 | Environmental Impacts and Policy Responses to Covid-19: A View from Latin America | Not empirical primary research |
| LÃ³pezHernÃ¡ndez 2020 | Aerosol Box 2.0: Adjustments and Improvements Made in Mexico for Intubating Patients During the Coronavirus Disease 2019 Pandemic | Not empirical primary research |
| LailaCÃntiaMota 2020 | LEITURA GEOGRÃ• FICA NO CONTEXTO DA COVID-19 EM RONDÃ”NIA AO NORTE DO BRASIL | Not empirical primary research |

| | | |
|---|---|---|
| Lamelas 2020 | [Cardiovascular disease in times of COVID-19]. | Not empirical primary research |
| Lana 2020 | The novel coronavirus (SARS-CoV-2) emergency and the role of timely and effective national health surveillance. | Not empirical primary research |
| Lancuba 2020 | SARS-CoV-2 in Argentina | Not empirical primary research |
| Landa-Ramirez 2020 | Psychological support during COVID-19 death notifications: Clinical experiences from a hospital in Mexico. | Not empirical primary research |
| Larenas-Linnemann 2020 | [Logistic precautions in preparation and administration of allergen immunotherapy during the COVID-19 pandemic in Mexico] | Not empirical primary research |
| Larreal 2020 | Pandemic of the new coronavirus SARSCoV-2 in Venezuela | Not empirical primary research |
| Larson 2020 | Modeling SARS-CoV-2 Positivity Using Laboratory Data: Timing Is Everything | Not focused in LAC countries |
| Lasco 2020 | Medical populism and the COVID-19 pandemic | Not empirical primary research |
| Laureano 2020 | The different tests for the diagnosis of COVID-19 - A review in Brazil so far. | Not empirical primary research |
| LefioCeledÃ³n 2020 | CrÃ³nica de la epidemia por covid-19: perplejidad, revelaciÃ³n y crisis | Not empirical primary research |
| Leite 2020 | GestÃ£o da forÃ§a de trabalho em saÃºde e COVID-19: DesinformaÃ§Ã£o e ausÃªncia de polÃticas pÃºblicas no Brasil) | Not focused in LAC countries |
| Lemos 2020 | Human anatomy education and management of anatomic specimens during and after COVID-19 pandemic: Ethical, legal and biosafety aspects | Not empirical primary research |
| Leonel 2020 | COVID-19, head and neck cancer, and the need of training of health students and practitioners regarding to tobacco control and patient counseling. | Not empirical primary research |
| Lev 2020 | Untangling factors associated with country-specific COVID-19 incidence, mortality and case fatality rates during the first quarter of 2020 | Not focused in LAC countries |
| Lew 2020 | Normalidad post-pandemia: Â¿una nueva normalidad socio-ambiental o adiÃ³s a la normalidad? | Not empirical primary research |
| Lima 2020 | Crisis, conditioning factors, and challenges in the coordination of Brazil's federative State in the context of COVID-19 | Not empirical primary research |
| Lima 2020 | Determinants of preterm infantsâ€™ deaths at the Neonatal Intensive Care Units in the Northeast Countryside in Brazil | Not produced in response to the COVID-19 pandemic |
| Lima-Costa 2020 | Envelhecimento no Brasil e coronavirus: iniciativa ELSI-COVID-19, Aging in Brazil and the novel coronavirus: the ELSI-COVID-19 initiative | Not empirical primary research |
| Llibre-Guerra 2020 | The impact of COVID-19 on Mental Health in the Hispanic Caribbean Region | Not empirical primary research |

| | | |
|---|---|---|
| Loayza-Alarico 2020 | Rapid response teams for epidemiological surveillance: Mitigation of the coronavirus pandemic in peru, 2020 | Not empirical primary research |
| Lopez-Alvarez 2020 | Genome Sequence of SARS-CoV-2 Isolate Cali-01, from Colombia, Obtained Using Oxford Nanopore MinION Sequencing. | Not empirical primary research |
| Lopez-Ortiz 2020 | From the handling of an outbreak by an unknown pathogen in Wuhan to the preparedness and response in the face of the emergence of Covid-19 in Mexico. | Not empirical primary research |
| Lorenz 2020 | Have measures against COVID-19 helped to reduce dengue cases in Brazil? | Not empirical primary research |
| Lucas 2020 | ReconstruÃ§Ã£o MamÃ¡ria em Pacientes OncolÃ³gicos durante a Pandemia da Covid-19 | Not empirical primary research |
| Luis 2020 | COVID-19 Physician Deaths in Peru: A Result of An Underfunded and Fragmented Healthcare System | Not empirical primary research |
| LuisEnrique 2020 | La salud pÃºblica y el cuidado materno fetal y neonatal en la pandemia por el virus SARS-CoV-2 | Not empirical primary research |
| Macedo-Perez 2020 | Dermatological findings in COVID-19 patients: Mexican experience. | Not empirical primary research |
| Machado 2020 | Brazilian oral medicine and oral histopathology services: the worrying reality in the COVID-19 Era | Not empirical primary research |
| Machado 2020 | Nursing in COVID-19 times in brazil: a look at work management | Not empirical primary research |
| Maciel 2020 | Tuberculosis and coronavirus: what do we know? | Not empirical primary research |
| Magalhaes 2020 | Oil spill + COVID-19: A disastrous year for Brazilian seagrass conservation | Not empirical primary research |
| Magallanes-Garza 2020 | Rapid improvement of a critically ill obstetric patient with SARS-CoV-2 infection after administration of convalescent plasma | Not empirical primary research |
| Mahase 2020 | Covid-19: Deaths in Mexico triple since reopening began in June | Not empirical primary research |
| Malay 2020 | Climatic influence on the magnitude of COVID-19 outbreak: a stochastic model-based global analysis | Not focused in LAC countries |
| Malerbi 2020 | Diabetic retinopathy screening and the COVID-19 pandemic in Brazil | Not empirical primary research |
| Malta 2020 | Coronavirus in Brazil: The heavy weight of inequality and unsound leadership | Not empirical primary research |
| MarcanoGonzÃ¡lez 2020 | Las ciencias y la pandemia del Covid-19 | Not empirical primary research |
| Marcilio 2020 | Epidemiologic Surveillance in an academic hospital during the COVID-19 pandemic in Sao Paulo, Brazil: the key role of epidemiologic engagement in operational processes | Not empirical primary research |
| MarcoTulioPacheco 2020 | Exponential phase of covid19 expansion is not driven by climate at global scale | Not focused in LAC countries |

| | | |
|---|---|---|
| Marcus 2020 | Effective Teaching and Examination Strategies for Undergraduate Learning During COVID-19 School Restrictions | Not empirical primary research |
| Maria 2020 | SARS-CoV2 (COVID-19) y las figuras fiscales-legales | Not empirical primary research |
| MariaGeraldade 2020 | MORADIA POPULAR E PANDEMIA DO COVID-19: reflexÃµes sobre as dificuldades de isolamento social | Not empirical primary research |
| Mariani 2020 | Thoracic surgery in a hospital dedicated to treating COVID-19: challenges and solutions. | Not empirical primary research |
| Marinelli 2020 | Safe Handling of Containers of Expressed Human Milk in all Settings During the SARS-CoV-2 (COVID-19) Pandemic. | Not empirical primary research |
| Marinho 2020 | A proposta de adoÃ§Ã£o de fila Ãºnica nas unidades de terapia intensiva e nas demais internaÃ§Ãµes hospitalares, durante a pandemia de Covid-19 no Brasil: consideraÃ§Ãµes teÃ³ricas do campo da economia da saÃºde sobre as alternativas disponÃveis | Not empirical primary research |
| Marois 2020 | Assessing the potential impact of COVID-19 on life expectancy | Not focused in LAC countries |
| Marques 2020 | Covid-19: nursing care for safety in the mobile pre-hospital service | Not empirical primary research |
| Marques 2020 | Violence against women, children, and adolescents during the COVID-19 pandemic: overview, contributing factors, and mitigating measures. | Not empirical primary research |
| MartÃ 2020 | Incidental findings of COVID-19 in F18-FDG PET/CT from asymptomatic patients with cancer in two healthcare institutions in BogotÃ¡, Colombia | Not empirical primary research |
| MartelliJunior 2020 | Brazilian oral medicine and oral pathology: We are here during the COVID-19 pandemic. | Not empirical primary research |
| Martinez-Portilla 2020 | A Spanish-translated clinical algorithm for management of suspected SARS-CoV-2 infection in pregnant women | Not empirical primary research |
| Martinez-Rivera 2020 | Overmedication in COVID-19 Context: A Report From Peru | Not empirical primary research |
| MartinezRodriguez 2020 | Comparison of expert recommendations on clinical nutrition for hospitalized patients with COVID-19 | Not focused in LAC countries |
| Martins 2020 | As implicaÃ§Ãµes da COVID-19 no sistema cardiovascular: prognÃ³stico e intercorrÃªncias | Not empirical primary research |
| Martins-Filho 2020 | No evidence supports the use of ether and chloroform inhalation for treating COVID-19 | Not empirical primary research |
| Mascarenhas 2020 | Simultaneous occurrence of COVID-19 and dengue: what do the data show? | Not empirical primary research |
| Matos 2020 | New Coronavirus (SARS-CoV-2): advances to flatten the curve the prison population | Not empirical primary research |
| Medina 2020 | Primary healthcare in times of COVID-19: what to do? | Not empirical primary research |

| | | |
|---|---|---|
| Medina-Mora 2020 | COVID-19 and alcohol in Mexico: A serious health crisis, strong actions on alcohol in response-Commentary on Stockwell et al | Not empirical primary research |
| Meleti 2020 | Covid-19 diffusion and its impact on dental practice in distant countries with similar ethnic background | Not empirical primary research |
| Melo 2020 | COVID-19 and hypertensive disease in Brazil: possibility of a perfect storm | Not empirical primary research |
| Melo 2020 | FRIENDLY PROTOCOL FOR OOCYTE DONORS: FERTILITY OPTIMIZATION IN TIMES OF SOCIAL ISOLATION DUE TO COVID-19? | Not produced in response to the COVID-19 pandemic |
| Mendes 2020 | The challenges of monitoring neurological manifestations associated with COVID-19 in Latin America: does the World Health Organization need changes? | Not empirical primary research |
| Mendez-Probst 2020 | CLINICAL AND EPIDEMIOLOGICAL CHARACTERISTICS OF PATIENTS DIAGNOSED WITH COVID-19 IN A TERTIARY CARE CENTER IN MEXICO CITY: A PROSPECTIVE COHORT STUDY | Not empirical primary research |
| Mendoza-Saldana 2020 | Sociodemographic aspects of the older population in front of COVID-19 pandemic in a developing country: the case of Peru | Not empirical primary research |
| Mendoza-Ticona 2020 | Clinical classification and early therapy for COVID-19. Cases reported from Hospital de Emergencias Villa El Salvador, Lima-Peru | Not empirical primary research |
| Menezes 2020 | Universal COVID-19 testing in the obstetric population: impacts on public health | Not empirical primary research |
| Meriem 2020 | Model of a Testing-and-Quarantine Strategy to Slow-Down the COVID-19 Outbreak in Guadeloupe | Not focused in LAC countries |
| Meriem 2020 | Monitoring the post-containment COVID-19 crisis in Guadeloupe: Early-warning signals of destabilisation through bootstrapped probability density functions | Not focused in LAC countries |
| Meriem 2020 | Prediction of the time evolution of the COVID-19 disease in Guadeloupe with a stochastic evolutionary model | Not focused in LAC countries |
| Meurer 2020 | Launching a public statewide tele(oral)medicine service in Brazil during COVID-19 pandemic | Not empirical primary research |
| Michail 2020 | Climatic influences on the worldwide spread of SARS-CoV-2 | Not focused in LAC countries |
| Millan-Onate 2020 | Successful recovery of COVID-19 pneumonia in a patient from Colombia after receiving chloroquine and clarithromycin. | Not empirical primary research |
| Miller 2020 | COVID-19 in Latin America: Novel transmission dynamics for a global pandemic?. | Not empirical primary research |
| MinasGerais 2020 | Plano de ContingÃªncia Operativo para infecÃ§Ã£o humana pelo SARS COV-2 (doenÃ§a pelo CoronavÃrus - COVID-19) da MacrorregiÃ£o Jequitinhonha | Not empirical primary research |

| | | |
|---|---|---|
| MinasGerais 2020 | Plano de ContingÃªncia Operativo para infecÃ§Ã£o humana pelo SARS COV-2 (doenÃ§a pelo CoronavÃrus - COVID-19) da MacrorregiÃ£o Nordeste | Not empirical primary research |
| MinasGerais 2020 | Plano de ContingÃªncia Operativo para infecÃ§Ã£o humana pelo SARS COV-2 (doenÃ§a pelo CoronavÃrus - COVID-19) da MacrorregiÃ£o Noroeste | Not empirical primary research |
| MinasGerais 2020 | Plano de ContingÃªncia Operativo para infecÃ§Ã£o humana pelo SARS COV-2 (doenÃ§a pelo CoronavÃrus - COVID-19) da MacrorregiÃ£o Norte | Not empirical primary research |
| MinasGerais 2020 | Plano de ContingÃªncia Operativo para infecÃ§Ã£o humana pelo SARS COV-2 (doenÃ§a pelo CoronavÃrus - COVID-19) da MacrorregiÃ£o Oeste | Not empirical primary research |
| MinasGerais 2020 | Plano de ContingÃªncia Operativo para infecÃ§Ã£o humana pelo SARS COV-2 (doenÃ§a pelo CoronavÃrus - COVID-19) da MacrorregiÃ£o Sudeste | Not empirical primary research |
| MinasGerais 2020 | Plano de ContingÃªncia Operativo para infecÃ§Ã£o humana pelo SARS COV-2 (doenÃ§a pelo CoronavÃrus - COVID-19) da MacrorregiÃ£o Sul | Not empirical primary research |
| MinasGerais 2020 | Plano de ContingÃªncia Operativo para infecÃ§Ã£o humana pelo SARS COV-2 (doenÃ§a pelo CoronavÃrus - COVID-19) da MacrorregiÃ£o TriÃ¢ngulo do Norte | Not empirical primary research |
| MinasGerais 2020 | Plano de ContingÃªncia Operativo para infecÃ§Ã£o humana pelo SARS COV-2 (doenÃ§a pelo CoronavÃrus - COVID-19) da MacrorregiÃ£o TriÃ¢ngulo do Sul | Not empirical primary research |
| MinasGerais 2020 | Plano de ContingÃªncia Operativo para infecÃ§Ã£o humana pelo SARS COV-2 (doenÃ§a pelo CoronavÃrus - COVID-19) da MacrorregiÃ£o Vale do AÃ§o | Not empirical primary research |
| Minayo 2020 | The pandemic exacerbates health inequalities | Not empirical primary research |
| Ministeriode 2020 | BoletÃn EpidemiolÃ³gico Semana 04 (del 19 al 25 de enero del 2020) | Not empirical primary research |
| Ministeriode 2020 | BoletÃn EpidemiolÃ³gico Semana 10 (del 01 al 07 de marzo del 2020) | Not empirical primary research |
| MinisteriodelPoderPopularparala 2020 | COVID-19: Recomendaciones para el cuidado integral de mujeres embarazadas y reciÃ©n nacidos en la RepÃºblica Bolivariana de Venezuela Dirigidas a proveedores de salud | Not empirical primary research |
| MinisteriodelPoderPopularparala 2020 | Normativa sanitaria de responsabilidad social ante la pandemia denominada Coronavirus (Covid-19) | Not empirical primary research |
| MinisteriodelPoderPopularparalaSalud 2020 | GuÃa para el manejo y tratamiento de contactos y pacientes con COVID-19 | Not empirical primary research |

| | | |
|---|---|---|
| Mira 2020 | Development of the platform "Be+ against COVID", a website and a mobile app to prevent and address the stress reactions of healthcare workers caring COVID-19 patients | Not focused in LAC countries |
| Miranda 2020 | Challenges in the Practice of Sexual Medicine in the Time of COVID-19 in Brazil | Not empirical primary research |
| MoacirJosÃ©da 2020 | Decision Trees: Predictions of Global Vulnerability to Coronavirus Outbreaks | Not focused in LAC countries |
| Molina 2020 | COVID-19 in Ecuador, how the pandemic strained the surgical healthcare systems over the edge | Not empirical primary research |
| Molina-Betancur 2020 | Coronavirus disease 2019 and slums in the Global South: lessons from Medellin (Colombia) | Not empirical primary research |
| MolinaPrendes 2020 | Social impact of COVID-19 on Brazil and Ecuador: where reality exceeds statistics | Not empirical primary research |
| Mondeja 2020 | SARS-CoV-2: High-Resolution Microscopy Study in Human Nasopharyngeal Samples | Not focused in LAC countries |
| Monteleone 2020 | A review of initial data on pregnancy during the COVID-19 outbreak: implications for assisted reproductive treatments. | Not empirical primary research |
| Montenegro-LÃ³pez 2020 | [Use of point-of-care technologies for the management of the COVID-19 pandemic in Colombia] | Not empirical primary research |
| Monteverde 2019 | Oxigenoterapia con cÃ¡nula nasal de alto flujo en lactantes con infecciÃ³n respiratoria baja aguda: Experiencia en hospitales de la Ciudad de Buenos Aires | Not produced in response to the COVID-19 pandemic |
| Montoya 2020 | COVID-19 in pediatric cancer patients in a resource-limited setting: National data from Peru | Not empirical primary research |
| MoorePerea 2020 | CirugÃa electiva en organizaciones no COVID durante la pandemia | Not empirical primary research |
| Moraes 2020 | Contributions to address violence against older adults during the Covid-19 pandemic in Brazil | Not empirical primary research |
| Moraes 2020 | COVID-19 nas instituiÃ§Ãµes de longa permanÃªncia para idosos: estratÃ©gias de rastreamento laboratorial e prevenÃ§Ã£o da propagaÃ§Ã£o da doenÃ§a | Not empirical primary research |
| Morales 2020 | Economic Packages for COVID-19 Recovery Must Invest in More Resilient Health Systems | Not empirical primary research |
| Moreira 2020 | Chest radiography and computed tomography findings from a Brazilian patient with COVID-19 pneumonia. | Not empirical primary research |
| MorejÃ³nGiraldoni 2020 | Surveillance and control actions before notification of confirmed cases to COVID-19. Aguada de Pasajeros, Cienfuegos. Cuba | Not empirical primary research |
| MorenoSoto 2020 | The feasibility of generalized face mask usage during the COVID-19 pandemic: a perspective from Latin America. | Not empirical primary research |
| Mosteiro-Diaz 2020 | Presenteeism in nurses: comparative study of Spanish, Portuguese and Brazilian nurses | Not produced in response to the COVID-19 pandemic |

| | | |
|---|---|---|
| Mota 2020 | Epidemiological surveillance and the Covid-19 pandemic in Brazil. Elements to understand the Brazilian response and the explosion of cases and deaths | Not empirical primary research |
| Moura 2020 | Epidemiological information on Covid-19: influence of cyberculture on popular engagement to control measures | Not empirical primary research |
| Moura 2020 | InformaÃ§Ãµes epidemiolÃ³gicas sobre a Covid-19: influÃªncia da cibercultura no engajamento popular Ã s medidas de controle | Not empirical primary research |
| Mujica 2020 | Scientific production on COVID-19 in Cuban medical journals during the first 90 days after the start of the pandemic | Not focused in LAC countries |
| Muniz 2020 | COVID-19 - Computed tomography findings in two patients in Petropolis, Rio de Janeiro, Brazil. | Not empirical primary research |
| Munoz 2020 | Covid-19 in latin america: A first glance to the mortality | Not empirical primary research |
| Nadia 2020 | COVID-19 pediatric mortality rates are heterogenous between countries | Not focused in LAC countries |
| Nakamura 2020 | Airport risk of importation and exportation of the COVID-19 pandemic | Not focused in LAC countries |
| Namasivayam-MacDonald 2020 | Speech-Language Pathology Management for Adults With COVID-19 in the Acute Hospital Setting: Initial Recommendations to Guide Clinical Practice | Not empirical primary research |
| Namendys-Silva 2020 | Health care workers on the frontlines of COVID-19 in Mexico | Not empirical primary research |
| NÃºÃ±ez-LÃ³pez 2021 | Migration rate estimation in an epidemic network | Not produced in response to the COVID-19 pandemic |
| Nascimento 2020 | Covid-19 and hypercoagulable state: A new therapeutic perspective | Not empirical primary research |
| Nascimento 2020 | Crise e oportunidades para a preservaÃ§Ã£o digital da informaÃ§Ã£o em saÃºde | Not empirical primary research |
| Nascimento 2020 | Neurological complications associated with SARS-CoV-2 (COVID-19) in Brazil: Organization of the NEUROCOVID-RIO group and preliminary findings (Opinion Article) | Not empirical primary research |
| Navarrete-Reyes 2020 | Staying in a Burning House: Perks and Perils of a Hotline in the Times of COVID-19. | Not empirical primary research |
| Navia 2020 | Adapting Plastic Surgery Residency Training During COVID-19: The Experience of a Chilean University Hospital | Not empirical primary research |
| Nct 2020 | Protection of Health Workers Against COVID-19 | Not focused in LAC countries |
| NCT04335279 2020 | Evaluation of the Scleroderma Patient-centered Intervention Network COVID-19 Home-isolation Activities Together Program | Not focused in LAC countries |
| NCT04365309 2020 | Protective Effect of Aspirin on COVID-19 Patients | Not focused in LAC countries |
| NCT04401475 2020 | A Randomized, Double-Blind, Placebo-Controlled Study to Evaluate the Safety and | Not focused in LAC countries |

| | | |
|---|---|---|
| | Efficacy of EB05 + SOC vs. Placebo + SOC in Adult Hospitalized Patients With Moderate to Severe COVID-19 Pneumonia | |
| Nepomuceno 2020 | Besides population age structure, health and other demographic factors can contribute to understanding the COVID-19 burden | Not empirical primary research |
| Neto 2020 | Fake news no cenÃ¡rio da pandemia de Covid-19 | Not empirical primary research |
| Neto 2020 | Inflammaging: Analysis of a Risk Profile for Gerontocide by COVID-19 in Brazil | Not produced in response to the COVID-19 pandemic |
| Neyra-Leon 2020 | The impact of COVID-19 in the healthcare workforce in Peru | Not empirical primary research |
| Ng 2020 | Challenges Implementing Preventive Measures at a Nursing-Skilled Facility in Mexico | Not empirical primary research |
| Ng 2020 | Solutions to prevent and address physician burnout during the pandemic in Mexico | Not empirical primary research |
| Nick 2020 | Reconstructing the global dynamics of under-ascertained COVID-19 cases and infections | Not focused in LAC countries |
| Novaes 2020 | Desafios na conduÃ§Ã£o de pesquisas sobre a COVID-19 envolvendo seres humanos no Brasil | Not empirical primary research |
| Nunciaroni 2020 | New Coronavirus: (Re)thinking the care process in Primary Health and Nursing | Not empirical primary research |
| Nunez-Velasco 2020 | Response to SARS-CoV-2 Pandemic in a Non-COVID-19 Designated Latin-American Neurosurgery Department. | Not empirical primary research |
| O'Driscoll 2020 | Age-specific mortality and immunity patterns of SARS-CoV-2 | Not focused in LAC countries |
| Oliveira 2020 | Food (in)security in Brazil in the context of the SARS-CoV-2 pandemic. | Not empirical primary research |
| Oliveira 2020 | Personal Protective Equipment in the coronavirus pandemic: training with Rapid Cycle Deliberate Practice | Not empirical primary research |
| Oliveira 2020 | What has the covid-19 pandemic taught us about adopting preventive measures? | Not empirical primary research |
| Olmos 2020 | Nuevo coronavirus (COVID-19) en poblaciÃ³n general y pediÃ¡trica: una revisiÃ³n epidemiolÃ³gica: Chile 2020 | Not empirical primary research |
| Olsen 2020 | Decreased Influenza Activity During the COVID-19 Pandemic - United States, Australia, Chile, and South Africa, 2020 | Not focused in LAC countries |
| OrellanaCenteno 2020 | El coronavirus 19 (COVID 19) en MÃ©xico | Not empirical primary research |
| Oreskovic 2020 | Violations of independence: ethnicity and COVID-19 in Brazil | Not empirical primary research |
| OrganizaciÃ³nPanamericanadela 2020 | AlocuciÃ³n de la Directora en el seminario virtual "Liderazgo en la crisis: cÃ³mo reducir el impacto de la COVID-19 en AmÃ©rica Latina y Ã• frica" | Not empirical primary research |
| OrganizaciÃ³nPanamericanadela 2020 | Colombia: con un trabajo articulado y coordinado Colombia busca no dejar a nadie atrÃ¡s en la lucha contra la COVID-19 | Not empirical primary research |

| | | |
|---|---|---|
| OrganizaciÃ³nPanamericanadela 2020 | COVID-19 - Informe de situaciÃ³n de PanamÃ¡ NÂ°25: Agosto 4, 2020 | Not empirical primary research |
| OrganizaciÃ³nPanamericanadela 2020 | COVID-19 - Informe de situaciÃ³n de PanamÃ¡ NÂ°26: Agosto 11, 2020 | Not empirical primary research |
| OrganizaciÃ³nPanamericanadela 2020 | COVID-19 - Informe de situaciÃ³n de PanamÃ¡ NÂ°27: Agosto 17, 2020 | Not empirical primary research |
| OrganizaciÃ³nPanamericanadela 2020 | COVID-19 - Informe de situaciÃ³n de PanamÃ¡ NÂ°28: Agosto 23, 2020 | Not empirical primary research |
| OrganizaciÃ³nPanamericanadela 2020 | COVID-19 - Informe de situaciÃ³n de PanamÃ¡ NÂ°29: Agosto 31, 2020 | Not empirical primary research |
| OrganizaciÃ³nPanamericanadela 2020 | COVID-19 - Informe de situaciÃ³n de PanamÃ¡ NÂ°30: Septiembre 6, 2020 | Not empirical primary research |
| OrganizaciÃ³nPanamericanadela 2020 | COVID-19 - Informe de situaciÃ³n de PanamÃ¡ NÂ°31: Septiembre 13, 2020 | Not empirical primary research |
| OrganizaciÃ³nPanamericanadela 2020 | COVID-19 - Informe de situaciÃ³n de PanamÃ¡ NÂ°32: Septiembre 20, 2020 | Not empirical primary research |
| OrganizaciÃ³nPanamericanadela 2020 | COVID-19 - Informe de situaciÃ³n de PanamÃ¡ NÂ°33: Septiembre 28, 2020 | Not empirical primary research |
| OrganizaciÃ³nPanamericanadela 2020 | COVID-19 - Informe de situaciÃ³n de PanamÃ¡ NÂ°34: Octubre 5, 2020 | Not empirical primary research |
| OrganizaciÃ³nPanamericanadela 2020 | COVID-19 - Informe de situaciÃ³n de PanamÃ¡ NÂ°35: Octubre 11, 2020 | Not empirical primary research |
| OrganizaciÃ³nPanamericanadela 2020 | COVID-19 - Informe de situaciÃ³n de PanamÃ¡ NÂ°36: Octubre 18, 2020 | Not empirical primary research |
| OrganizaciÃ³nPanamericanadela 2020 | COVID-19 - Informe de situaciÃ³n de PanamÃ¡ NÂ°38: Noviembre 01, 2020 | Not empirical primary research |
| OrganizaciÃ³nPanamericanadela 2020 | COVID-19 - Reporte de situaciÃ³n 27: Paraguay, 25 de septiembre de 2020 | Not empirical primary research |
| OrganizaciÃ³nPanamericanadela 2020 | COVID-19 reporte de situaciÃ³n 20: Paraguay, 07 de agosto de 2020 | Not empirical primary research |
| OrganizaciÃ³nPanamericanadela 2020 | COVID-19 reporte de situaciÃ³n 21: Paraguay, 14 de agosto de 2020 | Not empirical primary research |
| OrganizaciÃ³nPanamericanadela 2020 | COVID-19 reporte de situaciÃ³n 22: Paraguay, 21 de agosto de 2020 | Not empirical primary research |
| OrganizaciÃ³nPanamericanadela 2020 | COVID-19 reporte de situaciÃ³n 23: Paraguay, 04 de septiembre de 2020 | Not empirical primary research |
| OrganizaciÃ³nPanamericanadela 2020 | COVID-19 reporte de situaciÃ³n 23: Paraguay, 28 de agosto de 2020 | Not empirical primary research |
| OrganizaciÃ³nPanamericanadela 2020 | COVID-19 reporte de situaciÃ³n 24: Paraguay, 04 de septiembre de 2020 | Not empirical primary research |
| OrganizaciÃ³nPanamericanadela 2020 | COVID-19 reporte de situaciÃ³n 25: Paraguay, 11 de septiembre de 2020 | Not empirical primary research |
| OrganizaciÃ³nPanamericanadela 2020 | COVID-19 reporte de situaciÃ³n 26: Paraguay, 18 de septiembre de 2020 | Not empirical primary research |
| OrganizaciÃ³nPanamericanadela 2020 | COVID-19 reporte de situaciÃ³n 28: Paraguay, 02 de octubre de 2020 | Not empirical primary research |
| OrganizaciÃ³nPanamericanadela 2020 | Monitoreo de la respuesta de paÃses Sudamericanos frente a la pandemia de COVID-19 | Not empirical primary research |
| OrganizaciÃ³nPanamericanadela 2020 | Reporte de situacion COVID-19 Colombia No. 131: 30 de agosto de 2020 | Not empirical primary research |

| | | |
|---|---|---|
| OrganizaciÃ³nPanamericanadela 2020 | Reporte de Situacion COVID-19 Colombia No. 159: 14 de octubre 2020 | Not empirical primary research |
| OrganizaciÃ³nPanamericanadela 2020 | Reporte de situacion COVID-19 Colombia No. 160: 15 de octubre 2020 | Not empirical primary research |
| OrganizaciÃ³nPanamericanadela 2020 | Reporte de Situacion COVID-19 Colombia No. 161: 18 de octubre 2020 | Not empirical primary research |
| OrganizaciÃ³nPanamericanadela 2020 | Reporte de Situacion COVID-19 Colombia No. 162: 19 de octubre 2020 | Not empirical primary research |
| OrganizaciÃ³nPanamericanadela 2020 | Respuesta de la OrganizaciÃ³n Panamericana de la Salud a la COVID-19 en las AmÃ©ricas | Not empirical primary research |
| OrganizaciÃ³nPanamericanadela 2020 | Salud y EconomÃa. Una convergencia necesaria para enfrentar el COVID-19 y retomar la senda hacia el desarrollo sostenible en AmÃ©rica Latina y el Caribe | Not empirical primary research |
| OrganizaciÃ³nPanamericanadela 2020 | SistematizaciÃ³n de la respuesta de la RepresentaciÃ³n en la RepÃºblica Dominicana a la emergencia por la COVID-19 en el perÃodo de enero a septiembre de 2020 | Not empirical primary research |
| Ornell 2020 | The impact of the COVID-19 pandemic on the mental health of healthcare professionals. | Not empirical primary research |
| Orsini 2020 | Danos psÃquicos durante pandemia por COVID-19 no Brasil | Not empirical primary research |
| Ortega 2020 | New needs because of Covid-19: challenges and trends in the Argentinian province of CÃ³rdoba | Not empirical primary research |
| Ortiz-Brizuela 2020 | Erratum: CLINICAL AND EPIDEMIOLOGICAL CHARACTERISTICS OF PATIENTS DIAGNOSED WITH COVID-19 IN A TERTIARY CARE CENTER IN MEXICO CITY: A PROSPECTIVE COHORT STUDY (Revista de investigacion clinica; organo del Hospital de Enfermedades de la Nutricion (2020) 72 3 (165-177)) | Not empirical primary research |
| Otoya-Tono 2020 | COVID-19: generalidades, comportamiento epidemiolÃ³gico y medidas adoptadas en medio de la pandemia en Colombia | Not empirical primary research |
| Otoya-Tono 2020 | COVID-19: Generalities, epidemiological behavior and measures adopted in the middle of the pandemic in Colombia | Not empirical primary research |
| PÃ©rez 2020 | La pandemia de COVID-19 y sus violencias en AmÃ©rica Latina | Not empirical primary research |
| PÃ©rezdelCaz 2020 | Organization of burn units during the COVID-19 pandemic: experience of 5 burn units | Not empirical primary research |
| Pablos-MÃ©ndez 2020 | Covid-19 in Latin America | Not empirical primary research |
| Palamim 2020 | COVID-19 in the Indigenous Population of Brazil | Not empirical primary research |
| PanAmericanHealth 2020 | Retrofit Measures for COVID-19, 10 June 2020 | Not empirical primary research |
| ParÃs-Pineda 2020 | PREVENCIÃ“N DE INFECCIONES RESPIRATORIAS EN INSTITUCIONES DE ATENCIÃ“N INTEGRAL EN EL NORTE DE SANTANDER | Not produced in response to the COVID-19 pandemic |

| | | |
|---|---|---|
| Pareja-Ramos 2020 | Importance of the implementation of the chest tomography to contribute to the early diagnosis and timely triage of patients with COVID-19 in Peruvian hospitals | Not empirical primary research |
| PariciodelCastillo 2020 | Child mental health and covid-19 pandemic: questions and challenges | Not empirical primary research |
| Passos 2020 | The Lancet: COVID-19 in Brazil: "So what"? | Not empirical primary research |
| Paula 2020 | Hand hygiene in high-complexity sectors as an integrating element in the combat of Sars-CoV-2 | Not produced in response to the COVID-19 pandemic |
| PaulaKwamme 2020 | ComunicaÃ§Ã£o, educaÃ§Ã£o e vigilÃ¢ncia popular em saÃºde em tempos de COVID-19 â€“ a experiÃªncia das comunidades de NiterÃ³i, RJ | Not empirical primary research |
| Pawlak 2020 | Impact of COVID-19 on endoscopy trainees: an international survey | Not focused in LAC countries |
| Paz-Ibarra 2020 | Management of diabetes mellitus in the era of COVID-19 | Not empirical primary research |
| Pearson 2020 | Nicaragua's surprising response to COVID-19 | Not empirical primary research |
| Pecho-Silva 2020 | Possibilities of "crushing" the transmission curve of COVID-19 in Latin America - We still have time | Not empirical primary research |
| PedrazaAlejo 2020 | Surveillance and control actions in a COVID-19 case at the JosÃ© Luis Chaviano Polyclinic in Cienfuegos, Cuba | Not empirical primary research |
| Penna 2020 | PNAD COVID-19: A powerful new tool for Public Health Surveillance in Brazil | Not empirical primary research |
| Penteado 2021 | Covid-19 effects on municipal solid waste management: What can effectively be done in the Brazilian scenario? | Not empirical primary research |
| PerazadeAparicio 2020 | Occupational health facing COVID-19 pandemic in Ecuador | Not empirical primary research |
| Pereira 2020 | The risk of COVID-19 transmission in favelas and slums in Brazil. | Not empirical primary research |
| Peres 2020 | COVID-19-related challenges in dental education: experiences from Australia, Brazil, and the USA | Not focused in LAC countries |
| Peres 2020 | Covid-19-related challenges in dental education: Experiences from Brazil, the USA, and Australia | Not empirical primary research |
| Perez 2020 | On Race and the Environment in the COVID-19 Pandemic | Not empirical primary research |
| Perez-Brumer 2020 | COVID-19 Policies can Perpetuate Violence Against Transgender Communities: Insights from Peru. | Not empirical primary research |
| Perez-Pedrogo 2020 | COVID-19 in Puerto Rico: Preliminary observations on social distancing and societal response toward a novel health stressor | Not empirical primary research |
| PerlaGarcia 2020 | Increase in Relative Humidity Increases the COVID-19 Contagion Rate and the Increase in UV Index Decreases It | Not focused in LAC countries |

| | | |
|---|---|---|
| Perondi 2020 | Setting up hospital care provision to patients with COVID-19: lessons learnt at a 2400-bed academic tertiary center in Sao Paulo, Brazil | Not empirical primary research |
| Peru 2020 | Uso de la telemedicina en covid: seguimiento y manejo de casos positivos | Not empirical primary research |
| Peru 2020 | Uso de tocilizumab para el tratamiento de pacientes con COVID-19 | Not empirical primary research |
| Peru 2020 | Uso de ventilacion multiple con un solo ventilador | Not empirical primary research |
| PessaValente 2020 | COVID-19 among health workers in Brazil: The silent wave. | Not empirical primary research |
| PeterOswaldoOjeda 2020 | Nueva alternativa para el tratamiento para Covid 19 en Ecuador | Not empirical primary research |
| PiauÃ 2020 | NOTA TÃ‰CNICA - NOVO CORONAVÃ• RUS | Not empirical primary research |
| Pilar 2020 | An effective COVID-19 response in South America: the Uruguayan Conundrum | Not empirical primary research |
| Pina-Reyna 2020 | Atencion de los sindromes coronarios agudos durante la contingencia sanitaria por brote de SARS-CoV-2, Care of acute coronary syndromes during the health contingency due to a SARS-CoV-2 outbreak | Not empirical primary research |
| Pinedo-Soria 2020 | Virtual medical education in Peru during COVID-19 | Not empirical primary research |
| Pinheiro 2020 | [Current situation of COVID-19 in South America] | Not empirical primary research |
| Pinheiro 2020 | Surgical cancer care in the COVID-19 era: front line views and consensus | Not empirical primary research |
| Pizan-Campos 2020 | Impact of covid-19 on the education of medical students in Peru | Not empirical primary research |
| Pizzichini 2020 | Measures of frequency: calculating prevalence and incidence in the era of COVID-19. | Not empirical primary research |
| Polidoro 2020 | Territories Under Siege: Risks of the Decimation of Indigenous and Quilombolas Peoples in the Context of COVID-19 in South Brazil | Not empirical primary research |
| Ponce-Torres 2020 | Carta a la editora - ParticipaciÃ³n de los estudiantes de medicina de LatinoamÃ©rica en la pandemia por COVID-19 | Not empirical primary research |
| Portada 2020 | The Final Frontier: China, Taiwan, and the United States in Strategic Competition for Central America | Not empirical primary research |
| Portella 2020 | Evidence map on the contributions of traditional, complementary and integrative medicines for health care in times of COVID-19 | Not empirical primary research |
| Poterico 2020 | Response to comment on "genetic variants and source of introduction of SARS-CoV-2 in South America" | Not empirical primary research |
| Prado 2020 | La complejidad del entorno en la gestiÃ³n de la seguridad ciudadana ante el Covid-19 | Not empirical primary research |
| Prado 2020 | The environment complexity in the management of citizen security by Covid-19 | Not empirical primary research |

| | | |
|---|---|---|
| Preiss 2020 | Challenges facing the COVID-19 pandemic in Brazil: lessons from short food supply systems. | Not empirical primary research |
| Proano 2020 | On the Macroeconomic and Social Impact of the Coronavirus Pandemic in Latin America and the Developing World. | Not empirical primary research |
| Queiroz 2020 | Management of inflammatory bowel disease patients in the COVID-19 pandemic era: a Brazilian tertiary referral center guidance. | Not empirical primary research |
| Quispe-Salcedo 2020 | El impacto del COVID-19 en la odontologÃa peruana | Not empirical primary research |
| Rabiller 2020 | Hospital Radiopharmacy in Argentina during the COVID-19 pandemic. Criteria and rationale for the organization of work | Not empirical primary research |
| Rafaell 2020 | Epidemiologia, polÃticas pÃºblicas e pandemia de Covid-19: o que esperar no Brasil? | Not empirical primary research |
| Raifman 2020 | Disparities in the Population at Risk of Severe Illness From COVID-19 by Race/Ethnicity and Income. | Not focused in LAC countries |
| Raiha 2020 | Simple discrete-time self-exciting models can describe complex dynamic processes: a case study of COVID-19 | Not focused in LAC countries |
| Raimundo 2020 | Vulnerability of micro-regions within Northeast Brazil to the new coronavirus (SARS-CoV-2) pandemic | Not empirical primary research |
| Raj 2020 | A machine learning aided global diagnostic and comparative tool to assess effect of quarantine control in Covid-19 spread | Not focused in LAC countries |
| RamÃrez-SÃ¡nchez 2020 | Los profesionales de la salud en los tiempos de la pandemia: personal de EnfermerÃa frente al COVID-19 | Not empirical primary research |
| Ramirez 2020 | Sars-cov-2 in the amazon region: A harbinger of doom for amerindians | Not empirical primary research |
| Rangel 2020 | â€œRodovÃrusâ€□ ou â€œCaronavÃrusâ€• ? Mapas da DistribuiÃ§Ã£o do Covid-19 na RegiÃ£o Sul do Brasil: IndÃcios da contaminaÃ§Ã£o por rodovias | Not empirical primary research |
| Raquel 2020 | Similitudes y diferencias entre las epidemias del pasado y la pandemia actual de covid-19 en Uruguay | Not empirical primary research |
| Rasha 2020 | Transformation ofÂ LifeÂ andÂ ResearchÂ withÂ COVID-19:Â ThroughÂ the EyesÂ ofÂ Three WomenÂ PhDÂ StudentsÂ from Brazil, Palestine and Sri Lanka | Not empirical primary research |
| Raskin 2020 | Genetics of COVID-19 | Not empirical primary research |
| RBR-7p6568 2019 | Oral hygiene in critically ill patients | Not produced in response to the COVID-19 pandemic |
| RedArgentinaPÃºblicadeEvaluaciÃ³ndeTecnologÃas 2020 | Uso de medidas de barrera especÃficas y bÃºsqueda activa para la prevenciÃ³n de la | Not empirical primary research |

| | | |
|---|---|---|
| | diseminaciÃ³n de SARS-COV2 en el Ã¡mbito quirÃºrgico | |
| Reis 2020 | O novo normal da Odontologia: revisÃ£o das recomendaÃ§Ãµes para retomada da assistÃªncia odontolÃ³gica durante a pandemia da COVID-19 | Not empirical primary research |
| Reis 2020 | Severe coronavirus infection in pregnancy: challenging cases report | Not empirical primary research |
| Reis 2020 | Uso dos Equipamentos de ProteÃ§Ã£o Individual no Atendimento OdontolÃ³gico Durante Surto da COVID-19 e Alternativas em PerÃodos de Desabastecimento: RevisÃ£o Integrativa | Not empirical primary research |
| RepÃºblica 2020 | BoletÃn especial No. 129: Enfermedad por Coronavirus 2019 (COVID-19) | Not empirical primary research |
| Resiere 2020 | Implementation of Medical and Scientific Cooperation in the Caribbean Using Blockchain Technology in Coronavirus (Covid-19) Pandemics | Not empirical primary research |
| Ribas 2020 | Coronavirus Disease 2019 (COVID-19) and healthcare-associated infections: Emerging and future challenges for public health in Brazil | Not empirical primary research |
| Ribas 2020 | Coronavirus Disease 2019 (COVID-19) and healthcare-associated infections: Emerging and future challenges for public health in Brazil. | Not empirical primary research |
| Ribeiro 2020 | Arboviral diseases and COVID-19 in Brazil: Concerns regarding climatic, sanitation, and endemic scenario. | Not empirical primary research |
| Ribeiro 2020 | Concerns about COVID-19 and tuberculosis in Brazil: Social and public health impacts | Not empirical primary research |
| Ribeiro 2020 | Covid-19 pandemic and the motivations for demanding health service in indigenous villages | Not empirical primary research |
| Ribeiro 2020 | Does Vitamin D play a role in the management of Covid-19 in Brazil?. | Not empirical primary research |
| Ribeiro 2020 | In the COVID-19 pandemic in Brazil, do brown lives matter? | Not empirical primary research |
| Ribeiro 2020 | IntervenÃ§Ãµes de restriÃ§Ã£o de mobilidade social durante a pandemia de COVID-19 e suas repercussÃµes psicossociais no Brasil | Not empirical primary research |
| Ribeiro 2020 | Interventions of mobility restrictions during COVID-19 pandemic and psychosocial repercussions in Brazil | Not empirical primary research |
| Ribeiro 2020 | The role of spirituality in the COVID-19 pandemic: a spiritual hotline project | Not empirical primary research |
| RibeiroConceiÃ§Ã£o 2020 | Covid-19 - An effective municipal coordination and articulation exercise: the NiterÃ³i experience | Not empirical primary research |
| RibeiroRibeiro 2020 | Besides the climate model, other variables driving the COVID-19 spread in Brazil. | Not empirical primary research |
| RibotReyes 2020 | Efectos de la COVID-19 en la salud mental de la poblaciÃ³n | Not empirical primary research |
| Richtmann 2020 | Fetal deaths in pregnancies with SARS-CoV-2 infection in Brazil: A case series | Not empirical primary research |
| Riechelmann 2020 | Evidence-based recommendations for gastrointestinal cancers during the COVID-19 | Not empirical primary research |

| | | |
|---|---|---|
| | pandemic by the Brazilian Gastrointestinal Tumours Group. | |
| Righetti 2020 | Physiotherapy Care of Patients with Coronavirus Disease 2019 (COVID-19) - A Brazilian Experience. | Not empirical primary research |
| RiodeJaneiro 2020 | Prevention measures and care for suspected cases of infection with the new coronavirus in maternity hospitals in the state of Rio de Janeiro | Not empirical primary research |
| RioGrandedoSul 2020 | Boas prÃ¡ticas para prevenÃ§Ã£o do CoronavÃ-rus (COVID-19) em supermercados | Not empirical primary research |
| RioGrandedoSul 2020 | COVID-19: Medidas de biosseguranÃ§a em estabelecimentos de saÃºde, funerÃ¡rios e congÃªneres e cuidados apÃ³s a morte | Not empirical primary research |
| RioGrandedoSul 2020 | OrientaÃ§Ãµes aos serviÃ§os da rede de atenÃ§Ã£o psicossocial sobre estratÃ©gias de prevenÃ§Ã£o de disseminaÃ§Ã£o do COVID-19 | Not empirical primary research |
| RioGrandedoSul 2020 | OrientaÃ§Ãµes para o cuidado em saÃºde Ã populaÃ§Ã£o em situaÃ§Ã£o de rua em razÃ£o da disseminaÃ§Ã£o do coronavÃrus | Not empirical primary research |
| RioGrandedoSul 2020 | OrientaÃ§Ãµes tÃ©cnicas para as maternidades em perÃodo da COVID-19 | Not empirical primary research |
| RioGrandedoSul 2020 | RecomendaÃ§Ãµes para contenÃ§Ã£o da infecÃ§Ã£o humana pelo novo CoronavÃrus (COVID-19) em povos indÃgenas no Estado do RS | Not empirical primary research |
| Riquelme 2020 | Recomendaciones de Asociaciones Chilenas de Endoscopia Digestiva (ACHED), de HepatologÃa (ACHHEP) y la AgrupaciÃ³n Chilena de Trabajo en Enfermedad de Crohn y Colitis Ulcerosa (ACTECCU) para el manejo de pacientes con patologÃas gastrointestinales en pandemia SARS-CoV-2 | Not empirical primary research |
| Rizo-Patron 2020 | Pediatric hospital demand in COVID-19 times in PerÃº | Not empirical primary research |
| Rizo-Patron 2020 | Pediatric hospital demand in the COVID-19 pandemic | Not empirical primary research |
| Rodrigues 2020 | GestÃ£o da pandemia coronavÃrus em um hospital: relato de experiÃªncia profissional | Not empirical primary research |
| Rodrigues 2020 | Pandemia de Covid-19 e a saÃºde dos refugiados no Brasil | Not empirical primary research |
| Rodriguez 2020 | Burnout in pediatric critical care medicine: more challenging days during the COVID-19 pandemic | Not empirical primary research |
| Rodriguez-Morales 2020 | Preparation and control of the coronavirus disease 2019 (COVID-19) in Latin America | Not empirical primary research |
| Rodriguez-Morales 2020 | Tracing New Clinical Manifestations in Patients with COVID-19 in Chile and Its Potential Relationship with the SARS-CoV-2 Divergence. | Not empirical primary research |
| Rodriguez-Santos 2020 | Argentine experience with telemedicine for venous care during the COVID-19 pandemic | Not empirical primary research |
| Rojulpote 2020 | COVID-19 and the Heart | Not focused in LAC countries |

| | | |
|---|---|---|
| Roman 2020 | Treatment of COVID-19 in peru and bolivia, and self-medication risks | Not focused in LAC countries |
| Romani 2020 | Adaptation of the editorial process of the peruvian journal of experimental medicine and public health during the COVID-19 pandemic | Not empirical primary research |
| Romani 2020 | Reusing N95 Respirators: Decontamination strategies applicable in COVID-19 pandemics in Peru | Not empirical primary research |
| Romero 2020 | GuÃa de buenas prÃ¡cticas en OdontologÃa para Uruguay durante la pandemia Covid-19 | Not empirical primary research |
| Romero 2020 | Limited genomic information in public databases to investigate SARS-COV-2 in Latin America | Not empirical primary research |
| RomeroHiller 2020 | La gestiÃ³n pÃºblica Venezolana en tiempos del Covid-19 | Not empirical primary research |
| Ronald 2020 | A SEIR-like model with a time-dependent contagion factor describes the dynamics of the Covid-19 pandemic | Not focused in LAC countries |
| Rosemary 2020 | The Impact of COVID-19 on Indigenous Peoples | Not focused in LAC countries |
| Rossato 2020 | Could the COVID-19 pandemic aggravate antimicrobial resistance? | Not empirical primary research |
| Rosselli 2020 | COVID-19 and potential years of life lost in Colombia | Not empirical primary research |
| RosselliMd 2020 | COVID-19 in Colombia: The first 90 days | Not empirical primary research |
| Roza 2020 | The Coronavirus Pandemic and the Impact on Donation and Transplantation in Brazil. | Not empirical primary research |
| Rubio 2020 | COVID-19 and Cancer in Cuba | Not empirical primary research |
| Russo 2020 | How the 'plates' of a health system can shift, change and adjust during economic recessions: A qualitative interview study of public and private health providers in Brazil's Sao Paulo and Maranhao states | Not produced in response to the COVID-19 pandemic |
| SÃ¡nchez 2020 | Aplicaciones de la CriomicroscopÃa ElectrÃ³nica en ProducciÃ³n de Vacunas contra el SARS-CoV-2 | Not empirical primary research |
| SÃ¡nchez 2020 | Pandemia de COVID-19 del 2020 | Not empirical primary research |
| SÃ¡nchez 2020 | Plasma de convaleciente para el tratamiento de la COVID-19: Protocolo para el acceso y uso en Venezuela | Not empirical primary research |
| SÃ¡nchez 2020 | Surgical education in Colombia in the era of COVIDÂ19 | Not empirical primary research |
| SÃ£oPaulo 2020 | Serological survey for SARS-COV-2: prevalence of infection among schoolchildren in the city of SÃ£o Paulo: results of phase 1 | Not empirical primary research |
| SÃ£oPauloSecretaria daSaÃºde 2020 | Epidemiological surveillance of pediatric multisystem inflammatory syndrome (SIM-P), temporarily associated with COVID-19 in the State of SÃ£o Paulo, epidemiological weeks 01 to 37, 2020 | Not empirical primary research |

| | | |
|---|---|---|
| SÃ£oPauloSecretaria daSaÃºde 2020 | Epidemiological surveillance of pediatric multisystem inflammatory syndrome (SIM-P), temporarily associated with COVID-19 in the State of SÃ£o Paulo, epidemiological weeks 01 to 41 of 2020 | Not empirical primary research |
| SÃ£oPauloSecretaria daSaÃºde 2020 | Epidemiological surveillance of pediatric multisystem inflammatory syndrome (SIM-P), temporarily associated with COVID-19 in the State of SÃ£o Paulo, epidemiological weeks 01 to 42, 2020 | Not empirical primary research |
| Sahai 2020 | ARIMA modelling & forecasting of COVID-19 in top five affected countries | Not empirical primary research |
| Saiz-Ã• lvarez 2020 | B Corps: A Socioeconomic Approach for the COVID-19 Post-crisis | Not produced in response to the COVID-19 pandemic |
| SalazarMather 2020 | Love in the time of COVID-19: negligence in the Nicaraguan response. | Not empirical primary research |
| SaldarriagaRivera 2020 | Recommendations on the management of adult patients with rheumatic diseases in the context of SARS-CoV-2/COVID-19 infection. Colombian Association of Rheumatology | Not empirical primary research |
| SaldarriagaRivera 2020 | Recommendations on the management of adult patients with rheumatic diseases in the context of SARS-CoV-2/COVID-19 infection. Colombian Association of Rheumatology | Not empirical primary research |
| Sales 2020 | COVID-19 health surveillance in Brazil: investigation of contacts by primary health care as a community protection strategy | Not empirical primary research |
| Sales 2020 | COVID-19 health surveillance in Brazil: seeking contacts in communities as a self-protection strategy | Not empirical primary research |
| SallesNeto 2020 | Forecast UTI: application for predicting intensive care unit beds in the context of the COVID-19 pandemic | Not empirical primary research |
| Sampaio 2020 | AvaliaÃ§Ã£o do Plano de AÃ§Ã£o Implementado pelo ServiÃ§o MÃ©dico de uma Unidade de ReferÃªncia em Cuidados Paliativos OncolÃ³gicos frente Ã Pandemia de Covid-19 | Not empirical primary research |
| Sanchez 2020 | COVID-19 in prisons: an impossible challenge for public health?. | Not empirical primary research |
| Sanchez 2020 | Mental health in the age of COVID-19, a Mexican experience | Not empirical primary research |
| SanchezDiaz 2020 | COVID-19 and Ethics: A Latin American Perspective | Not empirical primary research |
| Sanchez-Duque 2020 | [Coronavirus disease 2019 (COVID-19) in Latin America: Role of primary care in preparedness and response]. | Not empirical primary research |
| Sanchez-Duque 2020 | Are we now observing an increasing number of coinfections between SARS-CoV-2 and other respiratory pathogens? | Not empirical primary research |
| SantaCatarina 2020 | Boletim epidemiolÃ³gico diÃ¡rio COVID-19 - 28 de setembro de 2020 | Not empirical primary research |

| | | |
|---|---|---|
| SantaCatarina 2020 | Boletim epidemiolÃ³gico diÃ¡rio COVID-19 Â 29 de setembro de 2020 | Not empirical primary research |
| SantaCatarina 2020 | Boletim EpidemiolÃ³gico semanal nÂº 01 Covid-19 (SARS-COV-2) Â 21 de maio de 2020 | Not empirical primary research |
| SantaCatarina 2020 | Boletim EpidemiolÃ³gico semanal nÂº 02 Covid-19 (SARS-COV-2) Â 27 de maio de 2020 | Not empirical primary research |
| SantaCatarina 2020 | Boletim EpidemiolÃ³gico semanal nÂº 03 Covid-19 (SARS-COV-2) Â 01 de junho de 2020 | Not empirical primary research |
| SantaCatarina 2020 | Boletim EpidemiolÃ³gico semanal nÂº 04 Covid-19 (SARS-COV-2) Â 08 de junho de 2020 | Not empirical primary research |
| SantaCatarina 2020 | Boletim EpidemiolÃ³gico semanal nÂº 05 Covid-19 (SARS-COV-2) Â 16 de junho de 2020 | Not empirical primary research |
| SantaCatarina 2020 | Boletim EpidemiolÃ³gico semanal nÂº 06 Covid-19 (SARS-COV-2) Â 22 de junho de 2020 | Not empirical primary research |
| SantaCatarina 2020 | Como proteger sua saÃºde mental em tempos de CoronavÃrus | Not empirical primary research |
| SantaCatarina 2020 | CoronavÃrus COVID-19: RecomendaÃ§Ãµes ao atendimento odontolÃ³gico no SUS em Santa Catarina | Not empirical primary research |
| SantaCatarina 2020 | Fluxo de atendimento aos casos suspeitos, provÃ¡veis e confirmados do novo CoronavÃrus (COVID-19) | Not empirical primary research |
| SantaCatarina 2020 | Medidas de enfrentamento Ã violÃªncia domÃ©stica no contexto da pandemia de COVID-19 | Not empirical primary research |
| SantaCatarina 2020 | Medidas de enfrentamento ao COVID-19 no sistema prisional | Not empirical primary research |
| SantaCatarina 2020 | OrientaÃ§Ã£o para profissionais de saÃºde no uso de prÃ¡ticas integrativas e complementares no perÃodo de pandemia do COVID-19: versÃ£o 1 | Not empirical primary research |
| SantaCatarina 2020 | OrientaÃ§Ãµes sobre o atendimento da gestante, puÃ©rpera e recÃ©m nascido no enfrentamento Ã pandemia de COVID-19 | Not empirical primary research |
| SantaCatarina 2020 | RecomendaÃ§Ãµes Ã s populaÃ§Ãµes indÃ-genas para o enfrentamento Ã pandemia de COVID-19 | Not empirical primary research |
| SantaCatarina 2020 | RecomendaÃ§Ãµes para os centros de atenÃ§Ã£o psicossocial (CAPS) para prevenÃ§Ã£o e controle de infecÃ§Ãµes pelo novo CoronavÃrus | Not empirical primary research |
| SantaCatarina 2020 | RecomendaÃ§Ãµes sobre o trabalho dos agentes comunitÃ¡rios de saÃºde (acs) na atenÃ§Ã£o primÃ¡ria Ã saÃºde para o enfrentamento da pandemia de COVID-19 | Not empirical primary research |
| Santaella-Tenorio 2020 | Sars-cov-2 diagnostic testing alternatives for latin america | Not empirical primary research |
| Santos 2000 | A mortalidade por infecÃ§Ã¤o respiratÃ³ria agudas no Brasil | Not produced in response to the COVID-19 pandemic |
| Santos 2020 | COVID-19 interferes in the disclosure of the first Brazilian GIAHS site | Not empirical primary research |

| | | |
|---|---|---|
| Santos 2020 | Um "fato social total": COVID-19 e povos indigenas no Brasil, A "total social fact": COVID-19 and indigenous peoples in Brazil | Not empirical primary research |
| Schtruk 2020 | COVID-19 infection in heart transplantation: Case reports | Not empirical primary research |
| Schuelter-Trevisol 2020 | Partnership between the academy and public and private health systems to fight COVID-19: an experience report in Tubarao, Santa Catarina, Brazil | Not empirical primary research |
| Schuelter-Trevisol 2020 | Partnership between the academy and public and private health systems to fight the COVID-19: an experience report in TubarÃ£o, Santa Catarina, Brazil | Not empirical primary research |
| Schulze-Koops 2020 | Actions required to assure continuous supply of anti-rheumatic medication during the SARS-CoV-2/COVID-19 pandemic. Response to: 'Treatment adherence behaviours in rheumatic diseases during pandemic COVID-19: a Latin American experience' by Pineda-Sic et a | Not empirical primary research |
| Scorsolini-Comin 2020 | Mentoring program with nursing students in the context of the pandemic of COVID-19 in Brazil | Not empirical primary research |
| Scorsolini-Comin 2020 | Programa de tutorÃa con estudiantes de enfermerÃa en el contexto de la pandemia de COVID-19 en Brasil | Not empirical primary research |
| ScrippsWhittierDiabetes 2020 | Dulce Digital-COVID Aware (DD-CA) Discharge Texting Platform for US/Mexico Border Hispanics With Diabetes + COVID-19 | Not focused in LAC countries |
| SecretarÃadeSalud 2020 | GuÃa para el funcionamiento de los equipos de respuesta rÃ¡pida en el contexto de COVID-19 Honduras | Not empirical primary research |
| SecretarÃadeSalud 2020 | Plan for the containment and response to coronavirus cases (COVID-19) in Honduras | Not empirical primary research |
| SecretarÃadeSalud 2020 | Plan para la contenciÃ³n y respuesta a casos de coronavirus (COVID-19) en Honduras | Not empirical primary research |
| SecretariadesaÃºdedoestadodopiauÃ 2020 | NOTA INFORMATIVA CORONAVÃ• RUS | Not empirical primary research |
| Segura 2020 | [Alerted but not panicked. The role of the media during the pandemic.]. | Not empirical primary research |
| Serafim 2020 | The impact of COVID-19 on Brazilian mental health through vicarious traumatization. | Not empirical primary research |
| Serdan 2020 | COVID-19 in Brazil: Historical cases, disease milestones, and estimated outbreak peak. | Not empirical primary research |
| SerraValdes 2020 | COVID-19. De la patogenia a la elevada mortalidad en el adulto mayor y con comorbilidades | Not empirical primary research |
| Shadmi 2020 | Health equity and COVID-19: global perspectives. | Not empirical primary research |
| Shahjouei 2020 | Risk of stroke in hospitalized SARS-CoV-2 infected patients: A multinational study | Not focused in LAC countries |

| | | |
|---|---|---|
| Shanbhag 2020 | Managing cancer care during the covid-19 pandemic-experience at a cancer department in a tertiary hospital in antigua and Barbuda | Not empirical primary research |
| Sigfrid 2020 | Addressing challenges for clinical research responses to emerging epidemics and pandemics: a scoping review. | Not produced in response to the COVID-19 pandemic |
| Sigua-RodrÃguez 2020 | COVID-19 y la odontologÃa: una revisiÃ³n de las recomendaciones y perspectivas para LatinoamÃ©rica | Not empirical primary research |
| Silberman 2020 | PolÃticas de planificaciÃ³n y gestiÃ³n de talento humano del Ministerio de Salud de Argentina durante la pandemia de COVID-19 | Not empirical primary research |
| Silva 2020 | A fragilidade do sistema de saÃºde brasileiro e a vulnerabilidade social diante da COVID-19 | Not empirical primary research |
| Silva 2020 | Fostering the common good in times of COVID-19: the Responsible Innovation in Health perspective | Not empirical primary research |
| Silva 2020 | Implementation of the body management protocol in the context of the new Coronavirus | Not empirical primary research |
| Silva 2020 | Protagonismo da Enfermagem Brasileira no combate ao COVID-19 | Not empirical primary research |
| Silva 2020 | SWOT-TOWS analysis to improve home health waste management during the COVID-19 pandemic | Not empirical primary research |
| Silva 2020 | The unacceptable tragedy of maternal mortality associated with COVID-19: (re)politicization of women's health and rights and the position of Brazilian nursing | Not empirical primary research |
| Silva 2020 | University participation in the production of molecular diagnostic tests for the novel coronavirus in Brazil: the response to health challenges | Not empirical primary research |
| SilvaFreire 2020 | Profissional residente no enfrentamento da COVID-19: relato de experiÃªncia no contexto da enfermagem intensiva | Not empirical primary research |
| Silveira 2020 | Coronary Artery Bypass Graft During the COVID-19 Pandemic | Not empirical primary research |
| Singer 2020 | Deadly Companions: COVID-19 and Diabetes in Mexico | Not empirical primary research |
| SistemaNacionaldeGestiÃ³ndeRiesgo 2020 | AnÃ¡lisis de la situaciÃ³n de salud y apertura econÃ³mica en el contexto de COVID-19 en Honduras | Not empirical primary research |
| Snyder 2020 | Exploring Why Adult Mexican Males Do Not Get Vaccinated: Implications for COVID-19 Preventive Actions | Not focused in LAC countries |
| Soares 2020 | De cuidador a paciente: na pandemia da Covid-19, quem defende e cuida da enfermagem brasileira? | Not empirical primary research |
| Soares 2020 | Surveillance of the sanitary sewage system and SARS-CoV-2 in Brazil: a necessary discussion | Not empirical primary research |
| SociedadeBrasileiradeMedicinadeFamÃ-liae 2020 | OrientaÃ§Ãµes para favelas e periferias sobre o Covid-19 | Not empirical primary research |

| | | |
|---|---|---|
| Solano-Sandi 2020 | Identification and analysis of ongoing registered clinical intervention trials on COVID-19 | Not focused in LAC countries |
| Solorzano-Santos 2020 | Measles outbreak during the COVID-19 pandemic in Mexico | Not empirical primary research |
| Soto 2020 | Rapid responses to the COVID-19 pandemic through science and global collaboration: The solidarity clinical trial | Not empirical primary research |
| Soto 2020 | Respuestas rápidas a la pandemia de COVID-19 a través de la ciencia y la colaboración global: el ensayo clínico Solidaridad | Not empirical primary research |
| Soto 2020 | The feasibility of generalized face mask usage during COVID-19 pandemic: A perspective from Latin America | Not empirical primary research |
| Soto 2020 | The use of drugs without demonstrated effect as a therapeutic strategy in COVID-19 in Peru | Not empirical primary research |
| Sousa 2020 | Enfermagem em contexto de pandemia no Brasil: docilidades dos corpos em questÃ£o | Not empirical primary research |
| Sousa 2020 | Reflections on the nursing process in the work of nurses in front of the COVID-19 pandemic | Not empirical primary research |
| Sousa 2020 | ReflexÃµes sobre o Processo de Enfermagem no trabalho de enfermeiras frente Ã pandemia da Covid-19 | Not empirical primary research |
| SouzaFilho 2020 | COVID-19: the importance of new technologies for physical activity as a public health strategy. | Not empirical primary research |
| Spanemberg 2020 | The impacts of the COVID-19 pandemic on the teaching of dentistry in Brazil | Not empirical primary research |
| SpanishSocietyof 2020 | Survey on COVID-19 Airway Management | Not focused in LAC countries |
| Srikanta 2020 | Examining the status of improved air quality due to COVID-19 lockdown and an associated reduction in anthropogenic emissions | Not focused in LAC countries |
| Stacciarini 2020 | Heroic efforts, sacrifices and leadership: It's all in a day's work for nurses before, during, and after the COVID-19 crisis | Not empirical primary research |
| Standley 2020 | Data and cooperation required for Venezuela's refugee crisis during COVID-19 | Not empirical primary research |
| Staneloni 2020 | DesafÃos en la era del COVID-19 | Not empirical primary research |
| Stein 2020 | COVID-19 Pandemic in Brazil: Institute for Health Metrics and Evaluation Projections and the Ensuing Evolution | Not focused in LAC countries |
| Stella 2020 | Documento de Consenso sobre manejo de los SÃ-ndromes Coronarios Agudos y la pandemia de COVID-19 | Not empirical primary research |
| Strasburg 2020 | COVID-19: Contingency actions for the provision of meals to employees of a university hospital in southern Brazil | Not empirical primary research |
| Subcomisiones 2020 | Trasplante de cÃ©lulas progenitoras hematopoyÃ©ticas (TCPH) Grupo Argentino de Trasplante de MÃ©dula Ã“sea y Terapia Celular (GATMO - TC) - SAH ComisiÃ³n de Infecciones en el Paciente Inmunocomprometido - SADI | Not empirical primary research |

| | | |
|---|---|---|
| | Registro de donantes de CPH y DirecciÃ³n CientÃfico | |
| SusanaRodriguez 2020 | Bronchiolitis in the year of COVID-19. | Not empirical primary research |
| Suzumura 2020 | Challenges for the development of alternative low-cost ventilators during COVID-19 pandemic in Brazil | Not empirical primary research |
| Suzumura 2020 | Desafios para o desenvolvimento de ventiladores alternativos de baixo custo durante a pandemia de COVID-19 no Brasil | Not empirical primary research |
| Sze 2020 | The need for improved discharge criteria for hospitalised patients with COVID-19-implications for patients in long term care facilities | Not focused in LAC countries |
| Tala 2020 | ConexiÃ³n en tiempos de COVID-19 | Not empirical primary research |
| Talal 2020 | Quantifying the Effects of Social Distancing on theSpread of COVID-19 | Not focused in LAC countries |
| Tannuri 2020 | Low incidence of COVID-19 in children and adolescent post-liver transplant at a Latin American reference center. | Not empirical primary research |
| Tanujit 2020 | Theta autoregressive neural network model for COVID-19 outbreak predictions | Not focused in LAC countries |
| Taylor 2020 | Covid-19 misinformation sparks threats and violence against doctors in Latin America | Not empirical primary research |
| Taylor 2020 | Covid-19: Venezuela's doctors refuse to be gagged | Not empirical primary research |
| Taylor 2020 | How Latin America is fighting covid-19, for better and worse | Not empirical primary research |
| Taylor 2020 | Uruguay is winning against covid-19. This is how | Not empirical primary research |
| Taype 2020 | Response of the emergency department of a tertiary hospital during the start of the COVID-19 pandemic in Peru | Not empirical primary research |
| Taype-Rondan 2020 | Regarding the lack of transparency in technical documents for the treatment of patients with COVID-19 in Peru | Not empirical primary research |
| Teheran 2020 | Potential negative effects of the free use of chloroquine to manage COVID-19 in Colombia. | Not empirical primary research |
| Teixeira 2020 | Clinical research protocol to evaluate the effectiveness and safety of individualized homeopathic medicine in the treatment and prevention of the COVID-19 epidemic | Not empirical primary research |
| Teixeira 2020 | COVID-19 and mass gatherings: emerging and future implications of the Brazilian carnival for public health | Not empirical primary research |
| Teixeira 2020 | Epidemiologia clÃnica homeopÃ¡tica na COVID-19: parte 1 | Not empirical primary research |
| Teixeira 2020 | Epidemiologia clÃnica homeopÃ¡tica na COVID-19: parte 2 | Not empirical primary research |
| Teixeira 2020 | Procedimento operacional padrÃ£o na assistÃªncia nutricional ao paciente com covid 19: relato de experiÃªncia | Not empirical primary research |

| | | |
|---|---|---|
| Teixeira 2020 | Protocolo de pesquisa clÃnica para avaliar a eficÃ¡cia e a seguranÃ§a de medicamento homeopÃ¡tico individualizado no tratamento e na prevenÃ§Ã£o da epidemia de COVID-19 | Not empirical primary research |
| Teles 2020 | COVID-19 Pandemic Triggers Telemedicine Regulation and Intensifies Diabetes Management Technology Adoption in Brazil. | Not empirical primary research |
| Tellez 2020 | Trends in peruvian scientific publications on COVID-19: A bibliometric analysis | Not empirical primary research |
| Tello-Ayllon 2020 | Evolution of the COVID19 pandemic in the andean regions of Peru | Not empirical primary research |
| ThaddeusGregory 2020 | The Prostitute, the City, and the Virus | Not empirical primary research |
| Thiago 2020 | Bridging Learning in Medicine and Citizenship During the COVID-19 Pandemic: A Telehealth-Based Strategy | Not empirical primary research |
| ThomÃ© 2020 | Ethical Considerations for Restrictive and Physical Distancing Measures in Brazil During COVID-19: Facilitators and Barriers | Not empirical primary research |
| Tobias 2020 | COVID-19 and Armed Conflict | Not focused in LAC countries |
| Torregrosa 2020 | Recomendaciones generales para los Servicios de CirugÃa en Colombia durante la pandemia COVID-19 (SARS-CoV-2) | Not empirical primary research |
| Torres 2020 | Malaria Situation in the Peruvian Amazon during the COVID-19 Pandemic | Not empirical primary research |
| TorresTorretti 2020 | [COVID-19 and the 3 "P": Pandemic, Pediatrics and impact in the Country] | Not empirical primary research |
| TrapaniJunior 2020 | Protocolo de cuidados no parto, no puerperio e no abortamento durante a pandemia de Covid-19, Childbirth, Puerperium and Abortion Care Protocol during the COVID-19 Pandemic | Not empirical primary research |
| Trevino-Becerra 2020 | Gaceta mÃ©dica de MÃ©xico in SARS-CoV-2 pandemic times | Not empirical primary research |
| Tumba 2020 | Temporal trend of hospitalizations for acute bronchiolitis in infants under one year of age in brazil between 2008 and 2015 | Not produced in response to the COVID-19 pandemic |
| Tzanno-Martins 2020 | Covid-19 pandemic: from carnival masks to surgical masks | Not empirical primary research |
| Ulloa-Gutierrez 2020 | Multisystem Inflammatory Syndrome (MIS-C) Surveillance and COVID-19 in Latin America | Not empirical primary research |
| UniversidadNacional AutÃ³nomade 2020 | Dispositivos mÃ©dicos prioritarios para los servicios de la salud de Honduras en el contexto del COVID-19 | Not empirical primary research |
| UniversidadNacional AutÃ³nomade 2020 | Propuesta integral para el desarrollo de Honduras frente a la pandemia COVID 19 | Not empirical primary research |
| Universityof 2020 | Impact of COVID-19 Pandemic in Perinatal Mental Health (RISEUP-PPD-COVID-19) | Not focused in LAC countries |
| UniversityofNew 2020 | Home-based Kidney Care in Native Americans of New Mexico (HBKC) - Enhancement | Not focused in LAC countries |
| Urrutia-Pereira 2020 | COVID-19 and air pollution: A dangerous association? | Not empirical primary research |

| | | |
|---|---|---|
| Utunen 2020 | Global Reach of an Online COVID-19 Course in Multiple Languages on OpenWHO in the First Quarter of 2020: Analysis of Platform Use Data. | Not focused in LAC countries |
| Utunen 2020 | Launch and global reach of an online COVID-19 course in multiple languages on OpenWHO in the first quarter of 2020 | Not focused in LAC countries |
| Valdez-Garcia 2020 | The role of the medicine student in COVID-19 pandemic. A shared responsibility | Not empirical primary research |
| ValdirMonteirodosSantos 2020 | Forecast of Complete Contamination by COVID-19 in the World, US and South America (April-28-2020) | Not focused in LAC countries |
| Valera 2020 | Orientaciones Ã©ticas para la toma de decisiones mÃ©dicas en el contexto de la pandemia de COVID-19 en Chile | Not empirical primary research |
| Vallinoto 2020 | The challenges of COVID-19 in the Brazilian Amazonian communities and the importance of seroepidemiological surveillance studies | Not empirical primary research |
| Vammen 2020 | Water Resources of Nicaragua and COVID-19: Between panic and apathy?. | Not empirical primary research |
| VanderlanNogueira 2020 | PANDEMIA DE COVID-19 E OS ESFORÃ‡OS DA CIÃŠNCIA PARA COMBATER O NOVO CORONAVÃ• RUS | Not empirical primary research |
| Vargas-Ponce 2020 | Influenza and covid-19 co-infection: Report of five cases in a peruvian hospital | Not empirical primary research |
| Vasquez-Chavesta 2020 | COVID-19 and dengue: Pushing the peruvian health care system over the edge. | Not empirical primary research |
| VelÃ¡squezJimÃ©nez 2020 | 2020, The Year of Nursing in Colombia and its Challenges in the Face of COVID-19 Pandemic | Not empirical primary research |
| Vidal 2020 | Use of mobile health in Cuba to fight COVID-19 | Not empirical primary research |
| Vieira 2020 | [The increase in domestic violence during the social isolation: what does it reveals?]. | Not empirical primary research |
| Vieira 2020 | Isolamento social e o aumento da violÃªncia domÃ©stica: o que isso nos revela? | Not empirical primary research |
| VielmaGuevara 2020 | Pandemia por el SARS-CoV-2: aspectos biolÃ³gicos, epidemiolÃ³gicos y clÃnicos | Not empirical primary research |
| Vigliar 2020 | COVID-19 Pandemic effect on Cytopathology Practice: Results from 23 Laboratories in 11 Countries | Not focused in LAC countries |
| Vijay 2020 | COVID-19 in the West Indies: Trinidad and Tobago Experience | Not empirical primary research |
| Vilela-Estrada 2020 | [Clinical manifestations and course of the first six reported cases of COVID-19 in the medical doctors of Peru] | Not empirical primary research |
| VillalobosDintrans 2020 | It Is Not Just Mortality: A Call From Chile for Comprehensive COVID-19 Policy Responses Among Older People | Not empirical primary research |
| Villulla 2020 | COVID-19 in Argentine agriculture: global threats, local contradictions and possible responses. | Not empirical primary research |
| Virag 2020 | An improved methodology for estimating the prevalence of SARS-CoV-2 | Not focused in LAC countries |

| | | |
|---|---|---|
| Vladimir 2020 | Pandemic in the Network Society. the Comparative Analysis of COVID-19 Spreading Features in Europe and Latin America | Not focused in LAC countries |
| Volpatto 2020 | AvaliaÃ§Ã£o de estratÃ©gias de relaxamento do distanciamento social para o Brasil e estado do Rio de Janeiro | Not focused in LAC countries |
| Walker 2020 | RAPID, RELEVANT CLINICAL GUIDELINES IN A PANDEMIC: ONE INSTITUTION'S EXPERIENCE | Not empirical primary research |
| Walton 2020 | Facility-Level Approaches for COVID-19 When Caseload Surpasses Surge Capacity. | Not empirical primary research |
| Werneck 2020 | A pandemia de COVID-19 no Brasil: cronica de uma crise sanitaria anunciada, The COVID-19 pandemic in Brazil: chronicle of a health crisis foretold | Not empirical primary research |
| Whitworth 2020 | COVID-19: A fast evolving pandemic | Not focused in LAC countries |
| Wickemeyer 2020 | Evolving Management of COVID-19: A Multi-institutional Otolaryngology Perspective. | Not focused in LAC countries |
| Wiesner 2020 | [Investigacion en cancer en el contexto de la pandemia de la enfermedad por SARS-CoV-2] | Not empirical primary research |
| Willem 2020 | SOCRATES: an online tool leveraging a social contact data sharing initiative to assess mitigation strategies for COVID-19. | Not focused in LAC countries |
| WilsonDiasda 2020 | IMPACTOS DA PANDEMIA DE CORONAVÃ• RUS EM UM CAPS INFANTOJUVENIL DO DISTRITO FEDERAL | Not focused in LAC countries |
| Wong 2020 | Argentina study moves needle away from convalescent plasma for COVID-19 | Not empirical primary research |
| Wong 2020 | Effects of national containment measures on decelerating the increase in daily new cases of COVID-19 in 54 countries and four epicenters of pandemic | Not focused in LAC countries |
| Wu 2020 | COVID-19's impact on private practice and academic dentistry in North America. | Not focused in LAC countries |
| XimenesNeto 2020 | CoordenaÃ§Ã£o do cuidado, vigilÃ¢ncia e monitoramento de casos da COVID-19 na AtenÃ§Ã£o PrimÃ¡ria Ã SaÃºde | Not empirical primary research |
| Yanez 2020 | Covid-19 in Peru: from supervised walks for children to the first case of Kawasaki-like syndrome. | Not empirical primary research |
| Yeo 2020 | Review of guidelines and recommendations from 17 countries highlights the challenges that clinicians face caring for neonates born to mothers with COVID-19 | Not empirical primary research |
| Yokoo 2020 | Quality and safety innovations in the Radiology Department during the COVID-19 pandemic: a Latin American experience | Not empirical primary research |
| Younan 2020 | Maximising the impact of social prescribing on population health in the era of COVID-19 | Not empirical primary research |
| YuriethGallardo 2020 | El posgrado frente a la COVID-19 en Granma | Not empirical primary research |

| | | |
|---|---|---|
| Zambrano 2020 | A pregnant woman with COVID-19 in Central America. | Not empirical primary research |
| Zavaleta 2020 | COVID-19: review Indigenous peoples' data. | Not empirical primary research |
| Zepeda 2020 | InfecciÃ³n por SARS-CoV-2 y enfermedad por coronavirus-2019 en pediatrÃa | Not empirical primary research |
| Zhao 2020 | Otolaryngology during COVID-19: Preventive care and precautionary measures. | Not empirical primary research |
| Zuniga-Blanco 2020 | New perspectives for the ambulatory or early treatment of COVID-19 | Not empirical primary research |
| Zylberman 2020 | Development of a hyperimmune equine serum therapy for covid-19 in Argentina | Not empirical primary research |
| | A SARS-CoV-2/COVID-19 em pacientes com cÃ¢ncer | Not focused in LAC countries |
| | Care of the rheumatic patient against the coronavirus (Covid-19). recommendations of the chilean society of rheumatology | Not empirical primary research |
| | Correction to: Strengths and limitations of mathematical models in pandemicsthe case of COVID-19 in Chile | Not empirical primary research |
| | COVID-19 in the Americas: Strategies that Mark the Difference | Not empirical primary research |
| | COVID-19 Outbreak Among College Students After a Spring Break Trip to Mexico - Austin, Texas, March 26-April 5, 2020 | Not empirical primary research |
| | Ecuador's New Life Birthing Centers at the Time of Covid-19 | Not empirical primary research |
| | EPIDEMIOLOGIA DO SURTO DE DOENÃ‡A POR CORONAVÃ• RUS (COVID-19) | Not empirical primary research |
| | Etiologia de las infecciones respiratorias agudas en Guatemala | Not produced in response to the COVID-19 pandemic |
| | 'Little flu' kills 60,000 and counting | Not empirical primary research |
| | MODELAGEM AMBIENTAL PARA COVID-19 (SARS-COV-2) EM SISTEMAS DE ESGOTAMENTO SANITÃ• RIO COMO INSTRUMENTO AUXILIAR NAS AÃ‡Ã•ES DE SAÃšDE PÃšBLICA | Not focused in LAC countries |
| | Reducing the Risks of New Coronavirus in Vulnerable Areas in Brazil | Not empirical primary research |
| | Will novel coronavirus (Covid-19) pandemic impact agriculture, food security and animal sectors? | Not empirical primary research |

73. Brasil. Ministério da Saúde. Fundação Oswaldo C. Saúde mental e atenção psicossocial na pandemia Covid-19: recomendações gerais. 2020.

74. Brasil. Ministério da Saúde. Fundação Oswaldo C. Saúde mental e atenção psicossocial na pandemia Covid-19: recomendações para gestores. 2020.

75. Brasil. Ministério da Saúde. Secretaria de Atenção Especializada à S. Fluxo de pacientes com sintomas respiratórios em unidades de urgência não hospitalares. 2020.

76. Brasil. Ministério da Saúde. Secretaria de Atenção Primária á S. Condutas para a doação de leite materno aos bancos de leite humano e postos de coleta de leite humano no contexto da infecção Covid 19 causada pelo novo Coronavírus (SARS-CoV-2). 2020.

77. Brasil. Ministério da Saúde. Secretaria de Atenção Primária á S. Coronavírus Covid-19: Recomendações Programa Academia da Saúde. 2020.

78. Brasil. Ministério da Saúde. Secretaria de Atenção Primária à S. Atenção à saúde do recém-nascido no contexto da infecção do novo Coronavírus (SARS-CoV-2). 2020.

79. Brasil. Ministério da Saúde. Secretaria de Atenção Primária à S. AtencÌ§aÌƒo aÌ€ sauÌ• de do receÌ• m-nascido no contexto da infeccÌ§aÌƒo pelo novo coronaviÌ• rus (SARS-CoV-2). 2020.

80. Brasil. Ministério da Saúde. Secretaria de Atenção Primária à S. Como registrar no e-SUS APS quando for realizado uma teleconsulta. 2020.

81. Brasil. Ministério da Saúde. Secretaria de Atenção Primária à S. Condutas para a doação de leite materno aos bancos de leite humano e postos de coleta de leite humano no contexto da infecção pelo Coronavírus (SARS-CoV-2). 2020.

82. Brasil. Ministério da Saúde. Secretaria de Atenção Primária à S. Infecção COVID-19 e os riscos às mulheres no ciclo gravídico-puerperal. 2020.

83. Brasil. Ministério da Saúde. Secretaria de Atenção Primária à S. Orientações gerais sobre a atuação do ACS frente à pandemia de COVID-19 e os registros a serem realizados no e-SUS APS. 2020.

84. Brasil. Ministério da Saúde. Secretaria de Atenção Primária à S. Orientações para o registro do FAST-TRACK da abordagem sindrômica de Síndrome Gripal nos atendimentos aos cidadãos com suspeita de H1N1 e COVID-19 no Prontuário Eletrônico do Cidadão (PEC) do e-SUS APS. 2020.

85. Brasil. Ministério da Saúde. Secretaria de Atenção Primária à S. Prevenção ao Covid-19 no âmbito das equipes de consultórios na rua. 2020.

86. Brasil. Ministério da Saúde. Secretaria de Atenção Primária à S. Recomendações para preveção, manejo e controle de casos de COVID-19 no âmbito do sistema socioeducativo. 2020.

87. Brasil. Ministério da Saúde. Secretaria de Atenção Primária à S. Suspensão temporária das consultas eletivas e atividades nos centros de referência da pessoa idosa em razão da contenção de aglomeração em população de risco para o Coronavírus (SARS-CoV-2). 2020.

88. Brasil. Ministério da Saúde. Secretaria de Ciência T e IE em S. Cabine de proteção para o tratamento de pacientes com COVID-19. 2020.

89. Brasil. Ministério da Saúde. Secretaria de Ciência TI e IE em S. Utilização do gás ozônio e da ozonioterapia no combate à disseminação da COVID-19 submetidas pela Sociedade Brasileira de Ozonioterapia Médica (SOBOM). 2020.

90. Brasil. Ministerio da S. Novo Coronavírus: atendimento a pessoas com suspeita de infecção pelo novo coronavírus (2019-nCoV) na Atenção Primária à Saúde. 2020.

91. Brasil. Ministério da S. Coronavírus CoViD-19: Atendimento odontológico no SUS. 2020.

92. Brasil. Ministério da S. Coronavírus Covid-19: Fast-track de teleatendimento para a atenção primária: fluxo rápido - versão 6. 2020.

93. Brasil. Ministério da S. Coronavírus Covid-19: Fast-track de teleatendimento para a atenção primária: fluxo rápido - versão 8. 2020.

94. Brasil. Ministério da S. Coronavírus Covid-19: Fast-track de teleatendimento para a atenção primária: fluxo rápido - versão 9. 2020.

95. Brasil. Ministério da S. Coronavírus CoViD-19: Fast-Track para a atenção primária em locais com transmissão comunitária fluxo rápido - versão 8. 2020.

96. Brasil. Ministério da S. Coronavírus CoViD-19: Fast-Track para a atenção primária em locais com transmissão comunitária fluxo rápido - versão 9. 2020.

97. Brasil. Ministério da S. Coronavírus CoViD-19: Fast-track para a atenção primária em locais com transmissão comunitária fluxo rápido: versão 6. 2020.

98. Brasil. Ministério da S. Coronavírus CoViD-19: Fluxo de manejo clínico de gestantes na atenção especializada. 2020.

99. Brasil. Ministério da S. Coronavírus Covid-19: Fluxo de manejo clínico na atenção primária à saúde em transmissão comunitária: versão 9. 2020.

100. Brasil. Ministério da S. Coronavírus CoViD-19: Fluxo de manejo clínico pediátrico na atenção especializada. 2020.

101. Brasil. Ministério da S. Coronavírus COVID-19: Nota Técnica: saúde mental e apoio e psicossocial na atenção especializada. 2020.

102. Brasil. Ministério da S. Coronavírus Covid-19: Orientações sobre amamentação. 2020.

103. Brasil. Ministério da S. Evitar transmissão intradomiciliar da doença pelo Coronavírus (COVID-19). 2020.

104. Brasil. Ministério da S. Fluxo de atendimento no hospital de referência para paciente referenciado de outros serviços de saúde. 2020.

105. Brasil. Ministério da S. Fluxograma para atendimento e detecção precoce de COVID-19 em hospital de referência para indivíduos por demanda espontânea. 2020.

106. Brasil. Ministério da S. Fluxograma para atendimento e detecção precoce de COVID-19 em pronto atendimento UPA 24 horas e unidade hospitalar não definida como referência. 2020.

107. Brasil. Ministério da S. Orientações do Ministério da Saúde para manuseio medicamentoso precoce de pacientes com diagnóstico da COVID-19. 2020.

108. Brasil. Ministério da S. Recomendações aos Agentes de Combate a Endemias (ACE) para adequação das ações de vigilância e controle de zoonoses frente à atual situação epidemiológica referente ao Coronavírus (COVID-19). 2020.

114. Caballero RL. Political will and intersectorality to confront COVID-19 in Cuba. Rev Habanera Ciencias Medicas. 2020;19: e_3241.

115. Cabanillas-Rojas W. BEHAVIOR AND SPREAD OF COVID-19 IN PERU: A FRAMEWORK FOR THE DESIGN BEHAVIORAL PUBLIC HEALTH INTERVENTIONS. 2020.

137. Carolina C. Chatting with my theoretical frameworks: Live sessions on Instagram as an opportunity to reflect in times of coronavirus. 2020;11.

148. Cavalcante TM, Mendes FL, Perez C de A, Carvalho AOR de, Teixeira APL, Viegas JRR. Como a Política Nacional de Controle do Tabaco Pode Ajudar no Enfrentamento da Pandemia da Covid-19? Rev bras cancerol. 2020;66: e-1023.

163. Christovam Reis dos Santos F, Otávio José Lemos C. IGREJA UNIVERSAL E O CORONAVIRUS: a dimensão geossimbólica e negação do isolamento social. 2020;5.

173. Conselho Nacional de Secretarias Municipais de S. Guia orientador para o enfrentamento da pandemia COVID-19 na Rede de Atenção à Saúde. 2020.

174. Conselho Nacional de Secretários de S. Second phase of the research “Evolution of the Prevalence of Infection by Covid-19 in Brazil: Population-Based Study.” CONASS Inf. 2020;253.

183. Cruz JR, Fernández A de, Peralta F, Cano F, Cáceres P. Etiologia de las infecciones respiratorias agudas en Guatemala. p. 21.

193. Daumas RP, Silva GAE, Tasca R, Leite I da C, Brasil P, Greco DB, et al. The role of primary care in the Brazilian healthcare system: limits and possibilities for fighting COVID-19. O Pap da

atencao primaria na rede atencao a saude no Bras limites e possibilidades no enfrentamento da COVID-19. 2020;36: e00104120. doi:https://dx.doi.org/10.1590/0102-311X00104120

206. de Sandes-Freitas TV, Regina Canito Brasil I, de Mattos Brito Oliveira ML, Seixas Studart E Neves Lunguinho M, Pimentel IRS, da Costa LATJ, et al. Lessons from SARS-CoV-2 screening

in a Brazilian Organ Transplant Unit. Transpl Infect Dis. 2020; e13376. doi:https://dx.doi.org/10.1111/tid.13376

207. Del Castillo R, Martinez D, Sarria GJ, Pinillos L, Garcia B, Castillo L, et al. Low-dose radiotherapy for COVID-19 pneumonia treatment: case report, procedure, and literature review. Strahlenther Onkol. 2020. doi:10.1007/s00066-020-01675-z

208. Dentzer S. Disruptive Innovation in rural cancer care: Opening the doors amid the pandemic. JCO Oncol Pract. 2020;16: 414–416. doi:http://dx.doi.org/10.1200/OP.20.00401

209. Devakumar D, Bhopal SS, Shannon G. COVID-19: the great unequaliser. J R Soc Med. 2020;113: 234–235. doi:http://dx.doi.org/10.1177/0141076820925434

210. Dias FLT, Mendonça FD, Pinto GM, Borges ISC, Oliveira SV de. Respiratory diseases in the Triângulo Mineiro: Epidemiological analysis and projective with the COVID-19 pandemic. J Heal Biol Sci. 2020;8.

228. Edesa Biotech I. A Randomized, Double-Blind, Placebo-Controlled Study to Evaluate the Safety and Efficacy of EB05 + SOC vs. Placebo + SOC in Adult Hospitalized Patients With Moderate to Severe COVID-19 Pneumonia. clinicaltrials.gov. 2020.

229. Eduardo Amaral H, Fernando Salgueiro P, Inácio Fernandes de A. Input-Output Analysis of COVID-19: Methodology for Assessing the Impacts of Lockdown Measures. 2020.

230. Effenberger M, Kronbichler A, Shin J Il, Mayer G, Tilg H, Perco P. Association of the COVID-19 pandemic with Internet Search Volumes: A Google TrendsTM Analysis. Int J Infect Dis. 2020;95: 192–197. doi:https://dx.doi.org/10.1016/j.ijid.2020.04.033

231. Elera-Fitzcarrald C, Ugarte-Gil MF, Alarcon GS. COVID-19 and Its Potential Effect on Patients With Rheumatic Diseases in Latin America. J Clin Rheumatol. 2020. doi:https://dx.doi.org/10.1097/RHU.0000000000001493

232. Eliseu José W, Lucas Manassi P, Ricardo D. Monitoramento do Coronavírus (Covid-19) nos municípios do Rio Grande do Sul, Brasil. SocArXiv. 2020.

233. Emanuelle Freitas G, Dandara de Oliveira R, Andrea Jacqueline Fortes F. Racial health inequalities and the COVID-19 pandemic Desigualdades raciais em saúde e a pandemia da Covid-19/ Racial health inequalities and the COVID-19 pandemic Las desigualdades raciales en la salud y la pandemia del Covid-19. Trab Educ e Saúde. 2020;18.

256. Fontanella MM, Saraceno G, Lei T, Bederson JB, You N, Rubiano AM, et al. Neurosurgical activity during COVID-19 pandemic: an expert opinion from China, South Korea, Italy, United

Stated of America, Colombia and United Kingdom. J Neurosurg Sci. 2020. doi:https://dx.doi.org/10.23736/S0390-5616.20.04994-2

271. Gaojun W, You-Gan W, Qibin D, Jinran W. Predication of Inflection Point and Outbreak Size of COVID-19 in New Epicentres. arXiv Appl. 2020.

279. García-Rey T. Colombia en tiempos de COVID-19. Acta otorrinolaringol cir cabeza cuello. 2020;48.

280. Garcia-Silva R, Hernandez-Dono S, Mena L, Granados J. MEXICAN HUMAN LEUKOCYTE ANTIGEN ALLELES MIGHT PREDICT CLINICAL OUTCOME IN SARS-COV-2 INFECTED PATIENTS. Rev Invest Clin. 2020;72: 178–179. doi:https://dx.doi.org/10.24875/RIC.20000135

281. Garrido RG, Almeida MP de. Impasses entre dignidade e saúde no manejo de cadáveres da COVID-19: identificar ou reconhecer? Comun ciênc saúde. 2020;31.

282. Garrido RG, Almeida MP de. Stalemates between dignity and health in the handling of COVID-19 corpses: identify or recognize? Comun ciênc saúde. 2020;31.

283. Garrido RG, Rodrigues RC. Restrição de contato social e saúde mental na pandemia: possíveis impactos das condicionantes sociais. J Heal Biol Sci. 2020;8: 1–9.

284. Garrido RG, Rodrigues RC. Restriction of social contact and mental health in pandemic: possible impacts of social constraints. J Heal Biol Sci. 2020;8: 1–9.

285. Gaspari CH, Assumpcao I, Freire R, Silva A, Santiso C, Jaccoud AC. The First 60 Days: Physical Therapy in a Neurosurgical Center Converted Into a COVID-19 Center in Brazil. Phys Ther. 2020. doi:http://dx.doi.org/10.1093/ptj/pzaa175

286. Gauna ME, Bernava JL. Recomendaciones diagnósticas y terapéuticas ante la Respuesta Inmune Trombótica Asociada a COVID-19 (RITAC). CorSalud. 2020;12.

294. Goiás . Secretaria da S. A Hidroxicloroquina eÌ• eficaz e segura no tratamento da COVID-19? 2020.

295. Goiás . Secretaria da S. Atualiza informações sobre o uso da Cloroquina como terapia adjuvante no tratamento de formas graves do COVID-19. 2020.

296. Goiás . Secretaria da S. COVID-19 em crianças e reabertura de escolas. 2020.

297. Goiás . Secretaria da S. Dispõe sobre recomendações para investigação e notificação da COVID19, no SINAN, na ficha de acidente de trabalho, por ser considerada doença ocupacional. 2020.

298. Goiás . Secretaria da S. Manejo ambiental de dengue e situação de pandemia do COVID-19. 2020.

299. Goiás . Secretaria da S. Multimorbidade e COVID-19. 2020.

300. Goiás . Secretaria da S. Orientações para a utilização de cloroquina e hidroxicloroquina no tratamento de pacientes com COVID-19. 2020.

301. Goiás . Secretaria da S. Orientações para o atendimento de gestantes e puérperas diante da pandemia do COVID-19. 2020.

302. Goiás . Secretaria da S. Recomendações dos Tutores Estaduais para a estratégia de Atenção Humanizada ao Recém-Nascido: Método Canguru, frente aos desafios enfrentados diante da pandemia decorrente de COVID-19. 2020.

303. Goiás . Secretaria da S. Recomendações gerais para implantação de medidas de prevenção e controle de casos de COVID-19 em empresas no estado de Goiás. 2020.

304. Goiás . Secretaria da S. Sobre serviços de somatoconservação, funerárias, cemitérios e crematórios no manejo de corpos de vítimas de casos suspeitos ou confirmados por COVID-19. 2020.

305. Goiás . Secretaria da S. Uso de máscaras pela população em geral. 2020.

312. Governo do Estado de Santa C. Guia de convívio responsável: regras do coronavírus. 2020.

321. Guimarães Jr PR. Vulnerabilidade das microrregiões do Estado de São Paulo à pandemia do novo coronavírus (SARS-CoV-2). 2020.

322. Gutierrez Cortes WA. Role of the Colombian geriatrician in the face of the pandemic by COVID-19. Rev Esp Geriatr Gerontol. 2020. doi:http://dx.doi.org/10.1016/j.regg.2020.06.010

323. Gutierrez-Aguilar R, Cordova-Lepe F, Munoz-Quezada MT, Gutierrez-Jara JP. [Model for a threshold of daily rate reduction of COVID-19 cases to avoid hospital collapse in Chile]. Medwave. 2020;20: e7871. doi:https://dx.doi.org/10.5867/medwave.2020.03.7871

324. Hallak J, Esteves SC. Concise practice recommendations for the provision of andrological services and assisted reproductive technology for male infertility patients during the SARS-

CoV-2 in Brazil. Int Braz J Urol. 2020;46: 1082–1089. doi:10.1590/S1677-5538.IBJU.2020.06.03

349. Humerez DC de, Ohl RIB, Silva MCN da. Mental health of brazilian nursing professionals in the context of the Covid-19 pandemic: action by the Nursing Federal Council. Cogitare enferm. 2020;25.

358. Institut P. COVID-19 Seroprevalence Study in French Guiana. clinicaltrials.gov. 2020.

359. Instituto de Efectividad Clínica y S. Barbijos y protectores faciales en población general en la pandemia por COVID-19. 2020.

412. López JAH, Ochoa-Martínez PY. Enseñanza virtual en educación física en primaria en México y la pandemia por COVID-19. Cienc act fís (Talca, En línea). 2020;21.

413. Lopez-Alvarez D, Parra B, Cuellar WJ. Genome Sequence of SARS-CoV-2 Isolate Cali-01, from Colombia, Obtained Using Oxford Nanopore MinION Sequencing. Microbiol Resour Announc. 2020;9. doi:https://dx.doi.org/10.1128/MRA.00573-20

414. López-Feldman A, Chávez C, Vélez MA, Bejarano H, Chimeli AB, Féres J, et al. Environmental Impacts and Policy Responses to Covid-19: A View from Latin America. Environ Resour Econ. 2020; 1–6. doi:10.1007/s10640-020-00460-x

415. Lopez-Ortiz E, Lopez-Ortiz G, Mendiola-Pastrana IR, Mazon-Ramirez JJ, Diaz-Quinonez JA. From the handling of an outbreak by an unknown pathogen in Wuhan to the preparedness and response in the face of the emergence of Covid-19 in Mexico. Gac Med Mex. 2020;156: 132–137. doi:https://dx.doi.org/10.24875/GMM.M20000346

416. Lorenz C, Dias Bocewicz AC, Corrêa de Azevedo Marques C, Reis Santana LM, Chiaravalloti-Neto F, Alves Gomes AH, et al. Have measures against COVID-19 helped to reduce dengue cases in Brazil? Travel Med Infect Dis. 2020; 101827. doi:10.1016/j.tmaid.2020.101827

417. Lucas F, Bergmann A, Bello M, Tonellotto F, Caiado Neto B. Reconstrução Mamária em Pacientes Oncológicos durante a Pandemia da Covid-19. Rev bras cancerol. 2020;66: e-1004.

418. Luis Enrique H-V, Lupe Antonieta V-Z. La salud pública y el cuidado materno fetal y neonatal en la pandemia por el virus SARS-CoV-2. 2020;5.

425. Magallanes-Garza GI, Valdez-Alatorre C, Dávila-González D, Martínez-Reséndez MF, Sánchez-Salazar SS, Castilleja-Leal F, et al. Rapid improvement of a critically ill obstetric

patient with SARS-CoV-2 infection after administration of convalescent plasma. Int J Gynaecol Obstet. 2020. doi:10.1002/ijgo.13467

435. Maria E, Mancilla R. SARS-CoV2 (COVID-19) y las figuras fiscales-legales. 2020;3.

448. Martins-Filho PR, Santos VS. No evidence supports the use of ether and chloroform inhalation for treating COVID-19. Rev panam salud pública. 2020;44.

468. Minas Gerais . Secretaria de S. Plano de Contingência Operativo para infecção humana pelo SARS COV-2 (doença pelo Coronavírus - COVID-19) da Macrorregião Jequitinhonha. 2020.

469. Minas Gerais . Secretaria de S. Plano de Contingência Operativo para infecção humana pelo SARS COV-2 (doença pelo Coronavírus - COVID-19) da Macrorregião Nordeste. 2020.

470. Minas Gerais . Secretaria de S. Plano de Contingência Operativo para infecção humana pelo SARS COV-2 (doença pelo Coronavírus - COVID-19) da Macrorregião Noroeste. 2020.

471. Minas Gerais . Secretaria de S. Plano de Contingência Operativo para infecção humana pelo SARS COV-2 (doença pelo Coronavírus - COVID-19) da Macrorregião Norte. 2020.

472. Minas Gerais . Secretaria de S. Plano de Contingência Operativo para infecção humana pelo SARS COV-2 (doença pelo Coronavírus - COVID-19) da Macrorregião Oeste. 2020.

473. Minas Gerais . Secretaria de S. Plano de Contingência Operativo para infecção humana pelo SARS COV-2 (doença pelo Coronavírus - COVID-19) da Macrorregião Sudeste. 2020.

474. Minas Gerais . Secretaria de S. Plano de Contingência Operativo para infecção humana pelo SARS COV-2 (doença pelo Coronavírus - COVID-19) da Macrorregião Sul. 2020.

475. Minas Gerais . Secretaria de S. Plano de Contingência Operativo para infecção humana pelo SARS COV-2 (doença pelo Coronavírus - COVID-19) da Macrorregião Triângulo do Norte. 2020.

476. Minas Gerais . Secretaria de S. Plano de Contingência Operativo para infecção humana pelo SARS COV-2 (doença pelo Coronavírus - COVID-19) da Macrorregião Triângulo do Sul. 2020.

477. Minas Gerais . Secretaria de S. Plano de Contingência Operativo para infecção humana pelo SARS COV-2 (doença pelo Coronavírus - COVID-19) da Macrorregião Vale do Aço. 2020.

503. Mota E, Glória Teixeira M. Epidemiological surveillance and the Covid-19 pandemic in Brazil. Elements to understand the Brazilian response and the explosion of cases and deaths. 2020.

519. Nct. A Randomized, Double-Blind, Placebo-Controlled Study to Evaluate the Safety and Efficacy of EB05 + SOC vs. Placebo + SOC in Adult Hospitalized Patients With Moderate to Severe COVID-19 Pneumonia. 2020.

520. Nct. Evaluation of the Scleroderma Patient-centered Intervention Network COVID-19 Home-isolation Activities Together Program. 2020.

521. Nct. Protection of Health Workers Against COVID-19. 2020.

522. Nct. Protective Effect of Aspirin on COVID-19 Patients. 2020.

530. Novaes ita CG, Silva RE da, Sallas J, Novaes LCG, Guilhem DB. Desafios na condução de pesquisas sobre a COVID-19 envolvendo seres humanos no Brasil. Comun ciênc saúde. 2020;31.

582. p RBR. Oral hygiene in critically ill patients. 2019.

583. p RBR. Oral hygiene in critically ill patients. 2019.

584. Pablos-Méndez A, Vega J, Aranguren FP, Tabish H, Raviglione MC. Covid-19 in Latin America. BMJ. 2020;370: m2939. doi:10.1136/bmj.m2939

585. Palamim CVC, Ortega MM, Marson FAL. COVID-19 in the Indigenous Population of Brazil. J racial Ethn Heal disparities. 2020. doi:http://dx.doi.org/10.1007/s40615-020-00885-6

586. Pan American Health O. Retrofit Measures for COVID-19, 10 June 2020. 2020.

603. Peres KG, Reher P, Castro RD de, Vieira AR. COVID-19-related challenges in dental education: experiences from Australia, Brazil, and the USA. 2020.

609. Perla Garcia R, Daniel Gonzalo Hernandez R, Luis Octavio Ramírez F, Veronica Velazquez S, Silvia Ruiz-Velasco A, Genaro Javier Delgado C, et al. Increase in Relative Humidity Increases

the COVID-19 Contagion Rate and the Increase in UV Index Decreases It. SSRN. 2020. doi:10.2139/ssrn.3688287

610. Perondi B, Miethke-Morais A, Montal AC, Harima L, Segurado AC. Setting up hospital care provision to patients with COVID-19: lessons learnt at a 2400-bed academic tertiary center in Sao Paulo, Brazil. Brazilian J Infect Dis. 2020. doi:http://dx.doi.org/10.1016/j.bjid.2020.09.005

611. Peru. EsSalud. Instituto de Evaluación de Tecnologías en Salud e I. Uso de la telemedicina en covid: seguimiento y manejo de casos positivos. 2020.

612. Peru. EsSalud. Instituto de Evaluación de Tecnologías en Salud e I. Uso de tocilizumab para el tratamiento de pacientes con COVID-19. 2020.

613. Peru. EsSalud. Instituto de Evaluación de Tecnologías en Salud e I. Uso de ventilacion multiple con un solo ventilador. 2020.

625. Ponce-Torres C. Carta a la editora - Participación de los estudiantes de medicina de Latinoamérica en la pandemia por COVID-19. Medwave. 2020;20.

647. Red Argentina Pública de Evaluación de Tecnologías S. Uso de medidas de barrera específicas y búsqueda activa para la prevención de la diseminación de SARS-COV2 en el ámbito quirúrgico. 2020.

648. Reis HLB dos, Boldrini NAT, Caldas JVJ, Paz APC da, Ferrugini CLP, Miranda AE. Severe coronavirus infection in pregnancy: challenging cases report. Rev Inst Med Trop Sao Paulo. 2020;62. doi:10.1590/s1678-9946202062049

649. Reis VP, Maia ABP, Bezerra AR, Castex D. O novo normal da Odontologia: revisão das recomendações para retomada da assistência odontológica durante a pandemia da COVID-19. Rev bras odontol. 2020;77: 1–9.

650. Reis VP, Maia ABP, Bezerra AR, Conde DC. Uso dos Equipamentos de Proteção Individual no Atendimento Odontológico Durante Surto da COVID-19 e Alternativas em Períodos de Desabastecimento: Revisão Integrativa. Rev bras odontol. 2020;77: 1–9.

651. República D, Ministerio de Salud P. Boletín especial No. 129: Enfermedad por Coronavirus 2019 (COVID-19). 2020.

652. Resiere D, Kallel H. Implementation of Medical and Scientific Cooperation in the Caribbean Using Blockchain Technology in Coronavirus (Covid-19) Pandemics. J Med Syst. 2020;44: 123. doi:http://dx.doi.org/10.1007/s10916-020-01589-4

653. Ribas RM. Coronavirus Disease 2019 (COVID-19) and healthcare-associated infections: Emerging and future challenges for public health in Brazil. Travel Med Infect Dis. 2020; 101675. doi:10.1016/j.tmaid.2020.101675

654. Ribas RM, Campos PA de, Brito CS de, Gontijo-Filho PP. Coronavirus Disease 2019 (COVID-19) and healthcare-associated infections: Emerging and future challenges for public health in Brazil. Travel Med Infect Dis. 2020; 101675. doi:https://dx.doi.org/10.1016/j.tmaid.2020.101675

655. Ribeiro Conceição M, Pascoal Freire R, Côrtes Macario F, Alves Torres Oliveira R, Maia Franco C. Covid-19 - An effective municipal coordination and articulation exercise: the Niterói experience. 2020.

669. Rio de Janeiro . Secretaria de Estado de S. Prevention measures and care for suspected cases of infection with the new coronavirus in maternity hospitals in the state of Rio de Janeiro. Nota técnica conjunta nº 01 SRUP/SGAIS SES/RJ. 2020; 14.

670. Rio Grande do Sul . Centro Estadual de Vigilância em S. Boas práticas para prevenção do Coronavírus (COVID-19) em supermercados. 2020. p. Good practices for Coronavirus prevention (COVID-19) in supermarkets.

671. Rio Grande do Sul . Centro Estadual de Vigilância em S. COVID-19: Medidas de biossegurança em estabelecimentos de saúde, funerários e congêneres e cuidados após a morte. 2020.

672. Rio Grande do Sul . Secretaria da S. Orientações aos serviços da rede de atenção psicossocial sobre estratégias de prevenção de disseminação do COVID-19. 2020.

673. Rio Grande do Sul . Secretaria da S. Orientações para o cuidado em saúde à população em situação de rua em razão da disseminação do coronavírus. 2020.

674. Rio Grande do Sul . Secretaria da S. Orientações técnicas para as maternidades em período da COVID-19. 2020.

675. Rio Grande do Sul . Secretaria da S. Recomendações para contenção da infecção humana pelo novo Coronavírus (COVID-19) em povos indígenas no Estado do RS. 2020.

676. Riquelme P A. Recomendaciones de Asociaciones Chilenas de Endoscopia Digestiva (ACHED), de Hepatología (ACHHEP) y la Agrupación Chilena de Trabajo en Enfermedad de Crohn y Colitis Ulcerosa (ACTECCU) para el manejo de pacientes con patologías gastrointestinales en pandemia SARS-CoV-2. Gastroenterol latinoam. 2020;31: 7–8.

677. Rizo-Patron E, Padilla J, Tantalean JA. Pediatric hospital demand in the COVID-19 pandemic. Acta Medica Peru. 2020;37: 376–381. doi:http://dx.doi.org/10.35663/amp.2020.373.1000

678. Rizo-Patron E, Padilla J, Tantalean JA. Pediatric hospital demand in COVID-19 times in Perú. 2020.

679. Rodrigues I de A, Cavalcante JR, Faerstein E. Pandemia de Covid-19 e a saúde dos refugiados no Brasil. 2020.

706. Sales CMM, Silva AI da, Maciel ELN. COVID-19 health surveillance in Brazil: seeking contacts in communities as a self-protection strategy. 2020.

707. Salles Neto LL de, Martins CB, Chaves AA, Konstantyner TCR de O, Yanasse HH, Campos CBL de, et al. Forecast UTI: application for predicting intensive care unit beds in the context of the COVID-19 pandemic. Forecast UTI Apl para Previs leitos unidades Ter intensiva no Context da pandemia COVID-19. 2020;29: e2020391. doi:https://dx.doi.org/10.5123/S1679-49742020000400023

708. Sampaio SG dos SM, Dias AM, Freitas R de. Avaliação do Plano de Ação Implementado pelo Serviço Médico de uma Unidade de Referência em Cuidados Paliativos Oncológicos frente à Pandemia de Covid-19. Rev bras cancerol. 2020;66: e-1158.

718. Santa Catarina . Secretaria da S. Como proteger sua saúde mental em tempos de Coronavírus. 2020.

719. Santa Catarina . Secretaria da S. Coronavírus COVID-19: Recomendações ao atendimento odontológico no SUS em Santa Catarina. 2020.

720. Santa Catarina . Secretaria da S. Fluxo de atendimento aos casos suspeitos, prováveis e confirmados do novo Coronavírus (COVID-19). 2020.

721. Santa Catarina . Secretaria da S. Medidas de enfrentamento à violência doméstica no contexto da pandemia de COVID-19. 2020.

722. Santa Catarina . Secretaria da S. Medidas de enfrentamento ao COVID-19 no sistema prisional. 2020.

723. Santa Catarina . Secretaria da S. Orientação para profissionais de saúde no uso de práticas integrativas e complementares no período de pandemia do COVID-19: versão 1. 2020.

724. Santa Catarina . Secretaria da S. Orientações sobre o atendimento da gestante, puérpera e recém nascido no enfrentamento à pandemia de COVID-19. 2020.

725. Santa Catarina . Secretaria da S. Recomendações às populações indígenas para o enfrentamento à pandemia de COVID-19. 2020.

726. Santa Catarina . Secretaria da S. Recomendações para os centros de atenção psicossocial (CAPS) para prevenção e controle de infecções pelo novo Coronavírus. 2020.

727. Santa Catarina . Secretaria da S. Recomendações sobre o trabalho dos agentes comunitários de saúde (acs) na atenção primária à saúde para o enfrentamento da pandemia de COVID-19. 2020.

740. São Paulo . Secretaria da S. Serological survey for SARS-COV-2: prevalence of infection among schoolchildren in the city of São Paulo: results of phase 1. 2020.

741. São Paulo Secretaria da Saúde. Centro de Vigilância E, quot, Prof. Alexandre V, quot. Epidemiological surveillance of pediatric multisystem inflammatory syndrome (SIM-P), temporarily associated with COVID-19 in the State of São Paulo, epidemiological weeks 01 to 37, 2020. 2020; 4.

742. São Paulo Secretaria da Saúde. Centro de Vigilância E, quot, Prof. Alexandre V, quot. Epidemiological surveillance of pediatric multisystem inflammatory syndrome (SIM-P), temporarily associated with COVID-19 in the State of São Paulo, epidemiological weeks 01 to 41 of 2020. 2020; 5.

743. São Paulo Secretaria da Saúde. Centro de Vigilância E, quot, Prof. Alexandre V, quot. Epidemiological surveillance of pediatric multisystem inflammatory syndrome (SIM-P), temporarily associated with COVID-19 in the State of São Paulo, epidemiological weeks 01 to 42, 2020. 2020; 5.

750. Scripps Whittier Diabetes I. Dulce Digital-COVID Aware (DD-CA) Discharge Texting Platform for US/Mexico Border Hispanics With Diabetes + COVID-19. clinicaltrials.gov. 2020.

751. Secretaría de Salud HN. Guía para el funcionamiento de los equipos de respuesta rápida en el contexto de COVID-19 Honduras. 2020. p. 27.

752. Secretaría de Salud hn. Plan for the containment and response to coronavirus cases (COVID-19) in Honduras. 2020; 38.

777. Soares AFS, Nunes BCR, Costa FCR, Silva LF de M, Souza e Souza LP. Surveillance of the sanitary sewage system and SARS-CoV-2 in Brazil: a necessary discussion. 2020.

778. Soares SSS, Souza NVD de O, Carvalho EC, Varella TCM y ML, Andrade KBS de, Pereira SRM, et al. De cuidador a paciente: na pandemia da Covid-19, quem defende e cuida da enfermagem brasileira? Esc Anna Nery Rev Enferm. 2020;24: e20200161–e20200161.

779. Sociedade Brasileira de Medicina de Família e C, Associação de Medicina de Família e Comunidade do Rio de J. Orientações para favelas e periferias sobre o Covid-19. 2020.

790. Spanish Society of A, Critical C. Survey on COVID-19 Airway Management. clinicaltrials.gov. 2020.

798. Subcomisiones S. Trasplante de células progenitoras hematopoyéticas (TCPH) Grupo Argentino de Trasplante de Médula Ósea y Terapia Celular (GATMO - TC) - SAH Comisión de Infecciones en el Paciente Inmunocomprometido - SADI Registro de donantes de CPH y Dirección Científico. 2020;24.

799. Susana Rodriguez M. Bronchiolitis in the year of COVID-19. Arch Argent Pediatr. 2020;118: 222–223. doi:https://dx.doi.org/10.5546/aap.2020.222

800. Suzumura EA, Zazula AD, Moriya HT, Fais CQA, Alvarado AL, Cavalcanti AB, et al. Challenges for the development of alternative low-cost ventilators during COVID-19 pandemic in Brazil. Rev Bras Ter intensiva. 2020;32: 444–457. doi:http://dx.doi.org/10.5935/0103-507X.20200075

801. Suzumura EA, Zazula AD, Moriya HT, Fais CQA, Alvarado AL, Cavalcanti AB, et al. Desafios para o desenvolvimento de ventiladores alternativos de baixo custo durante a pandemia de

COVID-19 no Brasil. Rev Bras Ter intensiva. 2020;32: 444–457. doi:10.5935/0103-507x.20200075

837. University of New M. Home-based Kidney Care in Native Americans of New Mexico (HBKC) - Enhancement. clinicaltrials.gov. 2020.

838. University of M. Impact of COVID-19 Pandemic in Perinatal Mental Health (RISEUP-PPD-COVID-19). clinicaltrials.gov. 2020.

851. Vidal MNV, Ledo MJV, Matar RP. Use of mobile health in Cuba to fight COVID-19. Rev Cuba salud pública. 2020;46: e:2545-e:2545.

870. Wilson Dias da C, Clara Correa L, Amanda Teixeira B, Gabrielle Silva M. IMPACTOS DA PANDEMIA DE CORONAVÍRUS EM UM CAPS INFANTOJUVENIL DO DISTRITO FEDERAL. 2020;1.

871. Wong CKH, Wong JYH, Tang EHM, Au CH, Lau KTK, Wai AKC. Effects of national containment measures on decelerating the increase in daily new cases of COVID-19 in 54 countries and four epicenters of pandemic. J Med Internet Res. 2020. doi:10.2196/19904

872. Wong S. Argentina study moves needle away from convalescent plasma for COVID-19. News - BIOCENTURY. 2020.

880. Yurieth Gallardo S, Madelín Fonseca A, Sucell Inés García R, Lisbeth Mesa G. El posgrado frente a la COVID-19 en Granma. 2020;24.
